# Pulse oximeter performance and skin pigment: comparison of 34 oximeters using current and emerging regulatory frameworks

**DOI:** 10.1101/2025.08.11.25332026

**Authors:** Caroline Hughes, Danni Chen, Tyler Law, Philip Bickler, John Feiner, Leonid Shmuylovich, Ella Behnke, Lily Ortiz, Gregory Leeb, Isabella Auchus, Fekir Negussie, Ronald Bisegerwa, René Vargas Zamora, Elizabeth Igaga, Kelvin Moore, Olubunmi Okunlola, Ellis Monk, Jana Lyn Fernandez, Odinakachukwu Ehie, Bernadette Wilks, Koyinsola Oyefeso, Deleree Schornack, Cornelius Sendagire, Michael S. Lipnick

## Abstract

**Background:** The International Organization for Standardization (ISO) and US Food and Drug Administration (FDA) are updating regulations for pulse oximeters to reduce performance disparities linked to skin pigment. We tested common oximeters with current and anticipated regulatory frameworks. We hypothesized that not all oximeters show more positive bias in darkly vs lightly pigmented participants and that few oximeters would ‘pass’ the anticipated FDA regulations.

**Methods:** We used a controlled desaturation protocol to test 34 oximeters across arterial oxygen saturations (SaO_2_) 70–100% in healthy adults. Based on what FDA and ISO had shared at the time of study design, we studied cohort sizes of ≥ 24 with ≥ 25% of participants being darkly pigmented. We used the subjective Monk Skin Tone (MST) scale and the objective individual typology angle (ITA) derived from a spectrophotometer to characterize skin pigment. The root mean square error (A_RMS_), bias (mean of SpO_2_ - SaO_2_ error), and skin pigment differential bias were calculated. Monte Carlo simulation explored potential impacts of participant selection on device passing.

**Results:** For cohorts of 24 participants, 28/34 oximeters passed 2017 ISO Standard (A_RMS_ ≤ 4%), 22/34 passed 2013 FDA guidance (A_RMS_ ≤ 3%), 21/34 oximeters passed both A_RMS_ and differential bias criteria for anticipated ISO standards, and 1/34 passed anticipated FDA criteria. More devices passed with cohorts > 24. Eleven oximeters had more positive bias in participants with dark vs. light (dorsal finger) pigmentation across 70–100% SaO_2_. Eighteen devices could pass or fail depending on cohorts selected for analysis.

**Conclusions:** Pulse oximeters show variable performance across manufacturers and models. Notably, only some devices show more positive bias in people with darker skin. Anticipated updates to ISO and FDA frameworks yield strikingly different assessments and require refinement of cohort sizes and differential bias criteria. Whether new guidelines will translate into improved real-world performance or reduced health disparities is yet to be determined.

## Introduction

Pulse oximeters are essential clinical tools that noninvasively estimate the percent of hemoglobin bound with oxygen (SpO_2_). However, long-standing concerns over variable performance across manufacturers and worse performance in people with darker skin have only recently received significant attention. Reports of oximeter inaccuracies in people with darker skin pigmentation began in the 1980s, and the COVID-19 pandemic brought this issue to the forefront as numerous studies reported not only positive bias (i.e. SpO_2_ overestimating true arterial blood functional oxygen saturation, SaO_2_) but also healthcare disparities (i.e. underrecognition or undertreatment of hypoxemia) in patients who self-identified as Black, Asian, Hispanic, or Native American.^1–4^ Laboratory studies in healthy participants have shown similar findings for some oximeters, especially at lower SaO_2_.^5–8^

In response to these concerns, the International Organization for Standardization (ISO) and US Food and Drug Administration (FDA) began updating pulse oximeter regulatory frameworks to improve performance and reduce potential disparities related to skin pigment. The FDA 510(k) premarket notification guidance for pulse oximeters and ISO 80601-2-61 standards were last updated in 2013 and 2017, respectively, and recommend manufacturers verify pulse oximeter performance using “10 or more healthy participants that vary in age and gender.”^9,10^ The 2013 FDA guidance also recommended at least 15% of the participants (i.e., the proportion of the US population in 2013 who identified as African American) should have “darkly pigmented” skin. As previously reported, these recommendations are likely underpowered to ensure equitable device performance.^11^ In 2024, the FDA released a discussion paper proposing larger verification study cohorts (24 participants) with more diversity. In 2025, FDA released a new draft guidance recommending even larger cohorts (150 participants), with more diversity of skin pigment, and additional statistical analyses.^12^ The ISO has shared some details of anticipated changes via public forums.^13^ Both agencies are expected to imminently release finalized versions of their new regulatory frameworks for pulse oximeters.^13–16^

Considering these upcoming changes and ongoing uncertainty about the extent to which oximeters on the market have bias related to skin color, we sought to independently test the performance of 34 commonly used pulse oximeters with both current and anticipated regulatory frameworks. We hypothesized that relatively few devices would pass anticipated frameworks, and that most but not all pulse oximeters would show more positive bias in people with dark vs. light pigmentation.

## Methods

This study was conducted at the University of California San Francisco (UCSF) Hypoxia Laboratory from 2022 to 2024 with UCSF IRB approval (#21-35637, ClinicalTrials.gov ID: NCT06142019). Written informed consent was obtained from all participants.

### Oximeter selection

We tested 34 pulse oximeters. Devices were chosen based on popularity in online marketplaces, clinician input from diverse settings, and global health donor procurement lists to capture commonly used devices across varied clinical settings and price points. Purchase prices varied from $10 to $5,999. A list of tested devices, including prices, form factors, wavelengths, and regulatory data is available in Table S4. For three devices (the Shenzhen PC-60NW, Masimo MightySat, and Acare AH-M1/MX), we purchased two of the same model and reported results by year of device purchase. For two additional devices (Masimo Rad97 and Masimo Rad G), we tested each with two different probes and reported each device-probe combination separately. All participants were monitored throughout the protocol with the lab’s ‘clinical monitor’ oximeter (Nellcor PM1000N, Medtronic, USA).

### Study demographics and skin pigment assessment

Participants were healthy adults 18–50 years old who were non-smoking, with no history of lung, cardiovascular, kidney, or liver disease and without hemoglobinopathy, anemia, clotting disorders, or Raynaud’s disease. Our enrollment targeted diversity of both skin pigment and sex assigned at birth.

Participants’ age, sex, and US National Institutes of Health (NIH) race were self-reported. Participants’ height, weight, and finger diameter were measured. Percent modulation of infrared light (an indicator of participant perfusion) was recorded from the clinical monitor and divided by 10 to approximate comparability with Masimo Perfusion Index.^8^

Our approach to skin pigment assessment is provided in the Supplemental Digital Content eMethods. Briefly, two research coordinators assigned subjective skin pigmentation data for each participant using the Monk Skin Tone (MST) scale.^17,18^ Coordinators also used the Konica Minolta CM 700-d (KM) spectrophotometer to derive individual typology angle (ITA), a frequently used surrogate for melanin^19,20^ at the study participant’s dorsal distal phalanx (DP) (the site for study oximeters) and forehead (the recommended site for using MST).^19^

We categorized participants into light, medium or dark bins based on previously proposed cutoffs: ‘light’ (MST 1–4 and ITA of > 30°), ‘medium’ (MST 5–7 and ITA between 30° and -30°), ‘dark’ (MST 8–10 and ITA of < -30° with ≥ 50% of these participants having ITA < -50°).^12,21,22^ We refer to participants with both MST 8–10 and ITA of < -50° as ‘very dark.’ Unless otherwise noted, we used the ‘dark’ definition above (not ‘very dark’) for analyses. In cases of discordance between ITA and MST, the participant was binned by ITA.

### Controlled desaturation protocol

Each participant underwent a controlled desaturation protocol as previously described and available online and in the Supplemental Digital Content eMethods. ^23–25^ Briefly, study investigators controlled partial pressures of inspired oxygen, carbon dioxide, and nitrogen to achieve six stable “plateaus” of targeted arterial functional oxygen saturation (SaO_2_) between SaO2 ∼70% and 100% (i.e. 2 plateaus in each decile 70-80%, 80-90%, and 90-100%). At each plateau, multiple arterial blood samples were collected ≥ 20 seconds apart from a radial artery catheter. At the time of each blood sample, pulse oximeter SpO_2_’s were recorded, and blood samples were immediately analyzed for SaO_2_ using Radiometer ABL90 Flex Plus (ABL) (Radiometer, Copenhagen, Denmark) blood gas analyzers. Of note, participants’ hands were not warmed by default, and most probes were randomly placed on all five finger digits (see the Supplemental Digital Content eMethods).

### Sample size and cohort composition

The cohort size and composition for the primary analysis were based on what had been shared publicly by the FDA and ISO at the time of study design.^13–16,26^ These criteria were: 1. ≥ 24 unique participants; 2. Each participant must contribute 16 to 30 data points; 3. Pooled SaO_2_ data must span at least 73% to 97% SaO_2_; 4. ≥ 90% of participants must provide ≥ 1 data point < 85%; 5. ≥ 69% of participants must provide ≥ 1 data point in the 70–80% decile; 6. The cohort must include ≥ 33% of each sex; 7. Using forehead skin data, ≥ 25% of participants must fall into each color bin (i.e. light, medium, or dark). Given uncertainties in anticipated recommendations for cohort size, we tested all devices in at least 24 participants and continued testing with as many participants as possible based on available resources.

### Statistical and sensitivity analyses

The primary analysis used 24 participant cohorts and skin pigment data from the forehead and dorsal distal phalanx (DP). We used two metrics to define a ‘passing device’ based on what was shared publicly by FDA and ISO at the time of study design (Table 2): 1. accuracy root mean square error (A_RMS_); and 2. differential bias. A_RMS_ is a performance threshold that measures the square root of the mean of the squared differences in SpO_2_ minus SaO_2_. Differential bias is defined in two ways: 1. For ITA, as the difference in SpO_2_ bias between two theoretical participants with an ITA difference of 100° (i.e., one participant with dark ITA -50° and another with light ITA 50°); and 2. For MST, as the difference between MST bins. We estimated the differential bias and 95% confidence interval (CI) using a linear mixed effects (LME) model. The anticipated ISO standard recommended differential bias thresholds (point estimate ≤ 4% for 70–85% SaO_2_ and ≤ 2% for 85–100% SaO_2_) for only ITA, while the anticipated FDA guidance recommended thresholds (95% CI < 3.5% for 70–85% SaO_2_ and < 1.5% for 85–100% SaO_2_) for both ITA and pairwise comparisons of MST bins 1–4, 5–7, and 8–10.^12,15,16^

**Table 1.** Baseline characteristics of participants.

| Covariates | Overall |
| --- | --- |
| Demographics |  |
| Age (years) | 25 [24,29] |
| BMI (kg/m<sup>2</sup>) | 23 [21,26] |
| Female | 80 (52%) |
| Male | 75 (48%) |
| Finger Diameter (mm) | 10 [9,12] |
| Percent Modulation | 2 [1,5] |
| NIH Race |  |
| Asian | 40 (26%) |
| Black/African American | 36 (23%) |
| Hispanic/Latino | 16 (10%) |
| White | 46 (30%) |
| Other/Multiple | 17 (11%) |
| MST (Forehead) |  |
| 1 | 5 (1%) |
| 2 | 41 (12%) |
| 3 | 17 (5%) |
| 4 | 57 (17%) |
| 5 | 74 (22%) |
| 6 | 37 (11%) |
| 7 | 20 (6%) |
| 8 | 65 (19%) |
| 9 | 21 (6%) |
| 10 | 4 (1%) |

| ITA |  |
| --- | --- |
| ITA (Forehead) | 18 [-19,33] |
| ITA (DP) | 11 [-27,27] |
| ITA Bin (Forehead) |  |
| ITA >30° | 111 (32%) |
| -30 <= ITA <= 30° | 158 (45%) |
| ITA <-30° | 79 (23%) |
| ITA <-50° | 42 (12%) |
| Categorical variables are presented as N (%), while continuous variables are summarized as median [Q1, Q3]. |  |
| BMI = Body Mass Index; MST = Monk Skin Tone Scale; ITA = Individual Typology Angle; NIH = National Institutes of Health;<br>DP = Dorsal Distal Phalanx |  |

This table presents the descriptive statistics for demographics and skin color data. Data are presented as median [Q1, Q3] for continuous variables and count (%) for categorical variables. Race was classified according to National Institutes of Health (NIH) categories. Skin tone was assessed using the Monk Skin Tone Scale (MST) and the Individual Typology Angle (ITA) at two anatomical sites: forehead and dorsal distal phalanx (DP), with repeated measures taken in participants who completed more than one study visit. Note that participants with ITA <-50° are a subset of participants with ITA <-30°.

**Table 2.** Summary of current and anticipated regulatory frameworks for pulse oximetry.

|  | ISO (2017) | Anticipated ISO | FDA (2013) | Anticipated FDA |
| --- | --- | --- | --- | --- |
| # participants for verification study cohorts | ≥10 | ≥ 24 | ≥10 | 150 |
| Diversity of biological sex |  | - ≥33% of each biological sex |  | In each MST group, at least 40% of participants are male, and at least 40% of participants are female |
| Diversity of skin pigment | Skin color should be described* | **MST at forehead:** ≥25% MST 1-3 (light), 4-7 (medium), 8-10 (dark)<br>**ITA at forehead:** ≥ 25% ITA > 30° (light), 30° ≥ ITA ≥ -30° (medium), < -30° (dark)<br>AND at least 50% of the participants in dark category have an ITA ≤ -50° (If a subject has ITA and MST that do not place this subject in the same light, medium, dark category, use the ITA categorization) | 2 of 10, or 15% of total should have dark skin* | **MST at forehead:** ≥25% MST 1-4, 5-7, 8-10<br>**ITA at the forehead:** at least 50% of the participants in MST group 8-10 have an ITA ≤ -50° at the forehead |
| **ARMS criteria between 70-100% SaO<sub>2</sub>** | ARMS <4%<br>"upper 95 % and lower 95 % limits of agreement shall at a minimum be provided" | Arms ≤ 3% with 95% CI disclosed | Arms ≤ 3% transmittance<br>Arms ≤ 3.5% reflectance | Arms < 3% including 95% CI for both transmittance and reflectance devices |
| **Statistical assessment of bias due to skin color** | None | For SaO<sub>2</sub> ranges 70-85% and 85-100%:<br>- Using linear regressions, estimate differential bias between values at an ITA of -50° and +50° at the sensor emitter site<br>- Absolute differential bias ≤ 2% for 85-100% SaO<sub>2</sub>, and ≤ 4% for 70-85% SaO<sub>2</sub> | None | For SaO<sub>2</sub> ranges 70-85% and 85-100%:<br>- Absolute difference in SpO<sub>2</sub> bias across ITA at the sensor emitter site<br>AND forehead MST levels < 1.5% for SaO<sub>2</sub> > 85% and < 3.5% for SaO<sub>2</sub> 70-85% |

**\*No specific recommendations provided on how skin color should be measured or described**

We assessed for bias related to pigment by calculating A_RMS_ and error (SpO_2_ minus SaO_2_) for each pigment bin (forehead and DP).^12^ Error and A_RMS_ were summarized as means with 95% CIs. The 95% CIs were determined using bootstrapping (random resampling with replacement) with 1,000 repetitions. Error across skin color bins was compared using an LME model, which included a random intercept to account for repeated measures within participants. A_RMS_ was computed for each participant and compared between groups of participants based on pigment binning using a Welch’s two-sample t-test. A two-sided p-value < 0.05 was considered significant. P-values are reported unadjusted for multiple comparisons in accordance with FDA guidance for independent pairwise analyses, which does not mandate multiplicity adjustments.

We conducted a secondary analysis for each device using the largest cohorts possible that still met cohort diversity criteria. Additional secondary analyses included the impact on device ‘passing’ of different anatomical pigment assessment sites, different definitions of pigment binning, and different statistical methods for estimating bias, including use of a linear regression (LR) model for differential bias instead of the LME model. The LR model regressed the participant-level bias on the ITA values and estimated the difference in predicted bias between ITA values of -50° and 50° based on the fitted model.

A Monte Carlo simulation explored if participant selection bias could influence ‘passing’ status. For each device, Monte Carlo simulation created 100 cohorts of 24 participants, with at least six darkly pigmented participants. If a device always passed or always failed in all simulated cohorts, it was determined to be less susceptible to selection bias (i.e. ‘cherrypicking’ participants).

The SpO_2_ from most study oximeters was recorded manually into a Research Electronic Data Capture (REDCap) Database at the time of each arterial blood sample, except for the Nellcor PM1000N and Masimo Rad97 which streamed data directly into Labview (National Instruments, Austin, TX). Most oximeters used a self-contained interface that displayed SpO_2_ data on the oximeter itself and could not directly stream data from the oximeter. All other physiologic data were collected into Labview, and all Labview data were merged into the REDCap Database. All analyses were performed with R 4.3.2 (R Core Team, 2023) and Python v3.9.6 (Python Software Foundation, 2024). De-identified data for this study are openly accessible through the Open Oximetry Data Repository via PhysioNet and accessible via the OpenOximetry.org website.^27^

## Results

We completed 348 desaturation studies in 155 participants (30,174 paired SpO_2_:SaO_2_ samples). Each device was tested in ≥ 24 unique participants (median 38, range 24–120) with > 540 SpO_2_:SaO_2_ pairs approximately equally distributed across SaO_2_ deciles, and with > 25% of data points in each decile coming from participants in each skin color bin (Table S3). Demographics are reported in Table 1.

### Overall oximeter performance

For cohorts of 24 participants, device A_RMS_ ranged from 1.69 to 8.04 (Figure 1). Of the 12 devices with A_RMS_ > 3% (i.e. 2013 FDA Guidance threshold), eight had 510(k) premarket clearances. All six devices with an A_RMS_ > 4% (i.e. 2017 ISO Standard threshold) reported a European Conformity (CE) marking (Figure 1 and Table S4).

**Figure 1.**
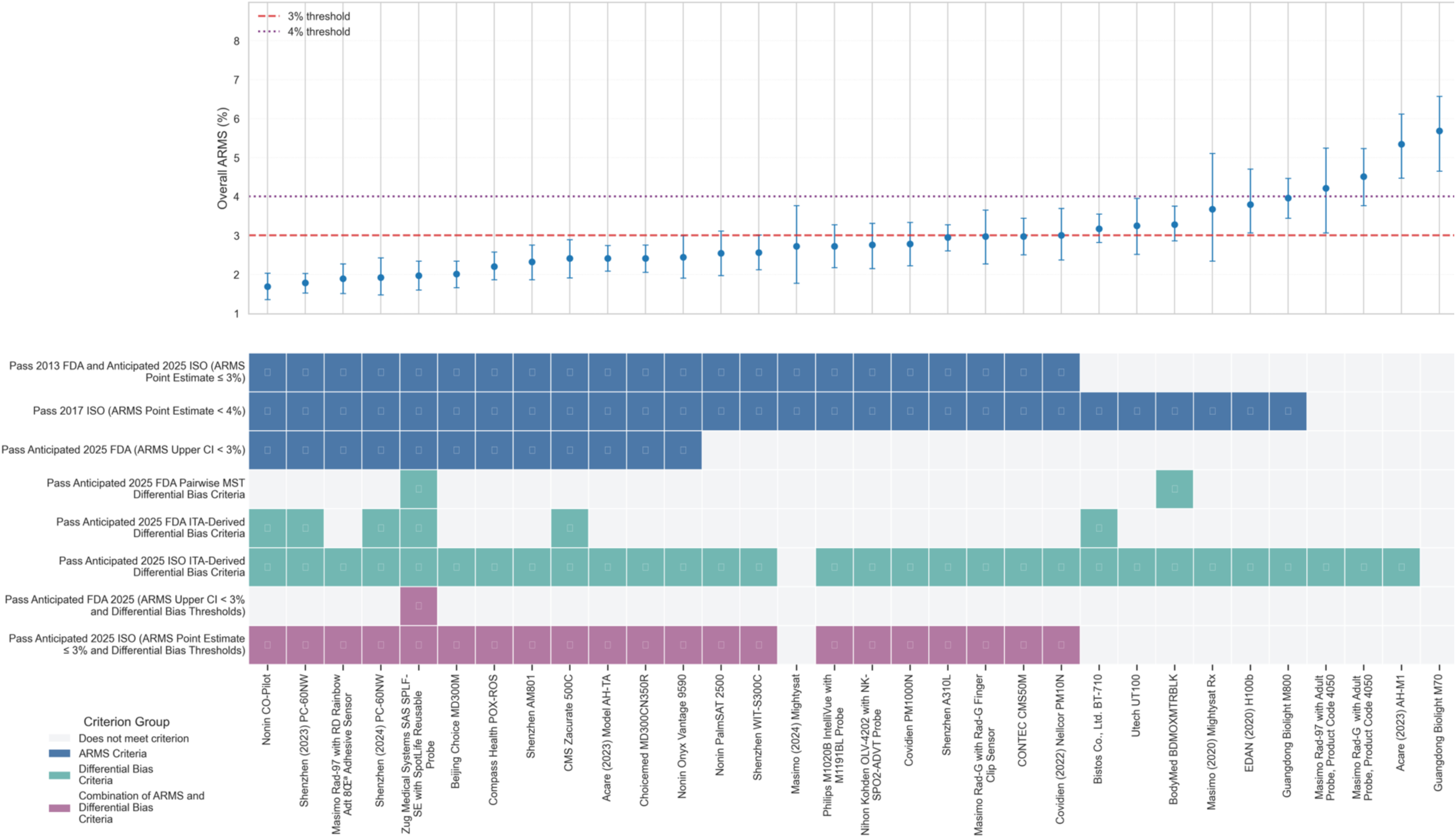
Device conformity with current and anticipated FDA and ISO performance thresholds for 24 participant cohorts and DP ITA. This figure displays devices from lowest to highest accuracy root mean square error (A_RMS_) with 95% confidence intervals (CI) for each device tested in a cohort of 24 participants. The boxes marked with colors indicate that the device meets the A_RMS_ or differential bias thresholds (differential bias and 95% CI < 3.5% for 70–85% SaO_2_ and < 1.5% for 85–100% SaO_2_ for 2025 FDA, and differential bias point estimate ≤ 4% for 70–85% SaO_2_ and ≤ 2% for 85–100% SaO_2_ for 2025 ISO). Differential bias was calculated with the Monk Skin Tone (MST) Scale and individual typology angle (ITA) as indicated. The 95% CI were determined using bootstrapping (random resampling with replacement) with 1,000 repetitions.

Device SpO_2_ bias ranged from -5.71 to 3.48 with a median of 0.97 (IQR: -0.04 – 1.67) (Table S2). We found 24 devices with overall positive bias, and 10 devices with overall negative bias.

### A_RMS_ by skin color bin

In cohorts of 24, we analyzed A_RMS_ by DP pigmentation across 70–100% SaO_2_ and found four devices with significantly different A_RMS_ between light and dark skin color bins (Table S1 and Figure 2). We found significantly higher A_RMS_ in the dark (DP) skin color bin for two devices at 70–80% SaO_2_, four devices at 80–90% SaO_2_, and two devices at 90–100% SaO_2_. One device showed significantly higher A_RMS_ in the light skin color bin (Table S1).

**Figure 2.**
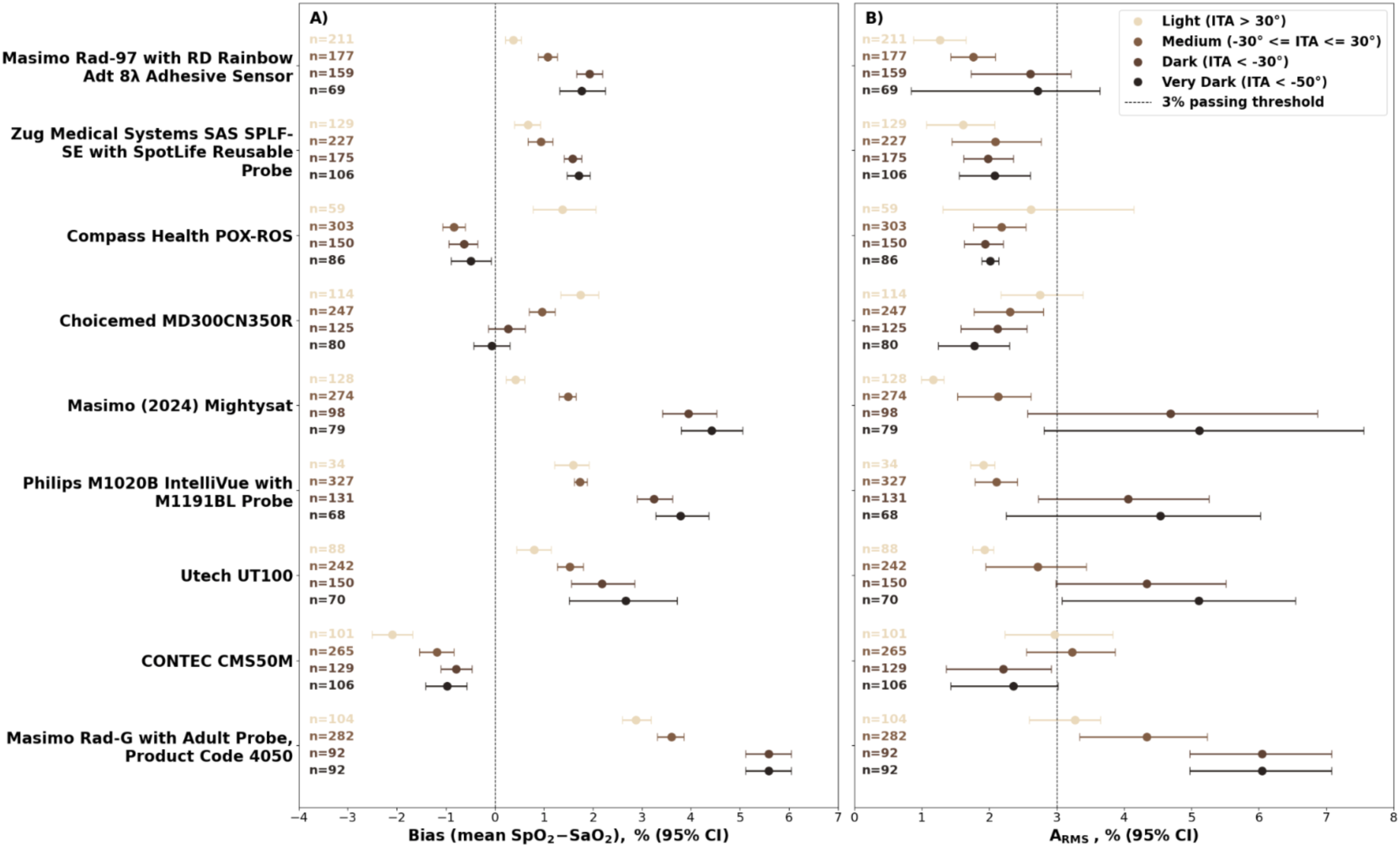
Distribution of bias and A_RMS_ across ITA bins at the DP site over 70-100% saturation range in 24 participant cohorts. This figure shows the distribution of pulse oximeter measured oxygen saturation (SpO₂) bias (mean of SpO₂ minus SaO₂ error, where SaO₂ is functional arterial oxygen saturation) and root mean square error (A_RMS_) in each skin color bin defined by Individual Typology Angle (ITA) measured at the dorsal distal phalanx (DP) site across the saturation range of 70-100%. Devices shown are selected with statistically significantly differences in either bias or A_RMS_ between light (ITA >30°) and dark (ITA <-30°) bins. Panel A displays the distribution of bias in closed circles with 95% confidence interval (CI) whiskers. Panel B shows the distribution of A_RMS_ in closed circles with 95% CI whiskers. Data are stratified into four ITA bins (top to bottom): ITA > 30°, -30° ≤ ITA ≤ 30°, ITA < -30°, and ITA < -50°; note that ITA < -50° is a subset of ITA < -30°. Sample size (n) for each bin is indicated at left in the corresponding color. The vertical dashed line in Panel A represents bias of 0 and in Panel B represents the A_RMS_ 3% threshold.

When analyzing A_RMS_ by forehead pigmentation in cohorts of 24, two devices showed significantly higher A_RMS_ in the dark vs. light color bin at 70–100% SaO_2_ (Table S6), with similar but not exact results to the DP site when broken down by SaO_2_ deciles.

With cohorts larger than 24, more oximeters demonstrated statistically significant A_RMS_ differences between light and dark bins (Table S1, Table S6, Table S8, Table S9). Two devices demonstrated statistically significant differences in A_RMS_ only when comparing light and ‘very dark’ color bins (Table S9).

### Bias by skin color bin

We found five devices with more positive bias in dark vs. light DP skin color bins across SaO_2_ 70–100% (median difference in bias (dark minus light) 1.56, IQR: 1.30 – 2.70), and two devices with more positive bias in the light bin (Table S2 and Figure 2). When analyzing by SaO_2_ deciles, we found significantly more positive bias in dark vs light skin color bins for four devices at 70–80% SaO_2_ (median difference 4.22, IQR: 3.40 – 4.74), four devices at 80–90% SaO_2_ (median difference 2.11 IQR: 1.60 – 2.85) and two devices at 90–100% SaO_2_ (median difference 1.36, range: 1.20 – 1.51). Bias by forehead pigmentation also varied (Table S7).

Analysis of larger cohort sizes revealed more devices (11/34) with significant differences in bias when comparing light vs. dark bins at the DP across SaO_2_ 70–100%. Four devices demonstrated significantly more positive bias when comparing ‘very dark’ vs light color bins, but not dark vs light (Table S10 and Figure 3).

**Figure 3.**
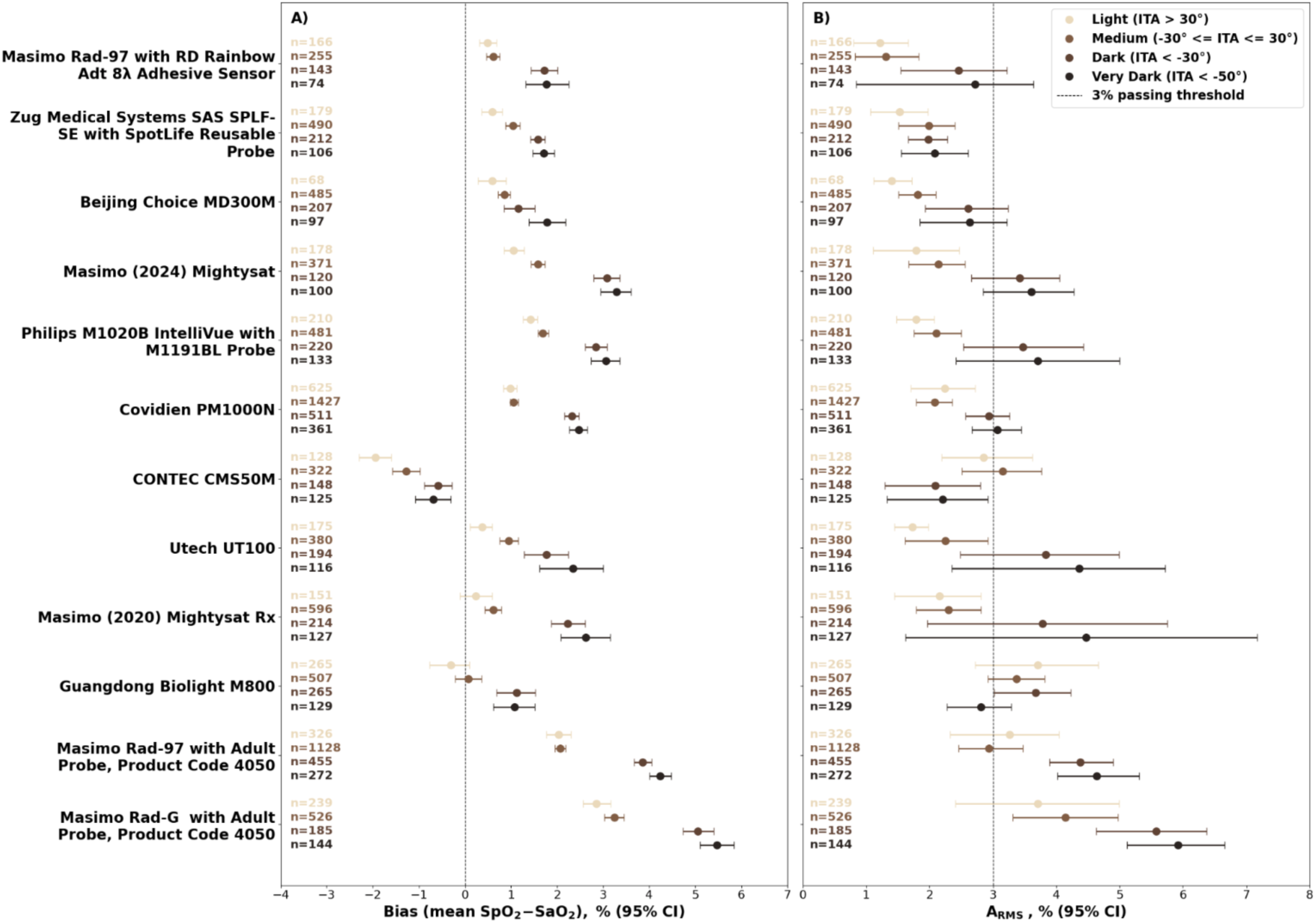
Distribution of bias and A_RMS_ across ITA bins at the DP site over 70-100% saturation range in maximum participant cohorts. This figure shows the distribution of pulse oximeter measured oxygen saturation (SpO₂) bias (mean of SpO₂ minus SaO₂ error, where SaO₂ is functional arterial oxygen saturation) and root mean square error (A_RMS_) in each skin color bin defined by Individual Typology Angle (ITA) measured at the dorsal distal phalanx (DP) site across the saturation range of 70-100%. Devices shown are selected with statistically significantly differences in either bias or A_RMS_ between light (ITA >30°) and dark (ITA <-30°) bins. Panel A displays the distribution of bias in closed circles with 95% confidence interval (CI) whiskers. Panel B shows the distribution of A_RMS_ in closed circles with 95% CI whiskers. Data are stratified into four ITA bins (top to bottom): ITA > 30°, -30° ≤ ITA ≤ 30°, ITA < -30°, and ITA < -50°; note that ITA < -50° is a subset of ITA < -30°. Sample size (n) for each bin is indicated at left in the corresponding color. The vertical dashed line in Panel A represents bias of 0 and in Panel B represents the A_RMS_ 3% threshold.

Figures 4-S13 contain the distribution of A_RMS_ and bias for all devices across all saturation ranges and both anatomical sites.

**Figure 4.**
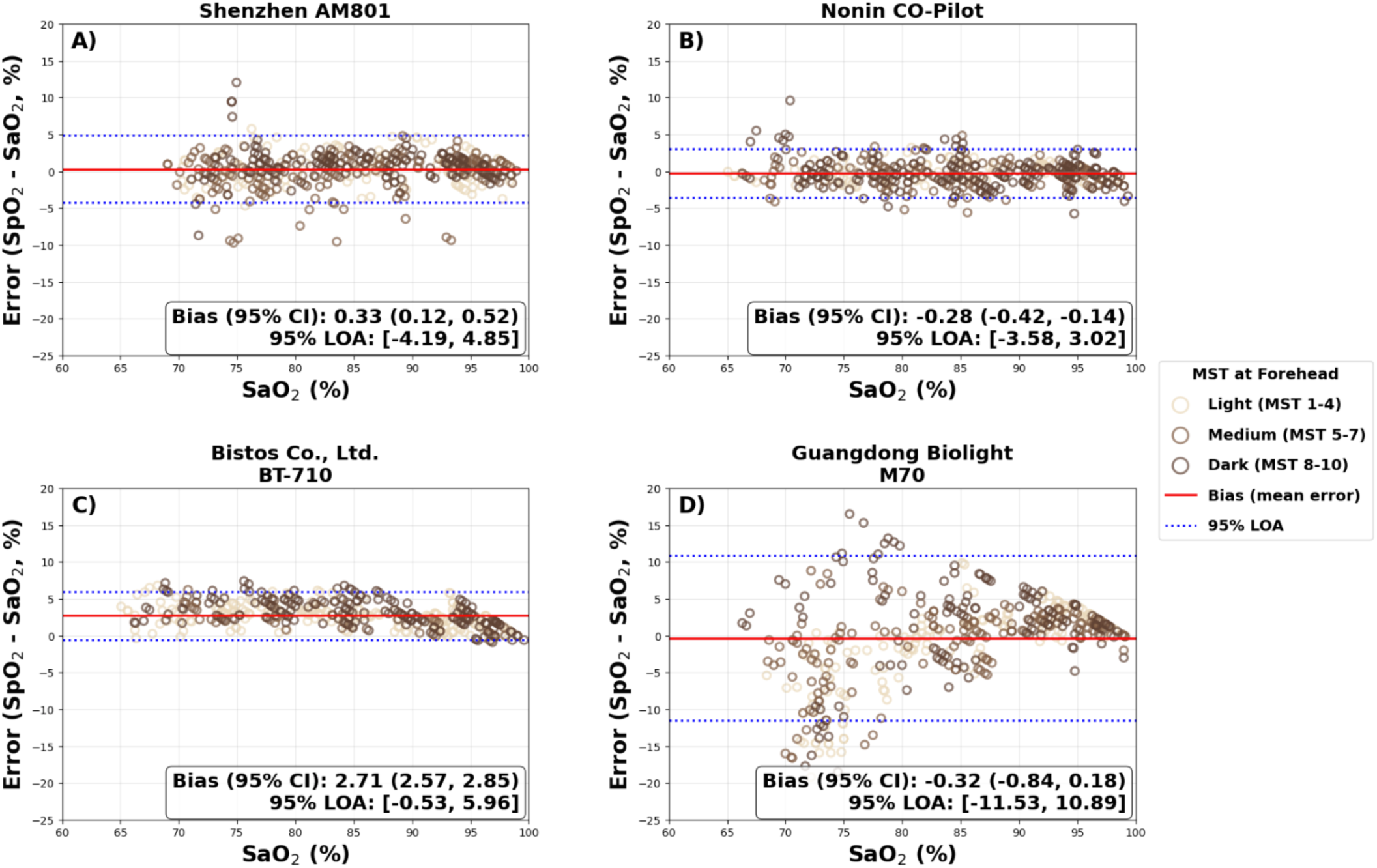
Modified bland altman plots for four selected devices in cohort of 24 participants. This figure shows the modified bland altman (BA) plots for four devices, showing pulse oximeter measured oxygen saturation (SpO₂) bias (mean of SpO₂ minus SaO₂ error, where SaO₂ is functional arterial oxygen saturation) and 95% limits of agreement (LOA). Data points are colored by Monk Skin Tone Scale (MST) at the forehead, as recommended by the U.S. Food and Drug Administration (FDA) draft 510(k) guidance. Panel A: Shenzhen Med-Link Electronics Tech Co., Ltd. AM801 (passing A_RMS_/failing Individual Typology Angle (ITA) differential bias); Panel B: Nonin Medical, Inc. Co-Pilot H500 (passing A_RMS_/passing ITA differential bias); Panel C: Bistos Co., Ltd. BT-710 (failing A_RMS_/passing ITA differential bias); Panel D: Guangdong Biolight Meditech Co., Ltd. M70 (failing A_RMS_/failing ITA differential bias). In each panel, the red solid horizontal line represents the SpO₂ bias; dashed lines are the 95% LOA, both have accounted for repeated measures.

### Differential bias

In cohorts of 24 using DP ITA, differential bias ranged from -1.39 to 9.17 at 70– 85% SaO_2_ (median 1.50, IQR: 0.00 – 2.53) and -1.12 to 2.18 at 85–100% SaO_2_ (median 0.42, IQR: -0.37 – 0.94) (Table S12).

With the larger cohort sizes, DP ITA differential bias ranged from -1.34 to 4.83 at 70–85% SaO_2_ (median 1.36, IQR: 0.01 – 2.33) and from -0.92 to 1.72 at 85–100% SaO_2_ (median of 0.38, IQR: -0.10 – 0.96) (Table S13). Forehead ITA differential bias was similar. The LME and LR models produced similar results.

### Old vs. anticipated regulatory frameworks

With cohorts of 24 participants, 28/34 devices passed 2017 ISO A_RMS_ criteria (≤4%), 22/34 passed 2013 FDA A_RMS_ (≤ 3%) criteria (which is also the anticipated ISO criteria), and 12/34 passed anticipated FDA A_RMS_ criteria (95% UCI < 3%). For differential bias, 1/34 oximeters passed the anticipated FDA criteria, and 32/34 passed the anticipated ISO criteria (Table S12 and Figure 1). Overall, 21/34 oximeters passed both the A_RMS_ and differential bias criteria for anticipated ISO standard, and 1/34 oximeters passed both criteria for anticipated FDA guidance.

With the larger cohort sizes, the number of devices passing both the anticipated A_RMS_ and differential bias criteria increased from 1/34 to 7/34 for FDA, and from 21/34 to 25/34 for ISO (Table S3 and Table S13).

### Impact of pigmentation definitions and measurement sites on bias

The use of race, MST or ITA with varied binning thresholds and different anatomical sites yielded different numbers of devices with significant differences in bias (Table S14). Analysis of light vs dark DP ITA in maximum cohort sizes identified the most devices with significant differences in bias (11), while analysis by race (white vs. Black/African American) identified the fewest.

### Monte Carlo Simulation

We found numerous devices could ‘pass’ or ‘fail’ A_RMS_ criteria depending on which participants were selected for cohort analysis (Figure S3).

## Discussion

We found that many pulse oximeters fail to meet current or anticipated regulatory performance recommendations, and some but not all oximeters had more positive bias in dark vs light participants. Anticipated ISO and FDA regulatory frameworks produced markedly different ‘passing’ rates for the same devices. Most devices passed the anticipated ISO standard, yet only one device passed the anticipated FDA guidance (in cohorts of 24 healthy adults).

Our findings corroborate prior studies demonstrating variable performance and positive bias in some oximeters on the market.^6,7,23,24,28,29^ In several instances, our findings differ from manufacturers’ reports (Figure 1 and Table S4), which may be partly explained by methodological differences. For example, we did not warm most participants’ hands and used all five digits. This was done in an effort to better reflect clinical reality and contrasts with most prior oximeter regulatory verification studies, which typically only used digits 2-4 and actively warmed participants’ hands to increase peripheral perfusion (a factor known to improve device performance and permitted by FDA and ISO).^8^ Additionally, we analyzed consecutively enrolled participants. Our Monte Carlo analysis suggests that had we ‘cherry-picked’ participants for analysis, a practice not explicitly prohibited for regulatory submissions, we could make poorly performing oximeters look better.

Consistent with our hypothesis, we found most (18/34) but not all oximeters exhibited more positive bias (i.e. SpO_2_ overestimating SaO_2_) in participants with dark vs. light skin (Table S10 and Table S11). Unexpectedly, several devices demonstrated more positive bias in participants with light vs. dark skin.

We confirmed our hypothesis that fewer devices pass anticipated regulatory frameworks, but the contrast in differential bias criteria between FDA and ISO is particularly noteworthy. Nearly all devices we tested passed the anticipated differential bias criteria for ISO, but only one passed anticipated FDA criteria (Figure 1). The anticipated ISO standard performed similarly to the 2013 FDA guidance. This is concerning given that devices cleared by 2013 FDA guidance were used in studies demonstrating oximeter performance and health disparity concerns^4,30^ ISO should tighten its recommendations and ensure differential bias thresholds reflect magnitudes of bias (e.g. <2%) seen in clinically relevant SpO2 ranges (e.g. 85-95%).^31–37^The FDA differential bias thresholds necessitate larger study cohorts for some devices to pass. This has generated concern that if studies become too costly or time-consuming, there could be negative global market implications (e.g. decreased device access and increased costs).^13,38^

Increasing the size of verification cohorts (from 10 to 24) and utilizing MST and ITA are good steps toward improving diversity. Our findings suggest that most devices will require cohorts > 24 to pass anticipated FDA criteria, but a cohort size of 150, as proposed in the January 2025 draft FDA guidance, is likely unnecessary. Additionally, we found the anatomical site for skin pigment assessment and definition of “dark” skin pigment also impacted bias, A_RMS_, and regulatory ‘passing’ rates (Table S1 and Table S15).

Without a better understanding of how performance criteria in the lab correlate with performance (or health disparities) in the clinical setting, the optimal regulatory performance thresholds will remain uncertain.

There was no device characteristic (e.g. cost, number of wavelengths, form factor etc.) that was clearly linked to performance, though the study was not designed for this purpose.

This study had several limitations. First, we generally tested only one oximeter per model. For devices with detachable probes, we replaced probes after approximately 24 participants. Thus, we cannot assess variability related to manufacturing differences or wear and tear. For the three oximeter models, we found different performance across manufactured years despite no obvious differences in hardware or software (Table S16, Figure 1, and Table S5). Our clinical monitor (PM1000N) had reasonably consistent performance in these same participant cohorts, suggesting this was not a methodological problem. Further work is needed to analyze variability across different probes of the same model, a potentially important source of error^13^ Second, devices were tested in comparable but not identical cohorts. For example, some cohorts (including those for two devices with the best A_RMS_) had higher participant perfusion (Table S17). Third, several aspects of our desaturation protocol could have influenced findings. For example, not all oximeters are designed for use on the 1st and 5th digits. Plateau stability was partially defined by stability of the clinical monitor, which may not always align with stability of tested oximeters. To mitigate this, we also used stability of calculated saturation (ScO_2_)^23^ based on exhaled CO_2_ and O_2_ partial pressures, to define plateau stability. Another protocol-related limitation is that SaO_2_ can vary by 1-3% across co-oximeter (i.e. blood gas analyzer) brands.^39^ We could not account for this factor because manufacturers generally do not disclose which co-oximeter was used in testing. Finally, there are also limitations in our skin pigment assessment protocol as previously reported.^40^ Although individuals with ITA < -50° represent a critical group for ensuring devices work equitably across all skin tones, we were unable to enroll a sufficient number of participants in this category to meaningfully assess its impact on bias analyses. Generalizability of our findings to the clinical setting, pediatric patients or devices we did not test should be done with caution.

## Conclusion

Clinicians should be aware that many oximeters do not meet current or anticipated regulatory performance recommendations, and some, but not all, devices show more positive bias in people with dark vs light skin pigment. While it may be uncertain to regulators how much differential bias is reasonable to allow, it is clear that some oximeters appear to have minimal or no significant bias related to pigment, and all manufacturers should aspire to this performance goal regardless of the thresholds regulators ultimately propose. Future studies are needed to optimize regulatory frameworks, though many improvements can be made immediately using available data. Despite performance limitations of regulatory-compliant oximeters, clinicians should continue to utilize these essential tools but use great caution when using absolute SpO_2_ cutoffs for providing or withholding treatments.

## Supporting information

eMethods

## Data Availability

All data produced are available online at https://openoximetry.org/data-repository/

## Acknowledgements

We are grateful to members of the Open Oximetry Collaborative Community who participated in the project’s open forum discussions on protocols and approaches to data analysis, including James Ramsay, Jenna Lester, Alex Pogorzelski, Margaret Akey, Daryl Dorsey, Bob Kopotic and Sandy Weininger (ISO/IEC Conveners of the Oximeters Medical Device Standards), and many others.

## Funding Statement

This study was conducted as part of the Open Oximetry Project funded by the Gordon and Betty Moore Foundation, Patrick J McGovern Foundation, PATH/UNITAID, and Robert Wood Johnson Foundation. Dr Ellis Monk’s time utilized for data analysis, reviewing and editing was funded by grant number: DP2MH132941.

## Conflicts of Interest

The UCSF Hypoxia Research Laboratory receives funding from multiple industry sponsors to test the sponsors’ devices for the purposes of product development and regulatory performance testing. This paper does not include data collected for sponsors. All data were collected from devices procured by the Hypoxia Research Laboratory for the purposes of independent research. No company provided any direct funding for this study, participated in study design, or was involved in analyzing data or writing the manuscript. None of the authors own stock or equity interests in any pulse oximeter companies.

## Abbreviations and Acronyms

A_RMS_: Accuracy root mean square error
CI: Confidence interval
UCI: Upper confidence interval
DP: Dorsal distal phalanx
FDA: US Food and Drug Administration
ISO: International Organization for Standardization
ITA: Individual Typology Angle
KM: Konica Minolta
LME: Linear mixed effects (model)
LR: Linear regression (model)
MST: Monk Skin Tone (Scale)
NIH: National Institutes of Health
SaO_2_: Functional arterial hemoglobin oxygen saturation
SpO_2_: Pulse oximeter indirect measure of arterial hemoglobin oxygen saturation
UCSF: University of California San Francisco

## Figures

**Figure S1.**
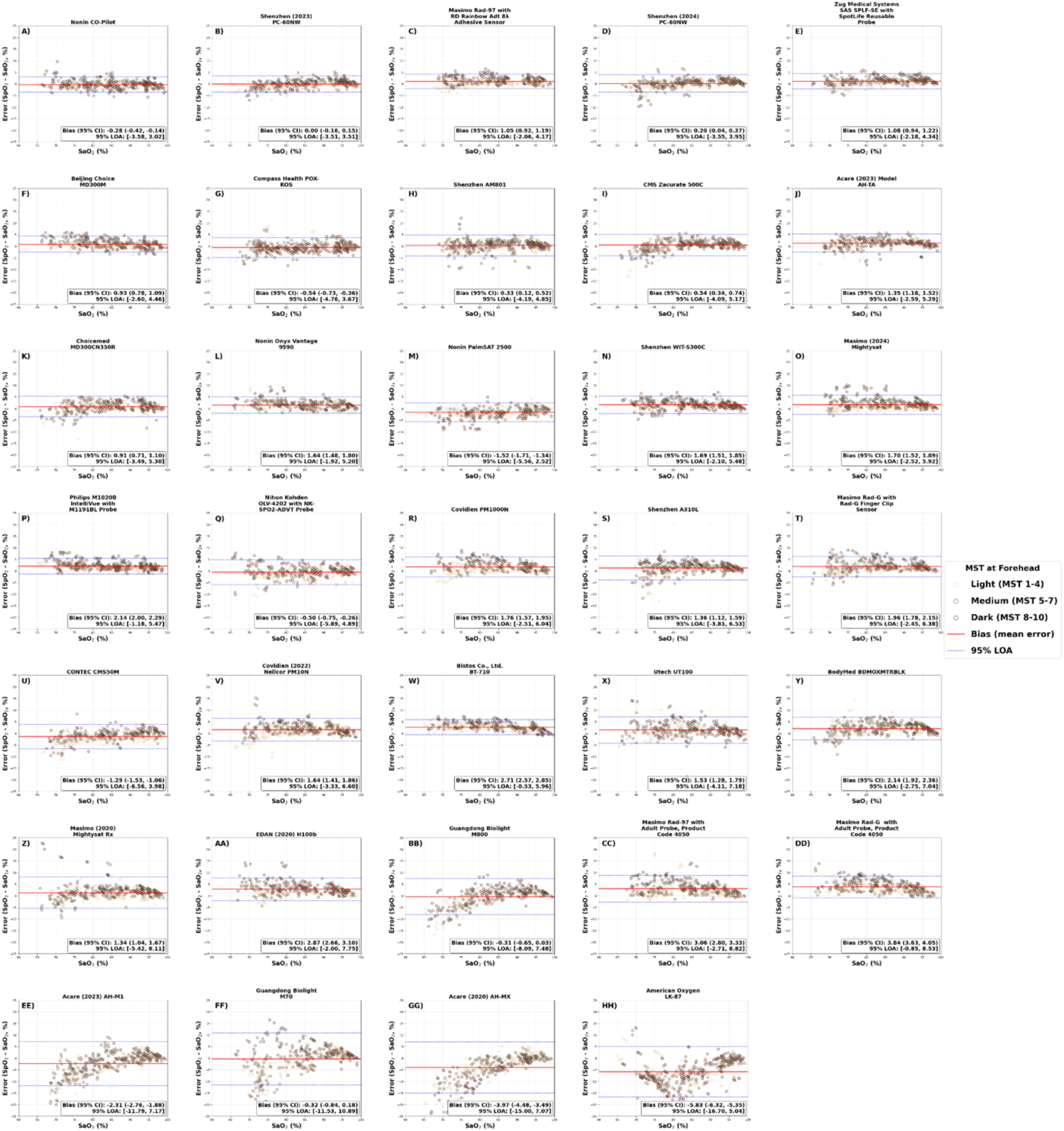
Modified bland altman plots for all 34 devices in cohorts of 24 participants. This figure shows the modified bland altman (BA) plots for all 34 devices, showing pulse oximeter measured oxygen saturation (SpO₂) bias (mean of SpO₂ minus SaO₂ error, where SaO₂ is functional arterial oxygen saturation) and 95% limits of agreement (LOA). Data points are colored by Monk Skin Tone Scale (MST) at the forehead, as recommended by the U.S. Food and Drug Administration (FDA) draft 510(k) guidance. In each panel, the red solid horizontal line represents the SpO₂ bias; dashed lines are the 95% LOA, both have accounted for repeated measures.

**Figure S2.**
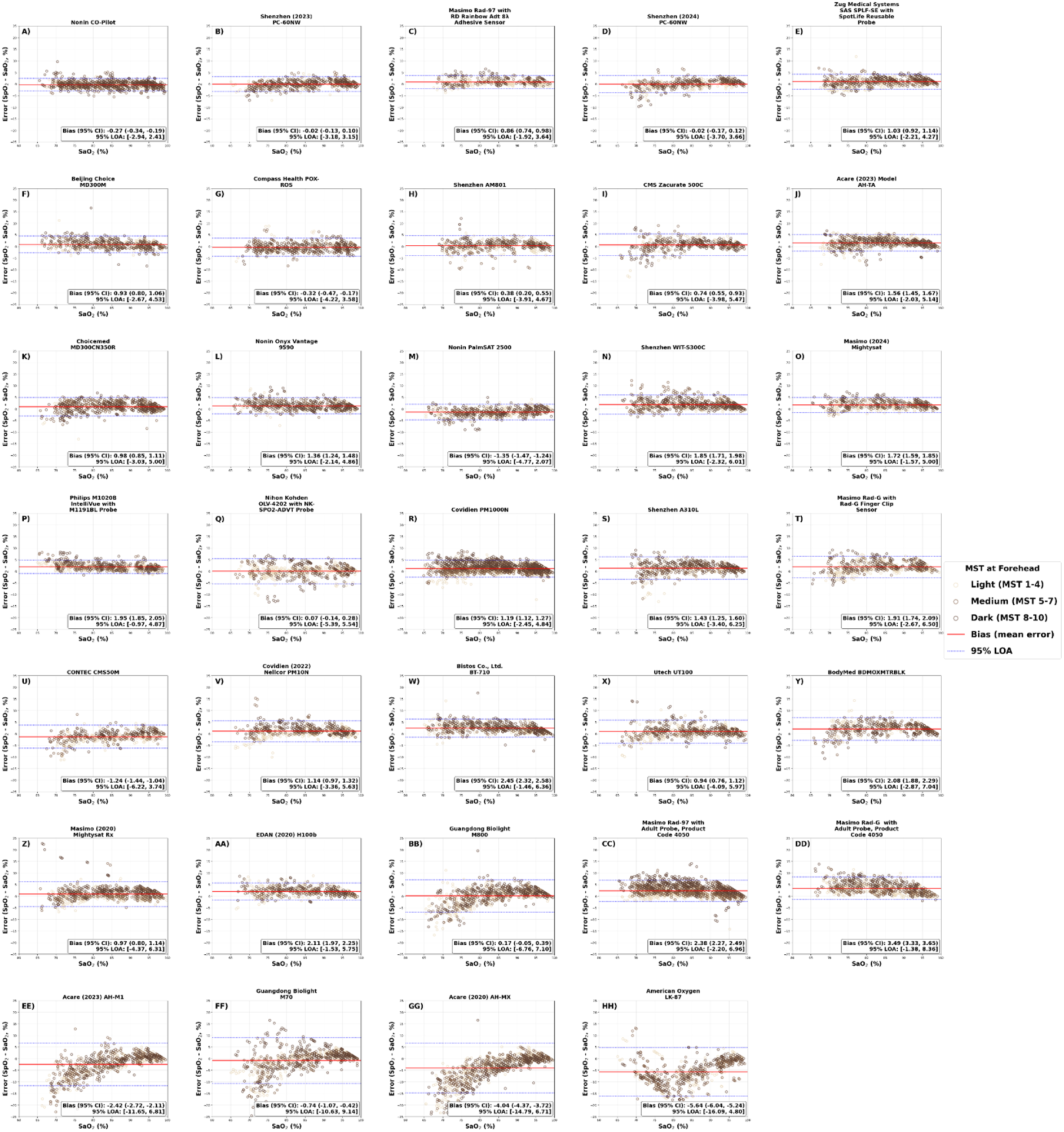
Modified bland altman plots for all 34 devices in maximum participants cohorts. This figure shows the modified bland altman (BA) plots for all 34 devices, showing pulse oximeter measured oxygen saturation (SpO₂) bias (mean of SpO₂ minus SaO₂ error, where SaO₂ is functional arterial oxygen saturation) and 95% limits of agreement (LOA). Data points are colored by Monk Skin Tone Scale (MST) at the forehead, as recommended by the U.S. Food and Drug Administration (FDA) draft 510(k) guidance. In each panel, the red solid horizontal line represents the SpO₂ bias; dashed lines are the 95% LOA, both have accounted for repeated measures.

**Figure S3.**
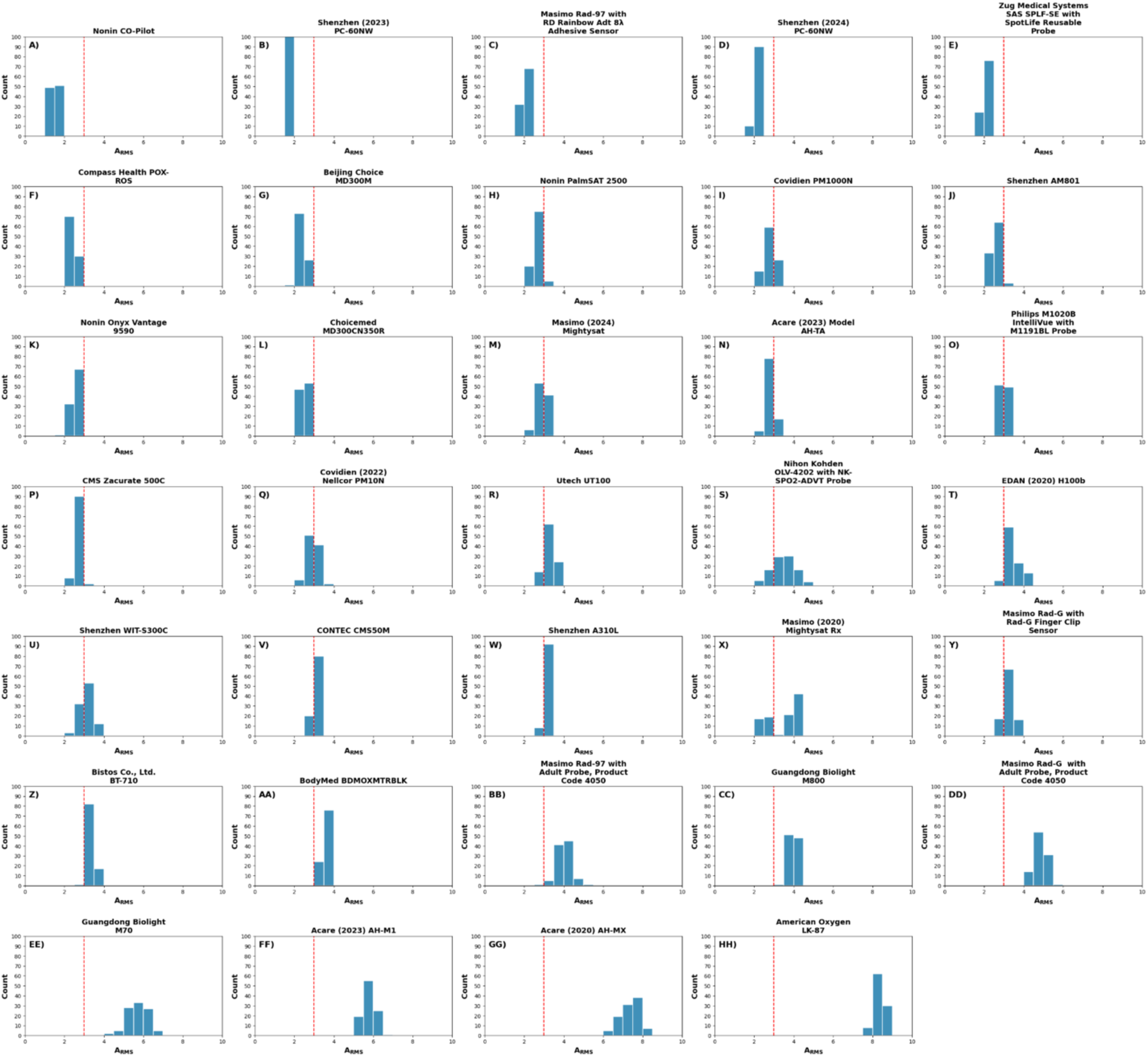
Monte Carlo simulation of A_RMS_ performance in cohorts of 24 participants. For each of 100 simulations, a cohort of 24 unique participants was randomly sampled without replacement and accuracy root mean square error (A_RMS_) was calculated; bootstrapping (random resampling with replacement) with 1,000 repetitions was then used to estimate the upper bound of the 95% confidence interval (CI) of A_RMS_ in that simulated cohort. The histogram displays, for each device, the number of simulated cohorts whose upper 95% CI for A_RMS_ fell below the 3% cutoff (red dashed line). Nine devices achieved an upper 95% CI for A_RMS_ below 3% in all 100 simulations, whereas eighteen devices met this criterion in some simulations but not others. Jaccard diversity scores across devices—quantifying the overlap of cohorts across the 100 simulations— had mean values ranging from 0.03 to 0.67 (median 0.35, interquartile range 0.31– 0.45), indicating adequate cohort diversity across simulations. All devices had at least thirty unique sessions used in the simulations (median 47.5 sessions).

**Figure S4.**
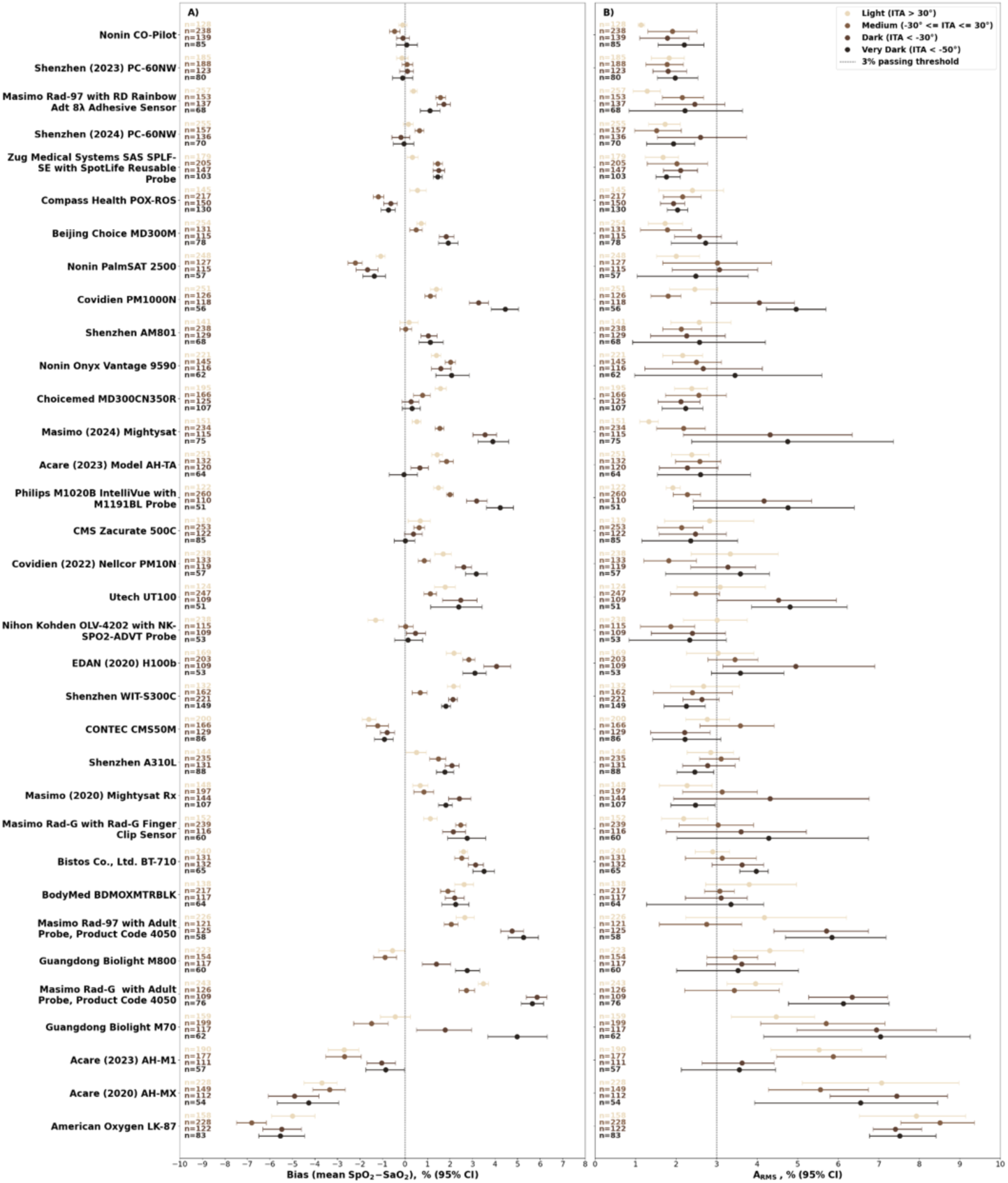
Distribution of bias and A_RMS_ across ITA bins at the forehead over 70–100% saturation range in 24 participants cohorts. This figure shows the distribution of pulse oximeter measured oxygen saturation (SpO₂) bias (mean of SpO₂ minus SaO₂ error, where SaO₂ is functional arterial oxygen saturation) and accuracy root mean square error (A_RMS_) in each skin color bin defined by Individual Typology Angle (ITA) measured at the forehead across the saturation range of 70–100%. Panel A displays the distribution of bias in closed circles with 95% confidence interval (CI) whiskers. Panel B shows the distribution of A_RMS_ in closed circles with 95% CI whiskers. Data are stratified into four ITA bins (top to bottom): ITA > 30°, -30° ≤ ITA ≤ 30°, ITA < -30°, and ITA < -50°; note that ITA < -50° is a subset of ITA < -30°. Sample size (n) for each bin is indicated at left in the corresponding color. The vertical dashed line in Panel A represents bias of 0 and in Panel B represents the A_RMS_ 3% threshold.

**Figure S5.**
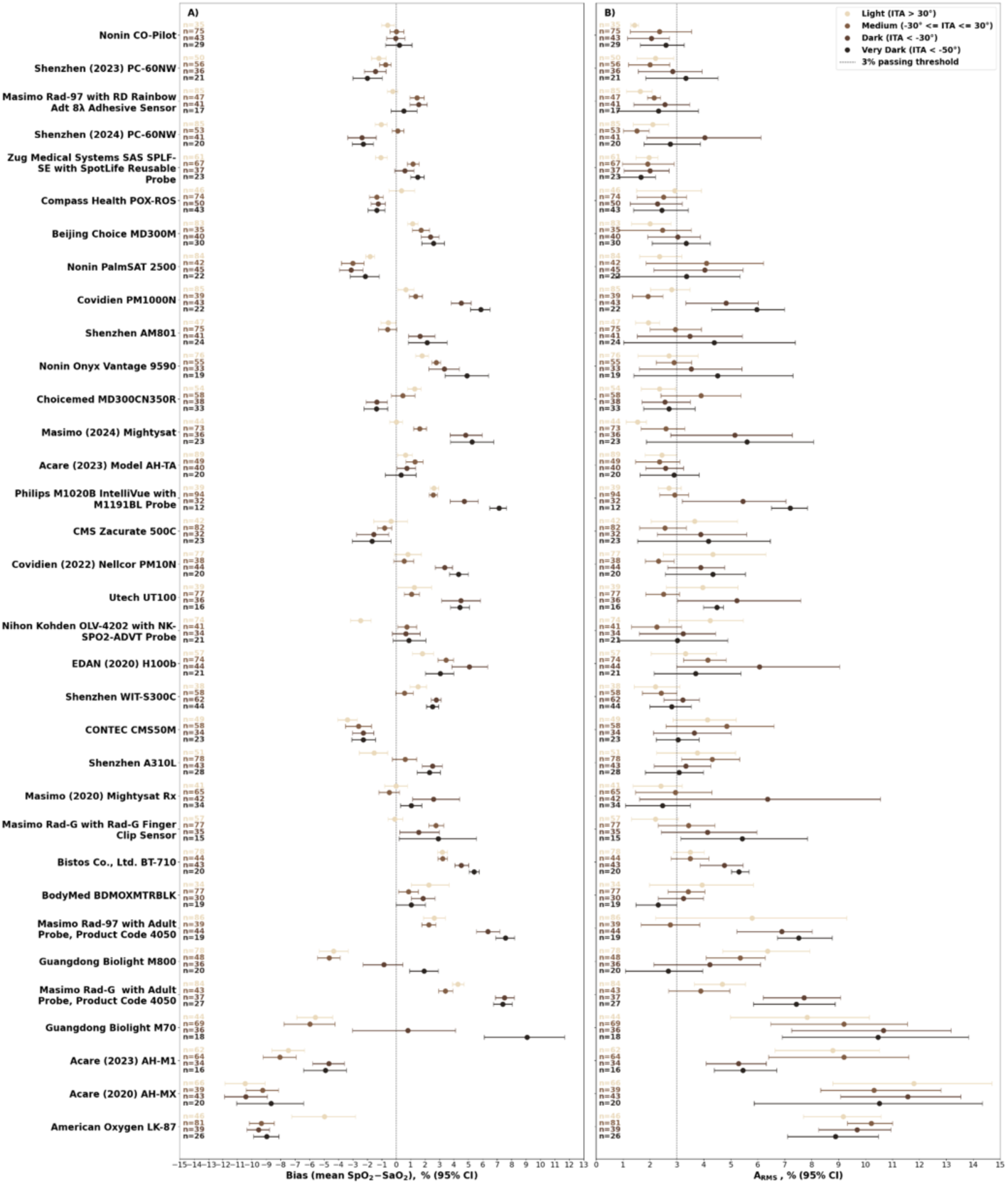
Distribution of bias and A_RMS_ across ITA bins at the forehead over 70–80% saturation range in 24 participants cohorts. This figure shows the distribution of pulse oximeter measured oxygen saturation (SpO₂) bias (mean of SpO₂ minus SaO₂ error, where SaO₂ is functional arterial oxygen saturation) and accuracy root mean square error (A_RMS_) in each skin color group defined by Individual Typology Angle (ITA) measured at the forehead across the saturation range of 70–80%. Panel A displays the distribution of bias in closed circles with 95% confidence interval (CI) whiskers. Panel B shows the distribution of A_RMS_ in closed circles with 95% CI whiskers. Data are stratified into four ITA bins (top to bottom): ITA > 30°, -30° ≤ ITA ≤ 30°, ITA < -30°, and ITA < -50°; note that ITA < -50° is a subset of ITA < -30°. Sample size (n) for each bin is indicated at left in the corresponding color. The vertical dashed line in Panel A represents bias of 0 and in Panel B represents the A_RMS_ 3% threshold.

**Figure S6.**
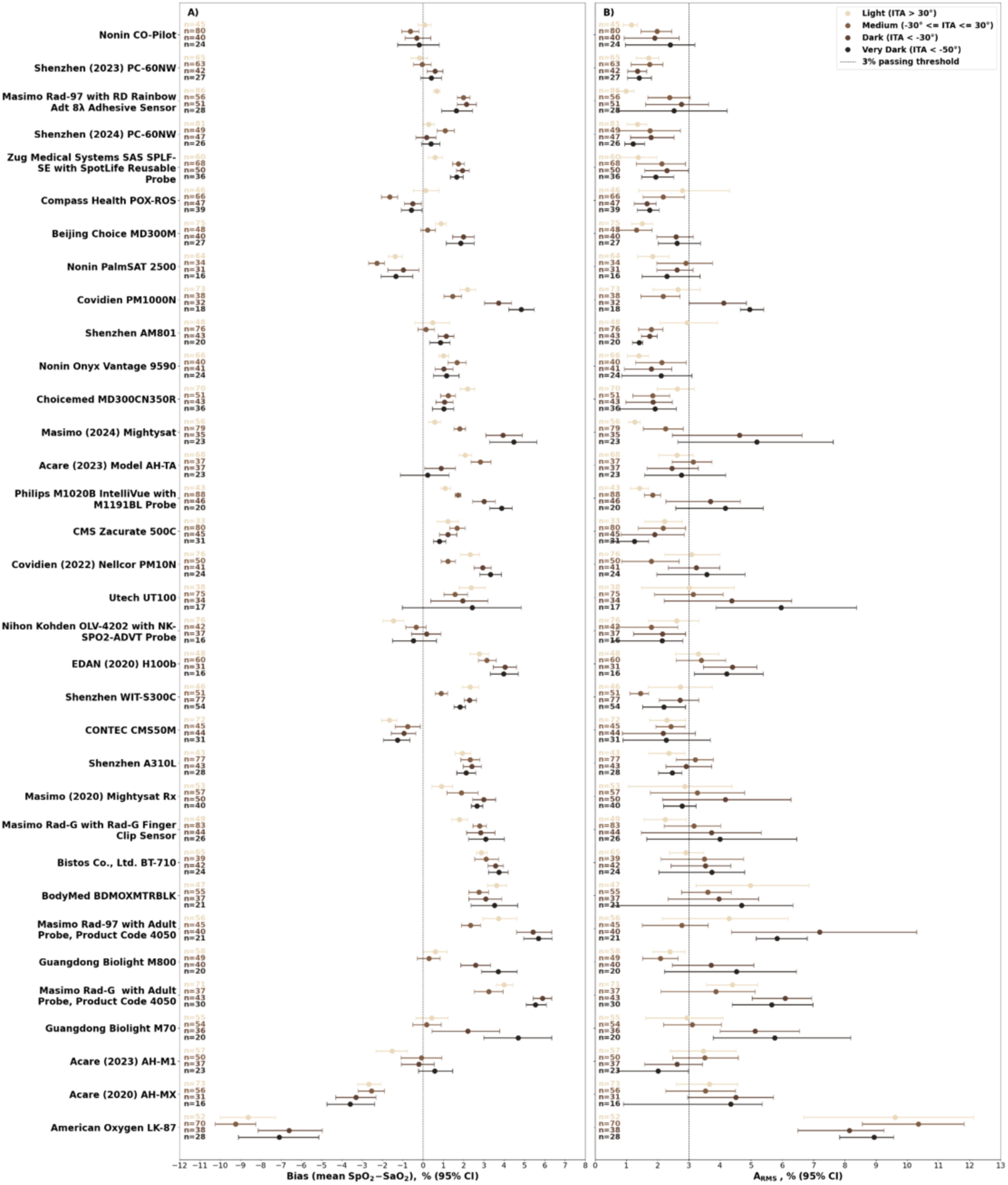
Distribution of bias and A_RMS_ across ITA bins at the forehead over 80–90% saturation range in 24 participants cohorts. This figure shows the distribution of pulse oximeter measured oxygen saturation (SpO₂) bias (mean of SpO₂ minus SaO₂ error, where SaO₂ is functional arterial oxygen saturation) and accuracy root mean square error (A_RMS_) in each skin color bin defined by Individual Typology Angle (ITA) measured at the forehead across the saturation range of 80–90%. Panel A displays the distribution of bias in closed circles with 95% confidence interval (CI) whiskers. Panel B shows the distribution of A_RMS_ in closed circles with 95% CI whiskers. Data are stratified into four ITA bins (top to bottom): ITA > 30°, -30° ≤ ITA ≤ 30°, ITA < -30°, and ITA < -50°; note that ITA < -50° is a subset of ITA < -30°. Sample size (n) for each bin is indicated at left in the corresponding color. The vertical dashed line in Panel A represents bias of 0 and in Panel B represents the A_RMS_ 3% threshold.

**Figure S7.**
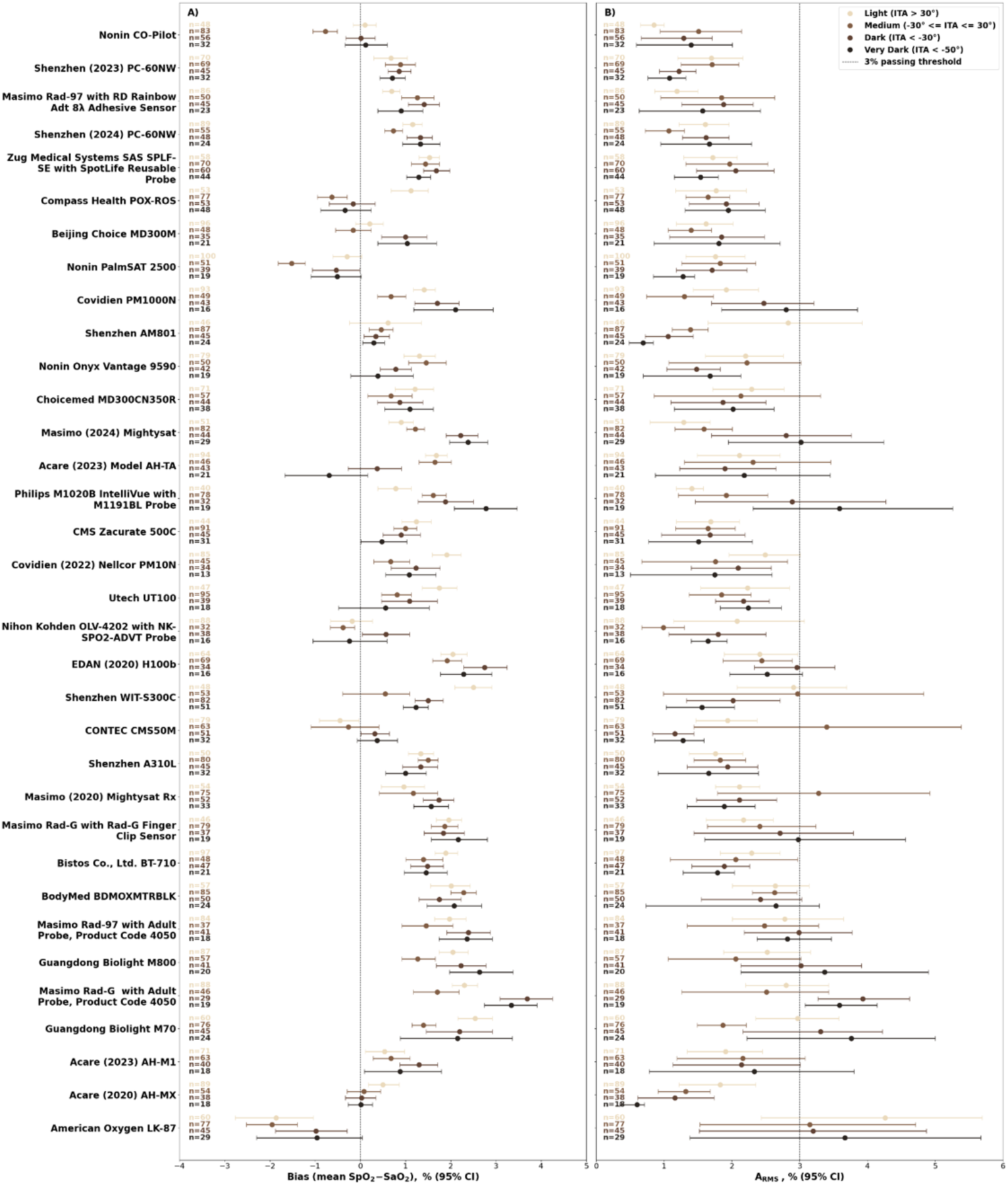
Distribution of bias and A_RMS_ across ITA bins at the forehead over 90–100% saturation range in 24 participants cohorts. This figure shows the distribution of pulse oximeter measured oxygen saturation (SpO₂) bias (mean of SpO₂ minus SaO₂ error, where SaO₂ is functional arterial oxygen saturation) and accuracy root mean square error (A_RMS_) in each skin color bin defined by Individual Typology Angle (ITA) measured at the forehead across the saturation range of 90–100%. Panel A displays the distribution of bias in closed circles with 95% confidence interval (CI) whiskers. Panel B shows the distribution of A_RMS_ in closed circles with 95% CI whiskers. Data are stratified into four ITA bins (top to bottom): ITA > 30°, -30° ≤ ITA ≤ 30°, ITA < -30°, and ITA < -50°; note that ITA < -50° is a subset of ITA < -30°. Sample size (n) for each bin is indicated at left in the corresponding color. The vertical dashed line in Panel A represents bias of 0 and in Panel B represents the A_RMS_ 3% threshold.

**Figure S8.**
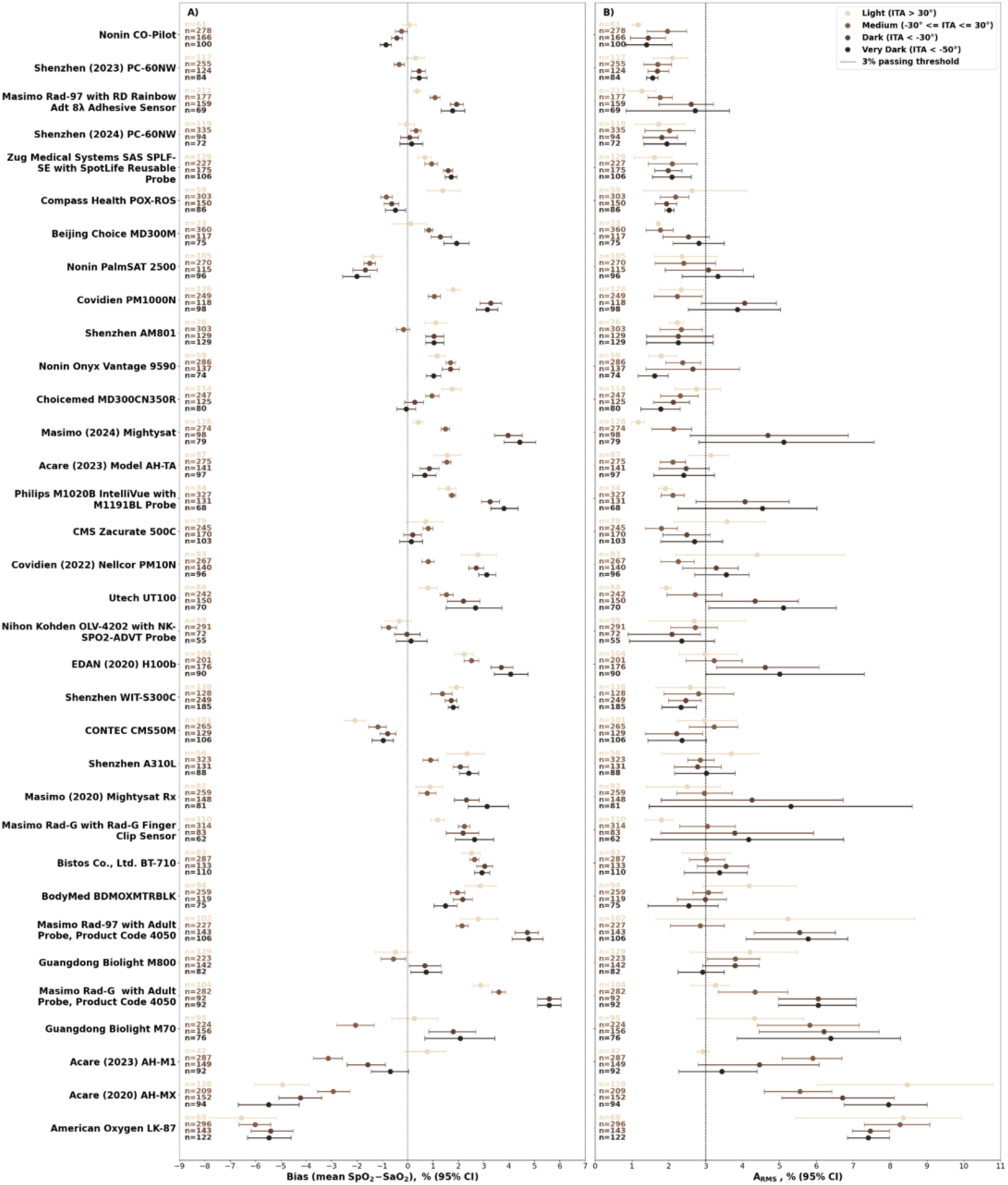
Distribution of bias and A_RMS_ across ITA bins at the DP site over 70–100% saturation range in 24 participants cohorts. This figure shows the distribution of pulse oximeter measured oxygen saturation (SpO₂) bias (mean of SpO₂ minus SaO₂ error, where SaO₂ is functional arterial oxygen saturation) and accuracy root mean square error (A_RMS_) in each skin color bin defined by Individual Typology Angle (ITA) measured at the dorsal distal phalanx (DP) site across the saturation range of 70–100%. Panel A displays the distribution of bias in closed circles with 95% confidence interval (CI) whiskers. Panel B shows the distribution of A_RMS_ in closed circles with 95% CI whiskers. Data are stratified into four ITA bins (top to bottom): ITA > 30°, -30° ≤ ITA ≤ 30°, ITA < -30°, and ITA < -50°; note that ITA < -50° is a subset of ITA < -30°. Sample size (n) for each bin is indicated at left in the corresponding color. The vertical dashed line in Panel A represents bias of 0 and in Panel B represents the A_RMS_ 3% threshold.

**Figure S9.**
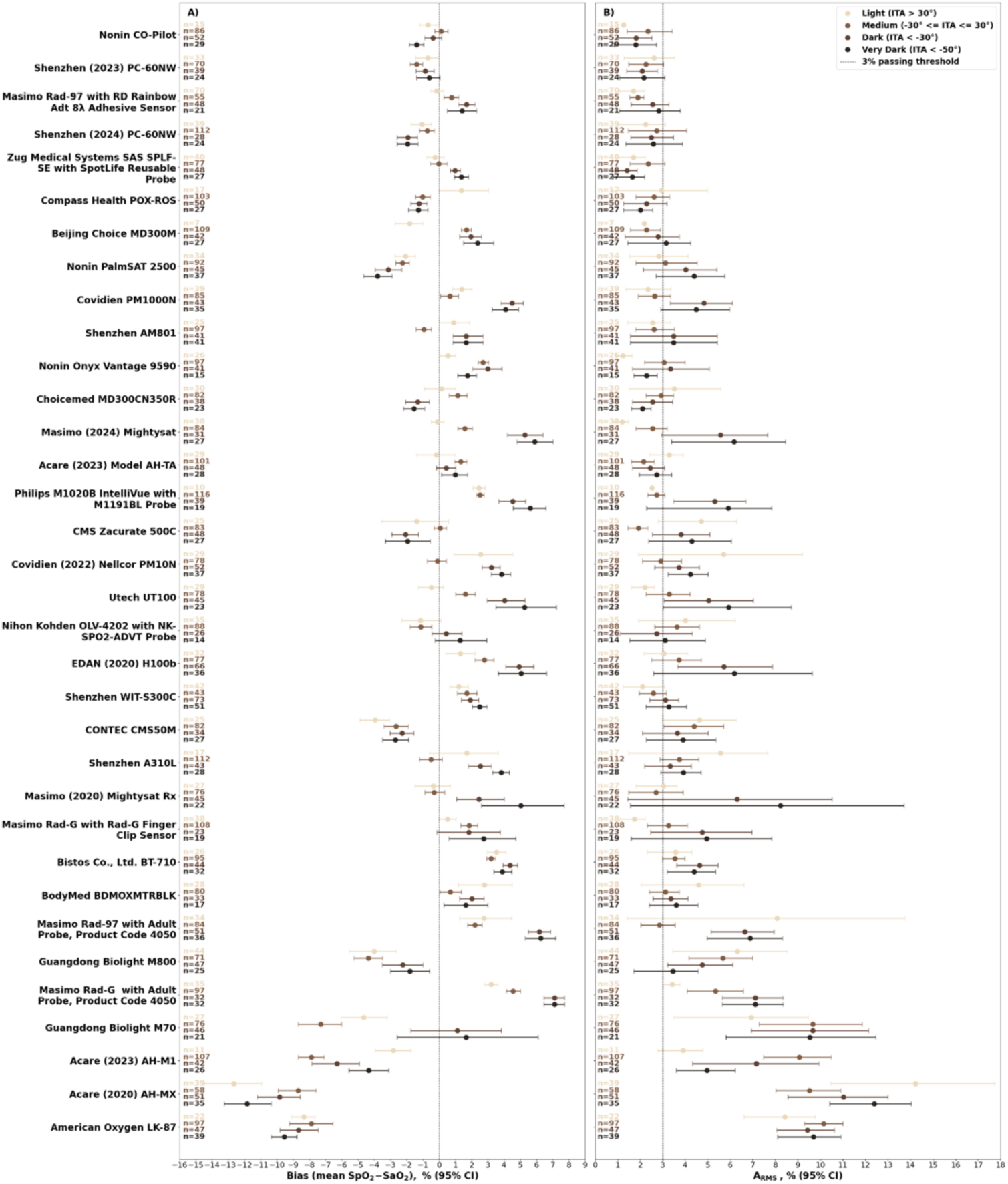
Distribution of bias and A_RMS_ across ITA bins at the DP site over 70–80% saturation range in 24 participants cohorts. This figure shows the distribution of pulse oximeter measured oxygen saturation (SpO₂) bias (mean of SpO₂ minus SaO₂ error, where SaO₂ is functional arterial oxygen saturation) and accuracy root mean square error (A_RMS_) in each skin color bin defined by Individual Typology Angle (ITA) measured at the dorsal distal phalanx (DP) site across the saturation range of 70–80%. Panel A displays the distribution of bias in closed circles with 95% confidence interval (CI) whiskers. Panel B shows the distribution of A_RMS_ in closed circles with 95% CI whiskers. Data are stratified into four ITA bins (top to bottom): ITA > 30°, -30° ≤ ITA ≤ 30°, ITA < -30°, and ITA < -50°; note that ITA < -50° is a subset of ITA < -30°. Sample size (n) for each bin is indicated at left in the corresponding color. The vertical dashed line in Panel A represents bias of 0 and in Panel B represents the A_RMS_ 3% threshold.

**Figure S10.**
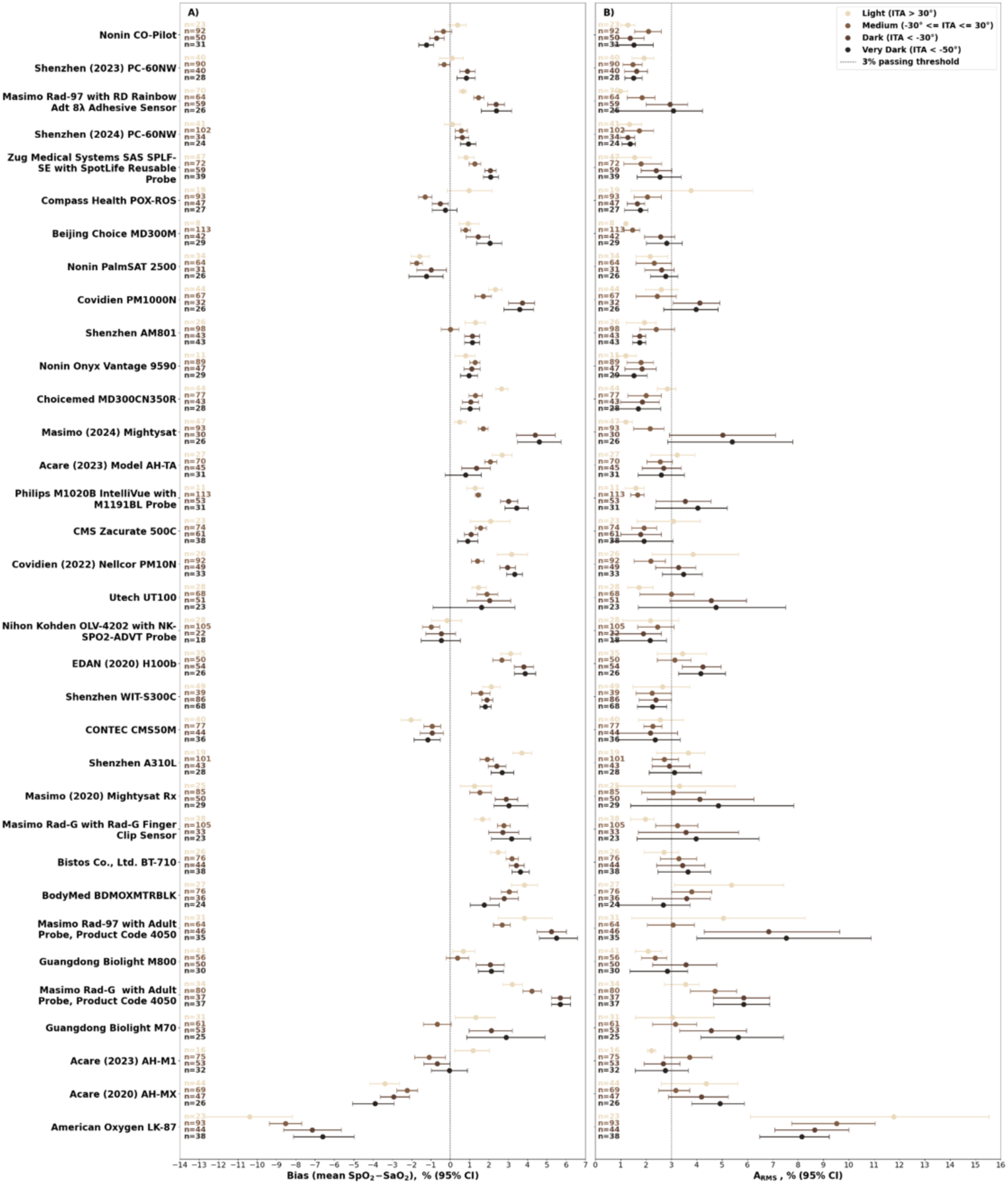
Distribution of bias and A_RMS_ across ITA bins at the DP site over 80–90% saturation range in 24 participants cohorts. This figure shows the distribution of pulse oximeter measured oxygen saturation (SpO₂) bias (mean of SpO₂ minus SaO₂ error, where SaO₂ is functional arterial oxygen saturation) and accuracy root mean square error (A_RMS_) in each skin color bin defined by Individual Typology Angle (ITA) measured at the dorsal distal phalanx (DP) site across the saturation range of 80–90%. Panel A displays the distribution of bias in closed circles with 95% confidence interval (CI) whiskers. Panel B shows the distribution of A_RMS_ in closed circles with 95% CI whiskers. Data are stratified into four ITA bins (top to bottom): ITA > 30°, -30° ≤ ITA ≤ 30°, ITA < -30°, and ITA < -50°; note that ITA < -50° is a subset of ITA < -30°. Sample size (n) for each bin is indicated at left in the corresponding color. The vertical dashed line in Panel A represents bias of 0 and in Panel B represents the A_RMS_ 3% threshold.

**Figure S11.**
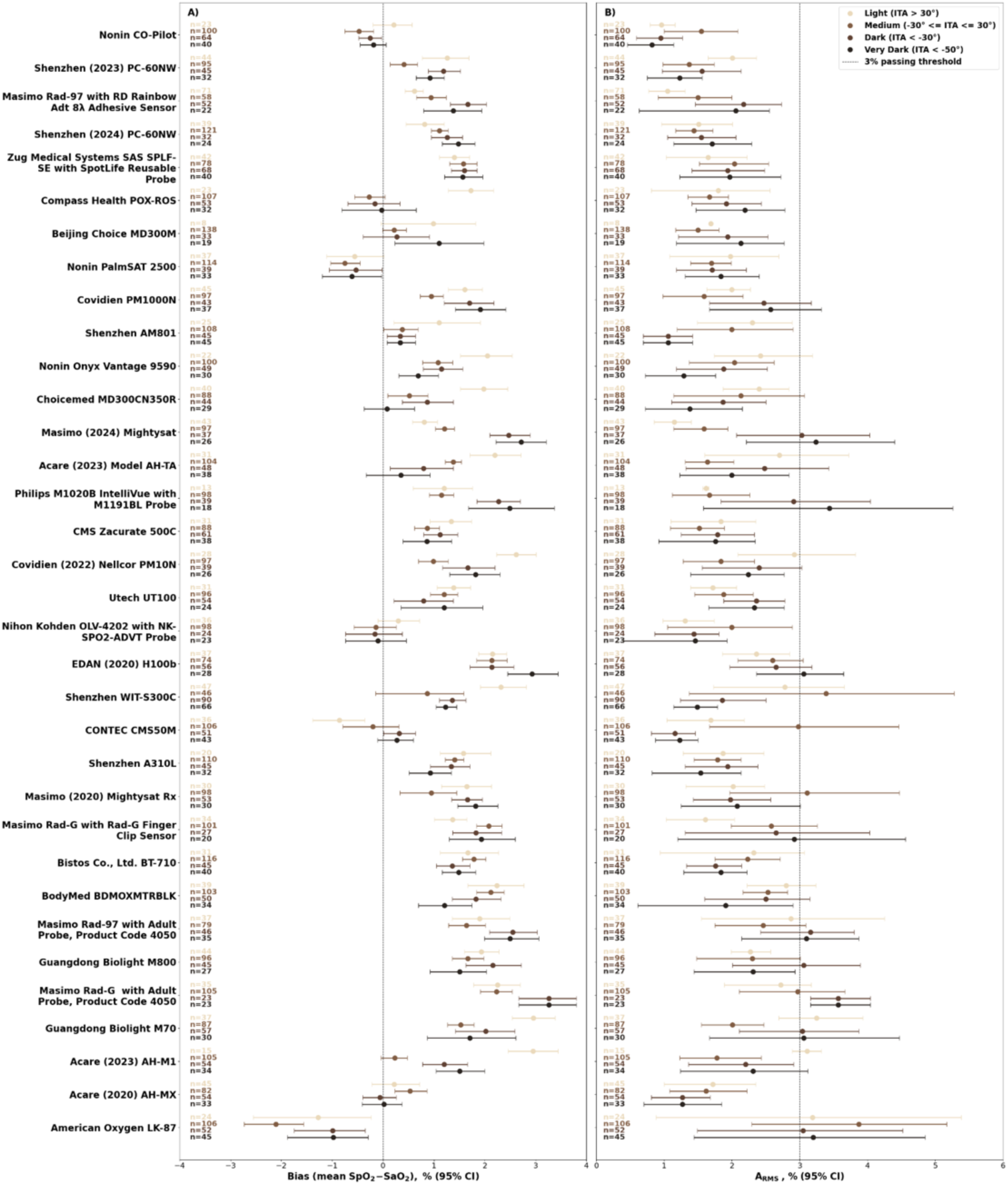
Distribution of bias and A_RMS_ across ITA bins at the DP site over 90–100% saturation range in 24 participants cohorts. This figure shows the distribution of pulse oximeter measured oxygen saturation (SpO₂) bias (mean of SpO₂ minus SaO₂ error, where SaO₂ is functional arterial oxygen saturation) and accuracy root mean square error (A_RMS_) in each skin color bin defined by Individual Typology Angle (ITA) measured at the dorsal distal phalanx (DP) site across the saturation range of 90–100%. Panel A displays the distribution of bias in closed circles with 95% confidence interval (CI) whiskers. Panel B shows the distribution of A_RMS_ in closed circles with 95% CI whiskers. Data are stratified into four ITA bins (top to bottom): ITA > 30°, -30° ≤ ITA ≤ 30°, ITA < -30°, and ITA < -50°; note that ITA < -50° is a subset of ITA < -30°. Sample size (n) for each bin is indicated at left in the corresponding color. The vertical dashed line in Panel A represents bias of 0 and in Panel B represents the A_RMS_ 3% threshold.

## Tables

**Table S1.**
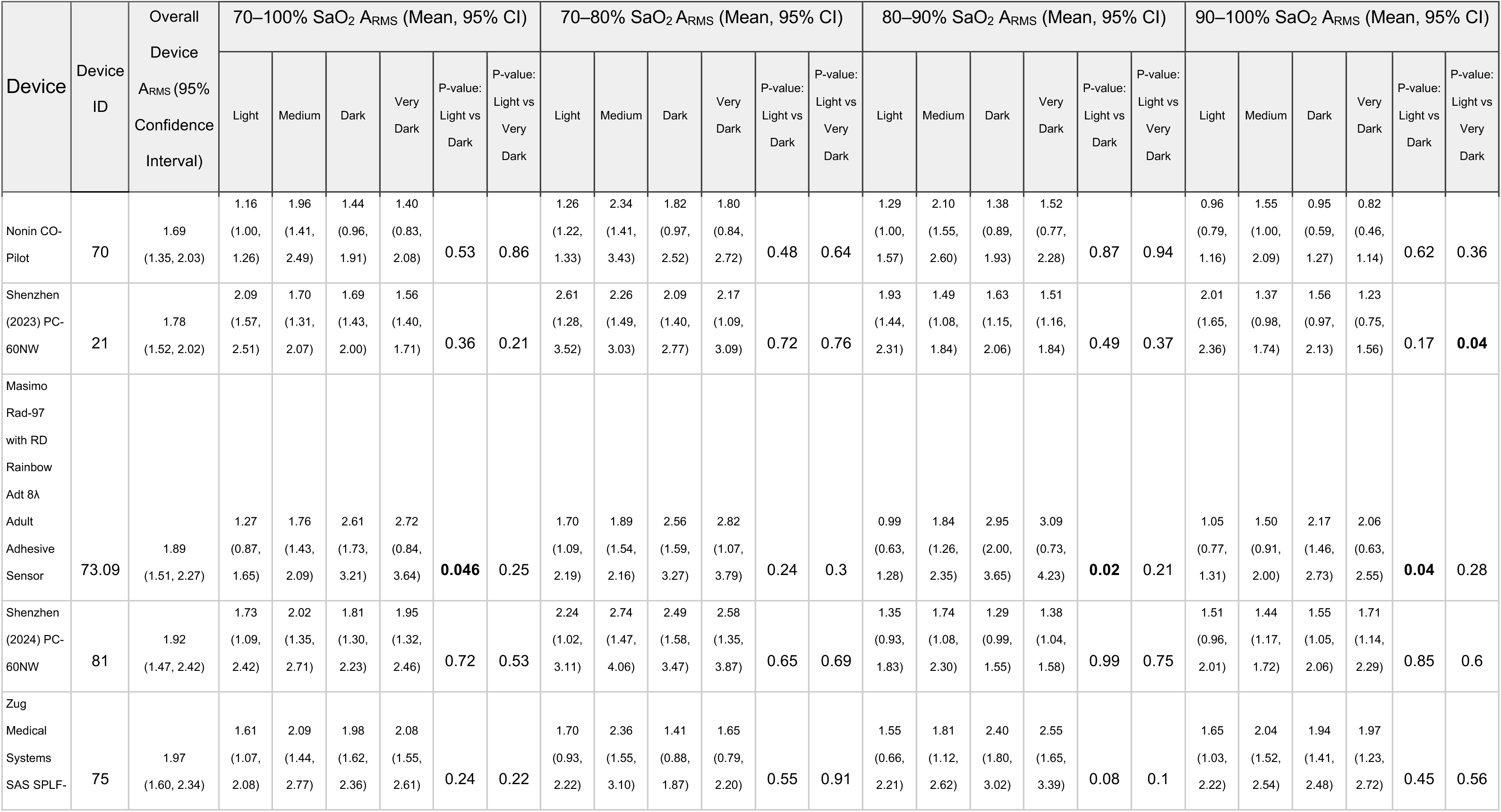
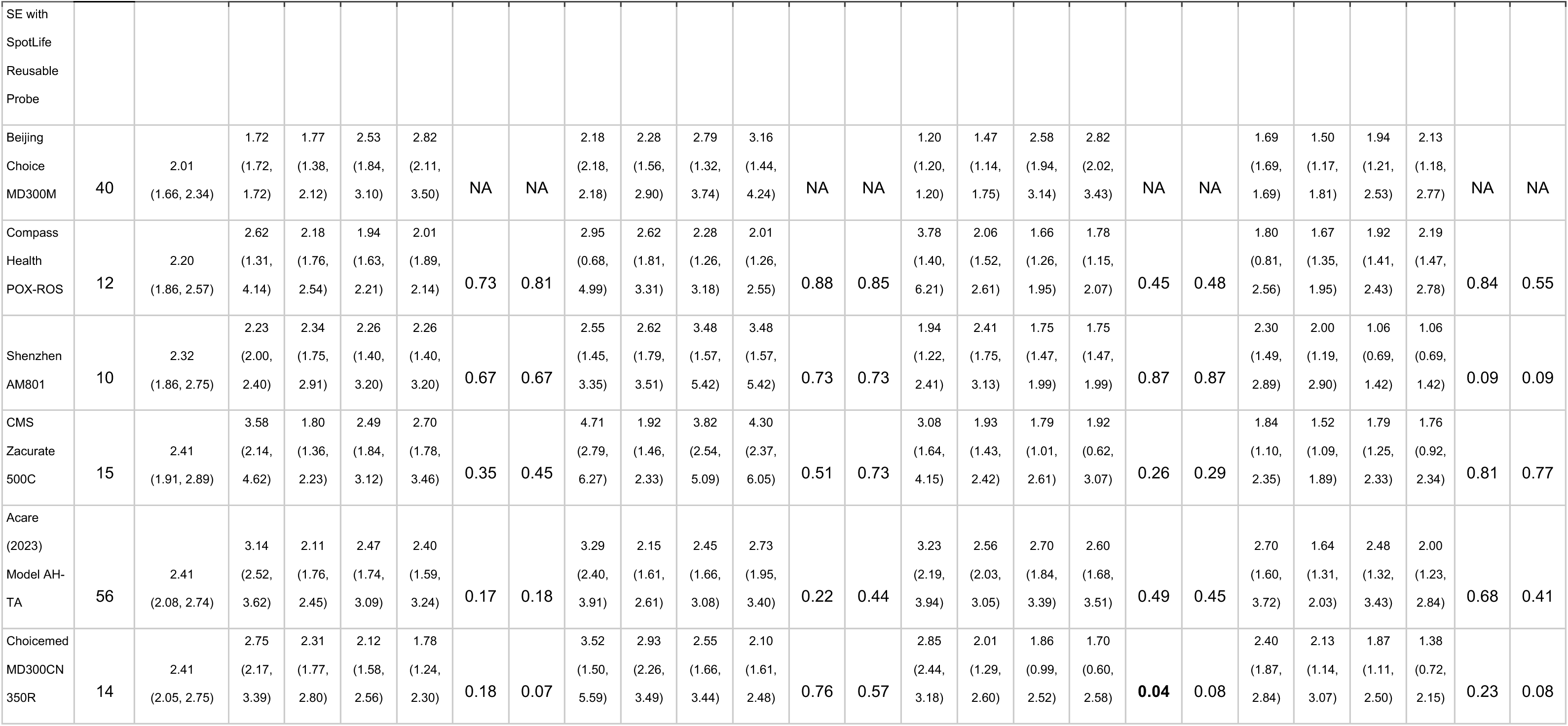
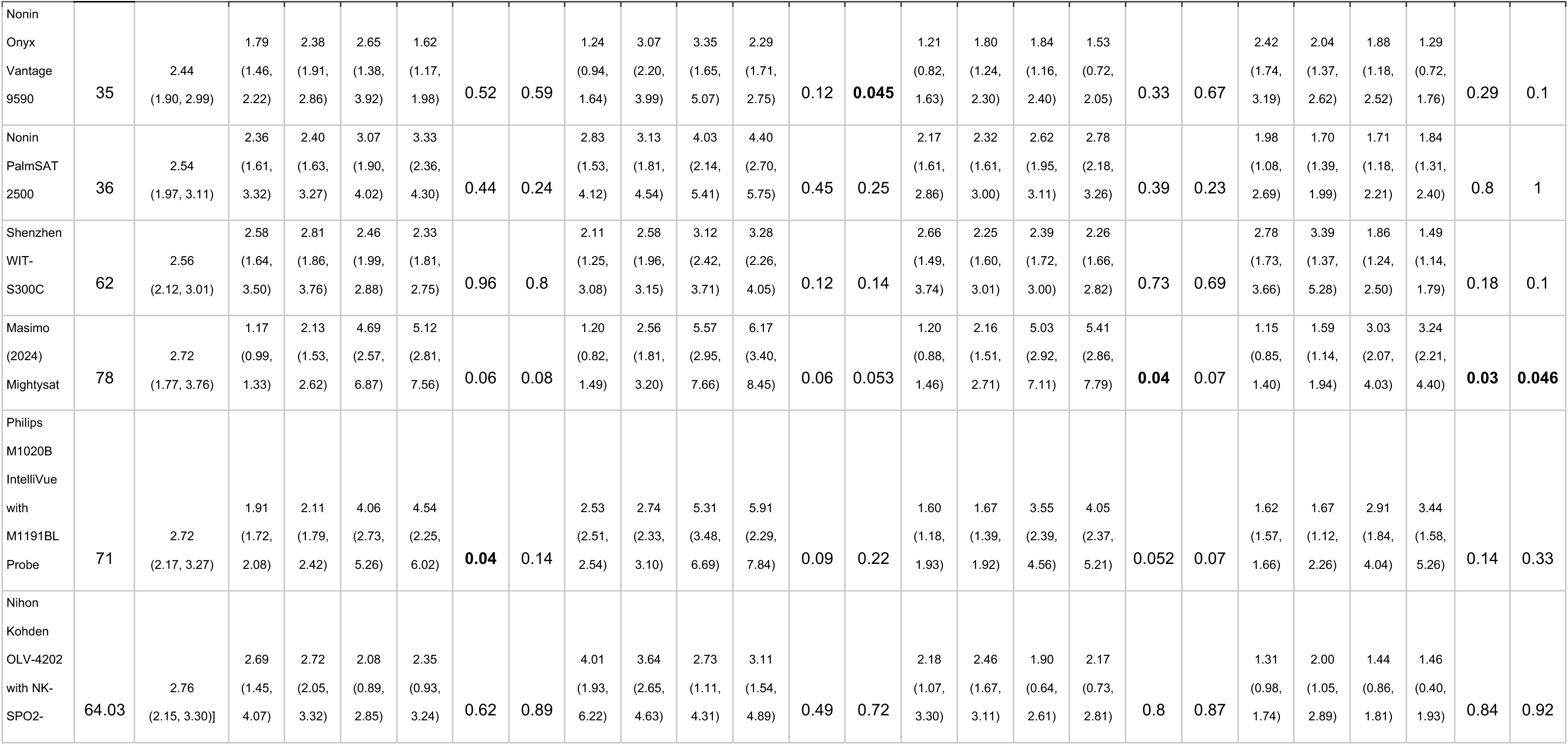
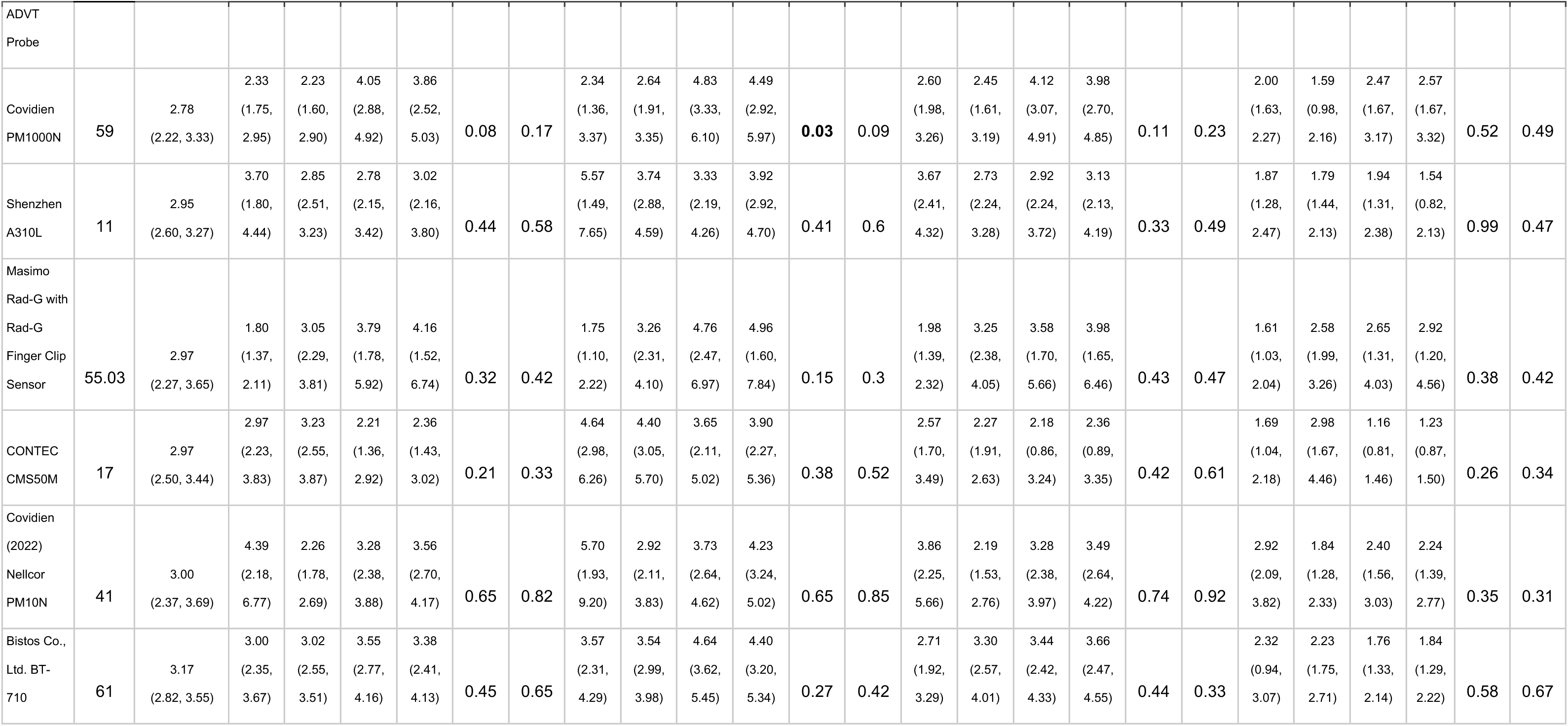
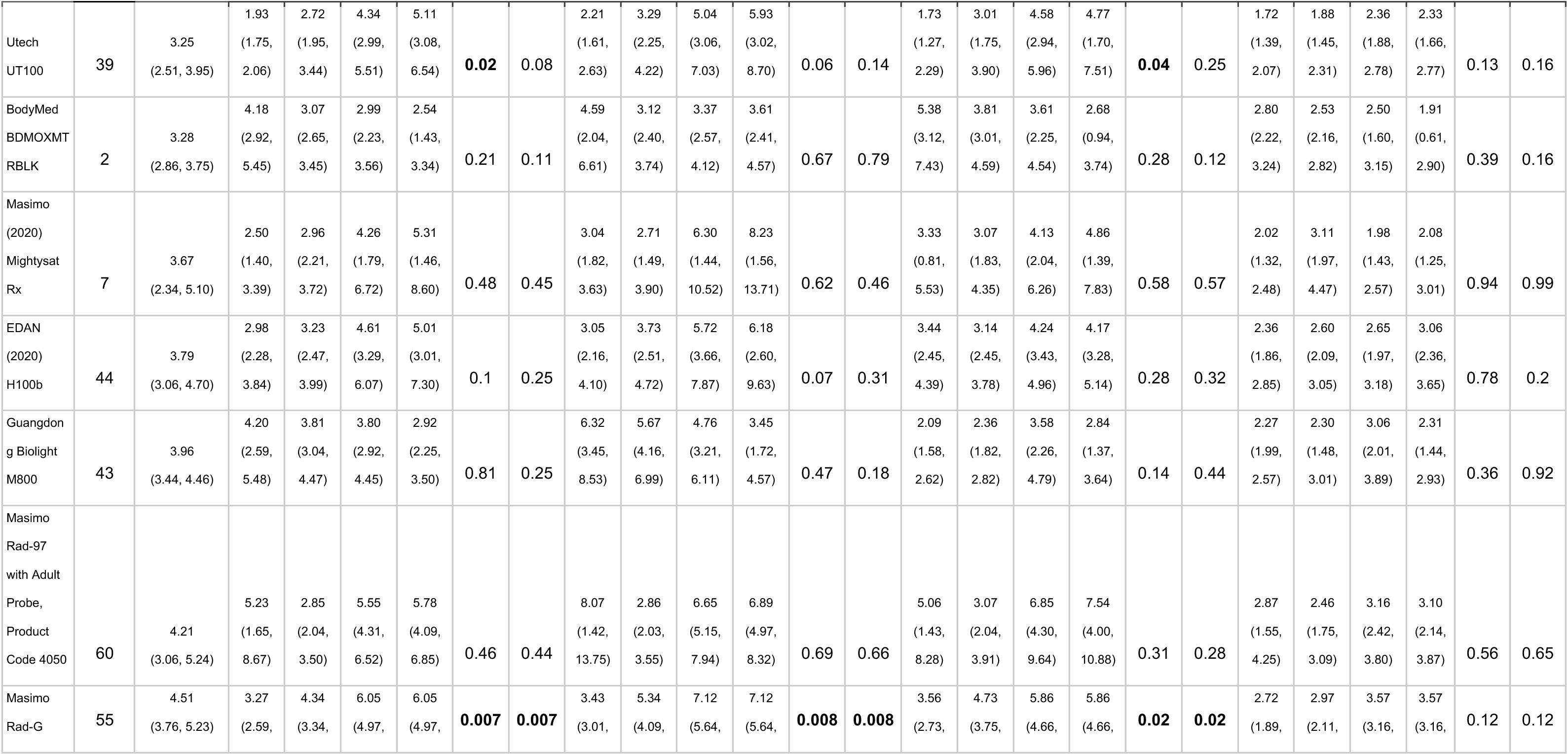
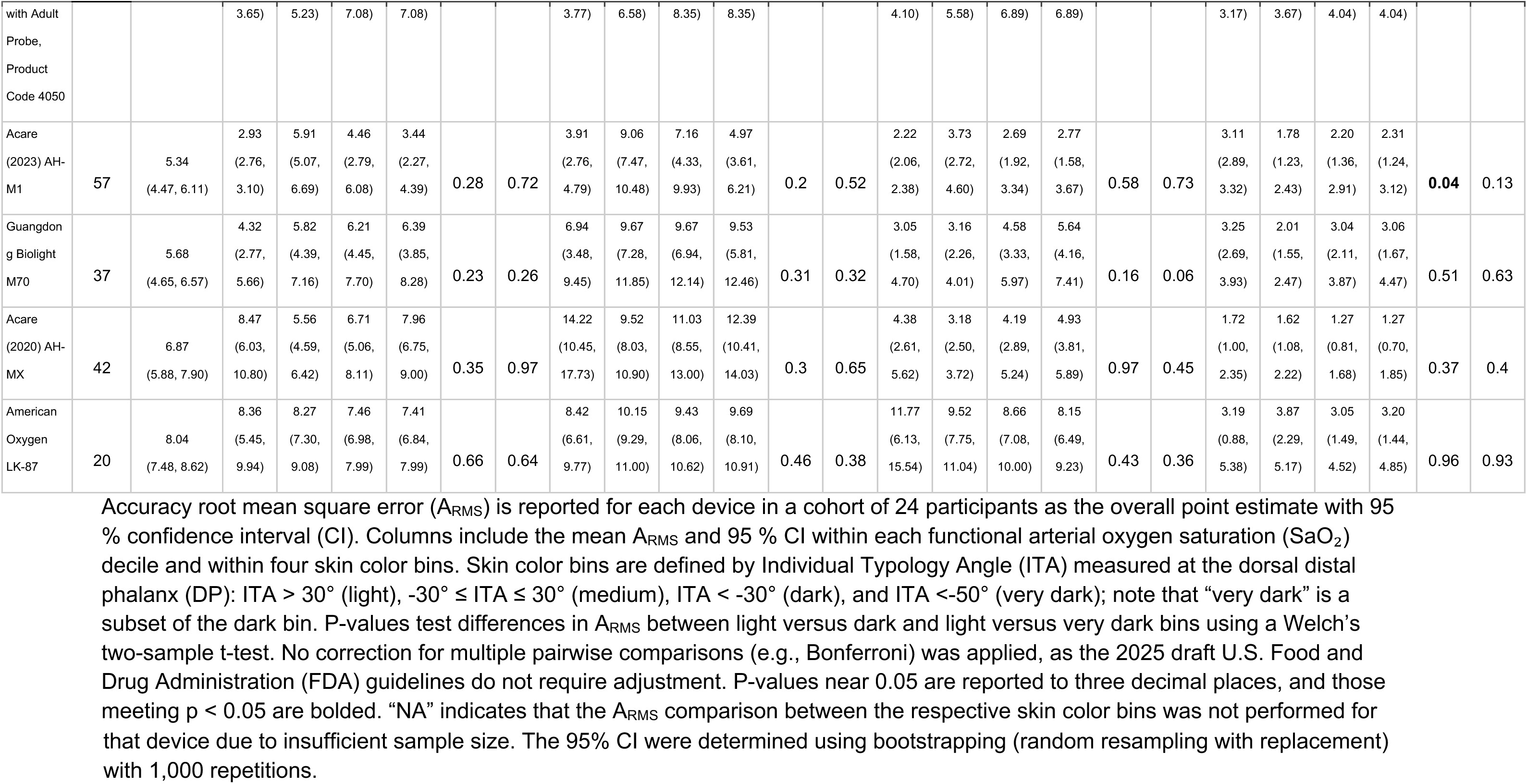
A_RMS_ at the DP for 24 participant cohorts.

**Table S2.**
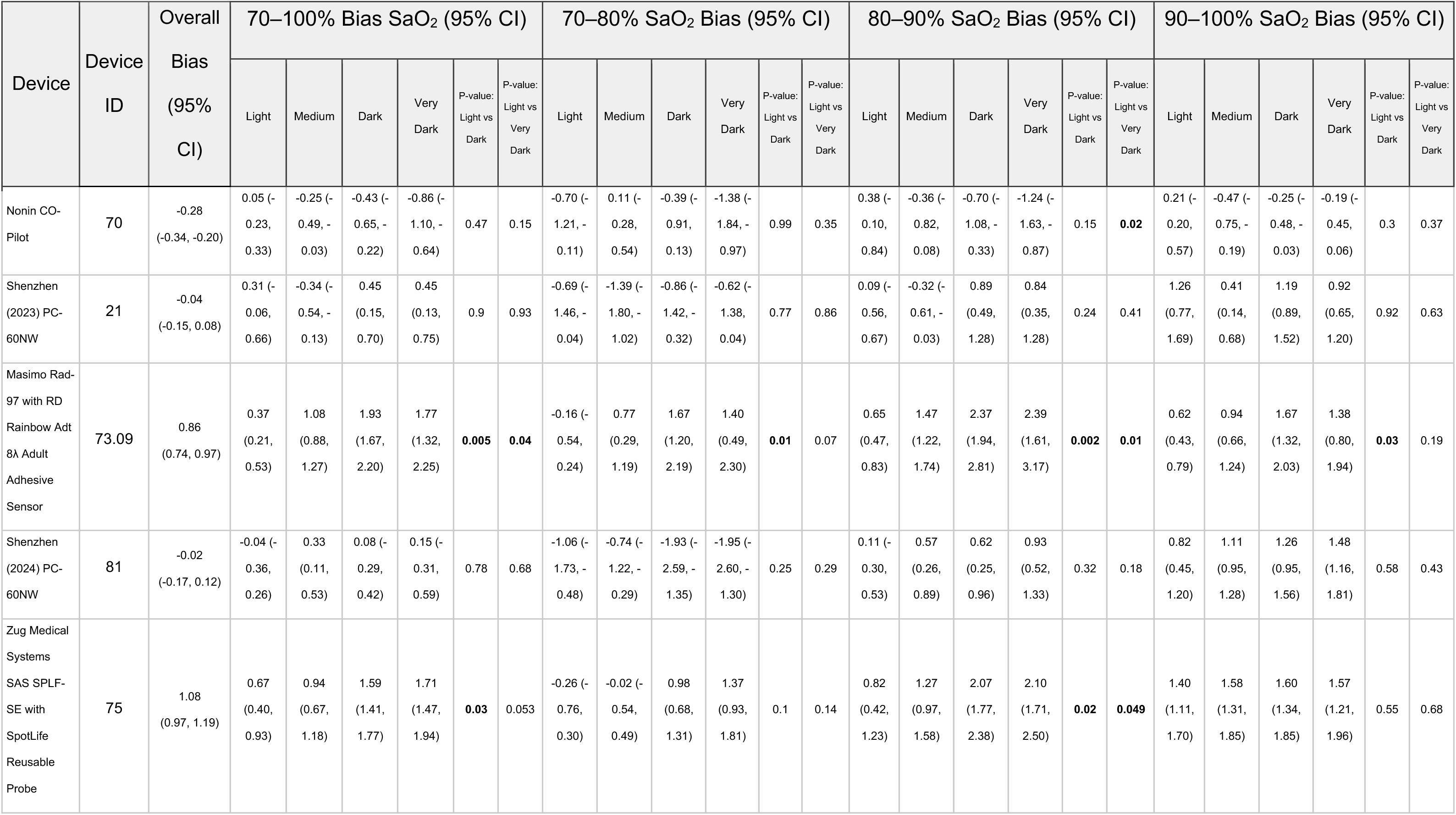
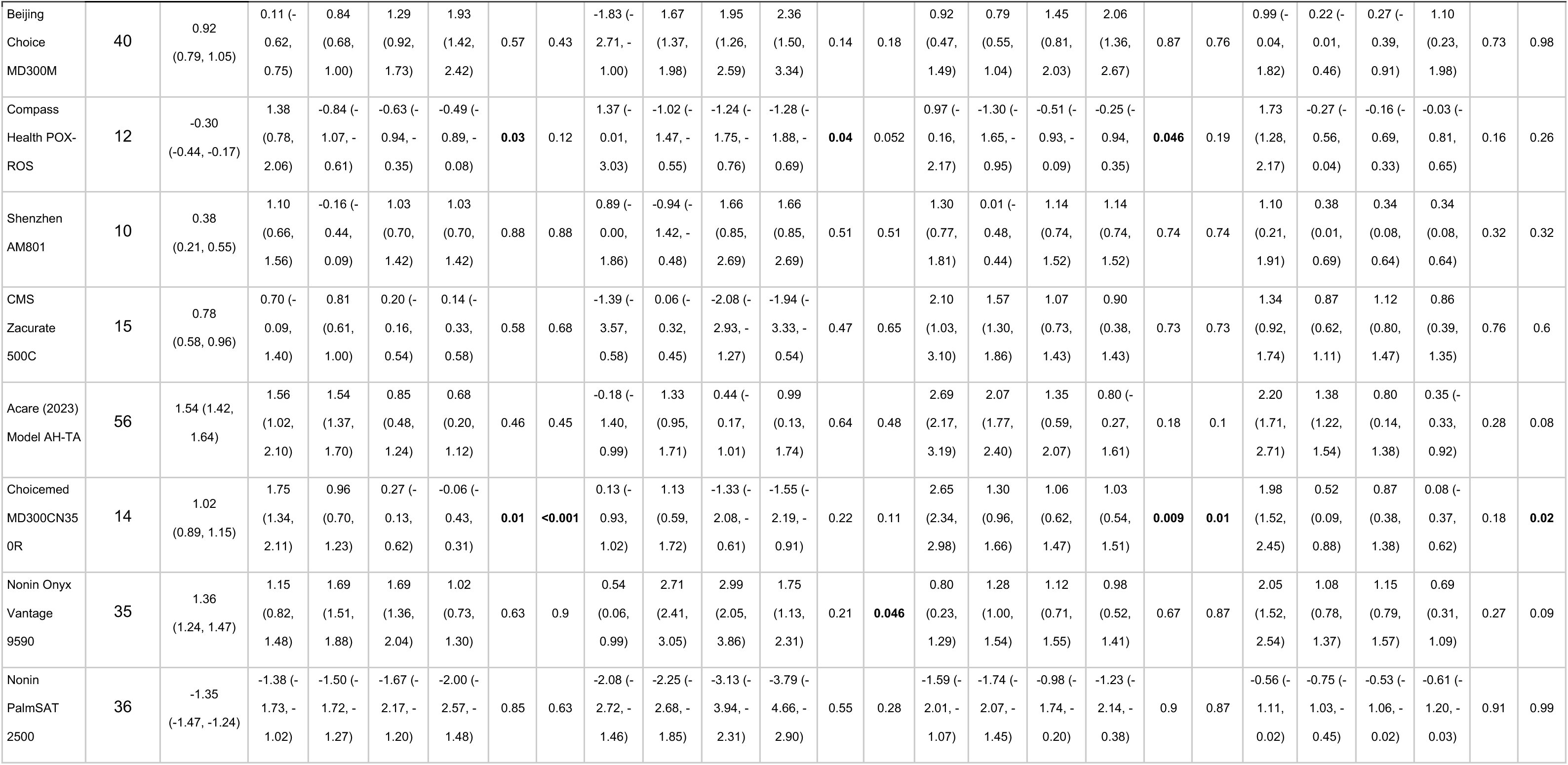
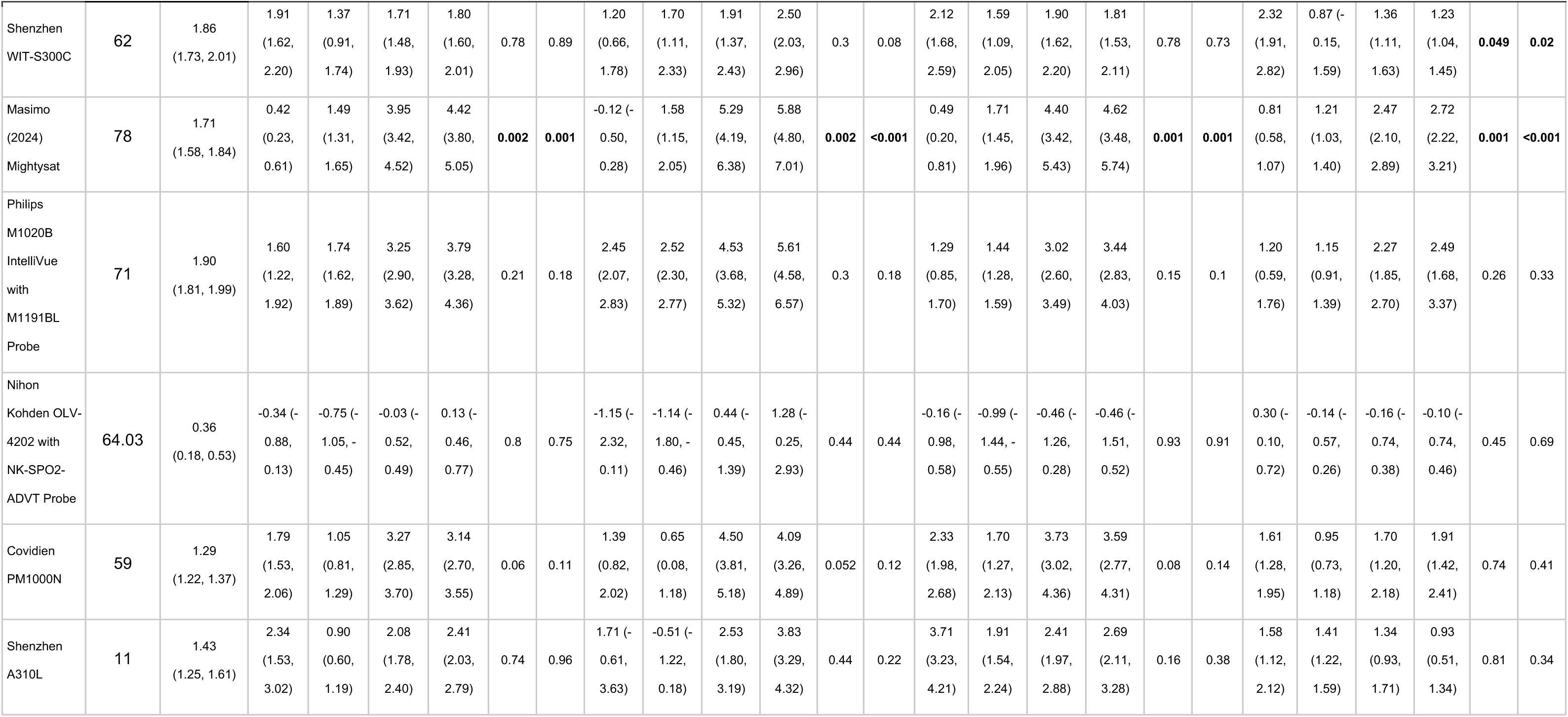
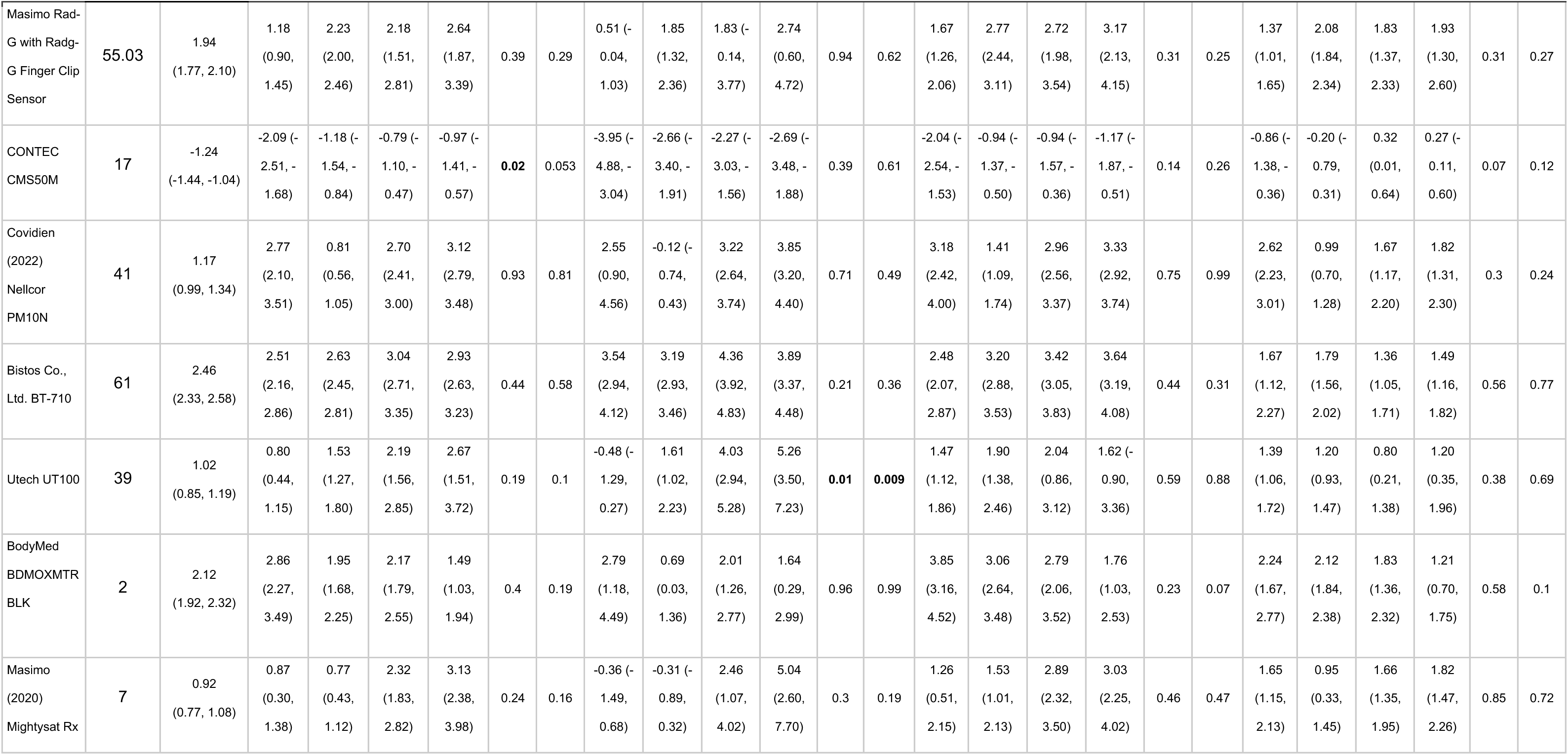
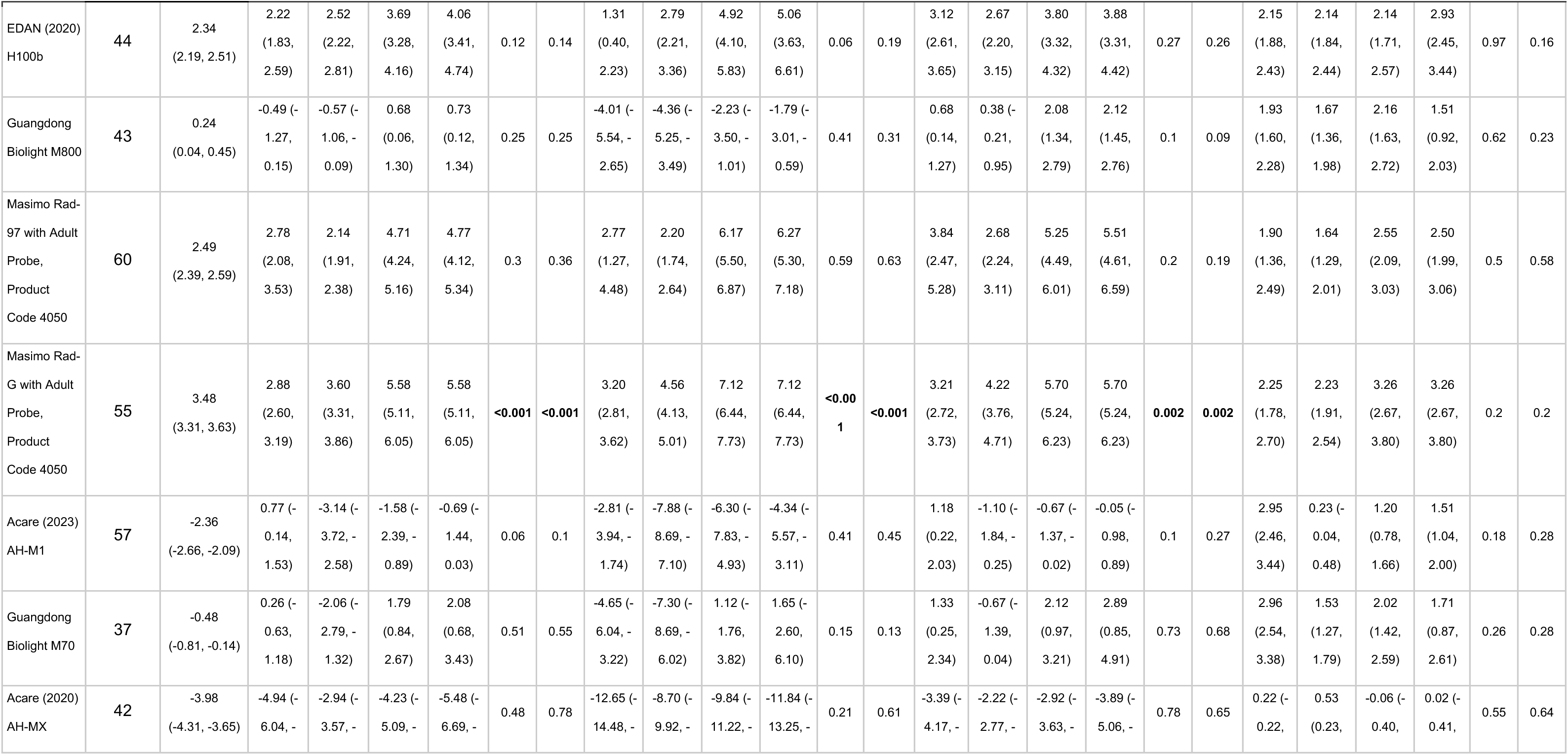
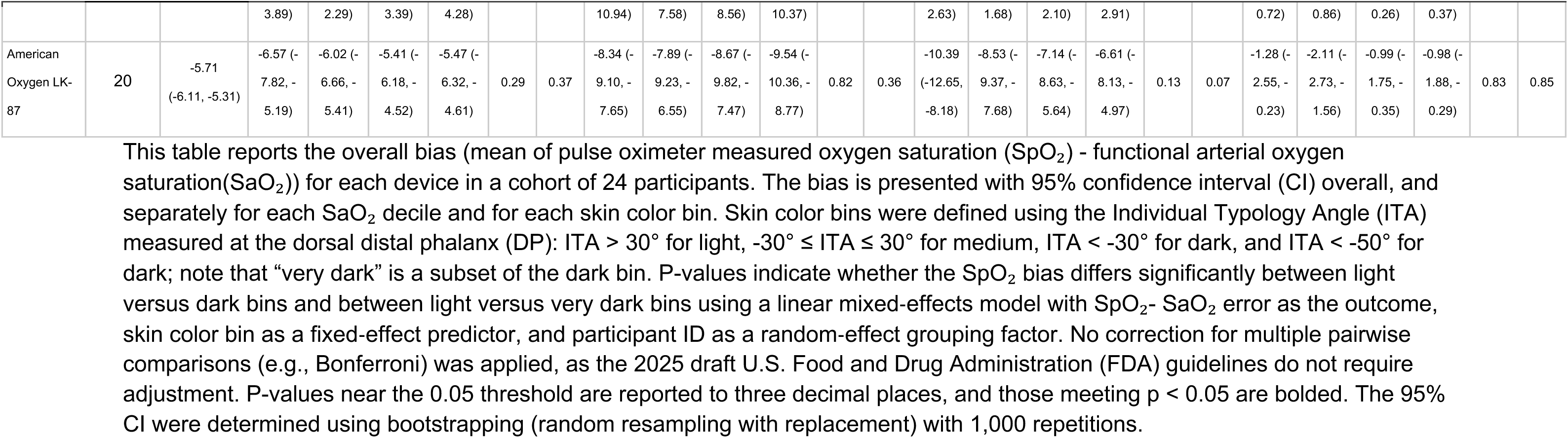
Bias (mean of SpO_2_ minus SaO_2_) at the DP for 24 participant cohorts.

**Table S3.**
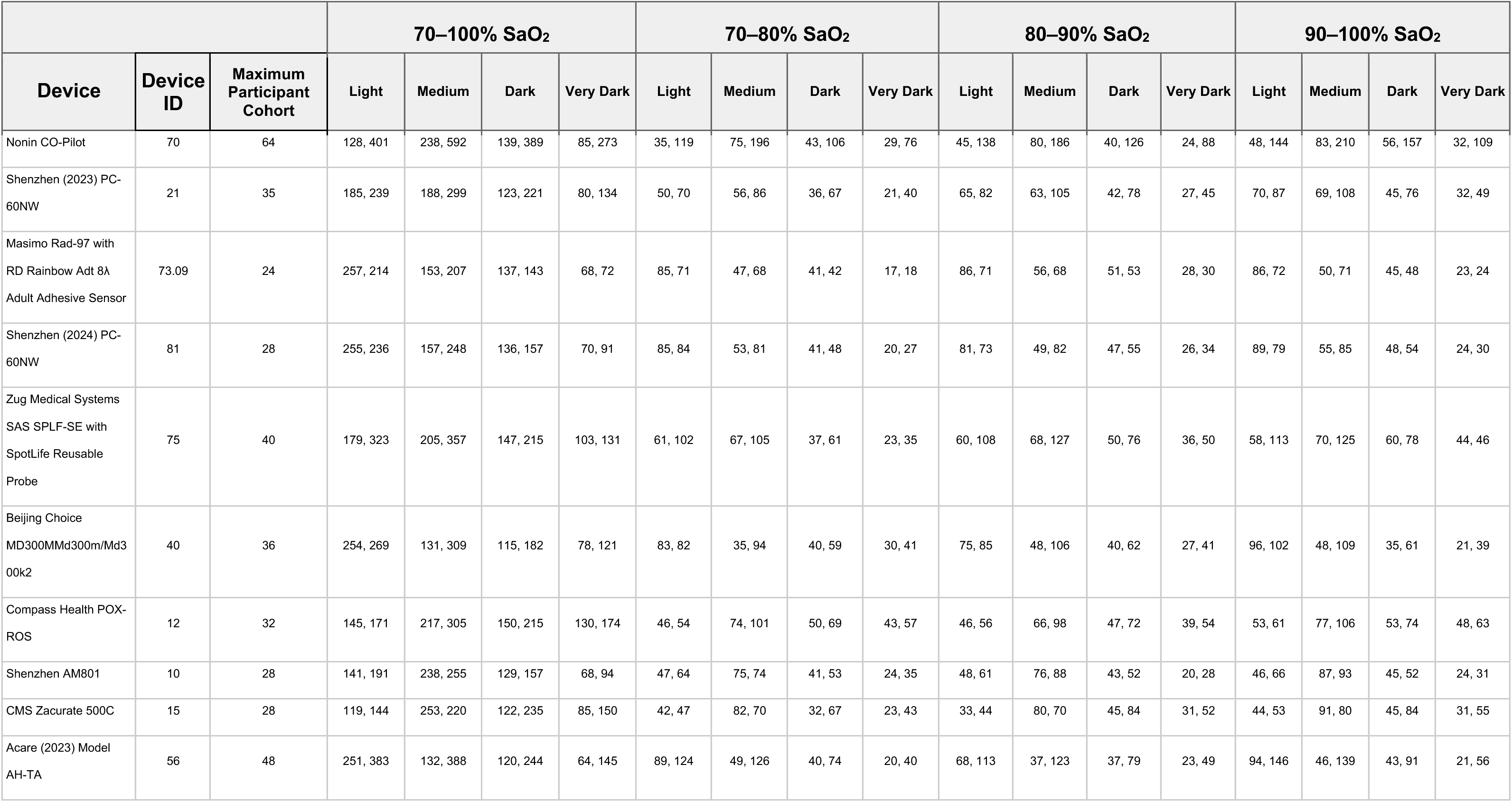
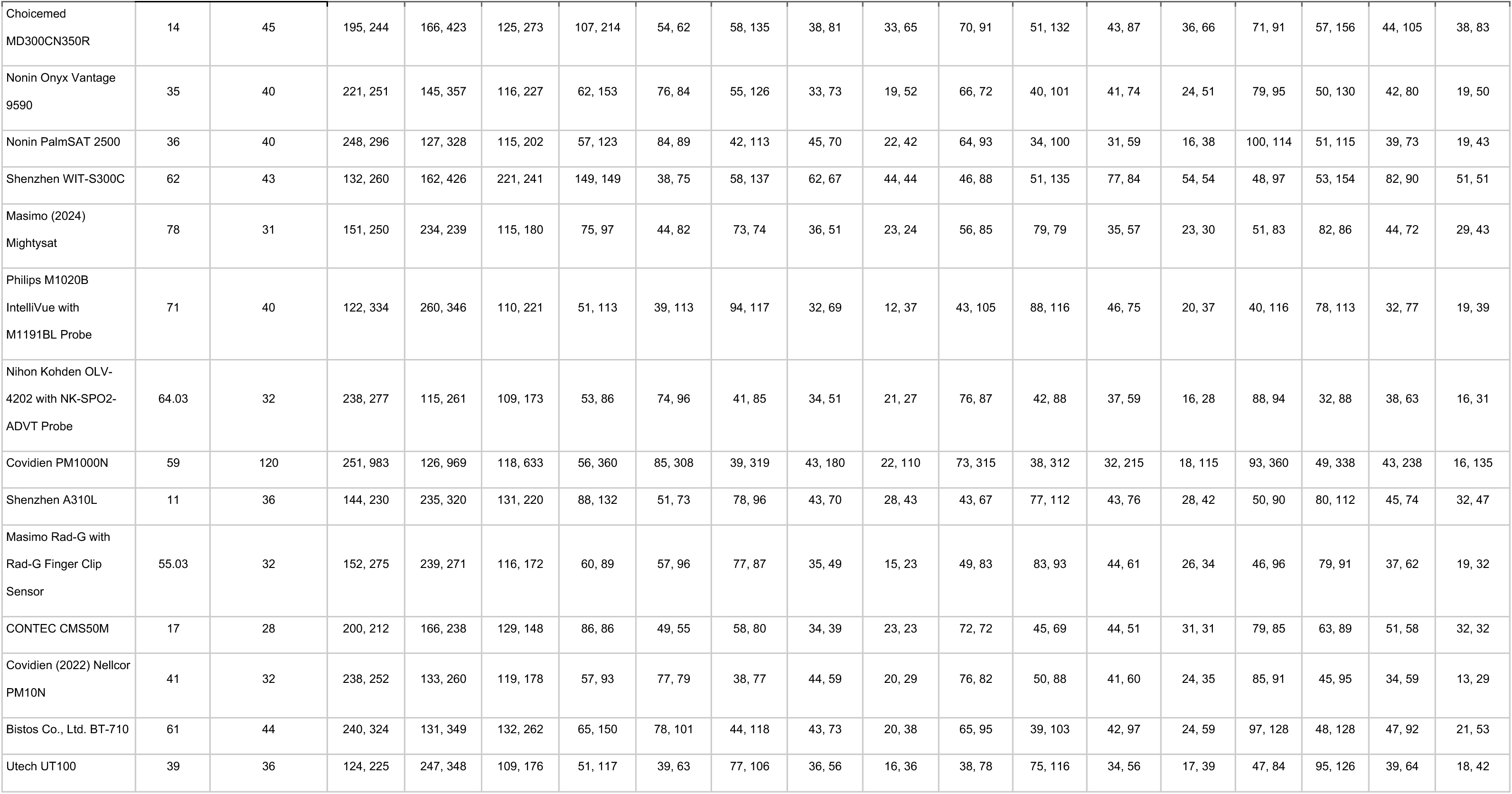
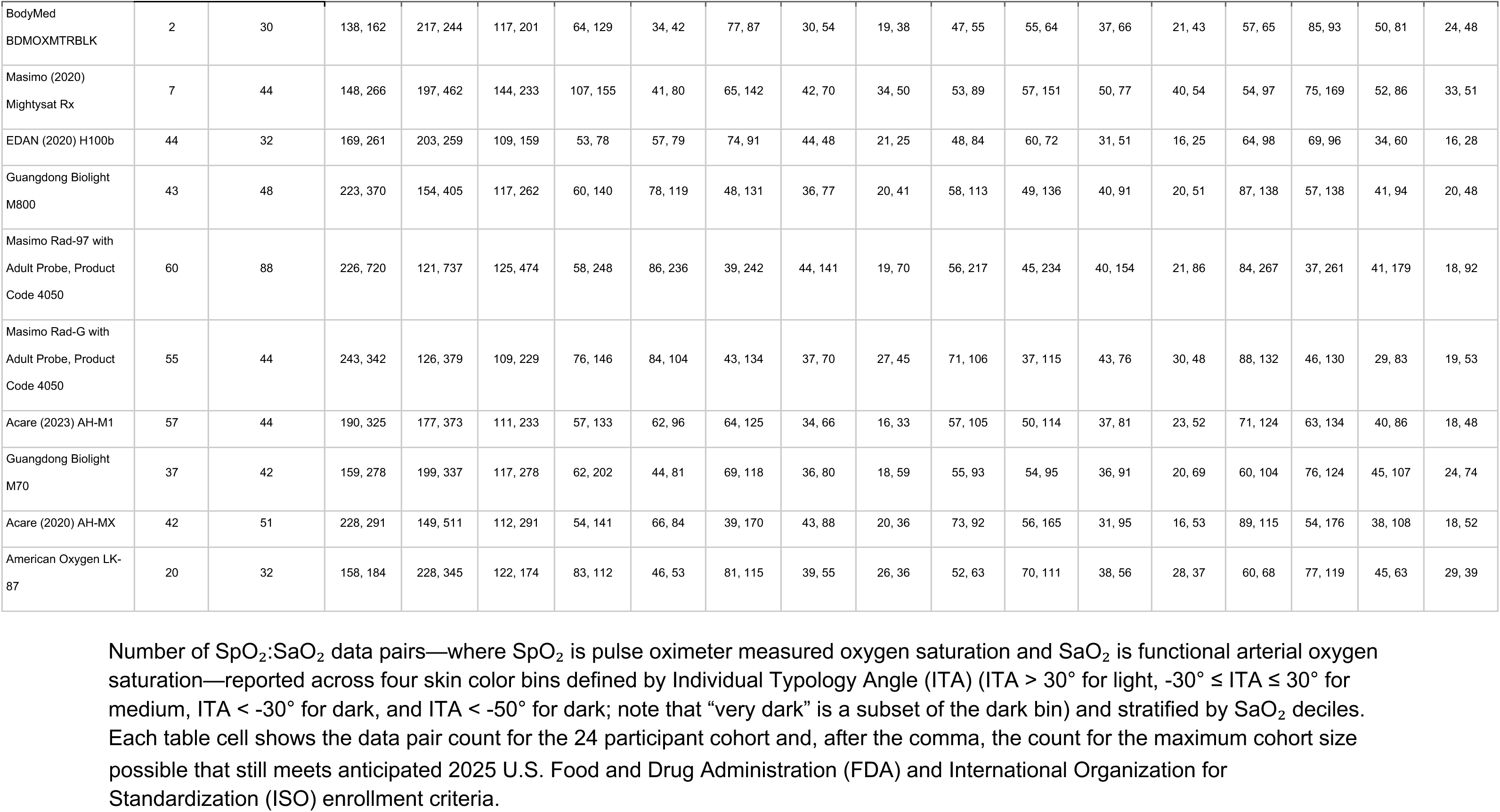
Number of SaO_2_ measurements (24 participant cohorts, max participant cohorts) stratified by skin color measured at the forehead.

**Table S4.** Complete list of devices, prices, specifications, and regulatory markings.

| Device | Device ID | Oximeter Type | Probe Name (if applicable) | Cost | 510k | CE Marking | ISO 80601-2-61 |
| --- | --- | --- | --- | --- | --- | --- | --- |
| Nonin CO-Pilot | 70 | Handheld | 8330AA Adult Reusable Sensor * | \$5,999 | ✓ | ✓ | ✓ |
| Shenzhen (2023) PC-60NW | 21 | Fingertip | | \$40 | ✓ | ✓ | ✓ |
| Masimo Rad-97 | 73.09 | Tabletop | RD Rainbow Adt 8λ Adult Adhesive Sensor * | \$1,730 | ✓ | ✓ | ✓ |
| Shenzhen (2024) PC-60NW | 81 | Fingertip | | \$40 | ✓ | | ✓ |
| Zug Medical Systems<br>SAS SPLF-SE | 75 | Tabletop | SpotLife Reusable Adult SpO2 Probe | \$1,125 | | | |
| Beijing Choice MD300M | 40 | Handheld | M-50E Probe | \$259 | ✓ | ✓ | |
| Compass Health POX-ROS | 12 | Fingertip | | \$21 | | | |
| Shenzhen AM801 | 10 | Fingertip | | \$30 | ✓ | | ✓ |
| CMS Zacurate 500C | 15 | Fingertip | | \$23 | | | |
| Acare (2023) Model AH-TA | 56 | Handheld | ASANR-D3 Probe | \$250 | ✓ | ✓ | ✓ |
| Choicemed<br>MD300CN350R | 14 | Fingertip | | \$30 | | ✓ | |
| Nonin Onyx Vantage 9590 | 35 | Fingertip | | \$200 | ✓ | ✓ | ✓ |
| Nonin PalmSAT 2500 | 36 | Handheld | 8000AA-1M Probe | \$500 | ✓ | ✓ | ✓ |
| Shenzhen WIT-S300C | 62 | Handheld | WTL403-01 Probe | \$250 | ✓ | ✓ | |
| Masimo (2024)<br>Mightysat | 78 | Fingertip | | \$299 | ✓ | ✓ | ✓ |
| Philips M1020B<br>IntelliVue | 71 | Tabletop | M1191BL Probe | \$4,470 | ✓ | ✓ | |
| Nihon Kohden OLV-4202 | 64.03 | Tabletop | NK-SPO2-ADVT Probe |  |  | ✓ |  |
| Covidien PM1000N | 59 | Tabletop | DS100A probe | \$2,000 | ✓ | | |
| Shenzhen A310L | 11 | Fingertip | | \$10 | ✓ | | |
| Masimo Rad-G Pulse<br>Oximeter | 55.03 | Handheld | Masimo Rad-G Reusable Finger Clip Sensor | \$485 | ✓ | ✓ | ✓ |
| CONTEC CMS50M | 17 | Fingertip | | \$20 | | ✓ | |
| Covidien (2022) Nellcor<br>PM10N | 41 | Handheld | DS100A Probe | \$500 | ✓ | ✓ | ✓ |
| Bistos Co., Ltd. BT-710 | 61 | Handheld | U403-01 Probe | \$350 | | ✓ | ✓ |
| Utech UT100 | 39 | Handheld | CSS029B-1 Probe | \$190 | | | |
| BodyMed<br>BDMOXMTRBLK | 2 | Fingertip | | \$25 | | | |
| Masimo (2020)<br>Mightysat Rx | 7 | Fingertip | | \$299 | ✓ | ✓ | ✓ |
| EDAN (2020) H100b | 44 | Handheld | Default DB9 Adult Reusable Finger Sensor | \$268 | ✓ | ✓ | ✓ |
| Guangdong Biolight<br>M800 | 43 | Handheld | Default Probe | \$329 | ✓ | ✓ | |
| Masimo Rad-97 | 60 | Tabletop | Reusable Adult Probe, Product Code 4050 | \$1,730 | ✓ | ✓ | ✓ |
| Masimo Rad-G | 55 | Handheld | Reusable Adult Probe, Product Code 4050 | \$485 | ✓ | ✓ | |
| Acare (2023) AH-M1 | 57 | Handheld | ASANR-D3 Probe | \$250 | ✓ | ✓ | |
| Guangdong Biolight M70 | 37 | Fingertip | | \$60 | ✓ | ✓ | |
| Acare (2020) AH-MX | 42 | Handheld | ASANR-D3 Probe | \$250 | ✓ | ✓ | |
| American Oxygen LK-87 | 20 | Fingertip | | \$25 | | ✓ | |

This table provides details on each of the 34 tested devices, including device type, probe name, cost (in U.S. dollars), and applicable regulatory markings or reported regulatory compliance. Regulatory compliance and markings were found by searching manufacturer websites, product literature, the FDA 510(k) Premarket Notification Database, and by looking at the labels on the devices themselves. Device probes with > 2 wavelengths are denoted by an asterisk (\*).

**Table S5.**
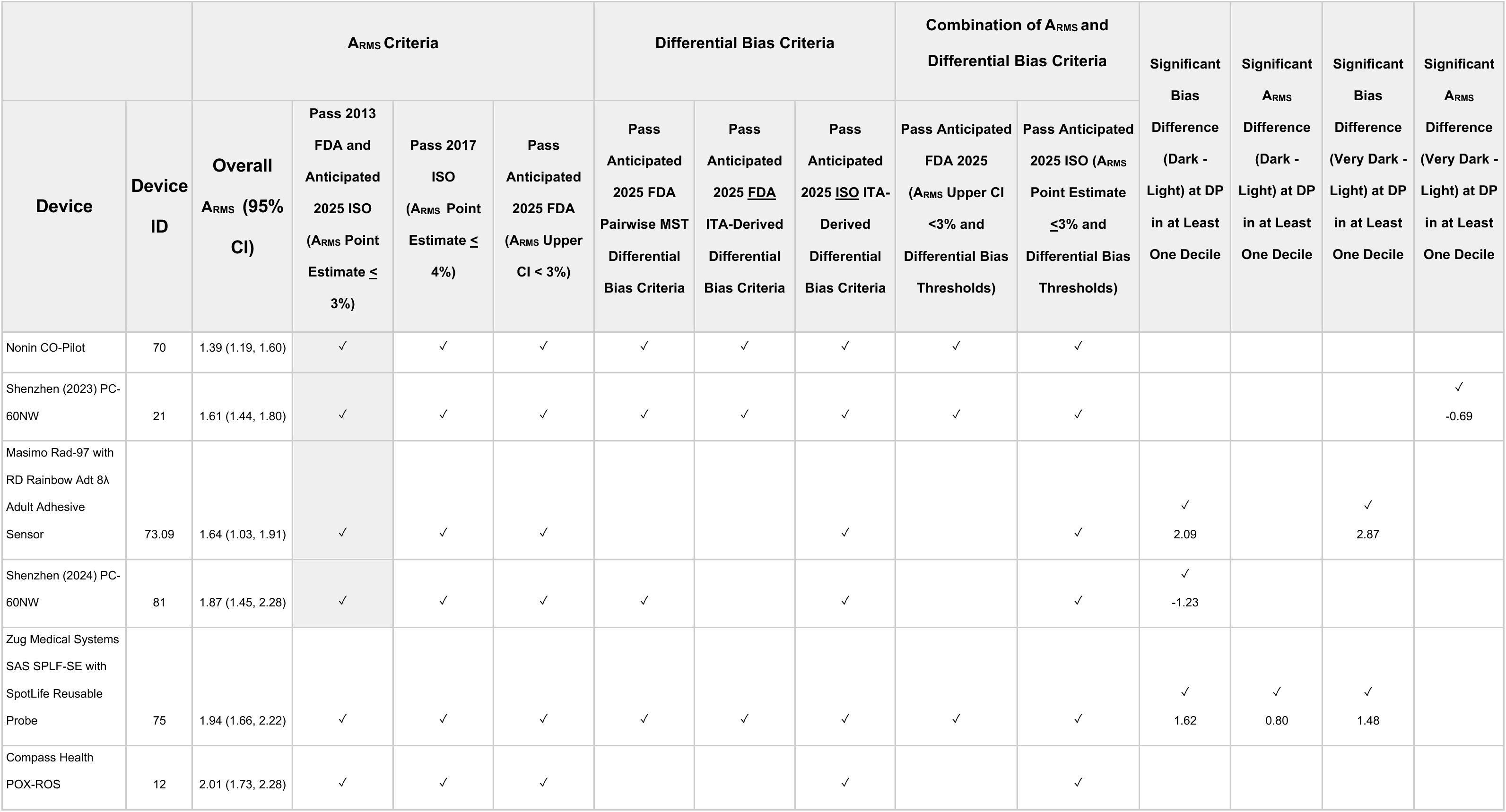
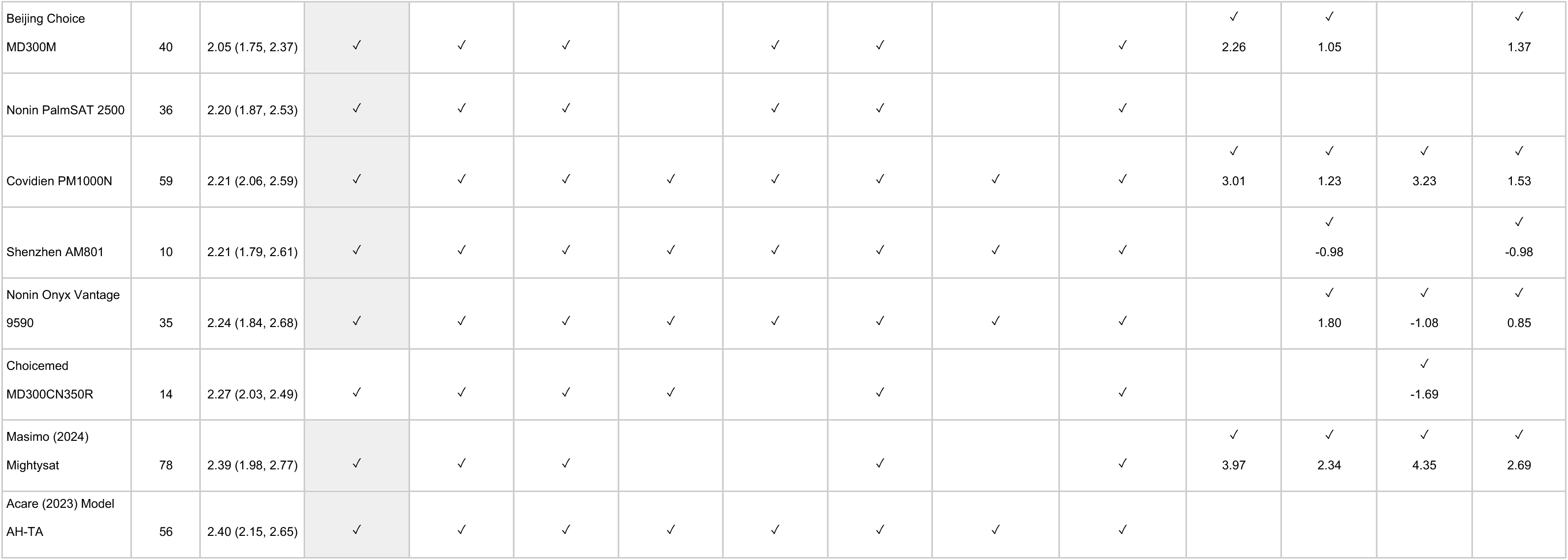
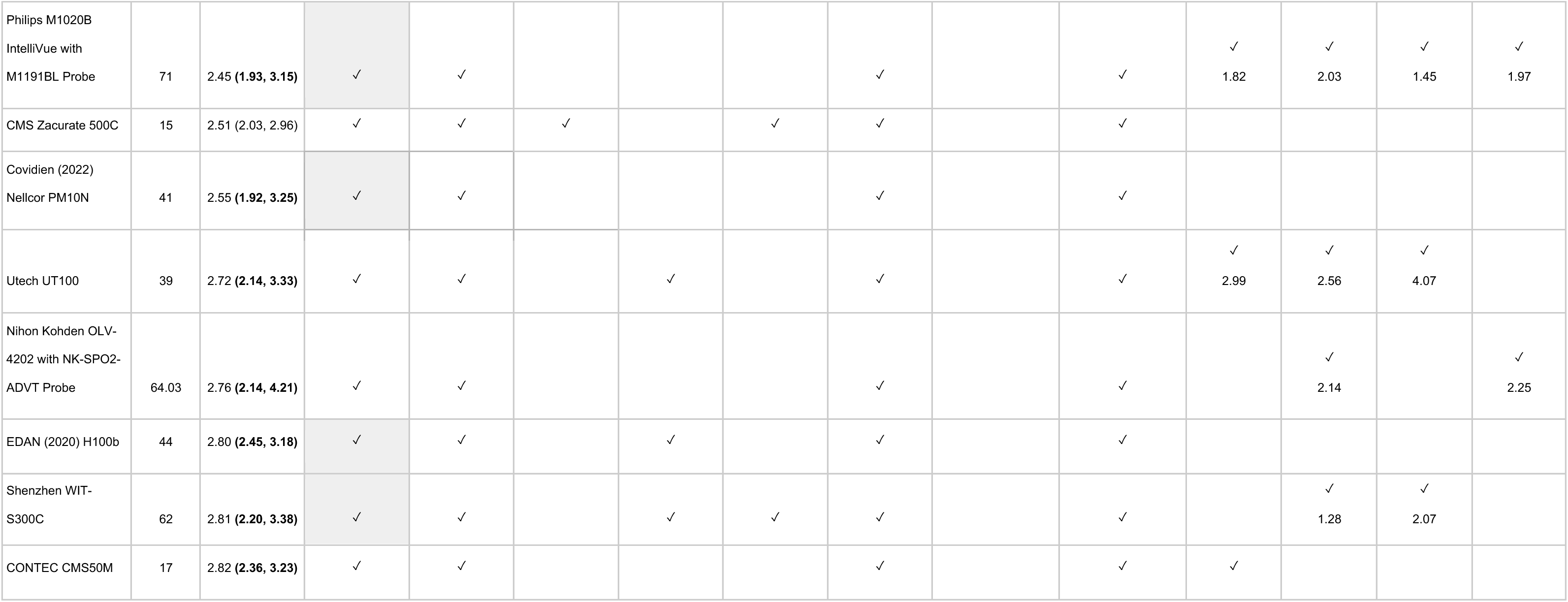
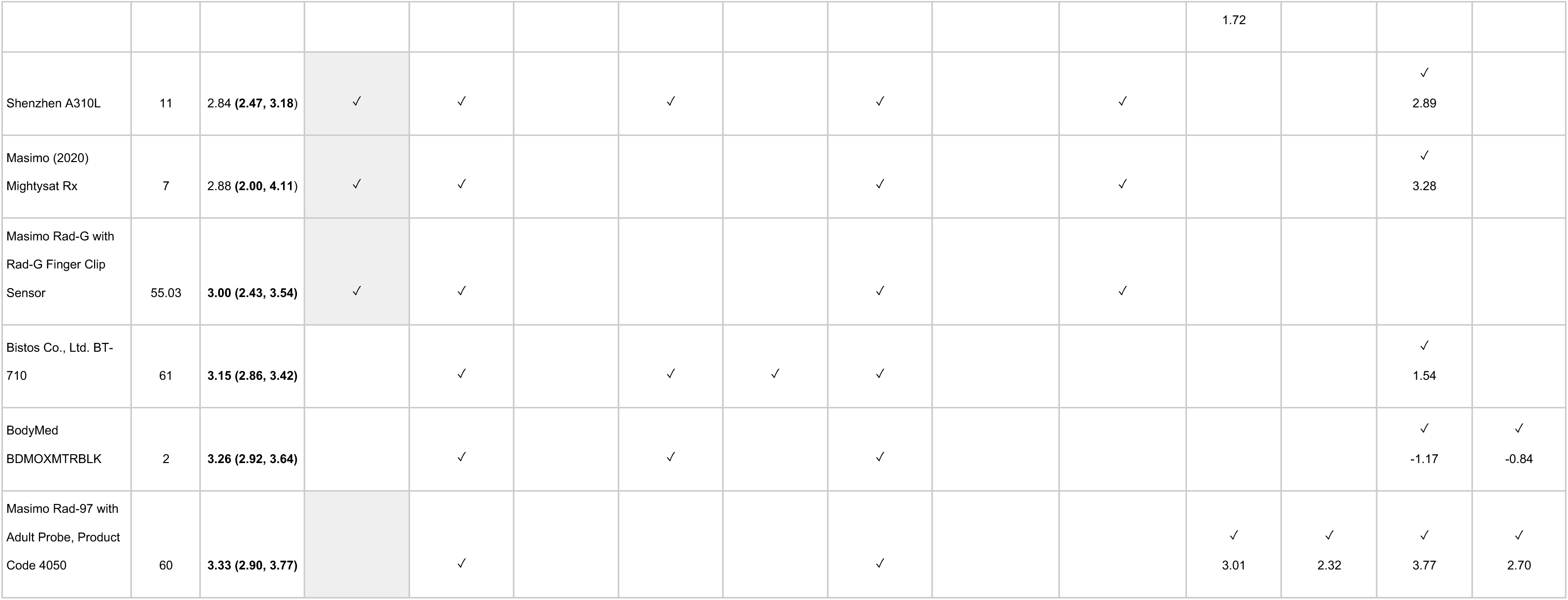
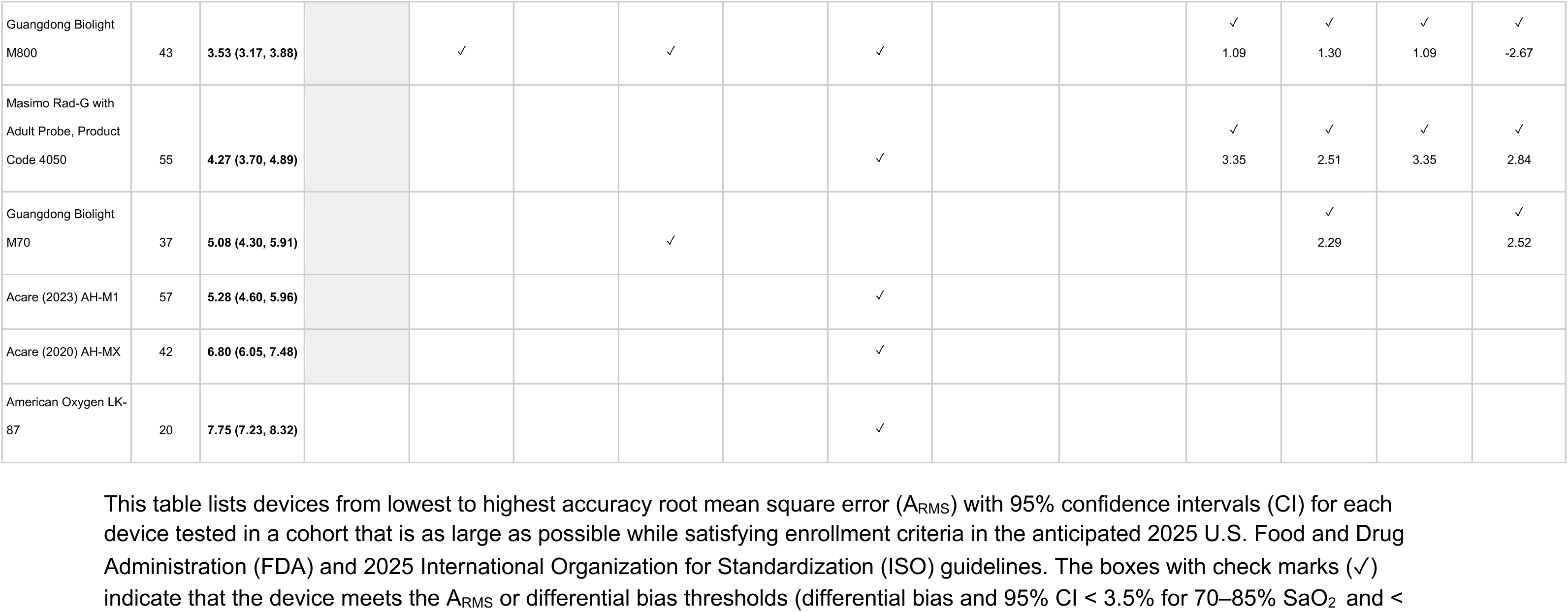
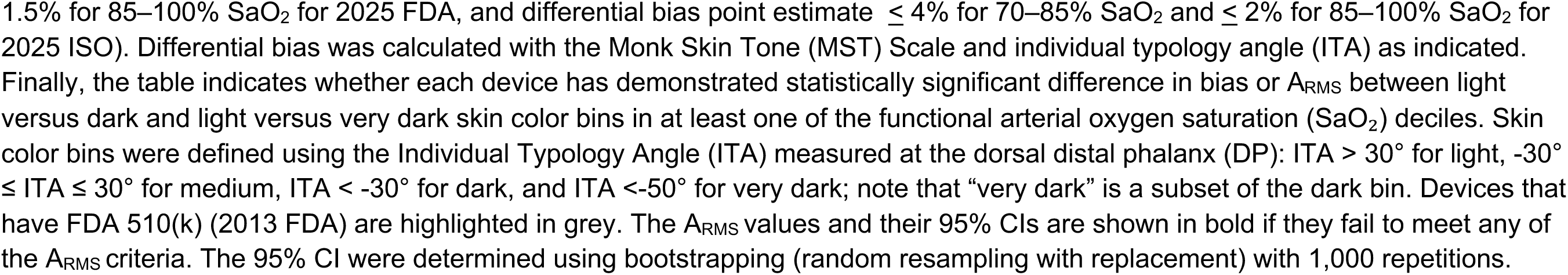
Device conformity with current and anticipated FDA and ISO performance thresholds for maximum cohort sizes and DP ITA.

**Table S6.**
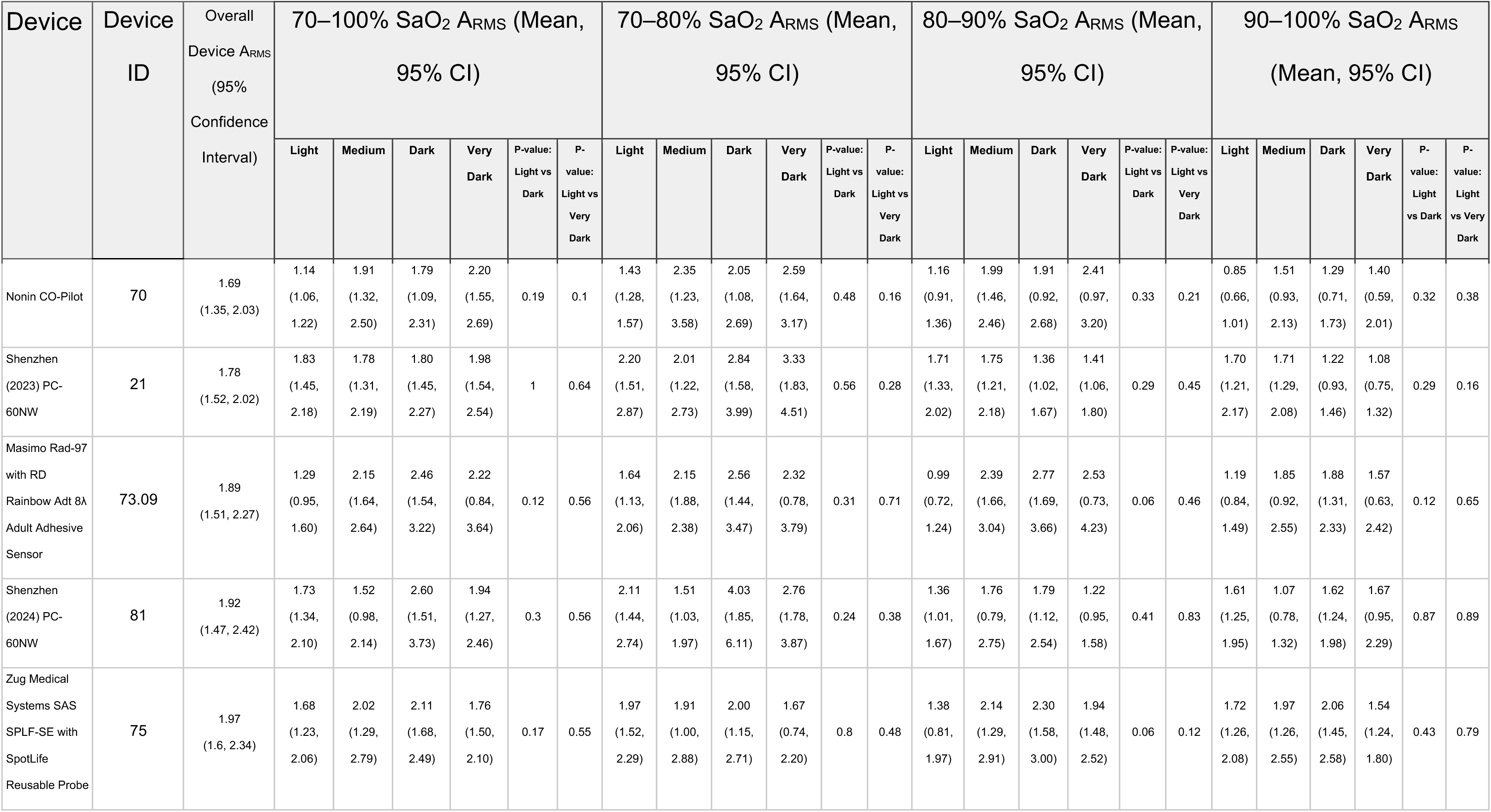
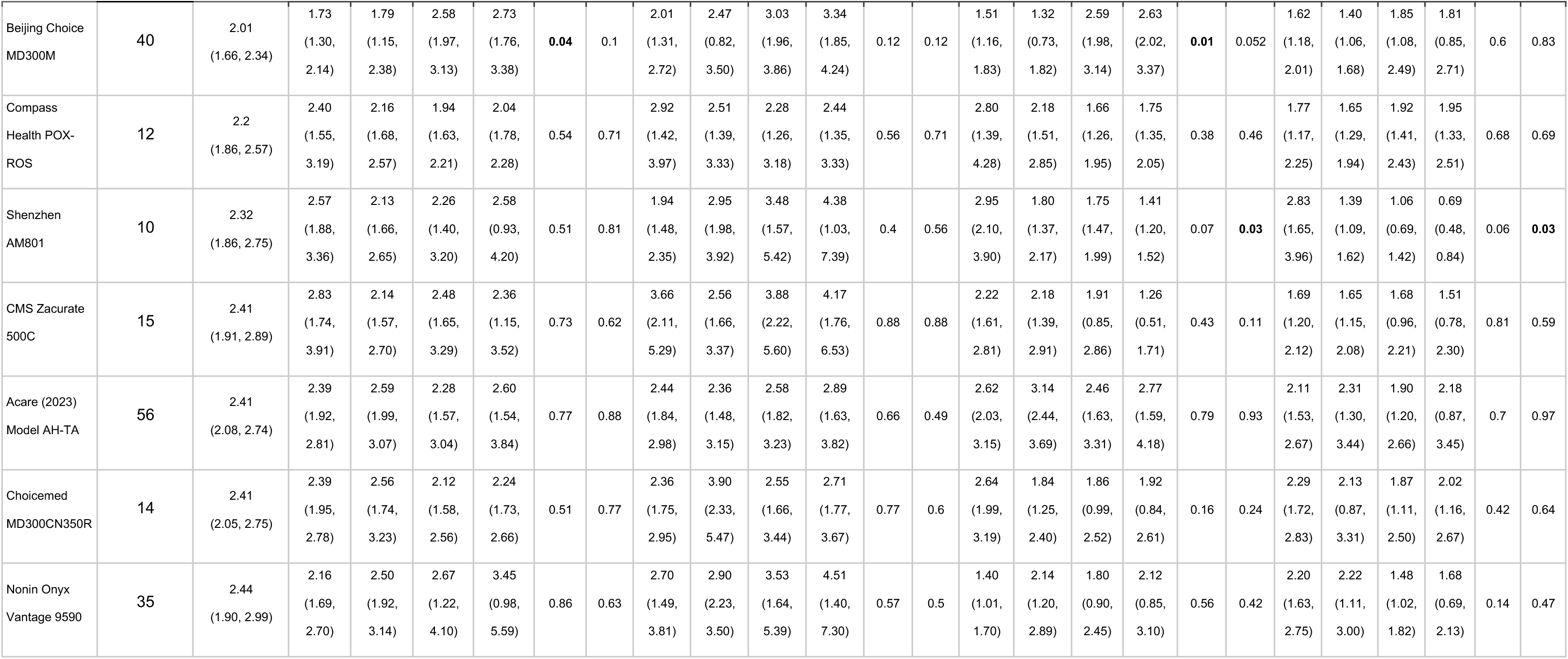
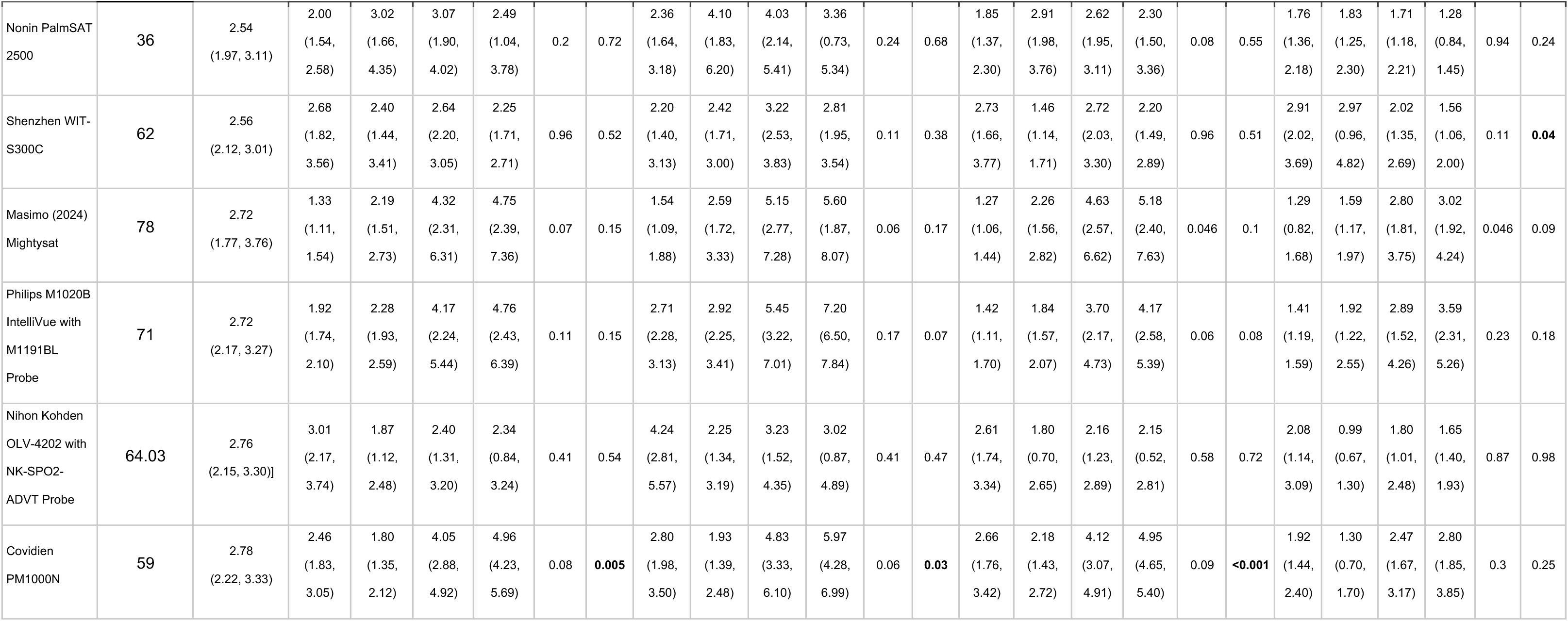
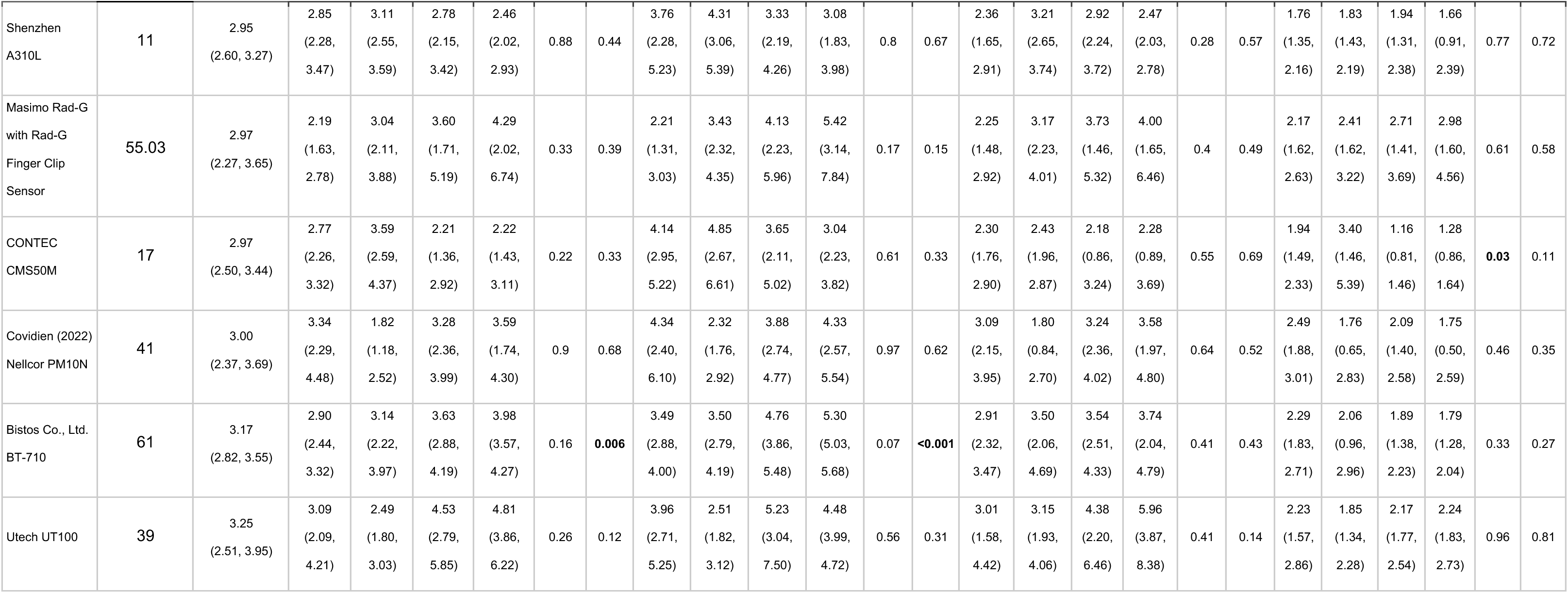
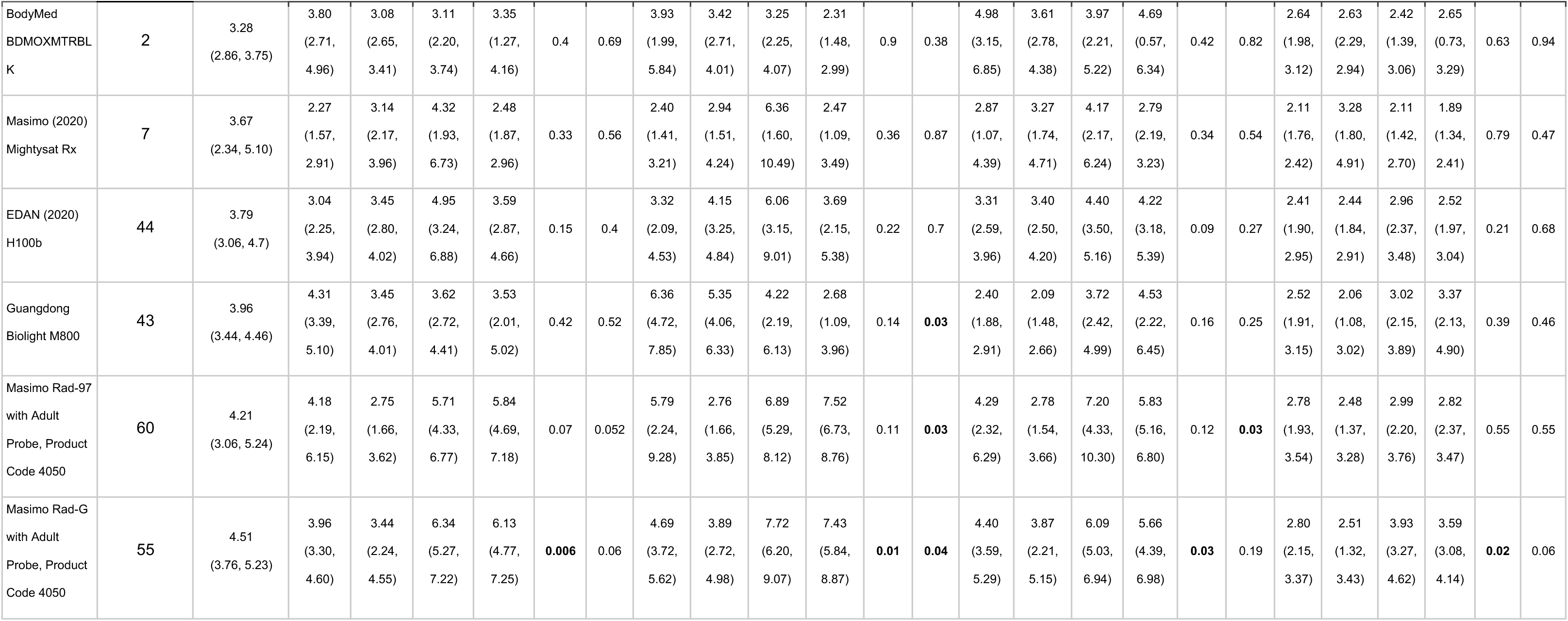
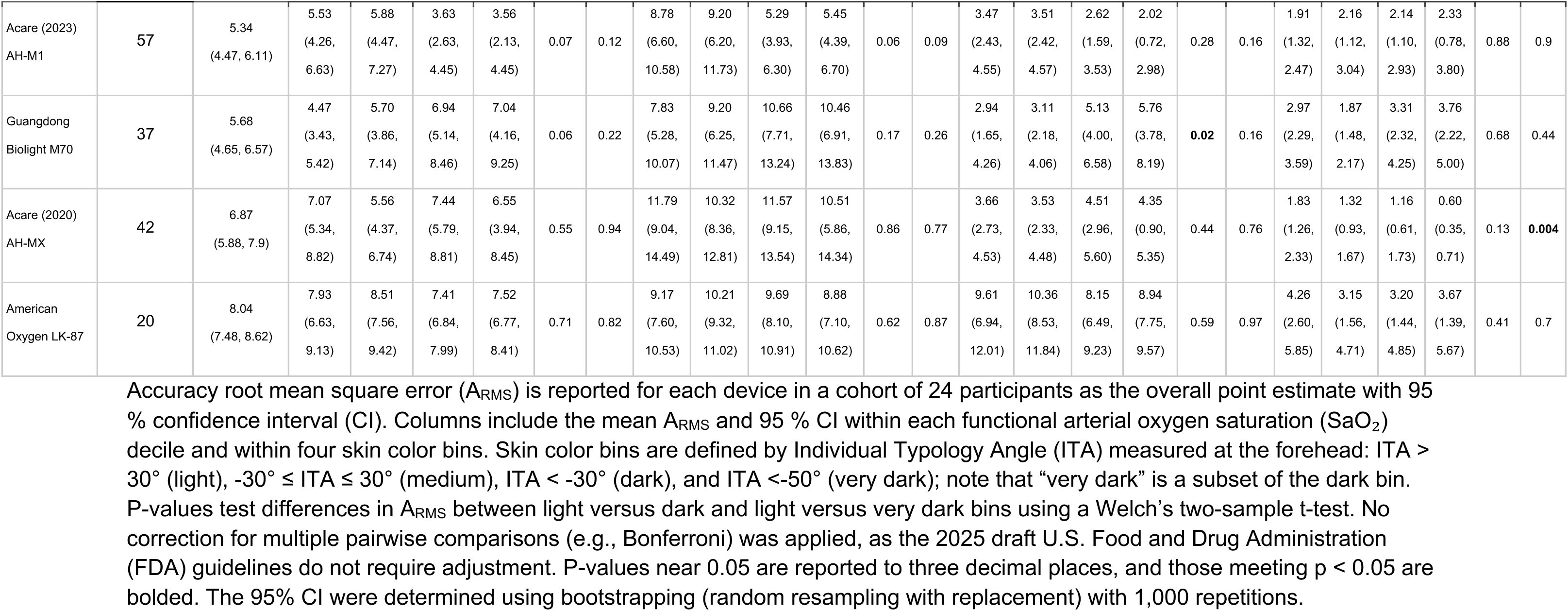
A_RMS_ at the forehead for 24 participant cohorts.

**Table S7.**
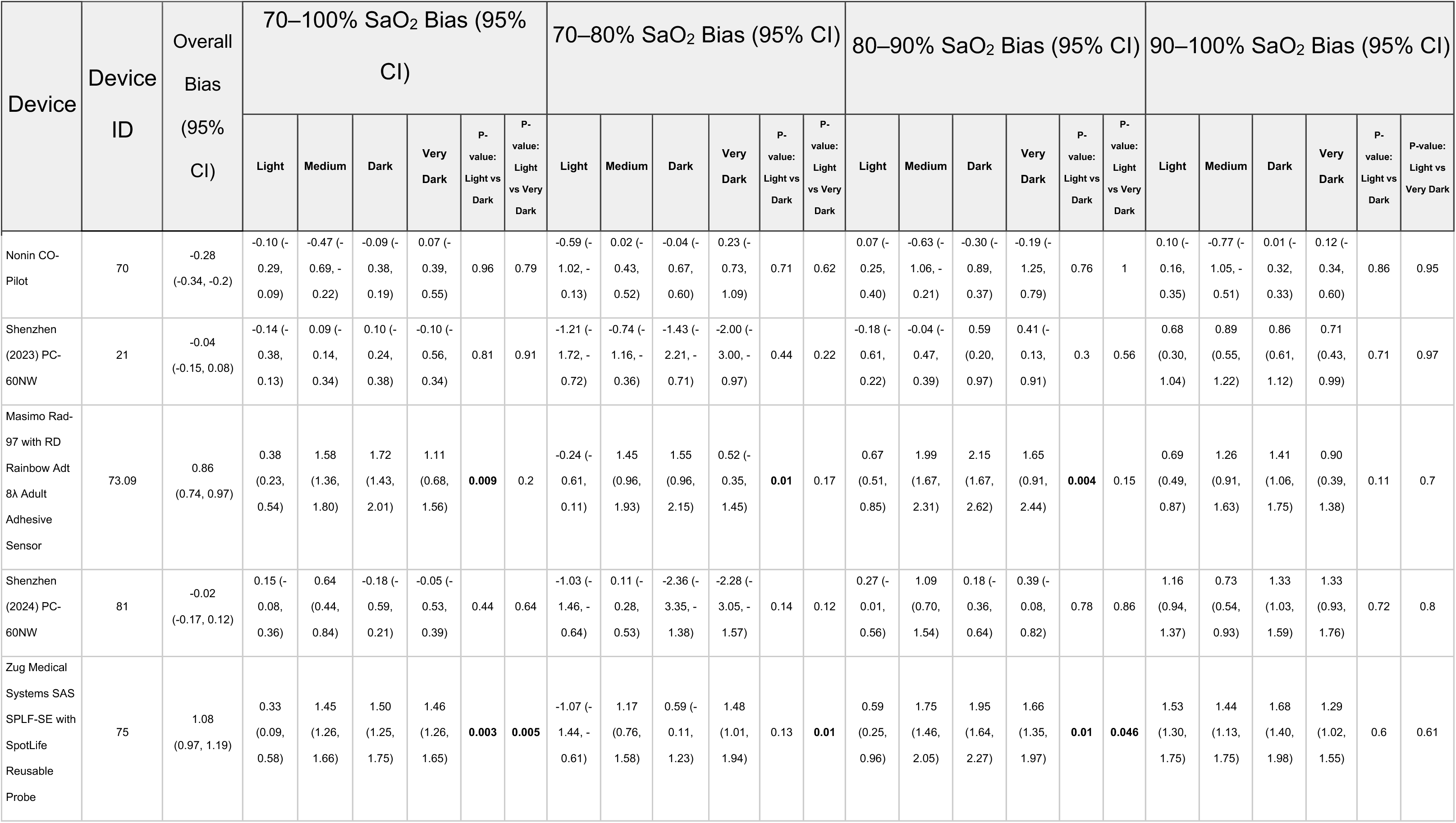
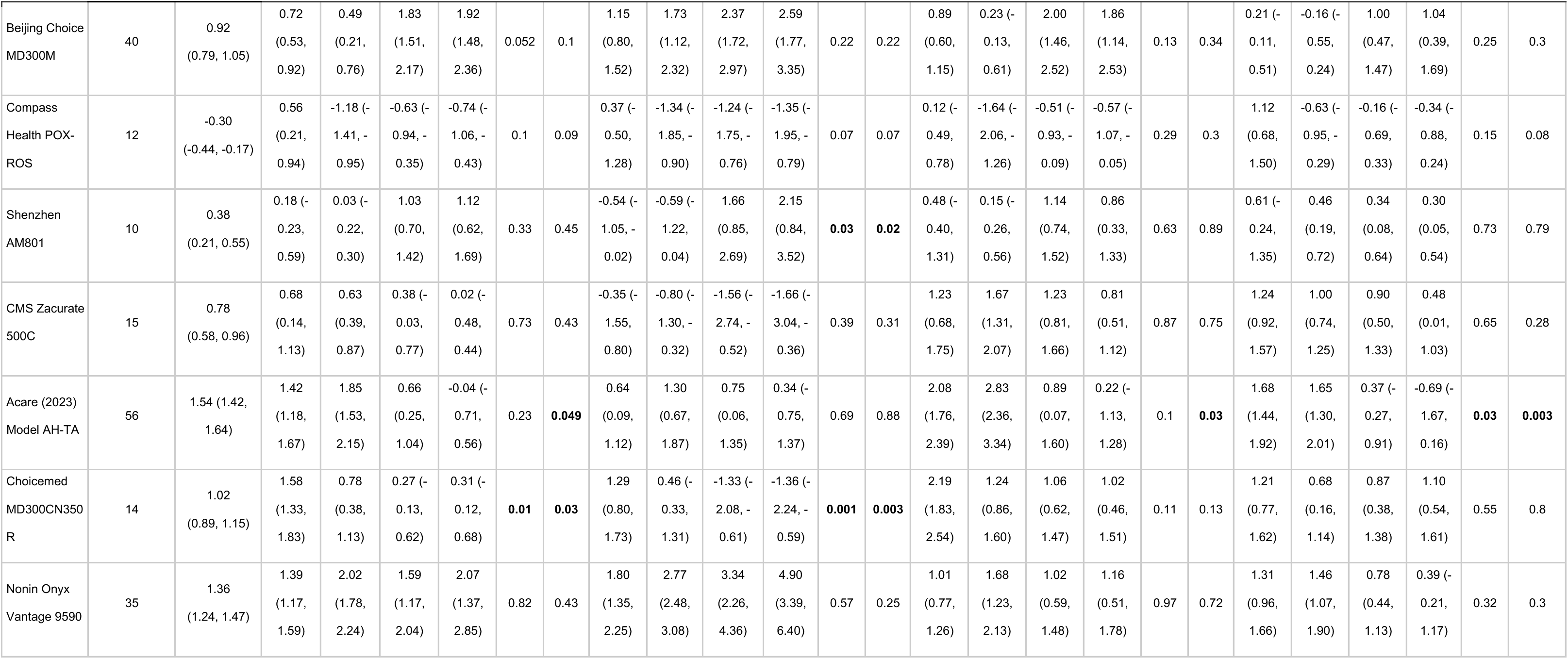
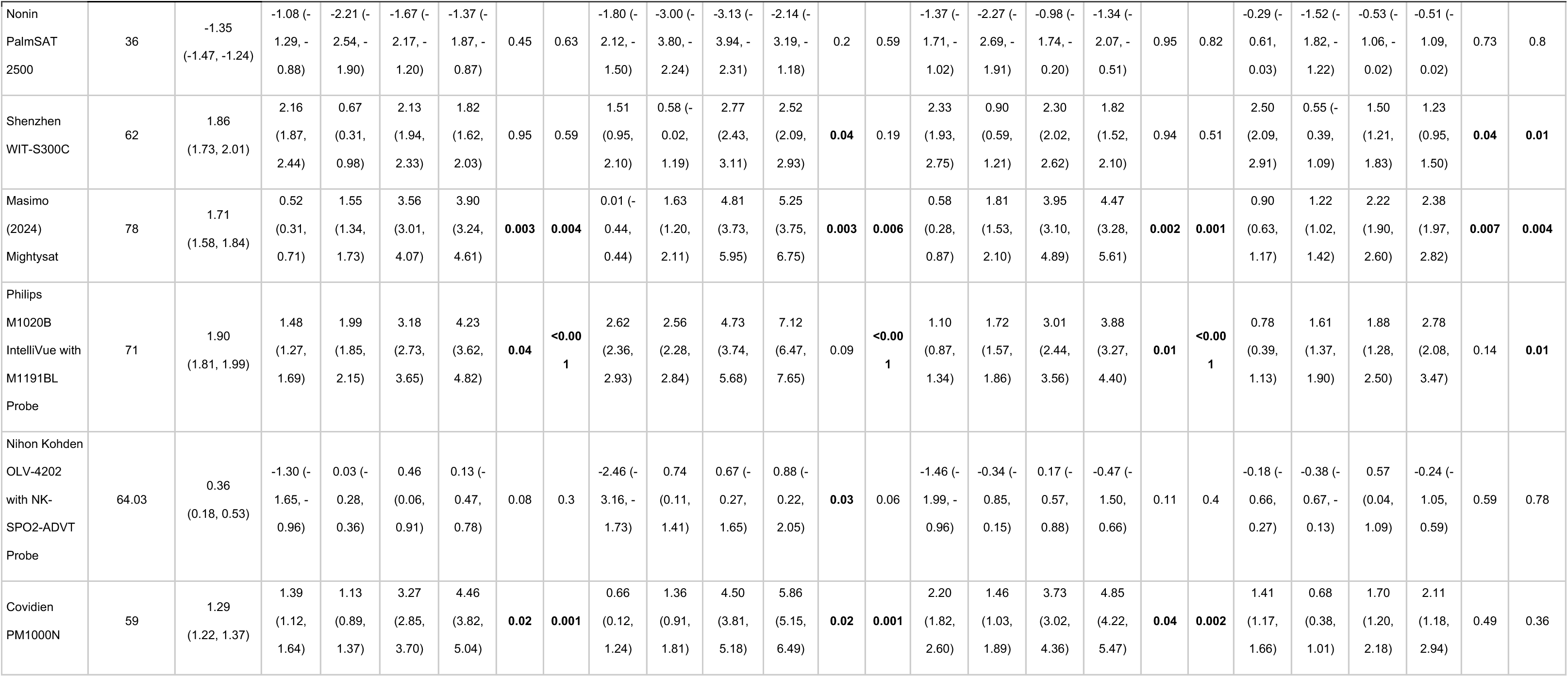
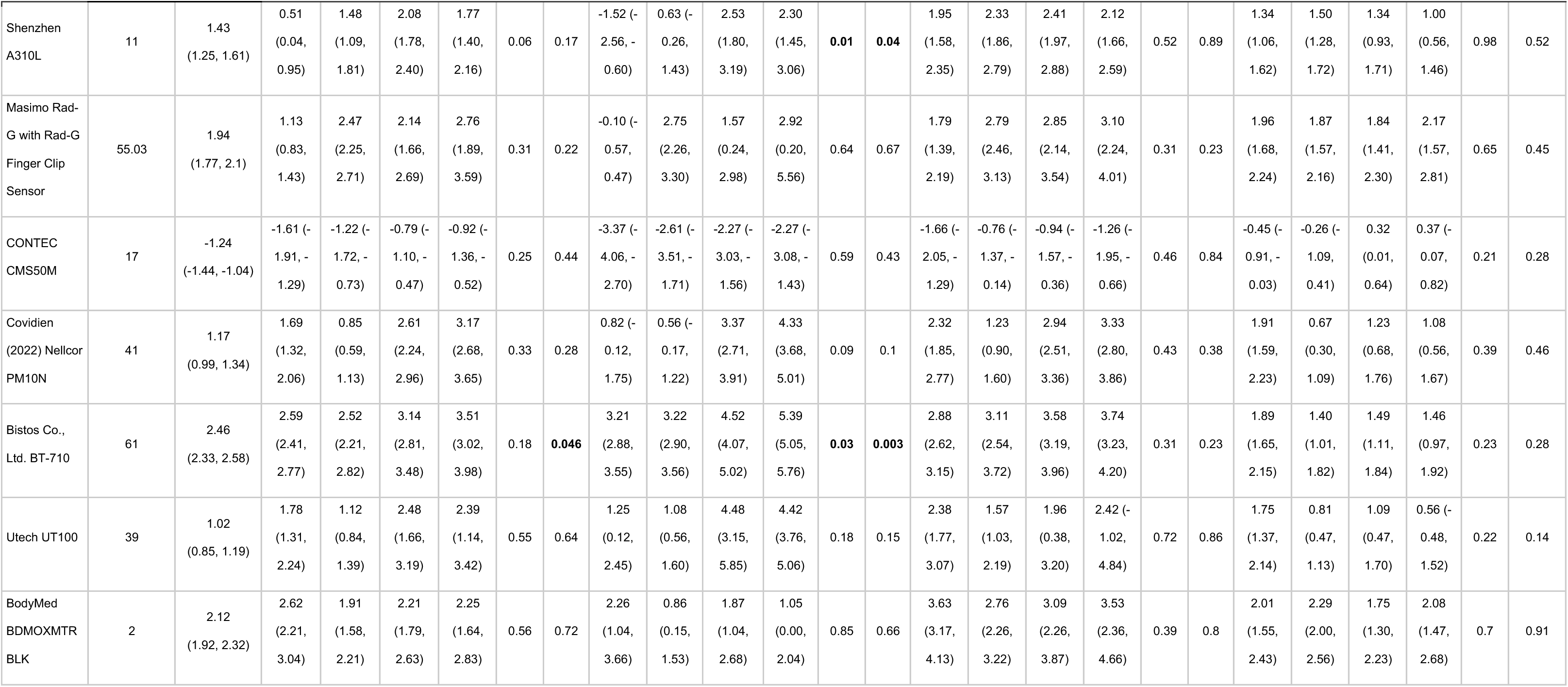
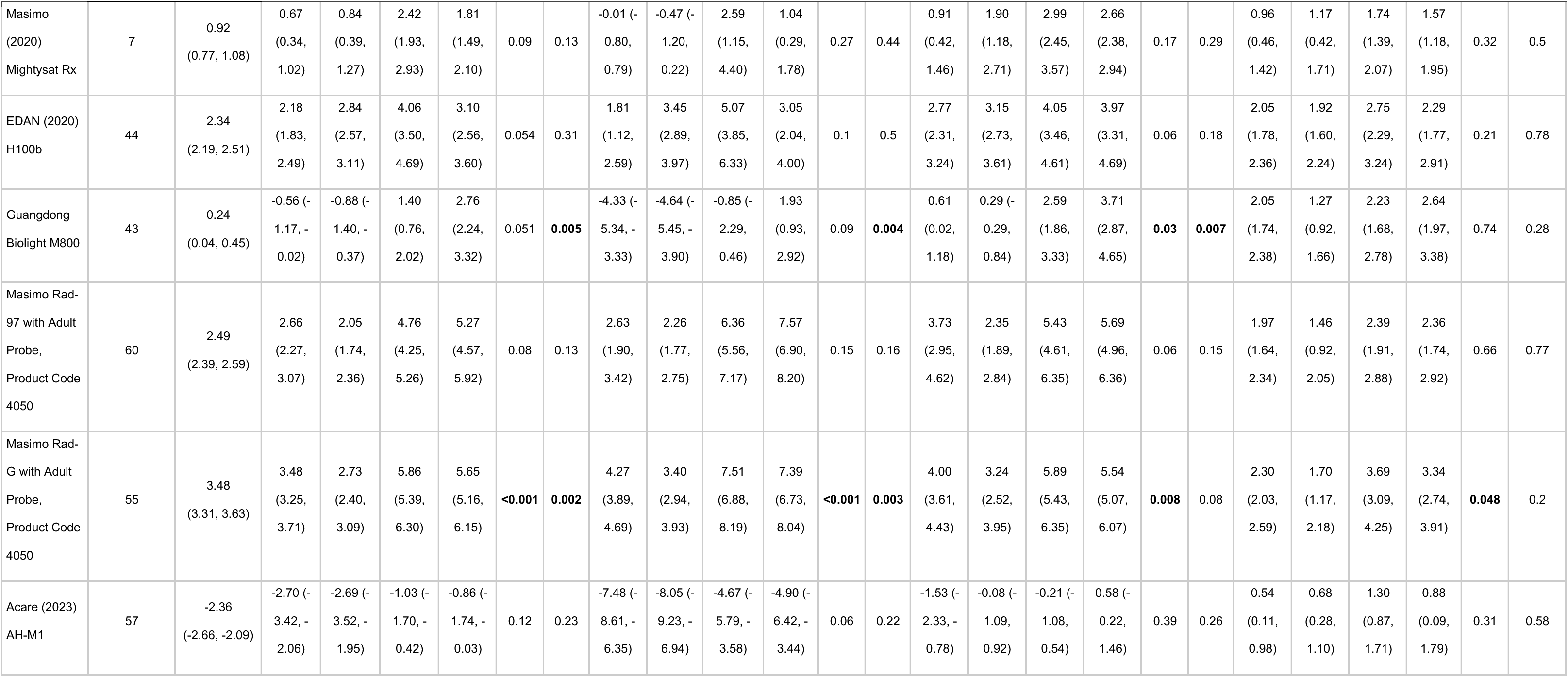
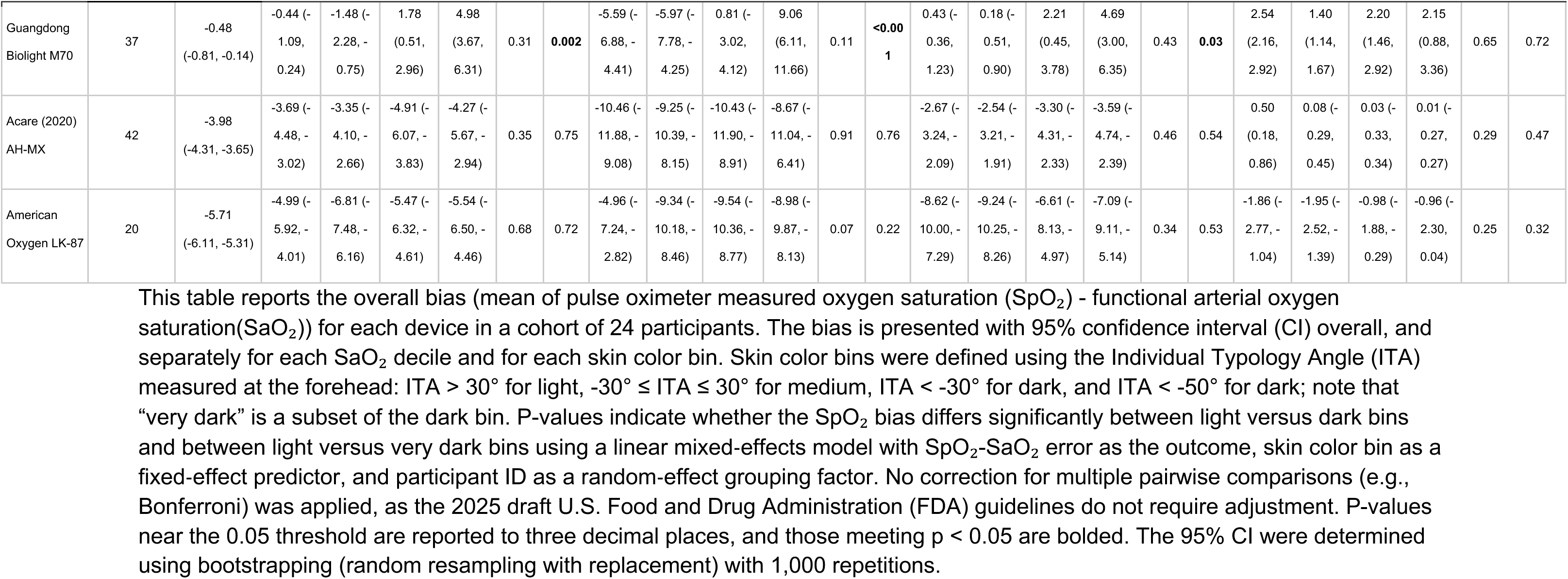
Bias (mean of SpO_2_ minus SaO_2_) at the forehead for 24 participant cohorts.

**Table S8.**
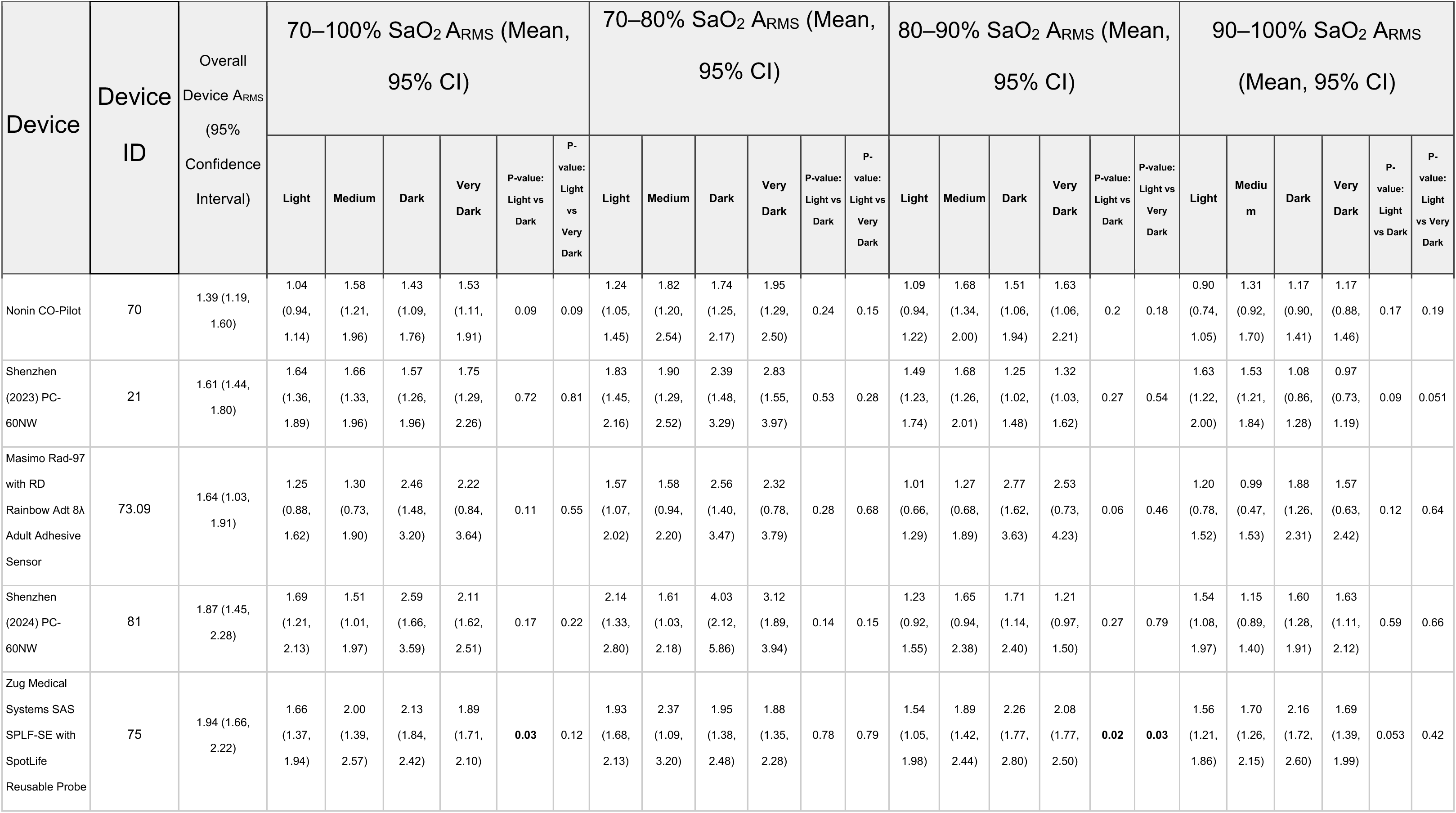
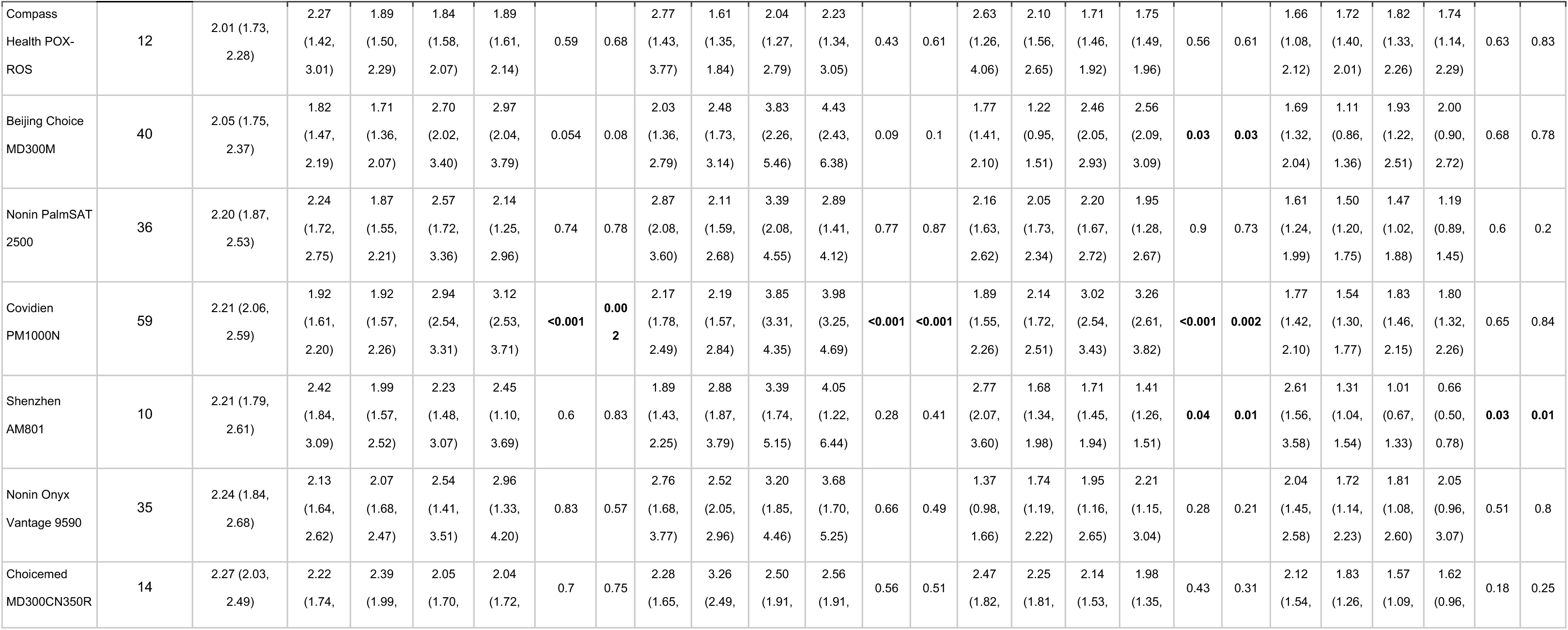
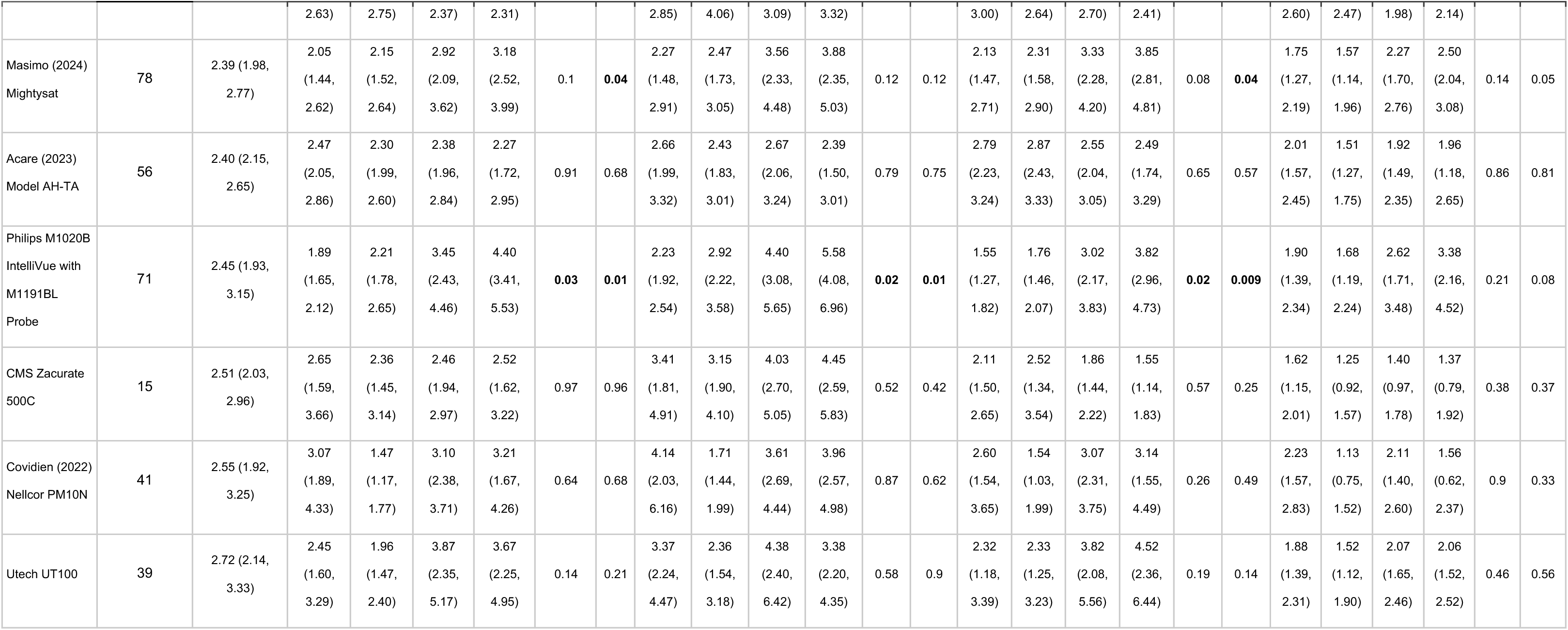
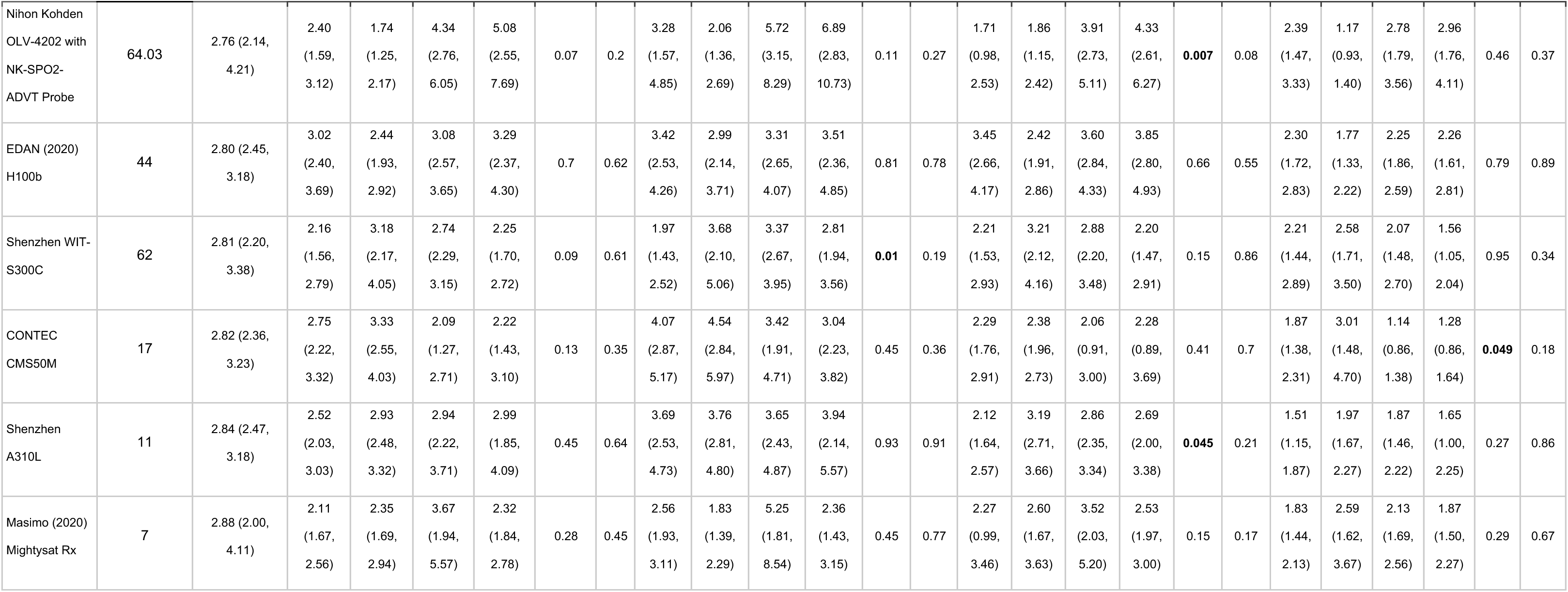
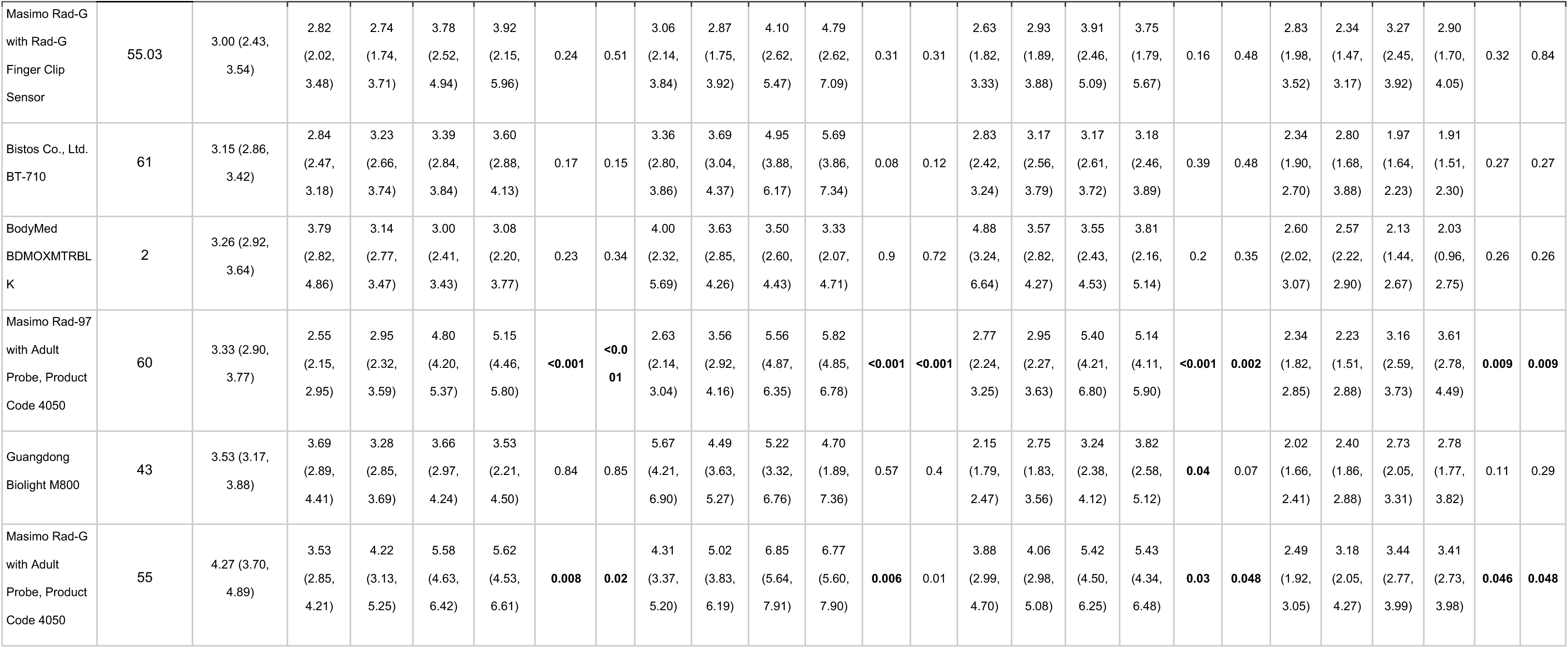
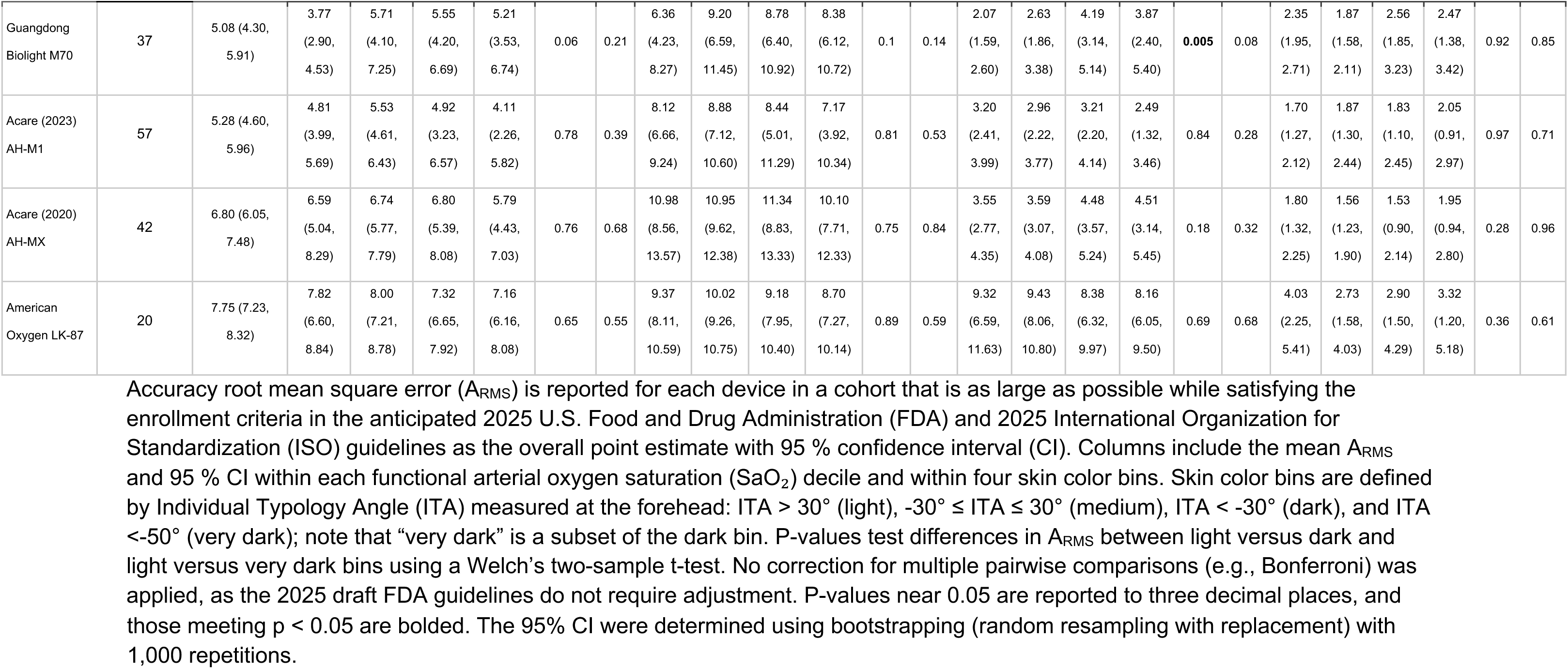
A_RMS_ at the forehead for maximum participant cohorts.

**Table S9.**
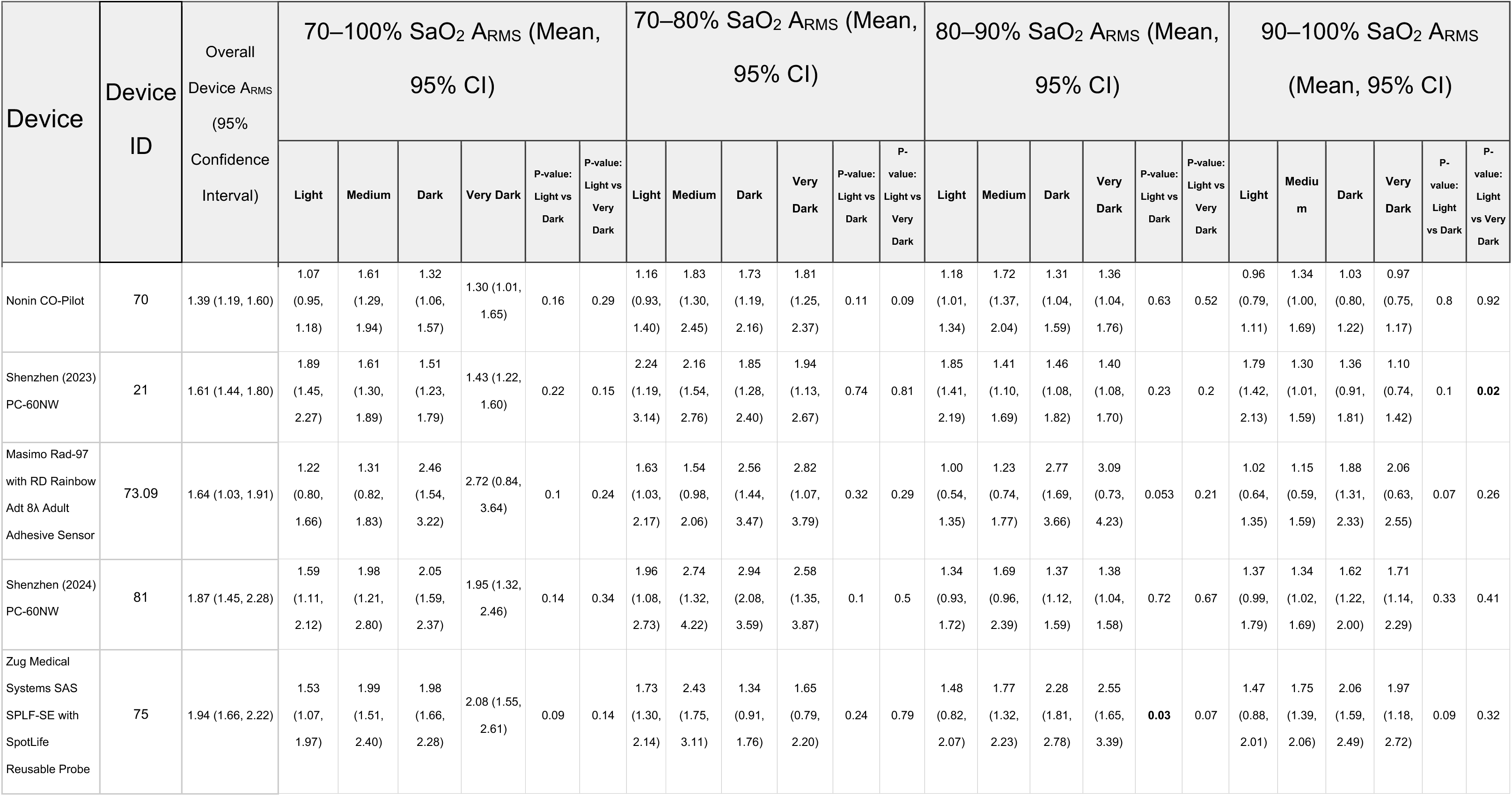
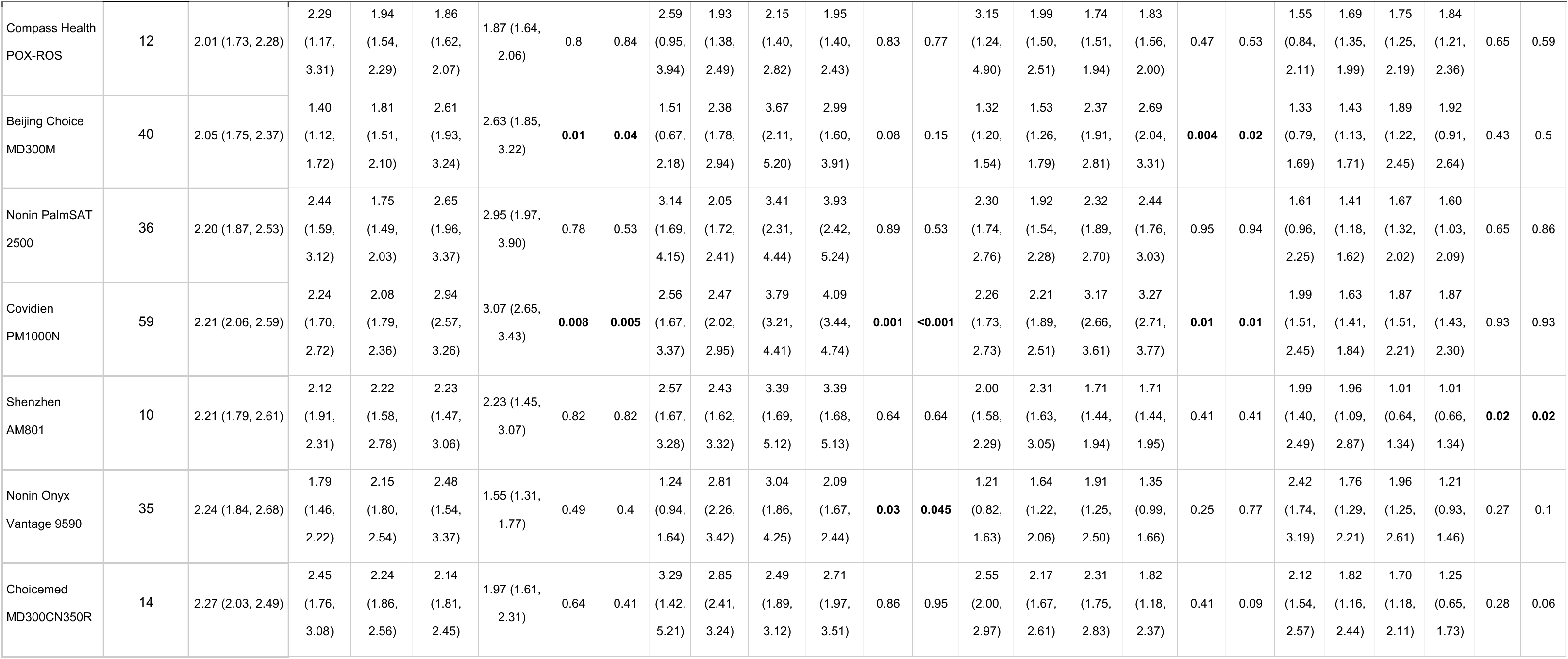
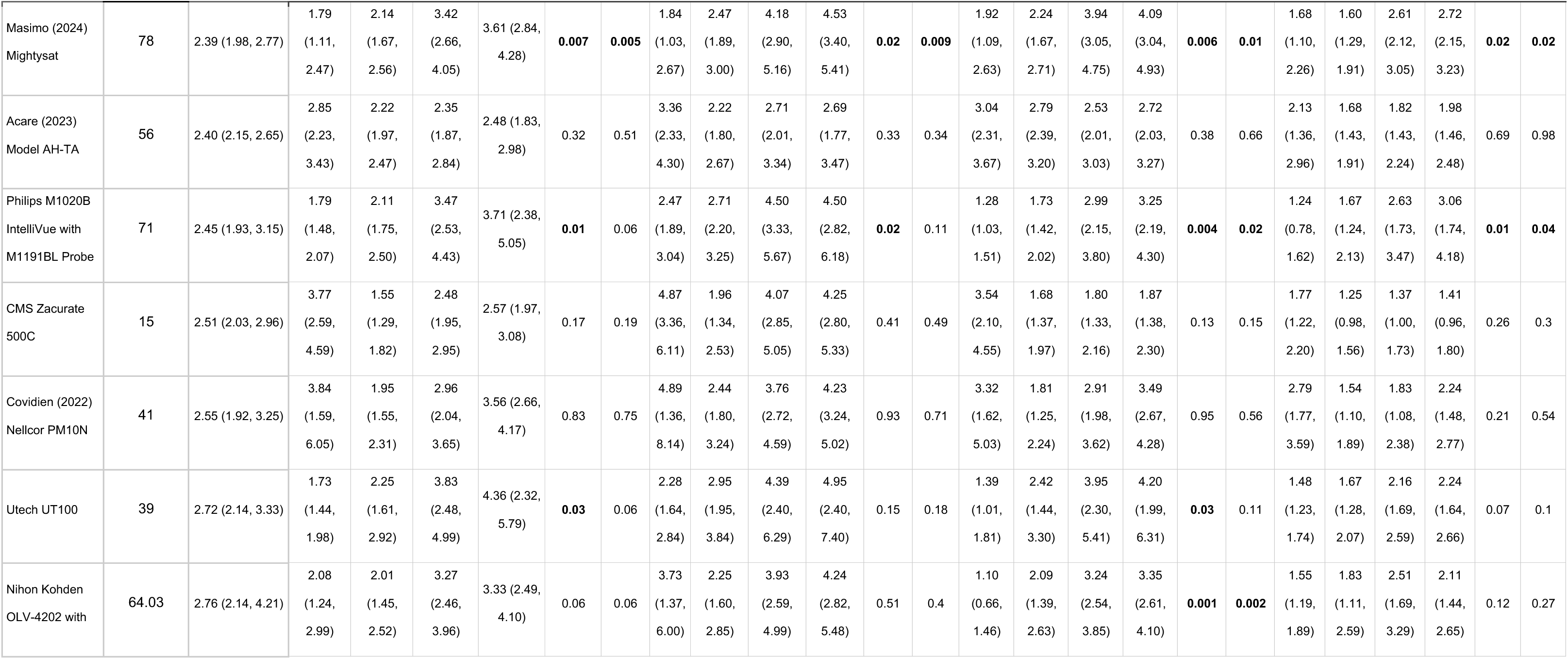
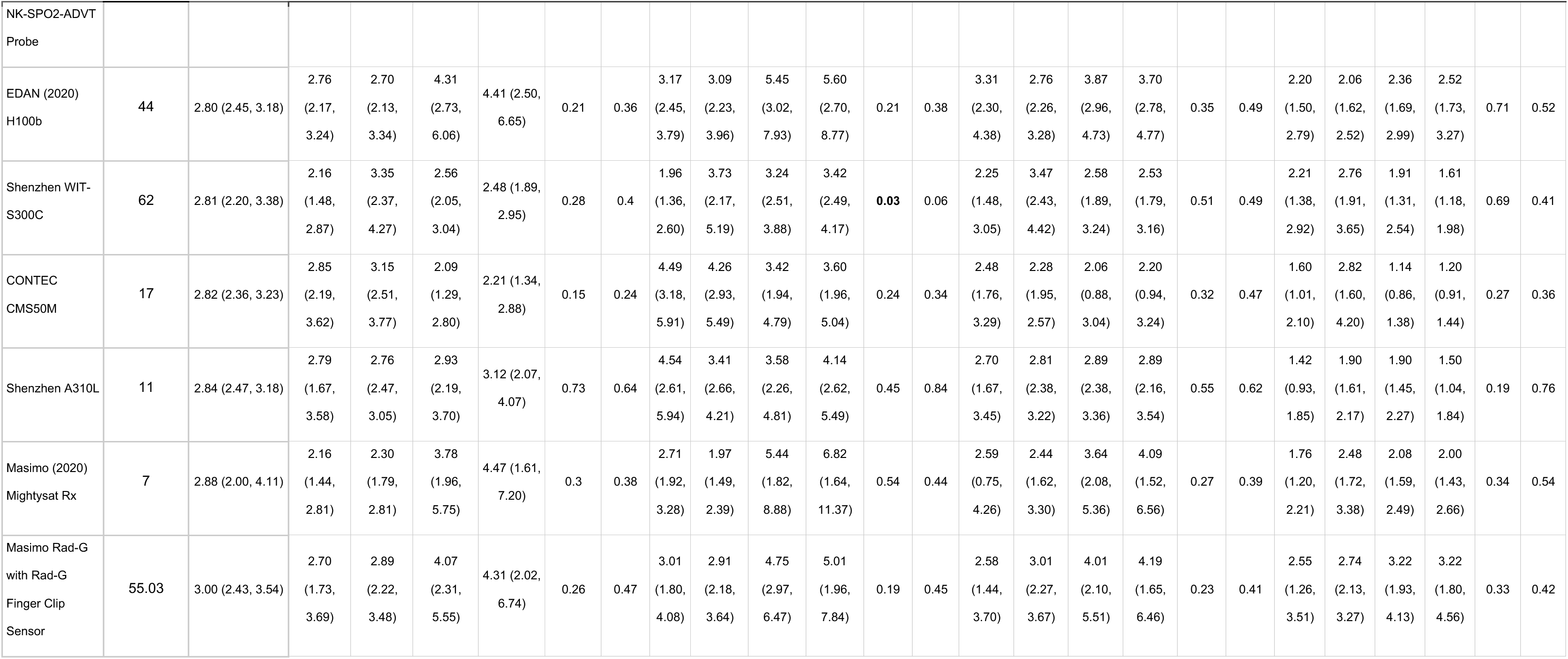
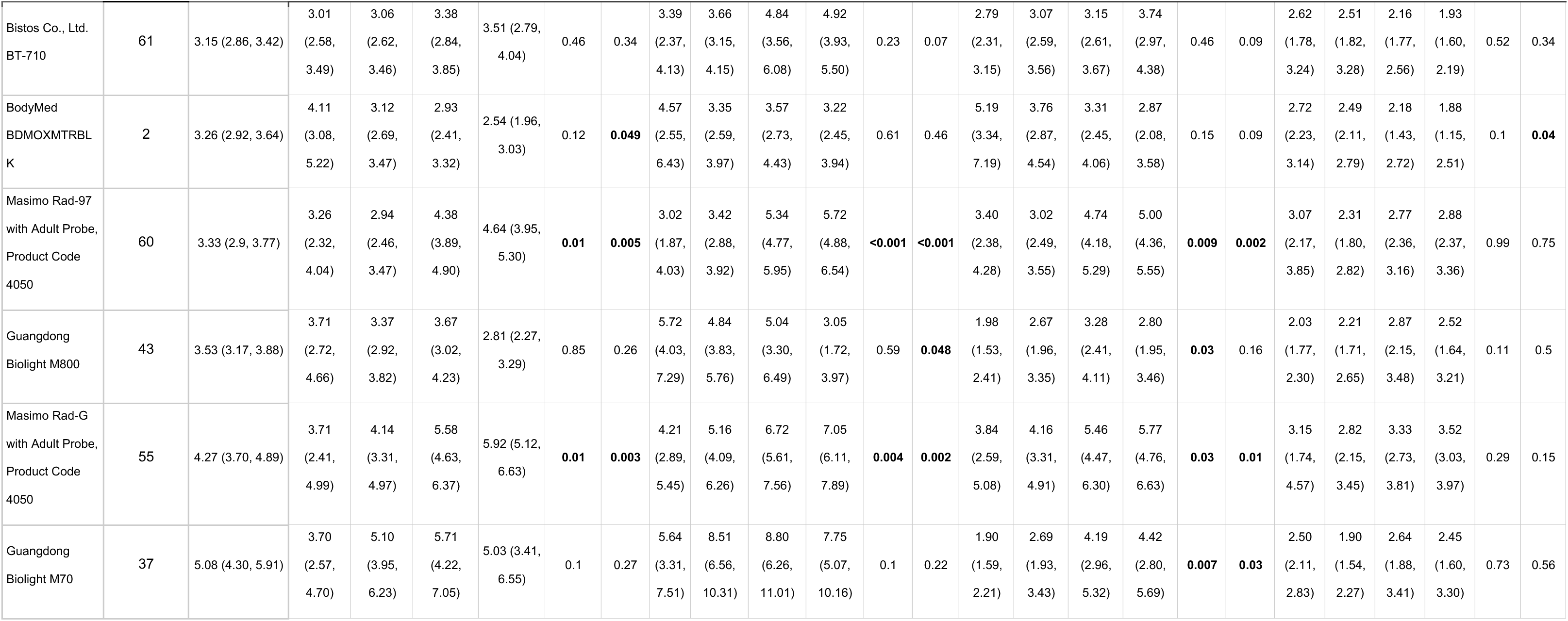
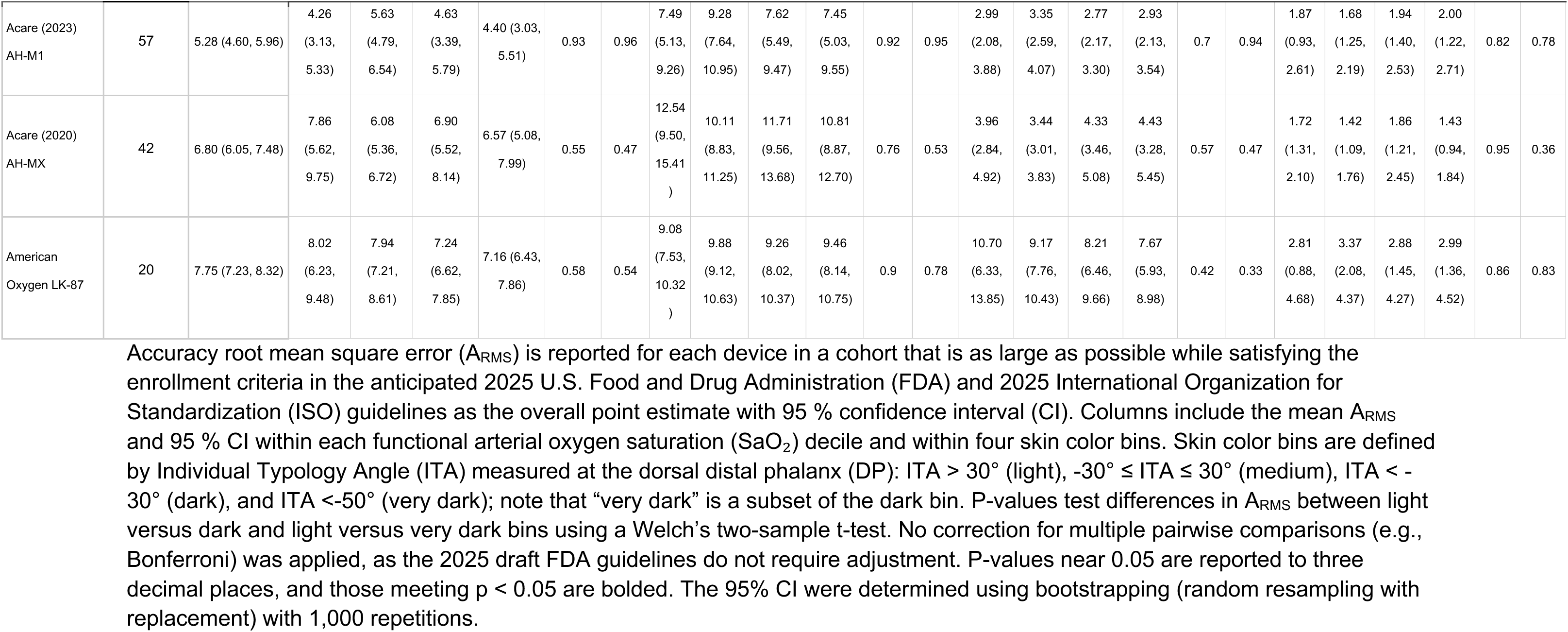
A_RMS_ at the DP for maximum participant cohorts.

| Device | Device ID | Overall Device A<sub>RMS</sub> (95% Confidence Interval) | 70–100% SaO<sub>2</sub> A<sub>RMS</sub> (Mean, 95% CI) |  |  |  |  |  |  | 70–80% SaO<sub>2</sub> A<sub>RMS</sub> (Mean, 95% CI) |  |  |  |  |  |  | 80–90% SaO<sub>2</sub> A<sub>RMS</sub> (Mean, 95% CI) |  |  |  |  |  |  | 90–100% SaO<sub>2</sub> A<sub>RMS</sub> (Mean, 95% CI) |  |  |
| --- | --- | --- | --- | --- | --- | --- | --- | --- | --- | --- | --- | --- | --- | --- | --- | --- | --- | --- | --- | --- | --- | --- | --- | --- | --- | --- |
|  |  |  | Light | Medium | Dark | Very Dark | P-value: Light vs Dark | P-value: Light vs Very Dark | Light | Medium | Dark | Very Dark | P-value: Light vs Dark | P-value: Light vs Very Dark | Light | Medium | Dark | Very Dark | P-value: Light vs Dark | P-value: Light vs Very Dark | Light | Medium | Dark | Very Dark | P-value: Light vs Dark | P-value: Light vs Very Dark |
| Nonin CO-Pilot | 70 | 1.39 (1.19, 1.60) | 1.07 (0.95, 1.18) | 1.61 (1.29, 1.94) | 1.32 (1.06, 1.57) | 1.30 (1.01, 1.65) | 0.16 | 0.29 | 1.16 (0.93, 1.40) | 1.83 (1.30, 2.45) | 1.73 (1.19, 2.16) | 1.81 (1.25, 2.37) | 0.11 | 0.09 | 1.18 (1.01, 1.34) | 1.72 (1.37, 2.04) | 1.31 (1.04, 1.59) | 1.36 (1.04, 1.76) | 0.63 | 0.52 | 0.96 (0.79, 1.11) | 1.34 (1.00, 1.69) | 1.03 (0.80, 1.22) | 0.97 (0.75, 1.17) | 0.8 | 0.92 |
| Shenzhen (2023) PC-60NW | 21 | 1.61 (1.44, 1.80) | 1.89 (1.45, 2.27) | 1.61 (1.30, 1.89) | 1.51 (1.23, 1.79) | 1.43 (1.22, 1.60) | 0.22 | 0.15 | 2.24 (1.19, 3.14) | 2.16 (1.54, 2.76) | 1.85 (1.28, 2.40) | 1.94 (1.13, 2.67) | 0.74 | 0.81 | 1.85 (1.41, 2.19) | 1.41 (1.10, 1.69) | 1.46 (1.08, 1.82) | 1.40 (1.08, 1.70) | 0.23 | 0.2 | 1.79 (1.42, 2.13) | 1.30 (1.01, 1.59) | 1.36 (0.91, 1.81) | 1.10 (0.74, 1.42) | 0.1 | **0.02** |
| Masimo Rad-97 with RD Rainbow Adt 8λ Adult Adhesive Sensor | 73.09 | 1.64 (1.03, 1.91) | 1.22 (0.80, 1.66) | 1.31 (0.82, 1.83) | 2.46 (1.54, 3.22) | 2.72 (0.84, 3.64) | 0.1 | 0.24 | 1.63 (1.03, 2.17) | 1.54 (0.98, 2.06) | 2.56 (1.44, 3.47) | 2.82 (1.07, 3.79) | 0.32 | 0.29 | 1.00 (0.54, 1.35) | 1.23 (0.74, 1.77) | 2.77 (1.69, 3.66) | 3.09 (0.73, 4.23) | 0.053 | 0.21 | 1.02 (0.64, 1.35) | 1.15 (0.59, 1.59) | 1.88 (1.31, 2.33) | 2.06 (0.63, 2.55) | 0.07 | 0.26 |
| Shenzhen (2024) PC-60NW | 81 | 1.87 (1.45, 2.28) | 1.59 (1.11, 2.12) | 1.98 (1.21, 2.80) | 2.05 (1.59, 2.37) | 1.95 (1.32, 2.46) | 0.14 | 0.34 | 1.96 (1.08, 2.73) | 2.74 (1.32, 4.22) | 2.94 (2.08, 3.59) | 2.58 (1.35, 3.87) | 0.1 | 0.5 | 1.34 (0.93, 1.72) | 1.69 (0.96, 2.39) | 1.37 (1.12, 1.59) | 1.38 (1.04, 1.58) | 0.72 | 0.67 | 1.37 (0.99, 1.79) | 1.34 (1.02, 1.69) | 1.62 (1.22, 2.00) | 1.71 (1.14, 2.29) | 0.33 | 0.41 |
| Zug Medical Systems SAS SPLF-SE with SpotLife Reusable Probe | 75 | 1.94 (1.66, 2.22) | 1.53 (1.07, 1.97) | 1.99 (1.51, 2.40) | 1.98 (1.66, 2.28) | 2.08 (1.55, 2.61) | 0.09 | 0.14 | 1.73 (1.30, 2.14) | 2.43 (1.75, 3.11) | 1.34 (0.91, 1.76) | 1.65 (0.79, 2.20) | 0.24 | 0.79 | 1.48 (0.82, 2.07) | 1.77 (1.32, 2.23) | 2.28 (1.81, 2.78) | 2.55 (1.65, 3.39) | **0.03** | 0.07 | 1.47 (0.88, 2.01) | 1.75 (1.39, 2.06) | 2.06 (1.59, 2.49) | 1.97 (1.18, 2.72) | 0.09 | 0.32 |

| Device | Device ID | Overall Device ARMS (95% Confidence Interval) | 70–100% SaO<sub>2</sub> ARMS (Mean, 95% CI) |  |  |  |  |  | 70–80% SaO<sub>2</sub> ARMS (Mean, 95% CI) |  |  |  |  |  | 80–90% SaO<sub>2</sub> ARMS (Mean, 95% CI) |  |  |  |  |  | 90–100% SaO<sub>2</sub> ARMS (Mean, 95% CI) |  |  |  |  |  |
| --- | --- | --- | --- | --- | --- | --- | --- | --- | --- | --- | --- | --- | --- | --- | --- | --- | --- | --- | --- | --- | --- | --- | --- | --- | --- | --- |
|  |  |  | Light | Medium | Dark | Very Dark | P-value: Light vs Dark | P-value: Light vs Very Dark | Light | Medium | Dark | Very Dark | P-value: Light vs Dark | P-value: Light vs Very Dark | Light | Medium | Dark | Very Dark | P-value: Light vs Dark | P-value: Light vs Very Dark | Light | Medium | Dark | Very Dark | P-value: Light vs Dark | P-value: Light vs Very Dark |
| Compass Health POX-ROS | 12 | 2.01 (1.73, 2.28) | 2.29 (1.17, 3.31) | 1.94 (1.54, 2.29) | 1.86 (1.62, 2.07) | 1.87 (1.64, 2.06) | 0.8 | 0.84 | 2.59 (0.95, 3.94) | 1.93 (1.38, 2.49) | 2.15 (1.40, 2.82) | 1.95 (1.40, 2.43) | 0.83 | 0.77 | 3.15 (1.24, 4.90) | 1.99 (1.50, 2.51) | 1.74 (1.51, 1.94) | 1.83 (1.56, 2.00) | 0.47 | 0.53 | 1.55 (0.84, 2.11) | 1.69 (1.35, 1.99) | 1.75 (1.25, 2.19) | 1.84 (1.21, 2.36) | 0.65 | 0.59 |
| Beijing Choice MD300M | 40 | 2.05 (1.75, 2.37) | 1.40 (1.12, 1.72) | 1.81 (1.51, 2.10) | 2.61 (1.93, 3.24) | 2.63 (1.85, 3.22) | **0.01** | **0.04** | 1.51 (0.67, 2.18) | 2.38 (1.78, 2.94) | 3.67 (2.11, 5.20) | 2.99 (1.60, 3.91) | 0.08 | 0.15 | 1.32 (1.20, 1.54) | 1.53 (1.26, 1.79) | 2.37 (1.91, 2.81) | 2.69 (2.04, 3.31) | **0.004** | **0.02** | 1.33 (0.79, 1.69) | 1.43 (1.13, 1.71) | 1.89 (1.22, 2.45) | 1.92 (0.91, 2.64) | 0.43 | 0.5 |
| Nonin PalmSAT 2500 | 36 | 2.20 (1.87, 2.53) | 2.44 (1.59, 3.12) | 1.75 (1.49, 2.03) | 2.65 (1.96, 3.37) | 2.95 (1.97, 3.90) | 0.78 | 0.53 | 3.14 (1.69, 4.15) | 2.05 (1.72, 2.41) | 3.41 (2.31, 4.44) | 3.93 (2.42, 5.24) | 0.89 | 0.53 | 2.30 (1.74, 2.76) | 1.92 (1.54, 2.28) | 2.32 (1.89, 2.70) | 2.44 (1.76, 3.03) | 0.95 | 0.94 | 1.61 (0.96, 2.25) | 1.41 (1.18, 1.62) | 1.67 (1.32, 2.02) | 1.60 (1.03, 2.09) | 0.65 | 0.86 |
| Covidien PM1000N | 59 | 2.21 (2.06, 2.59) | 2.24 (1.70, 2.72) | 2.08 (1.79, 2.36) | 2.94 (2.57, 3.26) | 3.07 (2.65, 3.43) | **0.008** | **0.005** | 2.56 (1.67, 3.37) | 2.47 (2.02, 2.95) | 3.79 (3.21, 4.41) | 4.09 (3.44, 4.74) | **0.001** | **<0.001** | 2.26 (1.73, 2.73) | 2.21 (1.89, 2.51) | 3.17 (2.66, 3.61) | 3.27 (2.71, 3.77) | **0.01** | **0.01** | 1.99 (1.51, 2.45) | 1.63 (1.41, 1.84) | 1.87 (1.51, 2.21) | 1.87 (1.43, 2.30) | 0.93 | 0.93 |
| Shenzhen AM801 | 10 | 2.21 (1.79, 2.61) | 2.12 (1.91, 2.31) | 2.22 (1.58, 2.78) | 2.23 (1.47, 3.06) | 2.23 (1.45, 3.07) | 0.82 | 0.82 | 2.57 (1.67, 3.28) | 2.43 (1.62, 3.32) | 3.39 (1.69, 5.12) | 3.39 (1.68, 5.13) | 0.64 | 0.64 | 2.00 (1.58, 2.29) | 2.31 (1.63, 3.05) | 1.71 (1.44, 1.94) | 1.71 (1.44, 1.95) | 0.41 | 0.41 | 1.99 (1.40, 2.49) | 1.96 (1.09, 2.87) | 1.01 (0.64, 1.34) | 1.01 (0.66, 1.34) | **0.02** | **0.02** |
| Nonin Onyx Vantage 9590 | 35 | 2.24 (1.84, 2.68) | 1.79 (1.46, 2.22) | 2.15 (1.80, 2.54) | 2.48 (1.54, 3.37) | 1.55 (1.31, 1.77) | 0.49 | 0.4 | 1.24 (0.94, 1.64) | 2.81 (2.26, 3.42) | 3.04 (1.86, 4.25) | 2.09 (1.67, 2.44) | **0.03** | **0.045** | 1.21 (0.82, 1.63) | 1.64 (1.22, 2.06) | 1.91 (1.25, 2.50) | 1.35 (0.99, 1.66) | 0.25 | 0.77 | 2.42 (1.74, 3.19) | 1.76 (1.29, 2.21) | 1.96 (1.25, 2.61) | 1.21 (0.93, 1.46) | 0.27 | 0.1 |
| Choicemed MD300CN350R | 14 | 2.27 (2.03, 2.49) | 2.45 (1.76, 3.08) | 2.24 (1.86, 2.56) | 2.14 (1.81, 2.45) | 1.97 (1.61, 2.31) | 0.64 | 0.41 | 3.29 (1.42, 5.21) | 2.85 (2.41, 3.24) | 2.49 (1.89, 3.12) | 2.71 (1.97, 3.51) | 0.86 | 0.95 | 2.55 (2.00, 2.97) | 2.17 (1.67, 2.61) | 2.31 (1.75, 2.83) | 1.82 (1.18, 2.37) | 0.41 | 0.09 | 2.12 (1.54, 2.57) | 1.82 (1.16, 2.44) | 1.70 (1.18, 2.11) | 1.25 (0.65, 1.73) | 0.28 | 0.06 |
|  |  |  | Light | Medium | Dark | Very Dark | P-value: Light vs Dark | P-value: Light vs Very Dark | Light | Medium | Dark | Very Dark | P-value: Light vs Dark | P-value: Light vs Very Dark | Light | Medium | Dark | Very Dark | P-value: Light vs Dark | P-value: Light vs Very Dark | Light | Medium | Dark | Very Dark | P-value: Light vs Dark | P-value: Light vs Very Dark |
| Masimo (2024) Mightysat | 78 | 2.39 (1.98, 2.77) | 1.79 (1.11, 2.47) | 2.14 (1.67, 2.56) | 3.42 (2.66, 4.05) | 3.61 (2.84, 4.28) | 0.007 | 0.005 | 1.84 (1.03, 2.67) | 2.47 (1.89, 3.00) | 4.18 (2.90, 5.16) | 4.53 (3.40, 5.41) | 0.02 | 0.009 | 1.92 (1.09, 2.63) | 2.24 (1.67, 2.71) | 3.94 (3.05, 4.75) | 4.09 (3.04, 4.93) | 0.006 | 0.01 | 1.68 (1.10, 2.26) | 1.60 (1.29, 1.91) | 2.61 (2.12, 3.05) | 2.72 (2.15, 3.23) | 0.02 | 0.02 |
| Acare (2023) Model AH-TA | 56 | 2.40 (2.15, 2.65) | 2.85 (2.23, 3.43) | 2.22 (1.97, 2.47) | 2.35 (1.87, 2.84) | 2.48 (1.83, 2.98) | 0.32 | 0.51 | 3.36 (2.33, 4.30) | 2.22 (1.80, 2.67) | 2.71 (2.01, 3.34) | 2.69 (1.77, 3.47) | 0.33 | 0.34 | 3.04 (2.31, 3.67) | 2.79 (2.39, 3.20) | 2.53 (2.01, 3.03) | 2.72 (2.03, 3.27) | 0.38 | 0.66 | 2.13 (1.36, 2.96) | 1.68 (1.43, 1.91) | 1.82 (1.43, 2.24) | 1.98 (1.46, 2.48) | 0.69 | 0.98 |
| Philips M1020B IntelliVue with M1191BL Probe | 71 | 2.45 (1.93, 3.15) | 1.79 (1.48, 2.07) | 2.11 (1.75, 2.50) | 3.47 (2.53, 4.43) | 3.71 (2.38, 5.05) | 0.01 | 0.06 | 2.47 (1.89, 3.04) | 2.71 (2.20, 3.25) | 4.50 (3.33, 5.67) | 4.50 (2.82, 6.18) | 0.02 | 0.11 | 1.28 (1.03, 1.51) | 1.73 (1.42, 2.02) | 2.99 (2.15, 3.80) | 3.25 (2.19, 4.30) | 0.004 | 0.02 | 1.24 (0.78, 1.62) | 1.67 (1.24, 2.13) | 2.63 (1.73, 3.47) | 3.06 (1.74, 4.18) | 0.01 | 0.04 |
| CMS Zacurate 500C | 15 | 2.51 (2.03, 2.96) | 3.77 (2.59, 4.59) | 1.55 (1.29, 1.82) | 2.48 (1.95, 2.95) | 2.57 (1.97, 3.08) | 0.17 | 0.19 | 4.87 (3.36, 6.11) | 1.96 (1.34, 2.53) | 4.07 (2.85, 5.05) | 4.25 (2.80, 5.33) | 0.41 | 0.49 | 3.54 (2.10, 4.55) | 1.68 (1.37, 1.97) | 1.80 (1.33, 2.16) | 1.87 (1.38, 2.30) | 0.13 | 0.15 | 1.77 (1.22, 2.20) | 1.25 (0.98, 1.56) | 1.37 (1.00, 1.73) | 1.41 (0.96, 1.80) | 0.26 | 0.3 |
| Covidien (2022) Nellcor PM10N | 41 | 2.55 (1.92, 3.25) | 3.84 (1.59, 6.05) | 1.95 (1.55, 2.31) | 2.96 (2.04, 3.65) | 3.56 (2.66, 4.17) | 0.83 | 0.75 | 4.89 (1.36, 8.14) | 2.44 (1.80, 3.24) | 3.76 (2.72, 4.59) | 4.23 (3.24, 5.02) | 0.93 | 0.71 | 3.32 (1.62, 5.03) | 1.81 (1.25, 2.24) | 2.91 (1.98, 3.62) | 3.49 (2.67, 4.28) | 0.95 | 0.56 | 2.79 (1.77, 3.59) | 1.54 (1.10, 1.89) | 1.83 (1.08, 2.38) | 2.24 (1.48, 2.77) | 0.21 | 0.54 |
| Utech UT100 | 39 | 2.72 (2.14, 3.33) | 1.73 (1.44, 1.98) | 2.25 (1.61, 2.92) | 3.83 (2.48, 4.99) | 4.36 (2.32, 5.79) | 0.03 | 0.06 | 2.28 (1.64, 2.84) | 2.95 (1.95, 3.84) | 4.39 (2.40, 6.29) | 4.95 (2.40, 7.40) | 0.15 | 0.18 | 1.39 (1.01, 1.81) | 2.42 (1.44, 3.30) | 3.95 (2.30, 5.41) | 4.20 (1.99, 6.31) | 0.03 | 0.11 | 1.48 (1.23, 1.74) | 1.67 (1.28, 2.07) | 2.16 (1.69, 2.59) | 2.24 (1.64, 2.66) | 0.07 | 0.1 |
| Nihon Kohden OLV-4202 with | 64.03 | 2.76 (2.14, 4.21) | 2.08 (1.24, 2.99) | 2.01 (1.45, 2.52) | 3.27 (2.46, 3.96) | 3.33 (2.49, 4.10) | 0.06 | 0.06 | 3.73 (1.37, 6.00) | 2.25 (1.60, 2.85) | 3.93 (2.59, 4.99) | 4.24 (2.82, 5.48) | 0.51 | 0.4 | 1.10 (0.66, 1.46) | 2.09 (1.39, 2.63) | 3.24 (2.54, 3.85) | 3.35 (2.61, 4.10) | 0.001 | 0.002 | 1.55 (1.19, 1.89) | 1.83 (1.11, 2.59) | 2.51 (1.69, 3.29) | 2.11 (1.44, 2.65) | 0.12 | 0.27 |

| Device | Device ID | Overall Device A<sub>RMS</sub> (95% Confidence Interval) | 70–100% SaO<sub>2</sub> A<sub>RMS</sub> (Mean, 95% CI) |  |  |  |  |  | 70–80% SaO<sub>2</sub> A<sub>RMS</sub> (Mean, 95% CI) |  |  |  |  |  | 80–90% SaO<sub>2</sub> A<sub>RMS</sub> (Mean, 95% CI) |  |  |  |  |  | 90–100% SaO<sub>2</sub> A<sub>RMS</sub> (Mean, 95% CI) |  |  |  |  |  |
| --- | --- | --- | --- | --- | --- | --- | --- | --- | --- | --- | --- | --- | --- | --- | --- | --- | --- | --- | --- | --- | --- | --- | --- | --- | --- | --- |
|  |  |  | Light | Medium | Dark | Very Dark | P-value: Light vs Dark | P-value: Light vs Very Dark | Light | Medium | Dark | Very Dark | P-value: Light vs Dark | P-value: Light vs Very Dark | Light | Medium | Dark | Very Dark | P-value: Light vs Dark | P-value: Light vs Very Dark | Light | Medium | Dark | Very Dark | P-value: Light vs Dark | P-value: Light vs Very Dark |
| NK-SPO2-ADVT Probe |  |  |  |  |  |  |  |  |  |  |  |  |  |  |  |  |  |  |  |  |  |  |  |  |  |  |
| EDAN (2020) H100b | 44 | 2.80 (2.45, 3.18) | 2.76 (2.17, 3.24) | 2.70 (2.13, 3.34) | 4.31 (2.73, 6.06) | 4.41 (2.50, 6.65) | 0.21 | 0.36 | 3.17 (2.45, 3.79) | 3.09 (2.23, 3.96) | 5.45 (3.02, 7.93) | 5.60 (2.70, 8.77) | 0.21 | 0.38 | 3.31 (2.30, 4.38) | 2.76 (2.26, 3.28) | 3.87 (2.96, 4.73) | 3.70 (2.78, 4.77) | 0.35 | 0.49 | 2.20 (1.50, 2.79) | 2.06 (1.62, 2.52) | 2.36 (1.69, 2.99) | 2.52 (1.73, 3.27) | 0.71 | 0.52 |
| Shenzhen WIT-S300C | 62 | 2.81 (2.20, 3.38) | 2.16 (1.48, 2.87) | 3.35 (2.37, 4.27) | 2.56 (2.05, 3.04) | 2.48 (1.89, 2.95) | 0.28 | 0.4 | 1.96 (1.36, 2.60) | 3.73 (2.17, 5.19) | 3.24 (2.51, 3.88) | 3.42 (2.49, 4.17) | 0.03 | 0.06 | 2.25 (1.48, 3.05) | 3.47 (2.43, 4.42) | 2.58 (1.89, 3.24) | 2.53 (1.79, 3.16) | 0.51 | 0.49 | 2.21 (1.38, 2.92) | 2.76 (1.91, 3.65) | 1.91 (1.31, 2.54) | 1.61 (1.18, 1.98) | 0.69 | 0.41 |
| CONTEC CMS50M | 17 | 2.82 (2.36, 3.23) | 2.85 (2.19, 3.62) | 3.15 (2.51, 3.77) | 2.09 (1.29, 2.80) | 2.21 (1.34, 2.88) | 0.15 | 0.24 | 4.49 (3.18, 5.91) | 4.26 (2.93, 5.49) | 3.42 (1.94, 4.79) | 3.60 (1.96, 5.04) | 0.24 | 0.34 | 2.48 (1.76, 3.29) | 2.28 (1.95, 2.57) | 2.06 (0.88, 3.04) | 2.20 (0.94, 3.24) | 0.32 | 0.47 | 1.60 (1.01, 2.10) | 2.82 (1.60, 4.20) | 1.14 (0.86, 1.38) | 1.20 (0.91, 1.44) | 0.27 | 0.36 |
| Shenzhen A310L | 11 | 2.84 (2.47, 3.18) | 2.79 (1.67, 3.58) | 2.76 (2.47, 3.05) | 2.93 (2.19, 3.70) | 3.12 (2.07, 4.07) | 0.73 | 0.64 | 4.54 (2.61, 5.94) | 3.41 (2.66, 4.21) | 3.58 (2.26, 4.81) | 4.14 (2.62, 5.49) | 0.45 | 0.84 | 2.70 (1.67, 3.45) | 2.81 (2.38, 3.22) | 2.89 (2.38, 3.36) | 2.89 (2.16, 3.54) | 0.55 | 0.62 | 1.42 (0.93, 1.85) | 1.90 (1.61, 2.17) | 1.90 (1.45, 2.27) | 1.50 (1.04, 1.84) | 0.19 | 0.76 |
| Masimo (2020) Mightysat Rx | 7 | 2.88 (2.00, 4.11) | 2.16 (1.44, 2.81) | 2.30 (1.79, 2.81) | 3.78 (1.96, 5.75) | 4.47 (1.61, 7.20) | 0.3 | 0.38 | 2.71 (1.92, 3.28) | 1.97 (1.49, 2.39) | 5.44 (1.82, 8.88) | 6.82 (1.64, 11.37) | 0.54 | 0.44 | 2.59 (0.75, 4.26) | 2.44 (1.62, 3.30) | 3.64 (2.08, 5.36) | 4.09 (1.52, 6.56) | 0.27 | 0.39 | 1.76 (1.20, 2.21) | 2.48 (1.72, 3.38) | 2.08 (1.59, 2.49) | 2.00 (1.43, 2.66) | 0.34 | 0.54 |
| Masimo Rad-G with Rad-G Finger Clip Sensor | 55.03 | 3.00 (2.43, 3.54) | 2.70 (1.73, 3.69) | 2.89 (2.22, 3.48) | 4.07 (2.31, 5.55) | 4.31 (2.02, 6.74) | 0.26 | 0.47 | 3.01 (1.80, 4.08) | 2.91 (2.18, 3.64) | 4.75 (2.97, 6.47) | 5.01 (1.96, 7.84) | 0.19 | 0.45 | 2.58 (1.44, 3.70) | 3.01 (2.27, 3.67) | 4.01 (2.10, 5.51) | 4.19 (1.65, 6.46) | 0.23 | 0.41 | 2.55 (1.26, 3.51) | 2.74 (2.13, 3.27) | 3.22 (1.93, 4.13) | 3.22 (1.80, 4.56) | 0.33 | 0.42 |
|  |  |  | Light | Medium | Dark | Very Dark | P-value: Light vs Dark | P-value: Light vs Very Dark | Light | Medium | Dark | Very Dark | P-value: Light vs Dark | P-value: Light vs Very Dark | Light | Medium | Dark | Very Dark | P-value: Light vs Dark | P-value: Light vs Very Dark | Light | Medium | Dark | Very Dark | P-value: Light vs Dark | P-value: Light vs Very Dark |
| Bistos Co., Ltd. BT-710 | 61 | 3.15 (2.86, 3.42) | 3.01 (2.58, 3.49) | 3.06 (2.62, 3.46) | 3.38 (2.84, 3.85) | 3.51 (2.79, 4.04) | 0.46 | 0.34 | 3.39 (2.37, 4.13) | 3.66 (3.15, 4.15) | 4.84 (3.56, 6.08) | 4.92 (3.93, 5.50) | 0.23 | 0.07 | 2.79 (2.31, 3.15) | 3.07 (2.59, 3.56) | 3.15 (2.61, 3.67) | 3.74 (2.97, 4.38) | 0.46 | 0.09 | 2.62 (1.78, 3.24) | 2.51 (1.82, 3.28) | 2.16 (1.77, 2.56) | 1.93 (1.60, 2.19) | 0.52 | 0.34 |
| BodyMed BDMOXMTRBL K | 2 | 3.26 (2.92, 3.64) | 4.11 (3.08, 5.22) | 3.12 (2.69, 3.47) | 2.93 (2.41, 3.32) | 2.54 (1.96, 3.03) | 0.12 | **0.049** | 4.57 (2.55, 6.43) | 3.35 (2.59, 3.97) | 3.57 (2.73, 4.43) | 3.22 (2.45, 3.94) | 0.61 | 0.46 | 5.19 (3.34, 7.19) | 3.76 (2.87, 4.54) | 3.31 (2.45, 4.06) | 2.87 (2.08, 3.58) | 0.15 | 0.09 | 2.72 (2.23, 3.14) | 2.49 (2.11, 2.79) | 2.18 (1.43, 2.72) | 1.88 (1.15, 2.51) | 0.1 | **0.04** |
| Masimo Rad-97 with Adult Probe, Product Code 4050 | 60 | 3.33 (2.9, 3.77) | 3.26 (2.32, 4.04) | 2.94 (2.46, 3.47) | 4.38 (3.89, 4.90) | 4.64 (3.95, 5.30) | **0.01** | **0.005** | 3.02 (1.87, 4.03) | 3.42 (2.88, 3.92) | 5.34 (4.77, 5.95) | 5.72 (4.88, 6.54) | **<0.001** | **<0.001** | 3.40 (2.38, 4.28) | 3.02 (2.49, 3.55) | 4.74 (4.18, 5.29) | 5.00 (4.36, 5.55) | **0.009** | **0.002** | 3.07 (2.17, 3.85) | 2.31 (1.80, 2.82) | 2.77 (2.36, 3.16) | 2.88 (2.37, 3.36) | 0.99 | 0.75 |
| Guangdong Biolight M800 | 43 | 3.53 (3.17, 3.88) | 3.71 (2.72, 4.66) | 3.37 (2.92, 3.82) | 3.67 (3.02, 4.23) | 2.81 (2.27, 3.29) | 0.85 | 0.26 | 5.72 (4.03, 7.29) | 4.84 (3.83, 5.76) | 5.04 (3.30, 6.49) | 3.05 (1.72, 3.97) | 0.59 | **0.048** | 1.98 (1.53, 2.41) | 2.67 (1.96, 3.35) | 3.28 (2.41, 4.11) | 2.80 (1.95, 3.46) | **0.03** | 0.16 | 2.03 (1.77, 2.30) | 2.21 (1.71, 2.65) | 2.87 (2.15, 3.48) | 2.52 (1.64, 3.21) | 0.11 | 0.5 |
| Masimo Rad-G with Adult Probe, Product Code 4050 | 55 | 4.27 (3.70, 4.89) | 3.71 (2.41, 4.99) | 4.14 (3.31, 4.97) | 5.58 (4.63, 6.37) | 5.92 (5.12, 6.63) | **0.01** | **0.003** | 4.21 (2.89, 5.45) | 5.16 (4.09, 6.26) | 6.72 (5.61, 7.56) | 7.05 (6.11, 7.89) | **0.004** | **0.002** | 3.84 (2.59, 5.08) | 4.16 (3.31, 4.91) | 5.46 (4.47, 6.30) | 5.77 (4.76, 6.63) | **0.03** | **0.01** | 3.15 (1.74, 4.57) | 2.82 (2.15, 3.45) | 3.33 (2.73, 3.81) | 3.52 (3.03, 3.97) | 0.29 | 0.15 |
| Guangdong Biolight M70 | 37 | 5.08 (4.30, 5.91) | 3.70 (2.57, 4.70) | 5.10 (3.95, 6.23) | 5.71 (4.22, 7.05) | 5.03 (3.41, 6.55) | 0.1 | 0.27 | 5.64 (3.31, 7.51) | 8.51 (6.56, 10.31) | 8.80 (6.26, 11.01) | 7.75 (5.07, 10.16) | 0.1 | 0.22 | 1.90 (1.59, 2.21) | 2.69 (1.93, 3.43) | 4.19 (2.96, 5.32) | 4.42 (2.80, 5.69) | **0.007** | **0.03** | 2.50 (2.11, 2.83) | 1.90 (1.54, 2.27) | 2.64 (1.88, 3.41) | 2.45 (1.60, 3.30) | 0.73 | 0.56 |

| Device | Device ID | Overall Device $A_{RMS}$ (95% Confidence Interval) | 70–100% $SaO_2$ $A_{RMS}$ (Mean, 95% CI) | | | | | | 70–80% $SaO_2$ $A_{RMS}$ (Mean, 95% CI) | | | | | | 80–90% $SaO_2$ $A_{RMS}$ (Mean, 95% CI) | | | | | | 90–100% $SaO_2$ $A_{RMS}$ (Mean, 95% CI) | | | | | |
| --- | --- | --- | --- | --- | --- | --- | --- | --- | --- | --- | --- | --- | --- | --- | --- | --- | --- | --- | --- | --- | --- | --- | --- | --- | --- | --- |
|  |  |  | Light | Medium | Dark | Very Dark | P-value: Light vs Dark | P-value: Light vs Very Dark | Light | Medium | Dark | Very Dark | P-value: Light vs Dark | P-value: Light vs Very Dark | Light | Medium | Dark | Very Dark | P-value: Light vs Dark | P-value: Light vs Very Dark | Light | Medium | Dark | Very Dark | P-value: Light vs Dark | P-value: Light vs Very Dark |
| Acare (2023) AH-M1 | 57 | 5.28 (4.60, 5.96) | 4.26<br>(3.13, 5.33) | 5.63<br>(4.79, 6.54) | 4.63<br>(3.39, 5.79) | 4.40 (3.03, 5.51) | 0.93 | 0.96 | 7.49<br>(5.13, 9.26) | 9.28<br>(7.64, 10.95) | 7.62<br>(5.49, 9.47) | 7.45<br>(5.03, 9.55) | 0.92 | 0.95 | 2.99<br>(2.08, 3.88) | 3.35<br>(2.59, 4.07) | 2.77<br>(2.17, 3.30) | 2.93<br>(2.13, 3.54) | 0.7 | 0.94 | 1.87<br>(0.93, 2.61) | 1.68<br>(1.25, 2.19) | 1.94<br>(1.40, 2.53) | 2.00<br>(1.22, 2.71) | 0.82 | 0.78 |
| Acare (2020) AH-MX | 42 | 6.80 (6.05, 7.48) | 7.86<br>(5.62, 9.75) | 6.08<br>(5.36, 6.72) | 6.90<br>(5.52, 8.14) | 6.57 (5.08, 7.99) | 0.55 | 0.47 | 12.54<br>(9.50, 15.41) | 10.11<br>(8.83, 11.25) | 11.71<br>(9.56, 13.68) | 10.81<br>(8.87, 12.70) | 0.76 | 0.53 | 3.96<br>(2.84, 4.92) | 3.44<br>(3.01, 3.83) | 4.33<br>(3.46, 5.08) | 4.43<br>(3.28, 5.45) | 0.57 | 0.47 | 1.72<br>(1.31, 2.10) | 1.42<br>(1.09, 1.76) | 1.86<br>(1.21, 2.45) | 1.43<br>(0.94, 1.84) | 0.95 | 0.36 |
| American Oxygen LK-87 | 20 | 7.75 (7.23, 8.32) | 8.02<br>(6.23, 9.48) | 7.94<br>(7.21, 8.61) | 7.24<br>(6.62, 7.85) | 7.16 (6.43, 7.86) | 0.58 | 0.54 | 9.08<br>(7.53, 10.32) | 9.88<br>(9.12, 10.63) | 9.26<br>(8.02, 10.37) | 9.46<br>(8.14, 10.75) | 0.9 | 0.78 | 10.70<br>(6.33, 13.85) | 9.17<br>(7.76, 10.43) | 8.21<br>(6.46, 9.66) | 7.67<br>(5.93, 8.98) | 0.42 | 0.33 | 2.81<br>(0.88, 4.68) | 3.37<br>(2.08, 4.37) | 2.88<br>(1.45, 4.27) | 2.99<br>(1.36, 4.52) | 0.86 | 0.83 |

Accuracy root mean square error ( $A_{RMS}$ ) is reported for each device in a cohort that is as large as possible while satisfying the enrollment criteria in the anticipated 2025 U.S. Food and Drug Administration (FDA) and 2025 International Organization for Standardization (ISO) guidelines as the overall point estimate with 95 % confidence interval (CI). Columns include the mean $A_{RMS}$ and 95 % CI within each functional arterial oxygen saturation ( $SaO_2$ ) decile and within four skin color bins. Skin color bins are defined by Individual Typology Angle (ITA) measured at the dorsal distal phalanx (DP): ITA > 30° (light), -30° ≤ ITA ≤ 30° (medium), ITA < -30° (dark), and ITA < -50° (very dark); note that “very dark” is a subset of the dark bin. P-values test differences in $A_{RMS}$ between light versus dark and light versus very dark bins using a Welch’s two-sample t-test. No correction for multiple pairwise comparisons (e.g., Bonferroni) was applied, as the 2025 draft FDA guidelines do not require adjustment. P-values near 0.05 are reported to three decimal places, and those meeting $p < 0.05$ are bolded. The 95% CI were determined using bootstrapping (random resampling with replacement) with 1,000 repetitions.

**Table S10.**
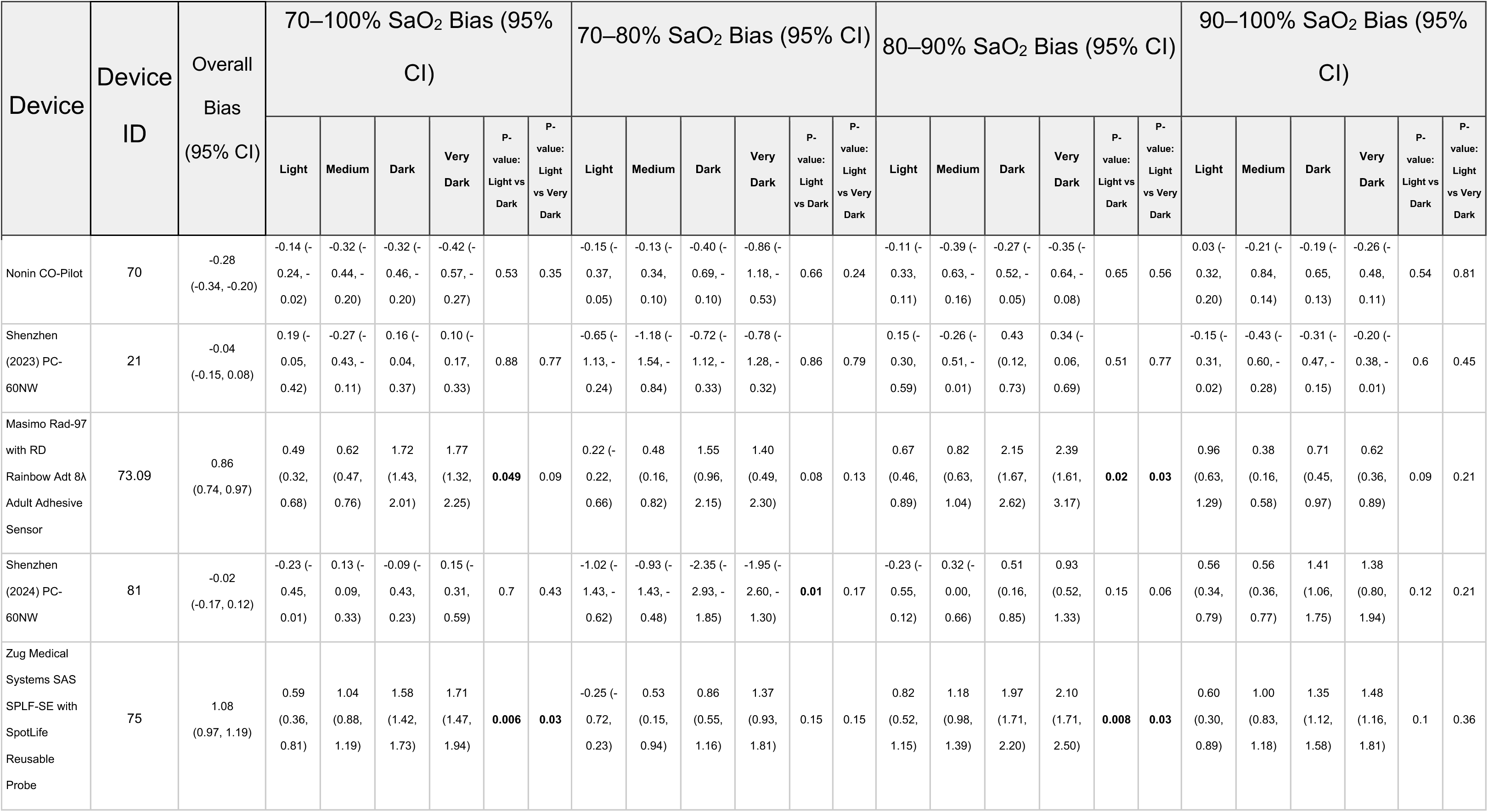
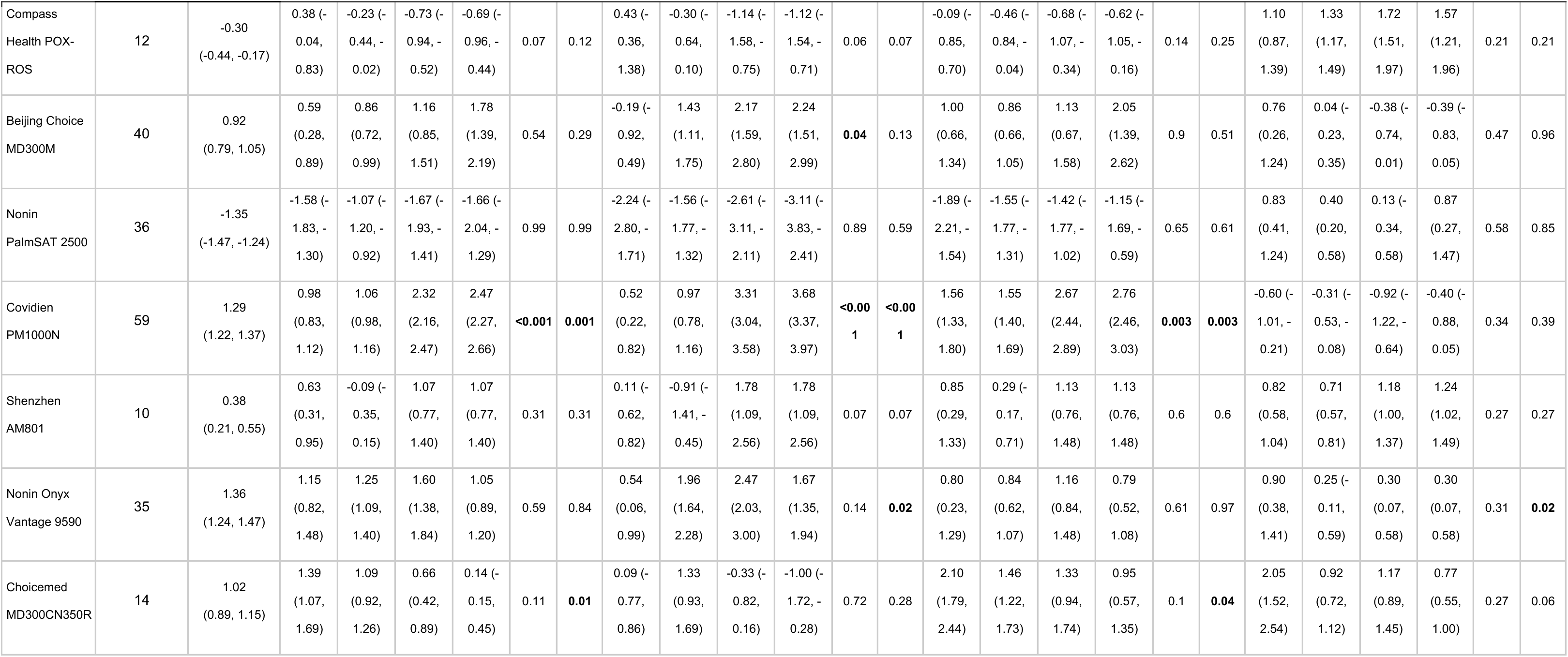
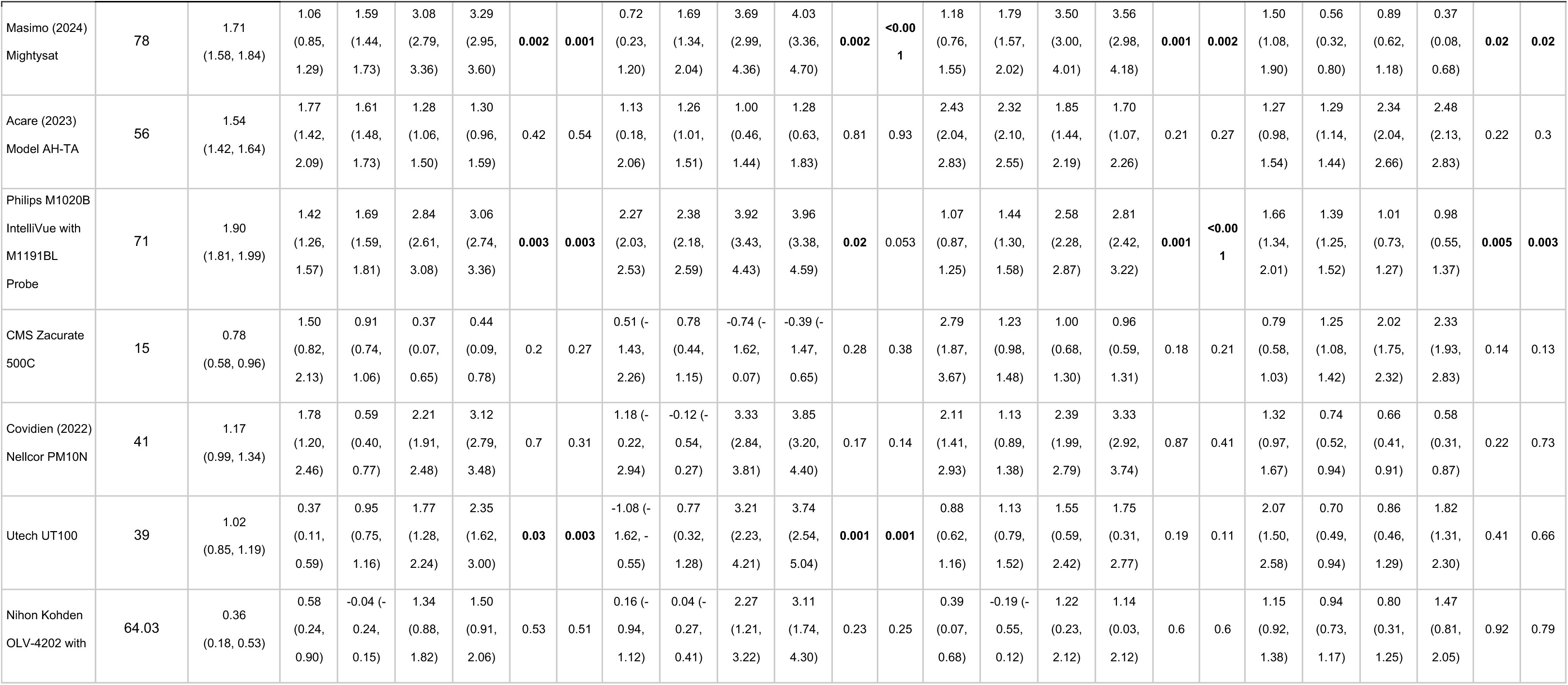
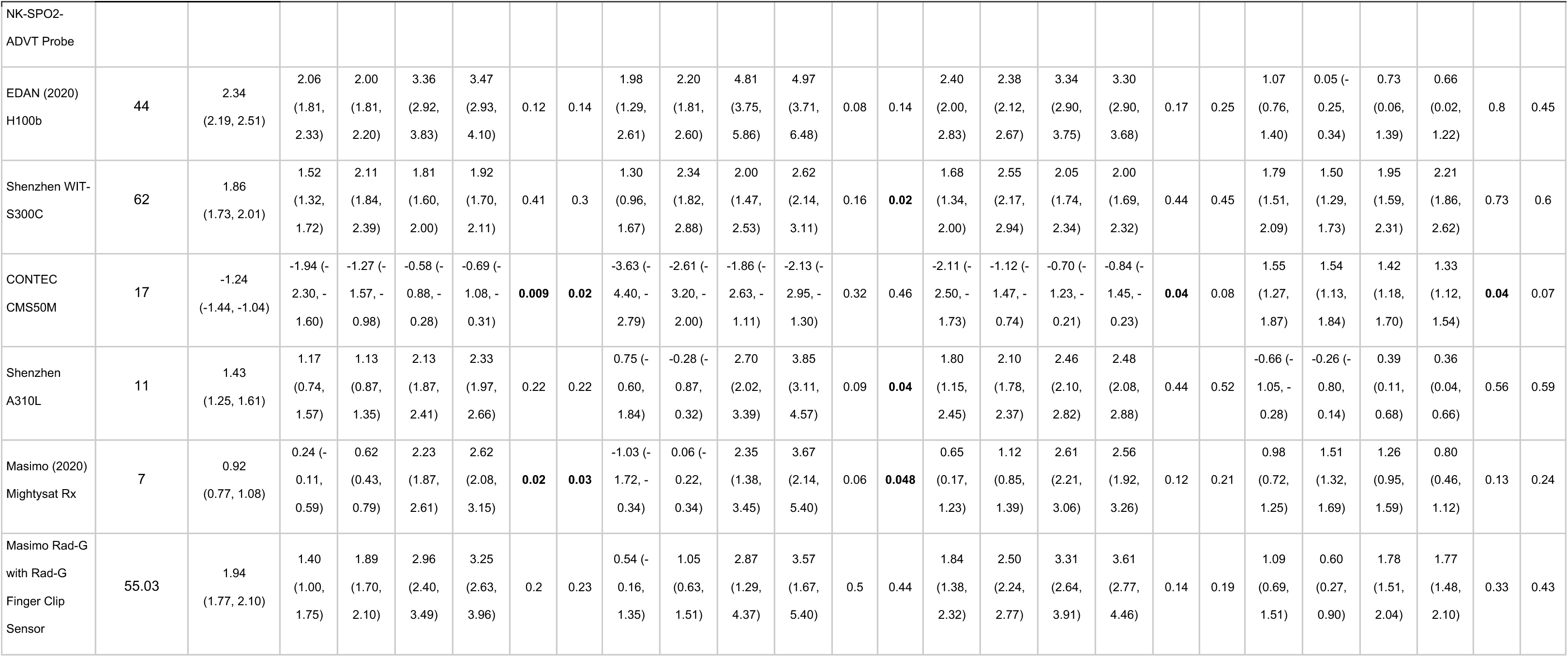
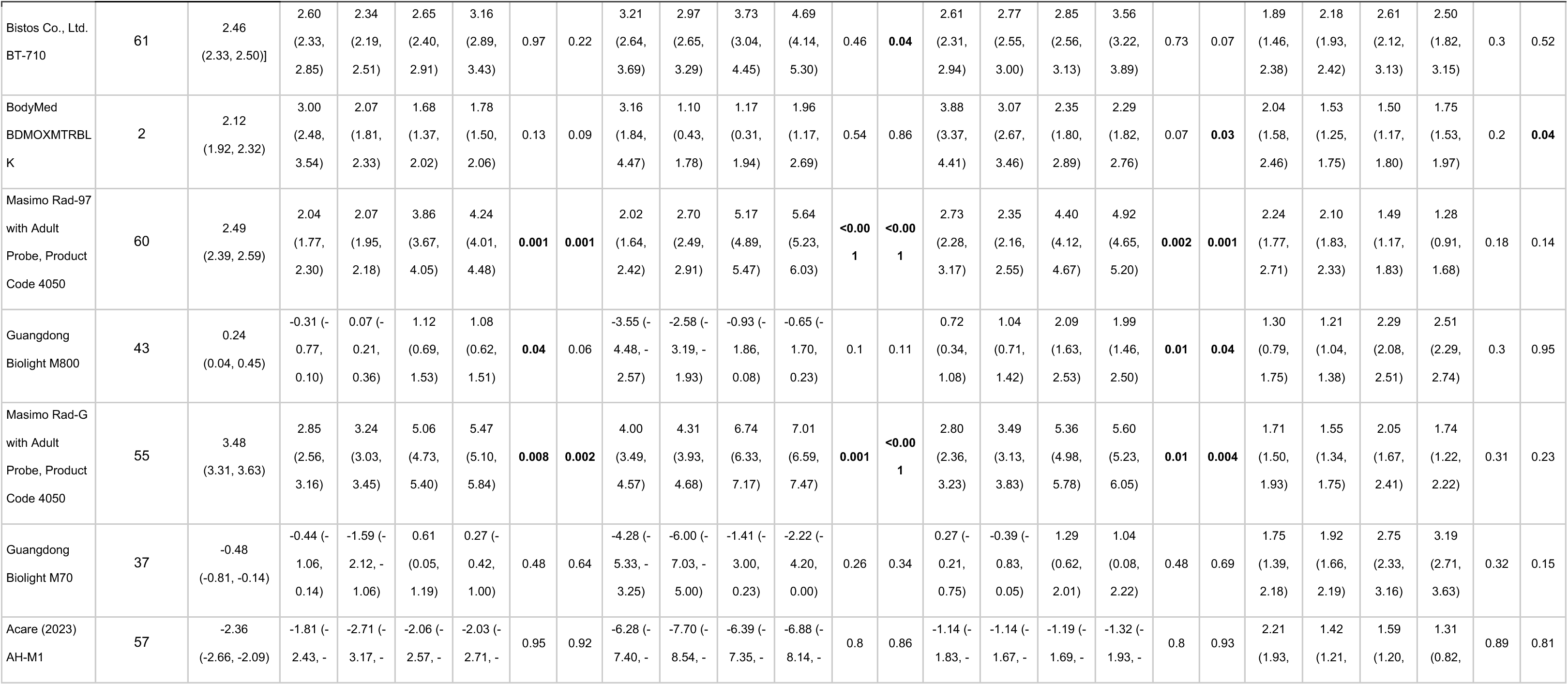
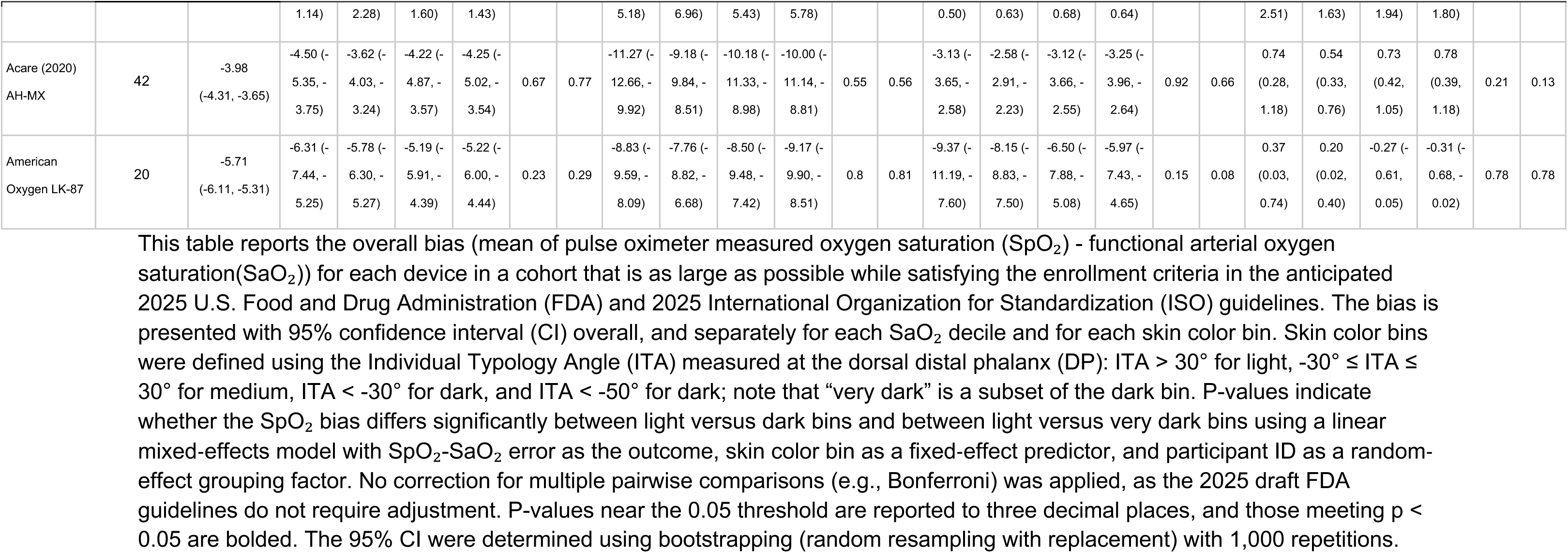
Bias (mean of SpO_2_ minus SaO_2_) at the DP for maximum participant cohorts.

**Table S11.**
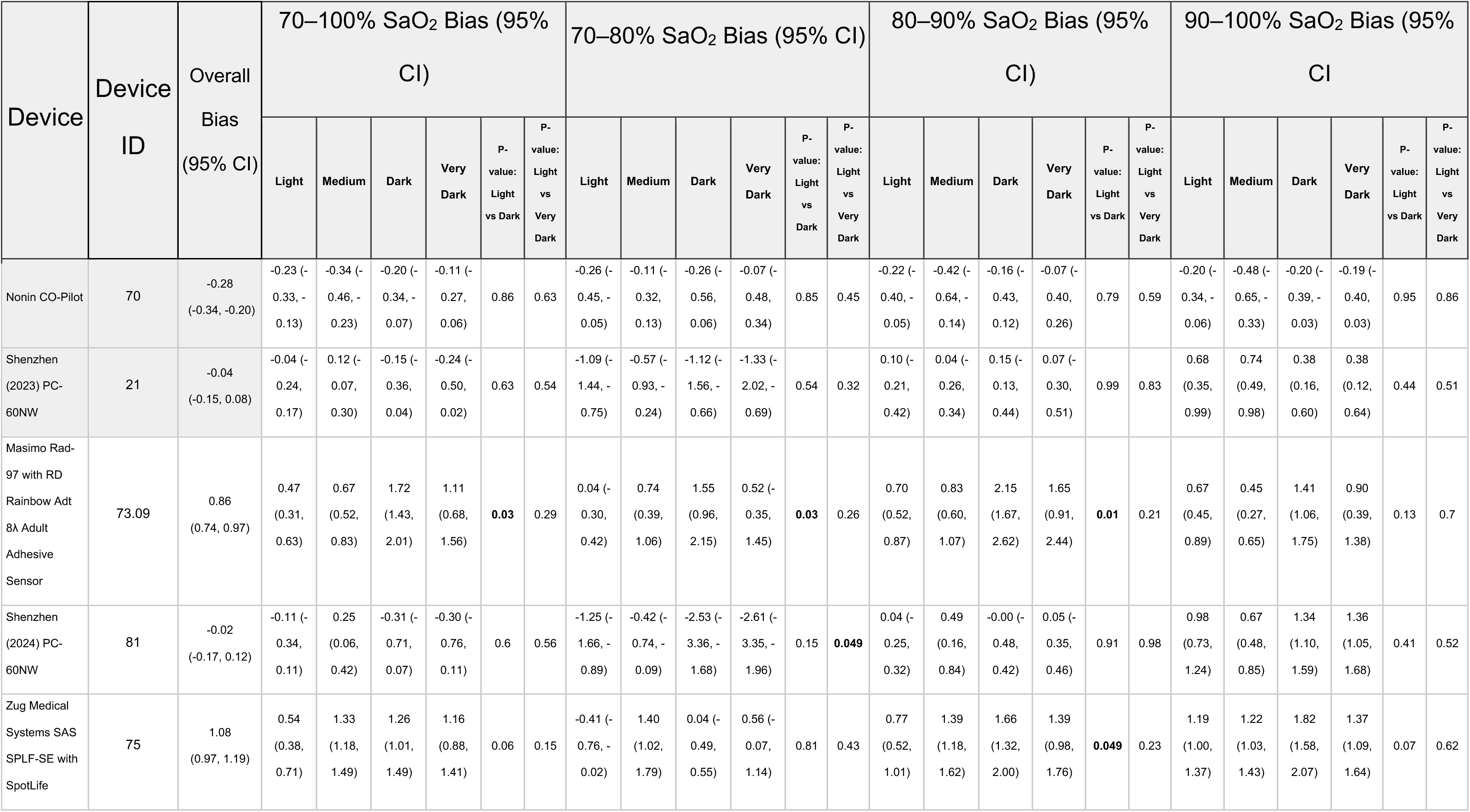
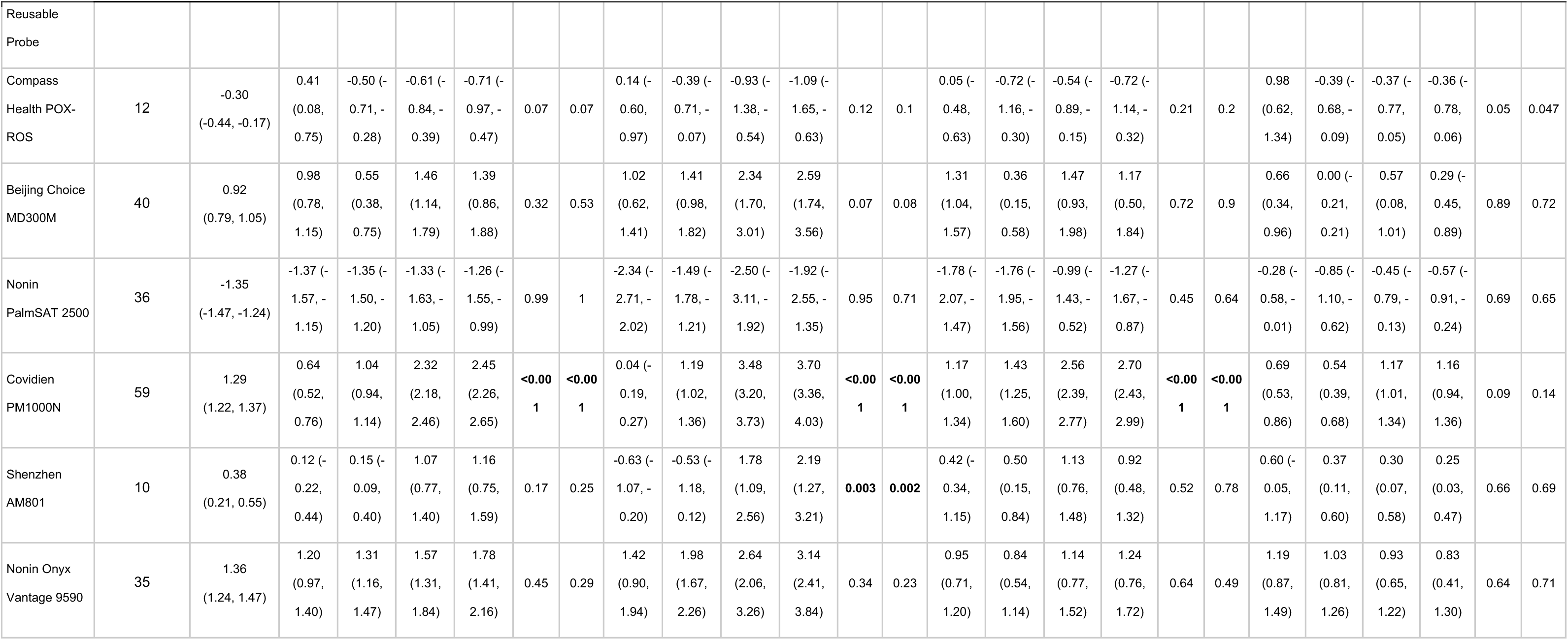
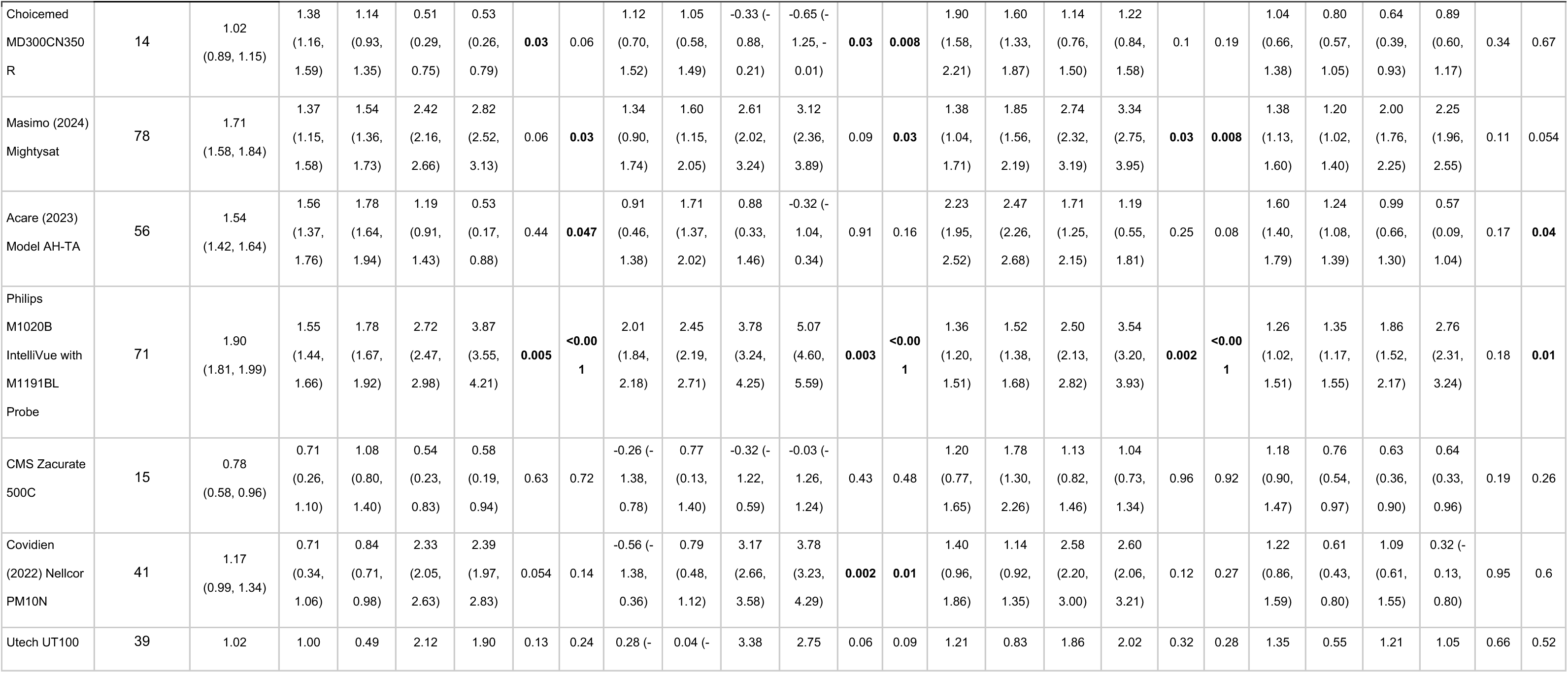
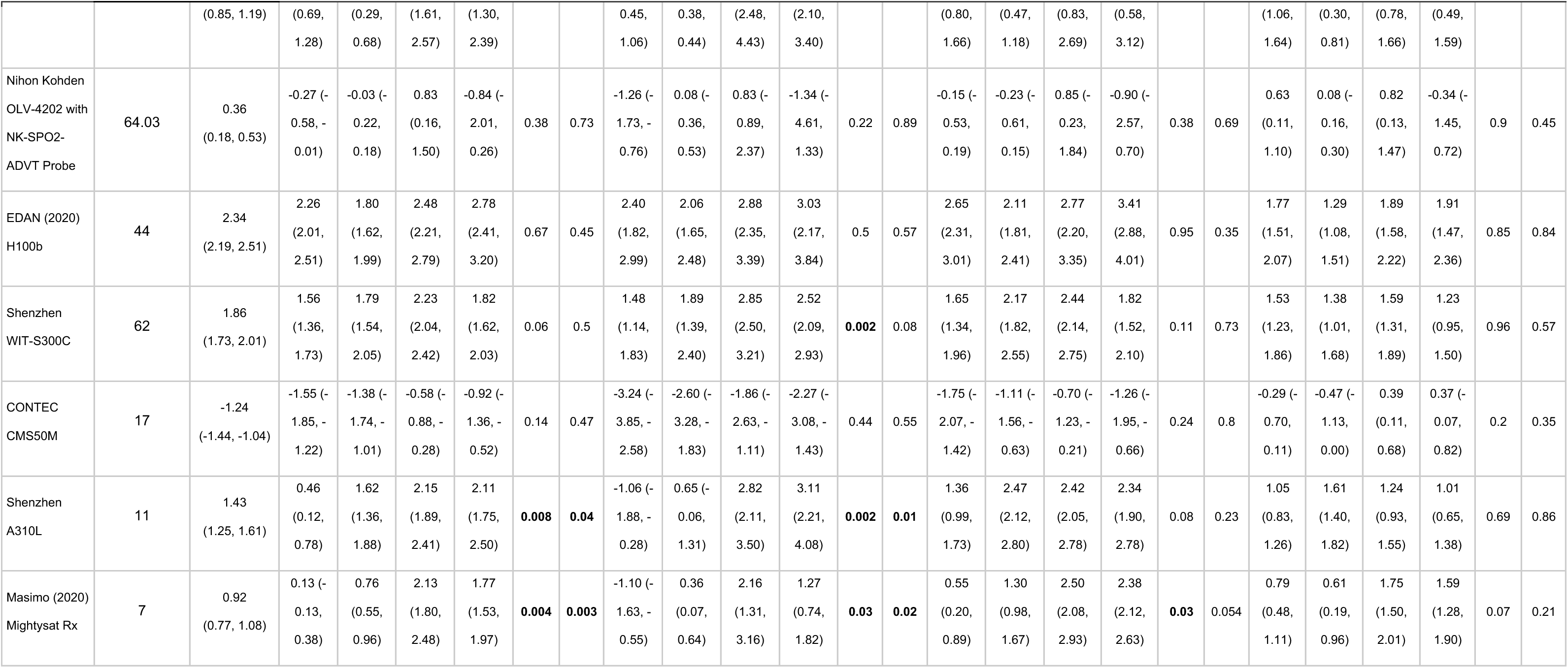
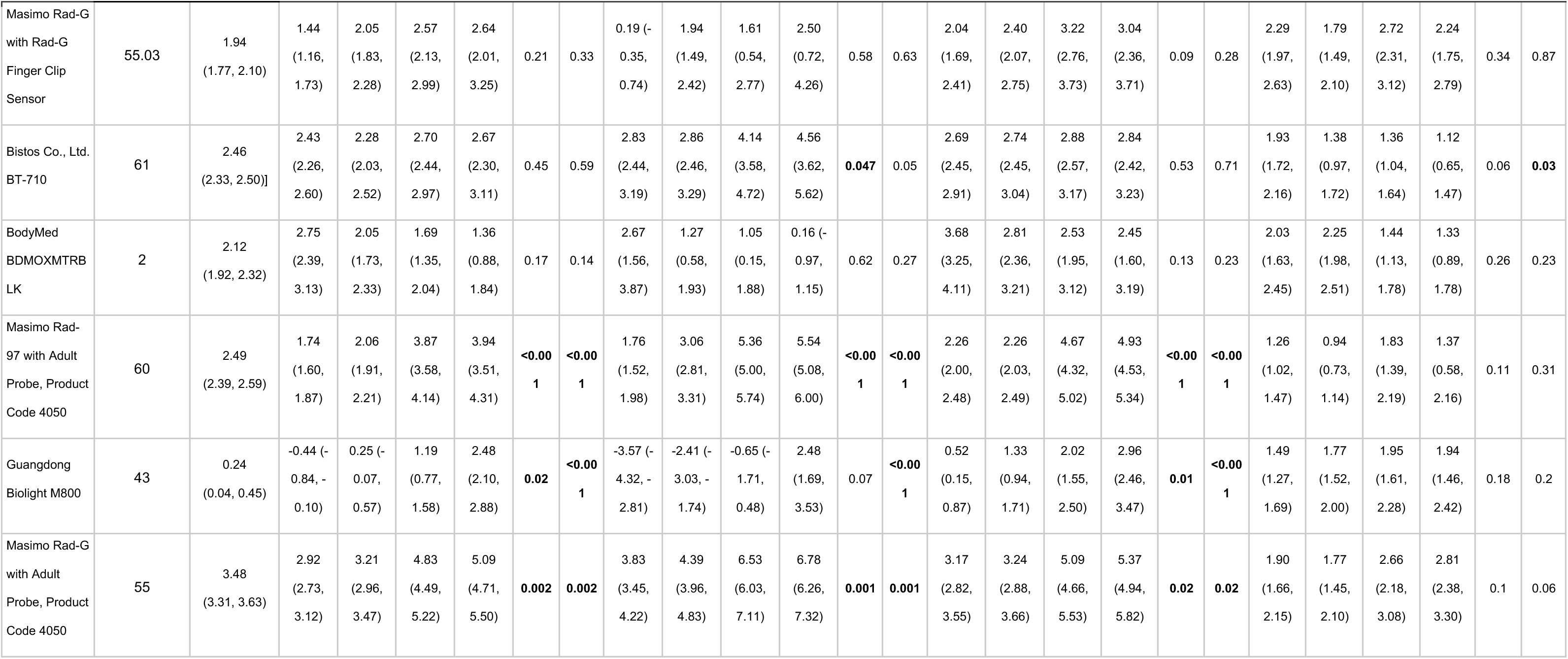
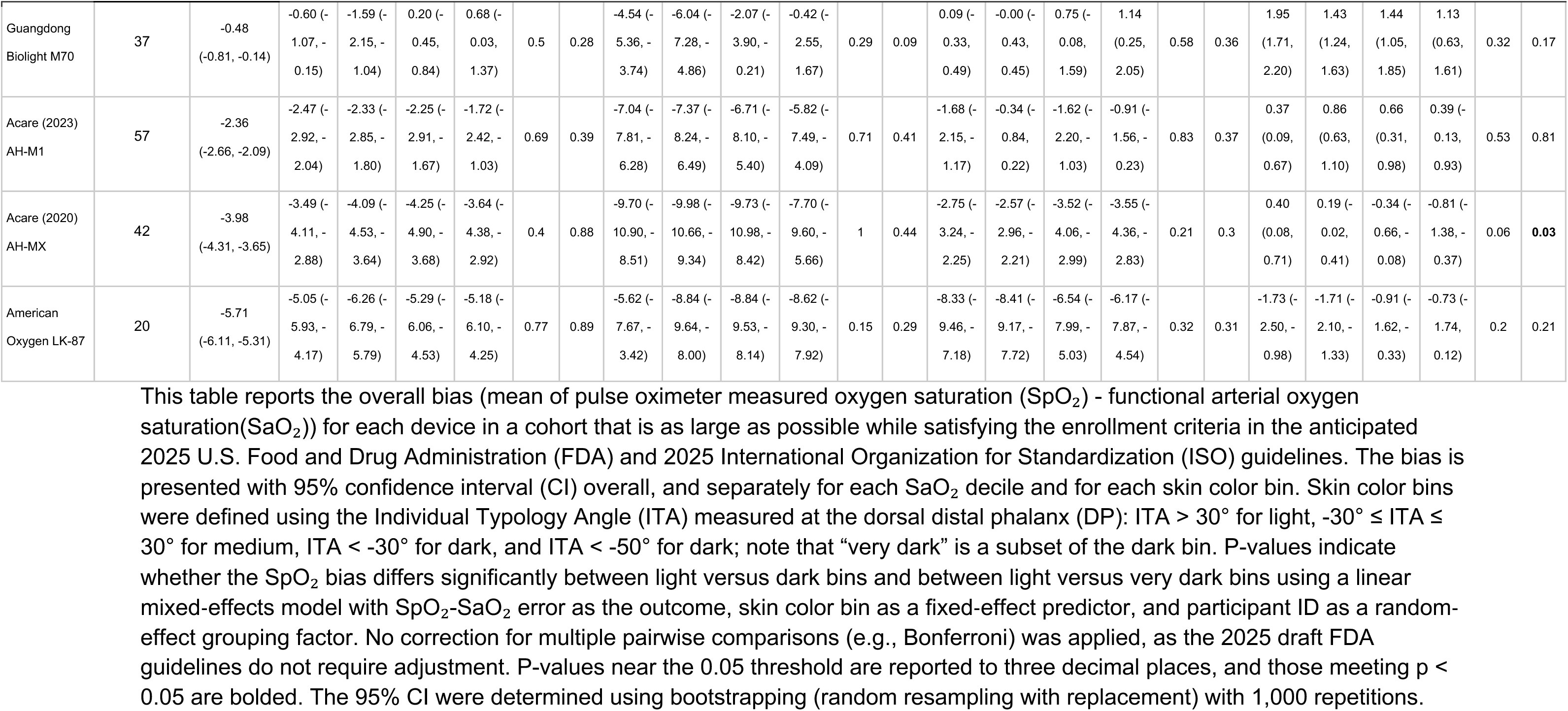
Bias (mean of SpO_2_ minus SaO_2_) at the forehead for maximum participant cohorts.

**Table S12.** Differential bias for 24 participant cohorts.

| Device | Device ID | Pass FDA 2025<br>Differential<br>Bias<br>Thresholds | Pass<br>Anticipated ISO<br>Differential<br>Bias<br>Thresholds | DP ITA-Derived<br>Differential<br>Bias (70–85%<br>SaO<sub>2</sub>) | MST 1–4 vs 5–7<br>Differential<br>Bias (70–85%<br>SaO<sub>2</sub>) | MST 5–7 vs 8–<br>10 Differential<br>Bias (70–85%<br>SaO<sub>2</sub>) | MST 1–4 vs 8–<br>10 Differential<br>Bias (70–85%<br>SaO<sub>2</sub>) | DP ITA-Derived<br>Differential<br>Bias (85–100%<br>SaO<sub>2</sub>) | MST 1–4 vs 5–<br>7 Differential<br>Bias (85–100%<br>SaO<sub>2</sub>) | MST 5–7 vs 8–<br>10 Differential<br>Bias (85–100%<br>SaO<sub>2</sub>) | MST 1–4 vs 8–<br>10 Differential<br>Bias (85–100%<br>SaO<sub>2</sub>) |
| --- | --- | --- | --- | --- | --- | --- | --- | --- | --- | --- | --- |
| Nonin CO-Pilot | 70 |  | ✓ | -0.41 (-2.07, 1.26) | -0.39 (-2.20, 1.42) | 0.57<br>(-1.02, 2.16) | 0.18 (-1.55, 1.91) | -0.32 (-1.47, 0.83) | -0.70<br>(-1.94, 0.53) | 0.28<br>(-0.81, 1.36) | -0.43<br>(-1.61, 0.75) |
| Shenzhen (2023) PC-60NW | 21 |  | ✓ | 0.19 (-0.97, 1.35) | -0.03 (-1.20, 1.13) | -0.02<br>(-1.26, 1.22) | -0.06 (-1.27, 1.15) | 0.29 (-0.75, 1.33) | -0.18<br>(-1.19, 0.83) | 0.66<br>(-0.42, 1.73) | 0.48<br>(-0.57, 1.52) |
| Masimo Rad-97 with<br>RD Rainbow Adt 8λ<br>Adult Adhesive<br>Sensor | 73.09 |  | ✓ | 1.87 (0.50, 3.24) | 2.21 (0.93, 3.50) | -0.67<br>(-2.24, 0.90) | 1.54 (0.18, 2.91) | 1.13 (0.18, 2.08) | 1.28<br>(0.33, 2.22) | -0.48<br>(-1.64, 0.69) | 0.80<br>(-0.21, 1.81) |
| Shenzhen (2024) PC-60NW | 81 |  | ✓ | -0.37 (-2.28, 1.53) | 1.02 (-0.44, 2.47) | -1.98<br>(-3.65, -0.30) | -0.96 (-2.49, 0.57) | 0.06 (-0.88, 1.00) | -0.19<br>(-0.99, 0.62) | 0.18<br>(-0.75, 1.10) | -0.01<br>(-0.85, 0.83) |
| Zug Medical Systems<br>SAS SPLF-SE with<br>SpotLife Reusable<br>Probe | 75 | ✓ | ✓ | 1.75 (0.37, 3.13) | 1.54 (0.21, 2.87) | -0.24<br>(-1.69, 1.21) | 1.30 (-0.04, 2.64) | 0.46 (-0.53, 1.44) | 0.50<br>(-0.47, 1.47) | -0.04<br>(-1.09, 1.01) | 0.46<br>(-0.50, 1.43) |
| Beijing Choice<br>MD300M | 40 |  | ✓ | 0.95 (-0.60, 2.50) | 0.91 (-0.49, 2.32) | 0.11<br>(-1.39, 1.61) | 1.02 (-0.45, 2.48) | 0.78 (-0.57, 2.14) | -0.40<br>(-1.66, 0.87) | 0.68<br>(-0.67, 2.03) | 0.28<br>(-1.03, 1.60) |
| Compass Health<br>POX-ROS | 12 |  | ✓ | -0.98 (-2.81, 0.86) | -0.75 (-2.67, 1.17) | 0.02<br>(-1.69, 1.73) | -0.73 (-2.84, 1.38) | -0.43 (-1.99, 1.13) | -1.10<br>(-2.65, 0.45) | 0.29<br>(-1.09, 1.68) | -0.81<br>(-2.52, 0.90) |
| Shenzhen AM801 | 10 |  | ✓ | 2.14 (0.61, 3.68) | -0.86 (-2.44, 0.73) | 1.81<br>(0.22, 3.40) | 0.95 (-0.78, 2.68) | -0.02 (-1.54, 1.49) | -0.19<br>(-1.71, 1.33) | 0.39<br>(-1.13, 1.90) | 0.20<br>(-1.45, 1.85) |
| CMS Zacurate 500C | 15 |  | ✓ | -0.86 (-2.97, 1.26) | -0.6 (-2.80, 1.59) | -0.66<br>(-2.78, 1.45) | -1.27 (-3.52, 0.99) | -0.29 (-1.39, 0.82) | -0.66<br>(-1.79, 0.48) | 0.26<br>(-0.84, 1.36) | -0.39<br>(-1.57, 0.78) |
| Acare (2023) Model<br>AH-TA | 56 |  | ✓ | -0.03 (-1.66, 1.59) | 0.29 (-1.25, 1.84) | -0.44<br>(-2.21, 1.33) | -0.15 (-1.85, 1.55) | -0.66 **(-2.06, 0.73)** | -0.09<br>(-1.33, 1.14) | -1.27<br>**(-2.68, 0.14)** | -1.37<br>**(-2.72, -0.02)** |
| Choicemed<br>MD300CN350R | 14 |  | ✓ | -1.39 (-2.96, 0.17) | -0.56 (-2.04, 0.93) | -1.05<br>(-2.71, 0.60) | -1.61 (-3.27, 0.06) | -1.12 **(-2.57, 0.33)** | 0.28<br>**(-1.20, 1.75)** | -0.47<br>**(-2.12, 1.18)** | -0.19<br>**(-1.84, 1.46)** |
| Nonin Onyx Vantage<br>9590 | 35 |  | ✓ | 0.27 (-1.43, 1.97) | 1.07 (-0.53, 2.67) | -0.50<br>(-2.24, 1.24) | 0.57 (-1.02, 2.17) | -0.59 **(-1.88, 0.70)** | 0.63<br>**(-0.63, 1.89)** | -0.49<br>**(-1.85, 0.88)** | 0.14<br>(-1.12, 1.40) |
| Nonin PalmSAT 2500 | 36 |  | ✓ | -0.97 (-2.89, 0.95) | **-2.12 (-3.96, -0.28)** | 1.15<br>(-0.88, 3.17) | -0.98 (-2.62, 0.67) | 0.1 **(-1.36, 1.57)** | -1.82<br>**(-3.13, -0.52)** | **1.55**<br>**(0.11, 2.99)** | -0.28<br>(-1.45, 0.90) |
| Shenzhen WIT-S300C | 62 |  | ✓ | 0.76 (-0.80, 2.33) | -0.63 (-2.76, 1.51) | 1.16<br>(-0.79, 3.11) | 0.53 (-1.07, 2.14) | -0.47 **(-1.53, 0.58)** | **-1.64**<br>**(-2.94, -0.33)** | 1.01<br>**(-0.18, 2.20)** | -0.63<br>**(-1.60, 0.35)** |
| Masimo (2024)<br>Mightysat | 78 |  |  | **4.63 (2.91, 6.35)** | 1.58 (-0.12, 3.28) | 2.38<br>**(0.62, 4.14)** | **3.96 (2.15, 5.77)** | **2.18 (1.04, 3.32)** | 0.61<br>**(-0.52, 1.73)** | 1.15<br>**(-0.01, 2.32)** | 1.76<br>**(0.56, 2.96)** |
| Philips M1020B<br>IntelliVue with<br>M1191BL Probe | 71 |  | ✓ | 1.88 (0.56, 3.21) | 0.00 (-1.47, 1.47) | 1.33<br>(-0.14, 2.81) | 1.33 (-0.27, 2.92) | 1.69 **(0.66, 2.72)** | 0.44<br>**(-0.69, 1.57)** | 1.10<br>**(-0.04, 2.23)** | **1.54**<br>**(0.31, 2.76)** |
| Nihon Kohden OLV-4202 with NK-SPO2-ADVT Probe | 64.03 |  | ✓ | **2.28 (-1.12, 5.67)** | **1.22 (-1.81, 4.24)** | 1.52<br>**(-1.77, 4.82)** | **2.74 (-0.70, 6.19)** | 0.75 **(-1.13, 2.63)** | -0.09<br>**(-1.77, 1.59)** | 1.01<br>**(-0.83, 2.84)** | 0.92<br>**(-1.00, 2.84)** |
| Covidien PM1000N | 59 |  | ✓ | 2.91 **(0.77, 5.04)** | 0.18 (-2.36, 2.72) | 2.17<br>**(-0.62, 4.96)** | 2.35 **(0.27, 4.44)** | 0.9 **(-0.22, 2.03)** | -0.67<br>**(-1.92, 0.58)** | 1.38<br>**(0.00, 2.75)** | 0.70<br>**(-0.33, 1.73)** |
| Shenzhen A310L | 11 |  | ✓ | **2.00 (-0.73, 4.74)** | -0.29 (-2.79, 2.21) | 2.23<br>**(-0.39, 4.85)** | **1.95 (-0.93, 4.83)** | -0.63 (-1.41, 0.15) | -0.22<br>(-0.98, 0.53) | -0.15<br>(-0.94, 0.65) | -0.37<br>(-1.24, 0.50) |
| Masimo Rad-G with<br>Rad-G Finger Clip<br>Sensor | 55.03 |  | ✓ | 1.24 (-1.58, 4.06) | 1.59 (-0.58, 3.76) | -1.07<br>(-3.50, 1.36) | 0.52 (-1.91, 2.95) | 0.99 (-0.65, 2.62) | 0.29<br>(-1.04, 1.63) | 0.29<br>(-1.21, 1.79) | 0.59<br>(-0.91, 2.08) |
| CONTEC CMS50M | 17 |  | ✓ | 0.60 (-1.38, 2.57) | 1.50 (-0.33, 3.33) | -0.61<br>(-2.73, 1.51) | 0.89 (-0.96, 2.74) | 0.9 (-0.5, 2.3) | 0.69<br>(-0.68, 2.05) | 0.20<br>(-1.38, 1.77) | 0.88<br>(-0.49, 2.25) |
| Covidien (2022)<br>Nellcor PM10N | 41 |  | ✓ | 2.54 (0.33, 4.75) | 0.96 (-1.39, 3.31) | 1.46<br>(-1.11, 4.03) | 2.42 (0.18, 4.66) | 0.28 (-1.18, 1.73) | -0.21<br>(-1.74, 1.32) | -0.12<br>(-1.80, 1.56) | -0.33<br>(-1.79, 1.14) |
| Bistos Co., Ltd. BT-710 | 61 |  | ✓ | 1.49 (0.31, 2.68) | 0.44 (-1.12, 2.00) | 0.75<br>(-0.90, 2.40) | 1.19 (0.09, 2.29) | 0.38 (-0.71, 1.47) | -0.45<br>(-1.85, 0.95) | 0.77<br>(-0.71, 2.25) | 0.32<br>(-0.66, 1.30) |
| Utech UT100 | 39 |  | ✓ | 3.05 (0.41, 5.69) | -0.12 (-2.71, 2.47) | 1.23<br>(-1.39, 3.84) | 1.11 (-1.51, 3.72) | -0.39 (-1.72, 0.94) | -0.43<br>(-1.57, 0.70) | -0.44<br>(-1.59, 0.71) | -0.88<br>(-2.02, 0.27) |
| BodyMed<br>BDMOXMTRBLK | 2 |  | ✓ | 0.09 (-2.52, 2.70) | -0.74 (-3.02, 1.54) | 0.92<br>(-1.36, 3.21) | 0.19 (-2.29, 2.66) | -0.58 (-1.7, 0.54) | 0.49<br>(-0.51, 1.49) | -0.22<br>(-1.22, 0.79) | 0.27<br>(-0.81, 1.36) |
| Masimo (2020)<br>Mightysat Rx | 7 |  | ✓ | 3.48 (0.03, 6.92) | -0.41 (-3.74, 2.92) | 2.43<br>(-0.90, 5.76) | 2.03 (-1.30, 5.36) | 0.95 (-0.8, 2.7) | 0.63<br>(-0.95, 2.21) | 0.68<br>(-0.91, 2.26) | 1.31<br>(-0.28, 2.89) |
| EDAN (2020) H100b | 44 |  | ✓ | 2.51 (0.24, 4.79) | 0.56 (-2.06, 3.18) | 0.96<br>(-1.71, 3.62) | 1.51 (-0.69, 3.72) | 0.67 (-0.33, 1.67) | 0.13<br>(-0.99, 1.24) | 0.21<br>(-0.93, 1.35) | 0.34<br>(-0.59, 1.27) |
| Guangdong Biolight<br>M800 | 43 |  | ✓ | 2.85 (0.20, 5.50) | -0.81 (-3.62, 2.01) | 2.38<br>(-0.56, 5.33) | 1.57 (-1.02, 4.17) | 0.66 (-0.86, 2.18) | -0.76<br>(-2.29, 0.78) | 1.03<br>(-0.57, 2.64) | 0.28<br>(-1.13, 1.69) |
| Masimo Rad-97 with<br>Adult Probe, Product<br>Code 4050 | 60 |  | ✓ | 3.43 (0.28, 6.57) | -1.68 (-4.91, 1.55) | 4.26<br>(0.76, 7.76) | 2.58 (-0.65, 5.81) | 1.42 (-0.06, 2.89) | -0.20<br>(-1.79, 1.39) | 1.33<br>(-0.40, 3.05) | 1.13<br>(-0.47, 2.72) |
| Masimo Rad-G with<br>Adult Probe, Product<br>Code 4050 | 55 |  | ✓ | 3.81 (2.08, 5.54) | -0.66 (-2.56, 1.24) | 3.23<br>(1.17, 5.29) | 2.57 (0.67, 4.47) | 1.47 (0.09, 2.86) | -0.60<br>(-1.90, 0.69) | 1.91<br>(0.49, 3.32) | 1.30<br>(-0.00, 2.61) |
| Acare (2023) AH-M1 | 57 |  | ✓ | 1.51 (-1.76, 4.79) | 0.67 (-2.38, 3.73) | 1.61<br>(-1.47, 4.70) | 2.28 (-1.06, 5.62) | 1.12 (-1.36, 3.59) | 0.16<br>(-2.29, 2.62) | 0.23<br>(-2.24, 2.69) | 0.39<br>(-2.28, 3.06) |
| Guangdong Biolight<br>M70 | 37 |  |  | 9.17 (3.15, 15.19) | 1.60 (-4.18, 7.38) | 4.59<br>(-1.88, 11.05) | 6.19 (-0.28, 12.65) | 0.51 (-1.4, 2.42) | -0.94<br>(-2.54, 0.67) | 1.23<br>(-0.57, 3.03) | 0.29<br>(-1.51, 2.10) |
| Acare (2020) AH-MX | 42 |  | ✓ | -0.27 (-3.62, 3.09) | -0.54 (-4.12, 3.03) | -1.71<br>(-5.50, 2.08) | -2.25 (-5.35, 0.85) | -0.86 (-2.15, 0.42) | 0.34<br>(-1.03, 1.72) | -1.18<br>(-2.65, 0.29) | -0.84<br>(-2.03, 0.35) |
| American Oxygen LK-87 | 20 |  | ✓ | -0.6 (-4.13, 2.93) | -1.38 (-5.01, 2.25) | 0.71<br>(-2.90, 4.32) | -0.67 (-4.61, 3.27) | 1.79 (-0.29, 3.87) | 0.23<br>(-1.90, 2.37) | 1.39<br>(-0.76, 3.54) | 1.63<br>(-0.71, 3.97) |

Point estimates and 95% confidence intervals (CI) for each differential bias criterion are reported per the anticipated 2025 U.S. Food and Drug Administration (FDA) and International Organization for Standardization (ISO) guidelines; a check mark (✓) indicates devices meeting the overall differential bias success criteria in each guideline. Columns display Individual Typology Angle (ITA)-derived differential bias at functional arterial oxygen saturation (SaO<sub>2</sub>) ranges 70–85% and 85–100%, as well as MST-derived differential bias at those same ranges. Point estimates (and their CIs) are bolded if they fail to meet any criterion (point estimate and 95% CI < 3.5% for 70–85% SaO<sub>2</sub> and < 1.5% for 85–100% SaO<sub>2</sub> for 2025 FDA, and point estimate ≤ 4% for 70–85% SaO<sub>2</sub> and ≤ 2% for 85–100% SaO<sub>2</sub> for 2025 ISO). Data reflect devices tested in a cohort size of 24 participants. 95% CIs for differential bias are calculated as described in eMethods, Supplemental Digital Content.

**Table S13.**
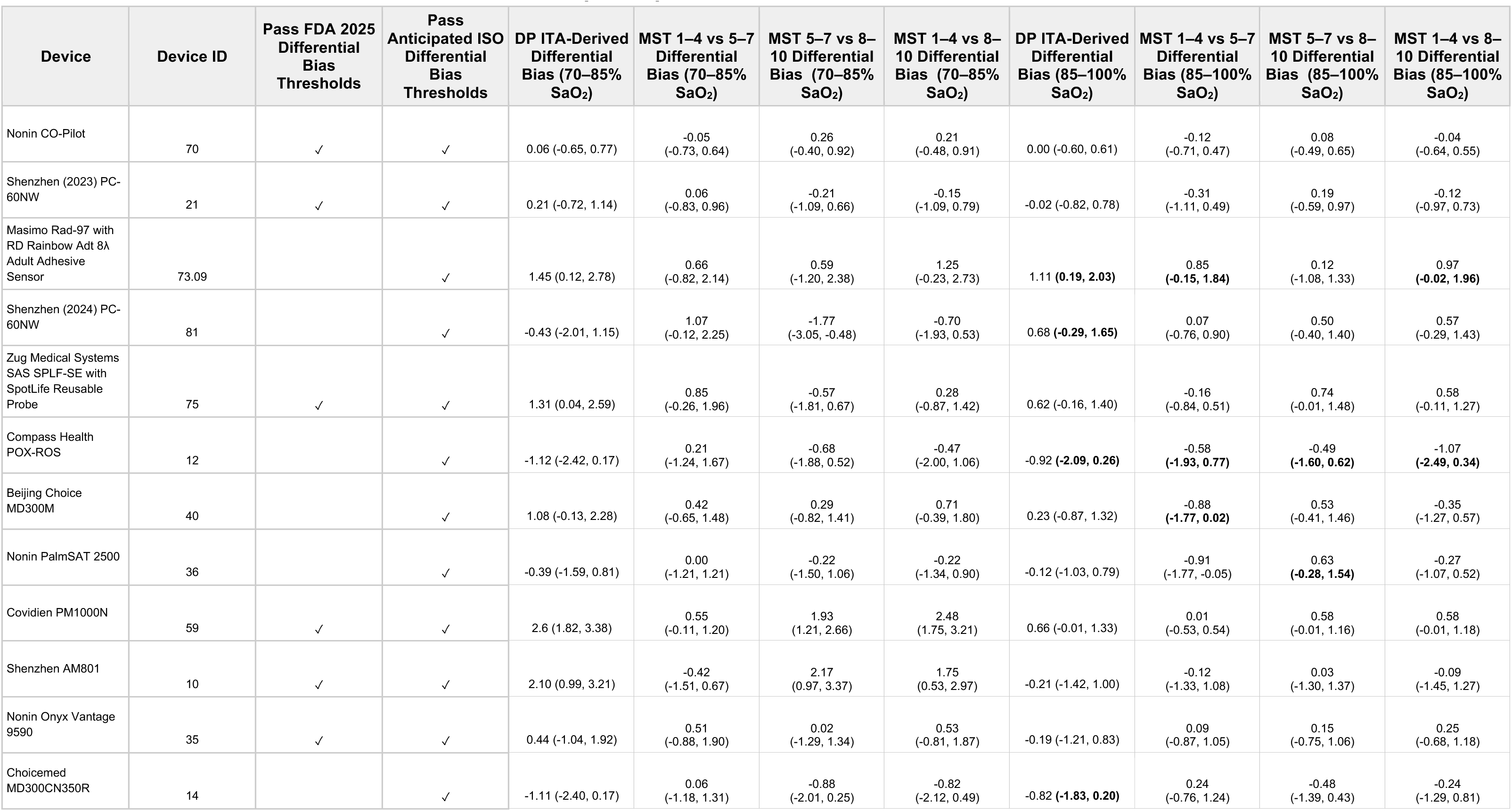
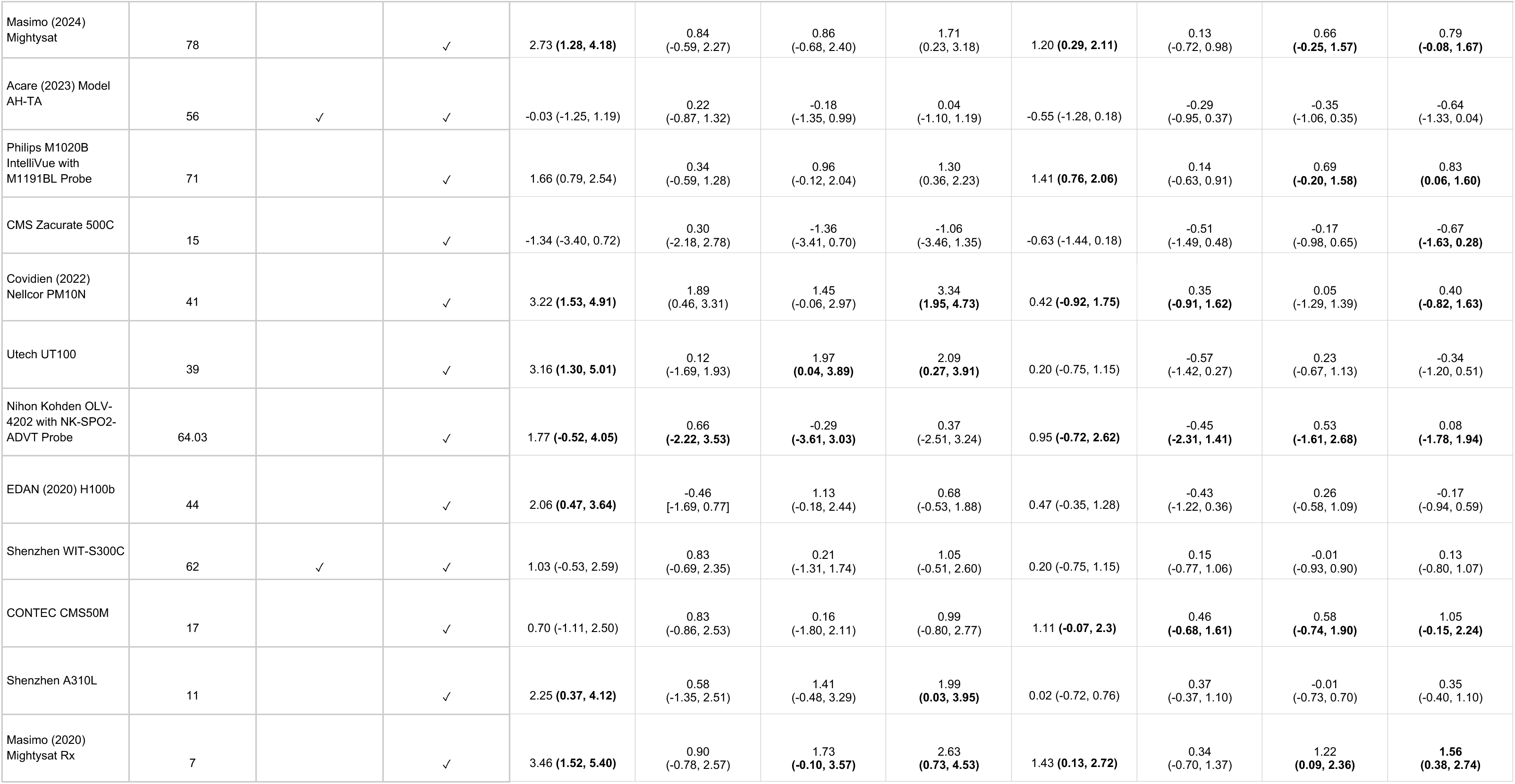
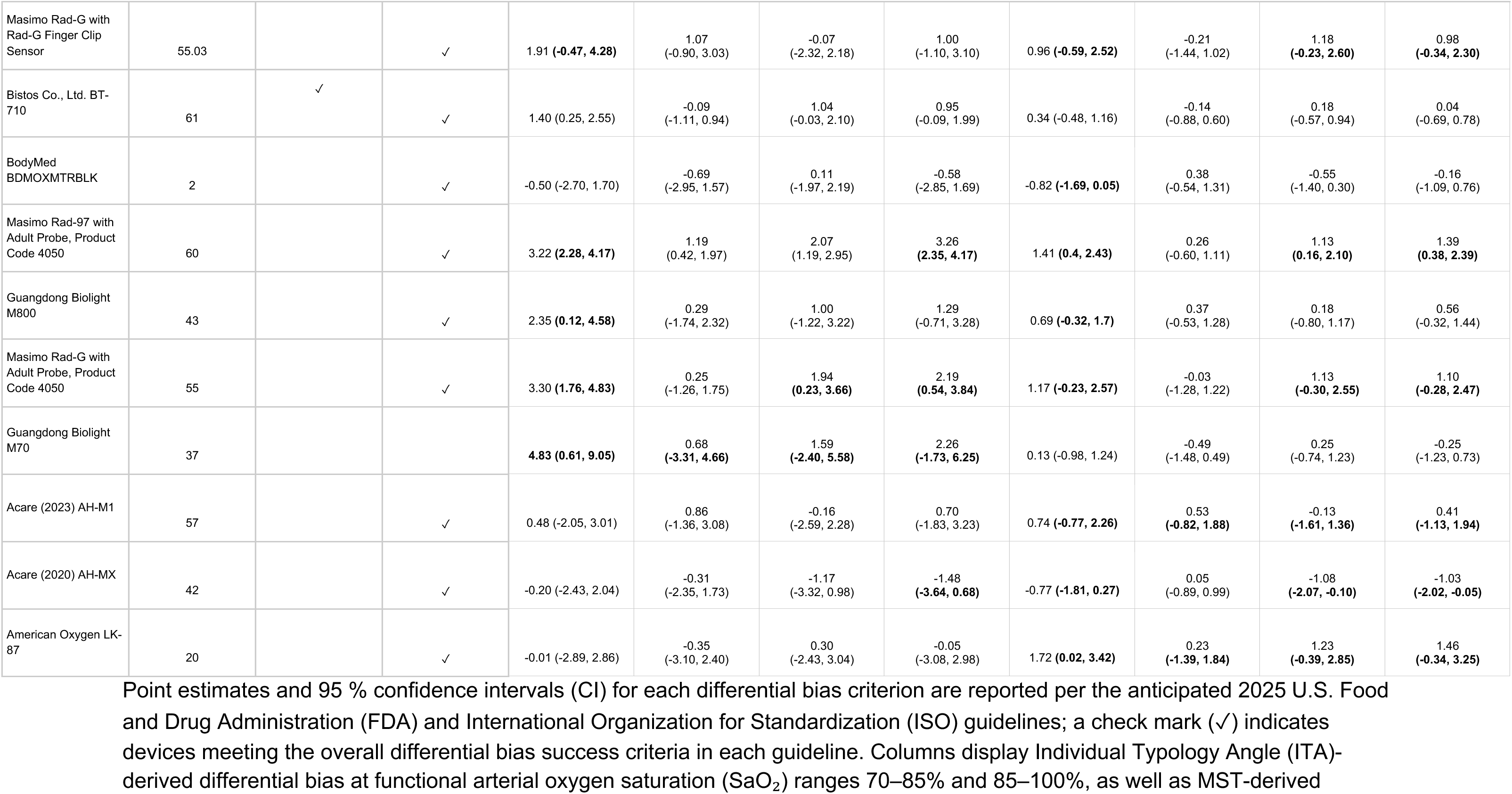
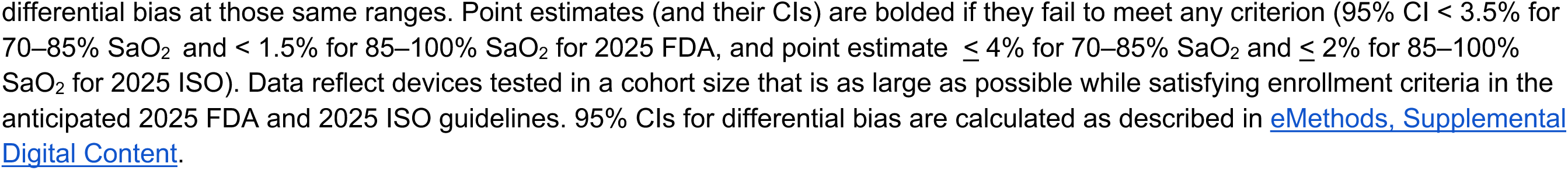
Differential bias for maximum participant cohorts.

**Table S14.** Count of devices with statistically significant differences in bias (mean of SpO_2_ minus SaO_2_) across different definitions of dark and anatomical sites in maximum participant cohorts.

|  | ITA>30° vs ITA<-30°<br>with half <-50°<br>(Light vs. Dark) |  | ITA>30° vs ITA<-50°<br>(Light vs. Very Dark) |  | MST 1–3 vs 8–<br>10 | MST 1–4 vs<br>8–10 | White vs<br>Black/African<br>American |
| --- | --- | --- | --- | --- | --- | --- | --- |
|  | Forehead | DP | Forehead | DP | Forehead | Forehead | Overall |
| **SaO<sub>2</sub> 70–**<br>**100%** | 9 | 11 | 9 | 10 | 9 | 8 | 7 |
| **SaO<sub>2</sub> 70–**<br>**80%** | 12 | 8 | 12 | 10 | 7 | 10 | 7 |
| **SaO<sub>2</sub> 80–**<br>**90%** | 9 | 9 | 6 | 10 | 9 | 9 | 8 |
| **SaO<sub>2</sub> 90–**<br>**100%** | 0 | 3 | 5 | 4 | 4 | 2 | 1 |

Number of devices showing a statistically significant bias difference between dark and light skin color bins at the forehead and dorsal distal phalanx (DP) sites, stratified by saturation ranges. Bias is defined as the mean SpO<sub>2</sub> - SaO<sub>2</sub> error, where SpO<sub>2</sub> is pulse oximeter measured oxygen saturation and SaO<sub>2</sub> is functional arterial oxygen saturation. Comparisons between light versus dark bins were made using a linear mixed-effects model (SpO<sub>2</sub> - SaO<sub>2</sub> error as the outcome, skin color bin as a fixed effect, participant ID as a random intercept) with $\alpha = 0.05$ . Light and dark bins are defined by either Individual Typology Angle (ITA), Monk Skin Tone Scale (MST), or National Institutes of Health (NIH) race. No correction for multiple comparisons (e.g., Bonferroni) was applied, per the 2025 draft U.S. Food and Drug Administration (FDA) guidelines. Results are based on cohorts that are as large as possible while satisfying enrollment criteria in the anticipated 2025 FDA and 2025 ISO (International Organization for Standardization) guidelines.

**Table S15.**
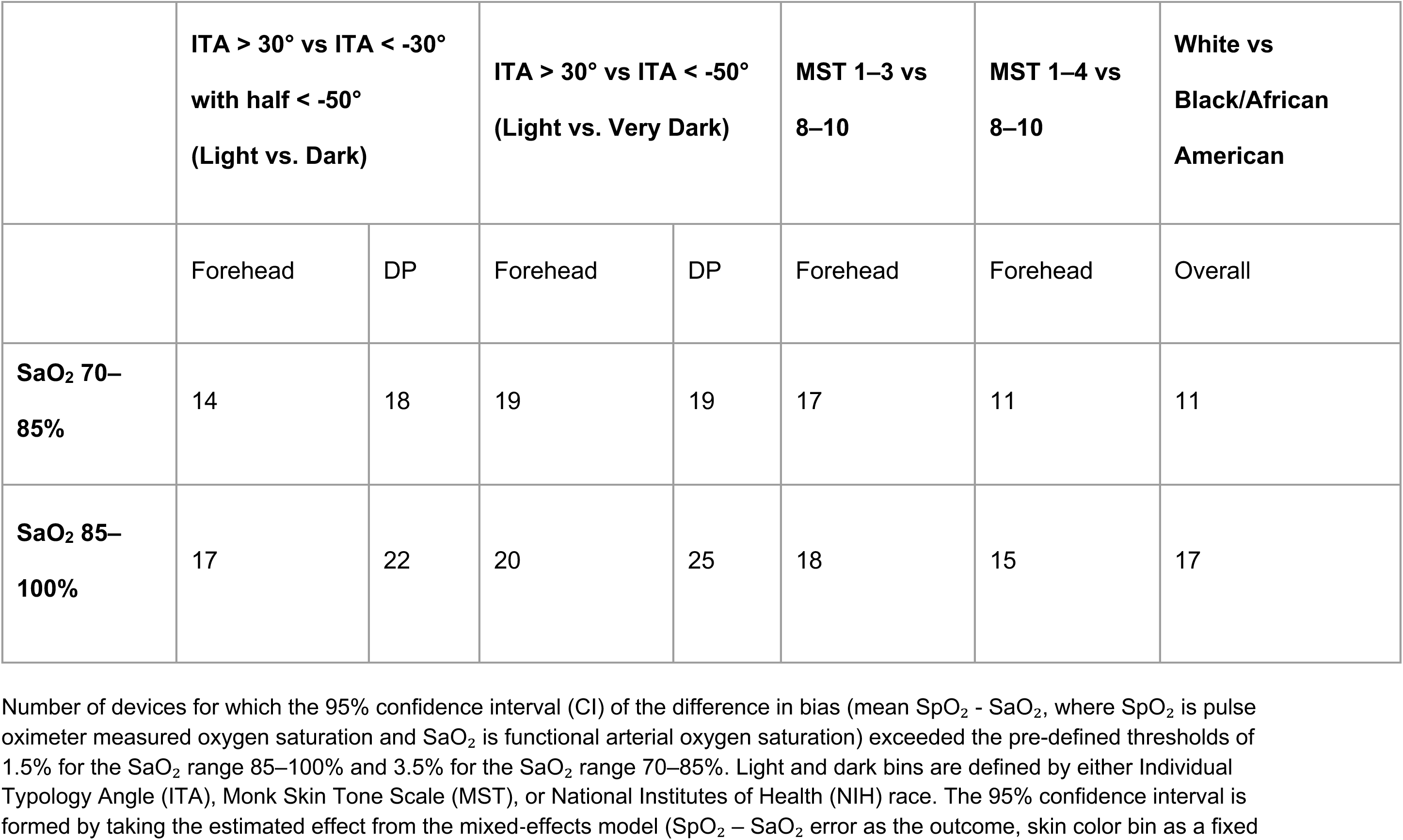
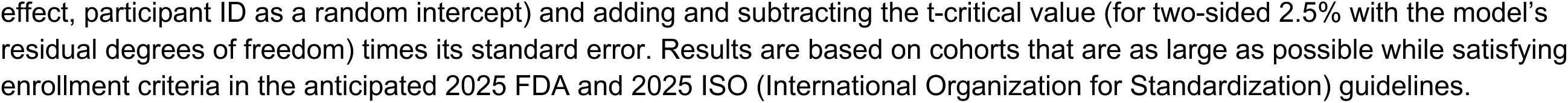
Count of devices with 95% CI of difference in bias (mean of SpO_2_ minus SaO_2_) exceeding pre-defined thresholds across different definitions of dark and anatomical sites in maximum participant cohorts.

**Table S16.**
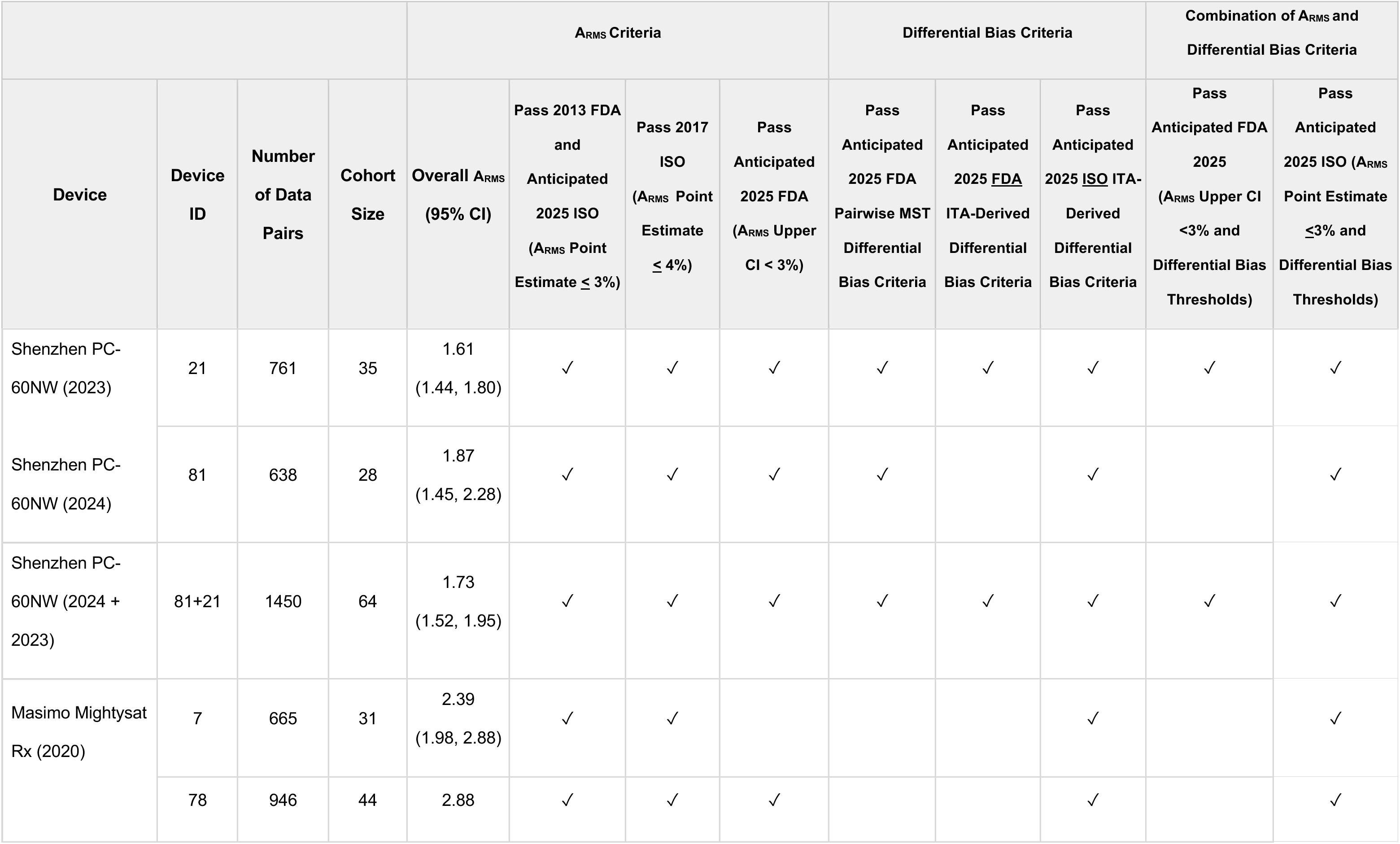
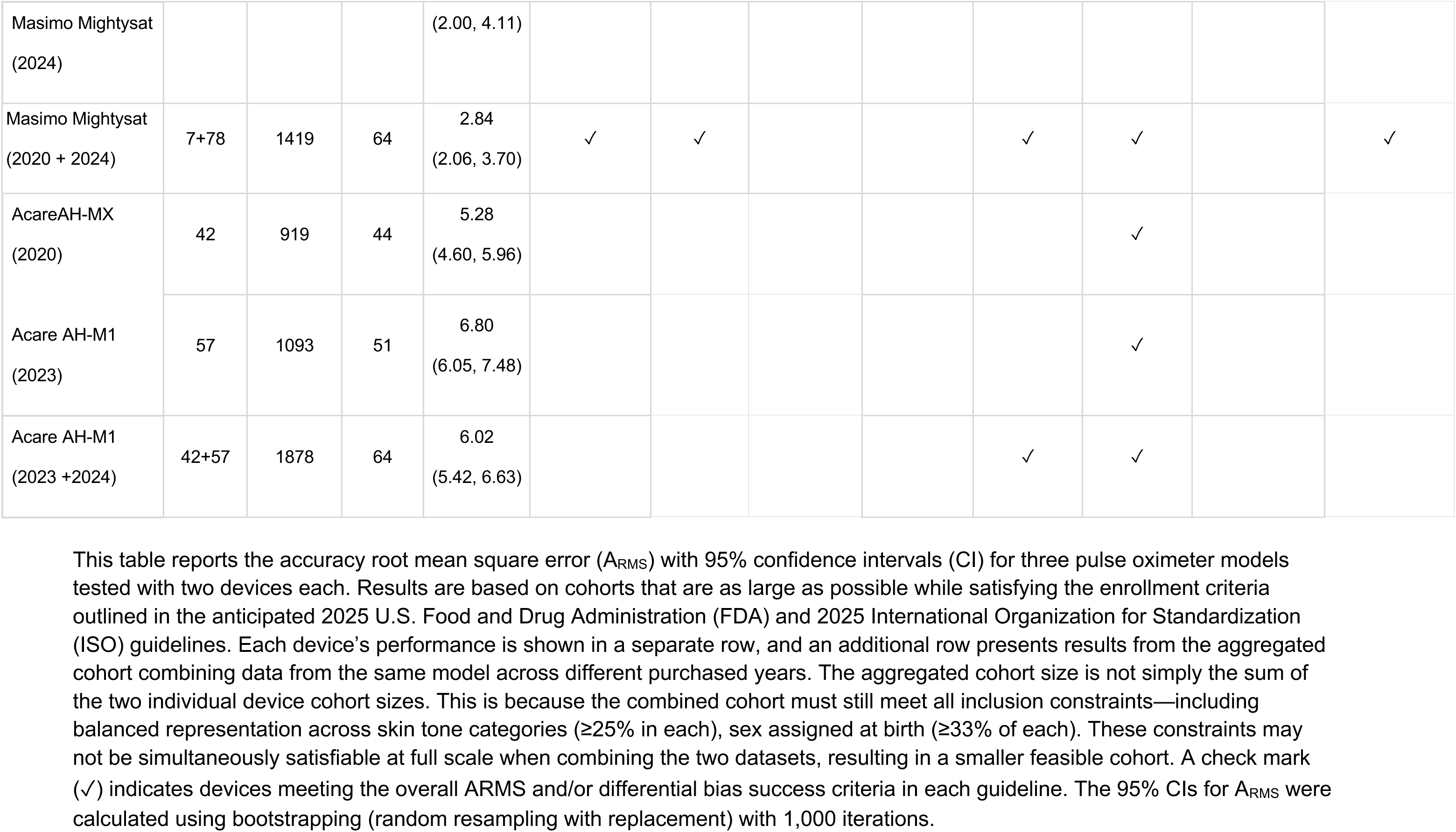
Reproducibility of device conformity with with regulatory frameworks.

**Table S17.** ITA and pulsatility amplitude for participant pigment bins in all device cohorts.

| Device | Device ID | Metric | 24 Participant Cohort Size |  |  |  | Maximum Participant Cohort |  |  |  |
| --- | --- | --- | --- | --- | --- | --- | --- | --- | --- | --- |
|  |  |  | Light | Medium | Dark | Very Dark | Light | Medium | Dark | Very Dark |
| Nonin CO-Pilot | 70 | ITA (Forehead) | 42.32 [39.37, 43.50] | 17.97 [-11.97, 20.78] | -53.32 [-66.82, -47.91] | -66.82 [-71.89, -54.63] | 40.97 [35.96, 45.29] | 18.03 [9.18, 24.39] | -56.14 [-59.99, -48.05] | -58.25 [-66.82, -56.14] |
|  |  | ITA (DP) | 20.94 [19.90, 30.22] | 11.33 [-28.79, 19.25] | -57.66 [-66.69, -37.96] | -57.66 [-84.81, -45.82] | 33.11 [23.79, 38.64] | 10.44 [-10.51, 19.25] | -58.91 [-66.83, -38.43] | -60.16 [-66.83, -45.82] |
|  |  | MST (Forehead) | 3 [1, 3] | 5 [5, 8] | 9 [8, 10] | 9 [8, 10] | 4 [1, 5] | 5 [2, 8] | 8 [8, 10] | 8 [8, 10] |
|  |  | MST (DP) | 2 [2, 4] | 5 [2, 8] | 9 [8, 10] | 9 [8, 10] | 2 [1, 5] | 5 [2, 8] | 8 [7, 10] | 8 [8, 10] |
|  |  | %Mod | 2.70 [1.15, 4.30] | 1.00 [0.70, 4.10] | 4.50 [1.40, 6.70] | 4.50 [2.10, 7.40] | 3.10 [2.00, 4.50] | 2.70 [0.90, 5.10] | 2.50 [1.20, 5.50] | 2.60 [1.20, 5.60] |
| Shenzhen (2023) PC-60NW | 21 | ITA (Forehead) | 39.37 [31.95, 43.48] | 16.59 [3.56, 20.71] | -58.94 [-60.45, -43.66] | -60.45 [-71.89, -58.94] | 39.37 [32.79, 43.50] | 18.03 [3.56, 25.95] | -51.48 [-60.45, -43.66] | -60.45 [-64.00, -54.63] |
|  |  | ITA (DP) | 26.31 [18.64, 30.22] | 8.24 [-1.84, 21.30] | -68.65 [-68.79, -37.96] | -68.65 [-84.81, -60.10] | 27.27 [18.64, 33.29] | 8.24 [-10.51, 21.82] | -61.80 [-68.65, -37.96] | -61.80 [-68.65, -44.20] |
|  |  | MST (Forehead) | 2 [1, 5] | 5 [2, 8] | 8 [8, 10] | 8 [8, 10] | 2 [1, 5] | 5 [2, 8] | 8 [8, 10] | 8 [8, 10] |
|  |  | MST (DP) | 2 [1, 6] | 5 [3, 8] | 9 [8, 9] | 9 [8, 9] | 2 [1, 6] | 5 [3, 8] | 8 [8, 9] | 8 [8, 9] |
|  |  | %Mod | 2.50 [1.10, 3.40] | 3.90 [0.70, 6.10] | 1.60 [0.70, 3.20] | 1.60 [0.40, 2.80] | 2.50 [1.50, 3.40] | 3.10 [0.90, 5.40] | 2.50 [1.20, 6.30] | 2.50 [1.50, 6.30] |
| Masimo Rad-97 with RD Rainbow Adt 8λ Adult Adhesive Sensor | 73.09 | ITA (Forehead) | 41.16 [37.97, 44.48] | 2.52 [-3.65, 20.77] | -53.55 [-68.03, -46.24] | -68.03 [-68.06, -53.55] | 41.96 [39.23, 44.48] | 20.77 [16.85, 24.83] | -53.55 [-68.03, -46.24] | -68.03 [-68.06, -53.55] |
|  |  | ITA (DP) | 39.68 [33.11, 44.68] | 4.10 [-13.01, 22.28] | -60.53 [-75.10, -36.92] | -60.53 [-82.47, -36.92] | 39.68 [34.58, 41.46] | 10.64 [-4.70, 24.11] | -60.53 [-75.10, -36.92] | -60.53 [-82.47, -36.92] |
|  |  | MST (Forehead) | 4 [2, 4] | 6 [4, 7] | 8 [7, 9] | 9 [8, 9] | 2 [2, 4] | 4 [2, 6] | 8 [7, 9] | 9 [8, 9] |
|  |  | MST (DP) | 2 [1, 4] | 7 [2, 7] | 8 [7, 10] | 10 [7, 10] | 2 [1, 4] | 4 [2, 7] | 8 [7, 10] | 10 [7, 10] |
|  |  | %Mod | 4.20 [2.90, 5.30] | 1.90 [1.00, 3.30] | 2.50 [1.60, 5.20] | 2.20 [1.10, 3.40] | 4.40 [2.40, 5.50] | 4.00 [2.70, 5.70] | 2.50 [1.60, 5.20] | 2.20 [1.10, 3.40] |
| Shenzhen (2024) PC-60NW | 81 | ITA (Forehead) | 40.97 [36.65, 45.52] | 19.24 [-2.77, 22.88] | -54.54 [-63.53, -32.14] | -63.53 [-73.37, -54.54] | 41.53 [38.52, 45.82] | 19.24 [-14.84, 22.88] | -50.02 [-63.53, -32.14] | -63.53 [-73.37, -54.54] |
|  |  | ITA (DP) | 28.17 [24.33, 36.70] | 3.36 [-26.37, 12.68] | -50.20 [-68.77, -24.57] | -50.20 [-70.41, -39.31] | 35.95 [28.10, 39.41] | 3.36 [-11.47, 15.82] | -39.31 [-68.77, -24.57] | -50.20 [-70.41, -39.31] |
|  |  | MST (Forehead) | 4 [1, 4] | 5 [5, 7] | 8 [8, 8] | 8 [8, 8] | 4 [1, 4] | 5 [3, 8] | 8 [8, 8] | 8 [8, 8] |
|  |  | MST (DP) | 2 [1, 5] | 5 [5, 7] | 8 [7, 9] | 8 [8, 8] | 2 [1, 4] | 7 [3, 8] | 8 [7, 9] | 8 [8, 8] |
|  |  | %Mod | 2.80 [1.60, 4.70] | 3.30 [2.40, 5.10] | 1.00 [0.60, 2.50] | 1.50 [0.60, 3.20] | 3.30 [1.80, 5.60] | 4.00 [2.40, 7.30] | 1.30 [0.60, 2.80] | 2.10 [0.70, 3.50] |
| Zug Medical Systems<br>SAS SPLF-SE with<br>SpotLife Reusable<br>Probe | 75 | ITA (Forehead) | 43.69 [40.97, 47.58] | 15.16 [4.21, 21.28] | -57.20 [-58.96, -39.60] | -58.25 [-58.96, -57.20] | 40.97 [36.65, 44.48] | 15.16 [1.00, 21.04] | -54.54 [-58.25, -39.60] | -58.25 [-63.53, -54.54] |
|  |  | ITA (DP) | 33.85 [27.27, 39.41] | 2.02 [-6.41, 10.64] | -61.09 [-67.15, -38.43] | -65.81 [-67.15, -61.09] | 30.29 [24.67, 35.95] | 6.07 [-11.47, 24.75] | -50.20 [-65.81, -38.85] | -50.20 [-67.15, -39.31] |
|  |  | MST (Forehead) | 2 [2, 4] | 5 [4, 7] | 8 [8, 9] | 8 [8, 9] | 4 [1, 4] | 5 [4, 8] | 8 [8, 9] | 8 [8, 9] |
|  |  | MST (DP) | 2 [1, 4] | 7 [4, 7] | 8 [7, 9] | 8 [8, 8] | 2 [1, 5] | 5 [2, 8] | 8 [7, 9] | 8 [8, 8] |
|  |  | %Mod | 2.00 [1.40, 3.80] | 4.20 [1.80, 6.20] | 1.50 [1.10, 2.50] | 1.50 [1.10, 2.42] | 2.30 [1.50, 4.40] | 3.90 [2.20, 6.00] | 1.50 [1.10, 2.80] | 1.50 [1.10, 2.90] |
| Compass Health<br>POX-ROS | 12 | ITA (Forehead) | 32.79 [31.54, 40.24] | 12.09 [3.35, 18.03] | -60.45 [-70.27, -51.44] | -66.82 [-70.27, -53.32] | 38.90 [31.95, 42.16] | 21.34 [7.62, 23.02] | -60.45 [-66.82, -51.44] | -64.00 [-66.82, -54.72] |
|  |  | ITA (DP) | 28.64 [11.48, 33.29] | 8.24 [-11.29, 19.25] | -57.66 [-72.04, -43.71] | -57.66 [-72.04, -45.82] | 28.64 [11.48, 30.29] | 8.24 [-1.84, 27.24] | -61.80 [-68.65, -45.82] | -61.80 [-68.65, -48.78] |
|  |  | MST (Forehead) | 5 [2, 5] | 6 [4, 7] | 8 [8, 10] | 9 [8, 10] | 5 [2, 5] | 5 [4, 8] | 8 [8, 10] | 8 [8, 10] |
|  |  | MST (DP) | 5 [1, 6] | 7 [4, 7] | 9 [8, 10] | 9 [8, 10] | 2 [1, 6] | 5 [4, 8] | 8 [8, 10] | 8 [8, 10] |
|  |  | %Mod | 0.50 [0.40, 1.10] | 2.00 [0.90, 5.00] | 1.50 [0.70, 5.20] | 1.80 [0.80, 5.80] | 0.70 [0.40, 1.30] | 3.00 [1.00, 5.40] | 2.20 [1.10, 6.10] | 2.40 [1.30, 5.60] |
| Beijing Choice<br>MD300M | 40 | ITA (Forehead) | 35.61 [32.78, 39.93] | 17.97 [9.18, 25.65] | -51.72 [-58.69, -45.26] | -58.69 [-71.89, -51.72] | 38.90 [34.88, 40.55] | 17.97 [9.18, 24.39] | -51.72 [-58.69, -46.24] | -58.69 [-71.89, -51.72] |
|  |  | ITA (DP) | 23.42 [15.00, 26.31] | 10.44 [6.69, 23.49] | -63.93 [-73.00, -44.50] | -63.93 [-84.81, -63.93] | 21.98 [14.82, 26.82] | 17.06 [-4.70, 26.63] | -56.96 [-63.93, -31.01] | -63.93 [-73.00, -44.50] |
|  |  | MST (Forehead) | 4 [1, 5] | 5 [5, 8] | 9 [8, 10] | 9 [9, 10] | 2 [1, 5] | 5 [4, 8] | 9 [8, 10] | 9 [8, 10] |
|  |  | MST (DP) | 4 [3, 5] | 5 [2, 8] | 8 [8, 10] | 8 [8, 10] | 4 [2, 5] | 5 [2, 8] | 8 [7, 10] | 9 [7, 10] |
|  |  | %Mod | 1.10 [0.80, 2.30] | 2.30 [0.90, 4.20] | 0.80 [0.50, 3.10] | 0.70 [0.30, 4.30] | 0.90 [0.70, 1.60] | 2.60 [1.20, 4.70] | 0.90 [0.60, 2.80] | 0.80 [0.40, 2.80] |
| Nonin PalmSAT 2500 | 36 | ITA (Forehead) | 38.03 [34.88, 42.67] | 11.06 [8.44, 12.82] | -51.23 [-51.72, -38.43] | -51.72 [-58.69, -51.23] | 39.76 [35.61, 43.69] | 15.79 [3.37, 25.65] | -51.72 [-58.96, -39.52] | -58.96 [-71.89, -51.72] |
|  |  | ITA (DP) | 25.48 [11.95, 36.13] | 8.17 [1.39, 22.26] | -63.93 [-68.35, -56.96] | -63.93 [-73.00, -44.50] | 25.48 [16.52, 32.46] | 5.56 [-18.99, 23.49] | -63.93 [-68.35, -44.50] | -63.93 [-73.00, -44.50] |
|  |  | MST (Forehead) | 2 [1, 5] | 5 [4, 8] | 9 [8, 9] | 9 [9, 9] | 2 [1, 5] | 6 [4, 8] | 9 [8, 10] | 9 [8, 10] |
|  |  | MST (DP) | 3 [2, 5] | 6 [2, 6] | 8 [7, 9] | 8 [8, 9] | 3 [2, 5] | 5 [4, 8] | 8 [7, 10] | 8 [8, 10] |
|  |  | %Mod | 1.00 [0.70, 1.50] | 1.80 [1.10, 2.80] | 0.60 [0.30, 1.60] | 0.50 [0.30, 0.70] | 1.00 [0.70, 1.50] | 2.40 [1.30, 4.00] | 1.00 [0.50, 2.50] | 1.20 [0.60, 3.60] |
| Covidien PM1000N | 59 | ITA (Forehead) | 38.03 [35.67, 42.67] | 10.74 [8.44, 21.49] | -39.52 [-51.72, -38.16] | -51.72 [-58.69, -51.23] | 39.37 [34.53, 43.50] | 14.29 [3.56, 21.04] | -51.46 [-59.99, -40.73] | -58.69 [-66.82, -53.55] |
|  |  | ITA (DP) | 28.92 [21.63, 41.76] | 14.12 [1.33, 22.26] | -63.93 [-68.35, -53.68] | -63.93 [-73.00, -44.50] | 31.16 [24.20, 36.15] | 16.20 [-3.60, 23.49] | -57.66 [-66.69, -43.71] | -61.80 [-67.37, -44.20] |
|  |  | MST (Forehead) | 4 [1, 5] | 5 [4, 8] | 8 [8, 9] | 9 [9, 9] | 4 [1, 5] | 5 [4, 8] | 8 [7, 10] | 8 [8, 10] |
|  |  | MST (DP) | 3 [2, 5] | 4 [4, 6] | 8 [7, 9] | 8 [8, 9] | 4 [1, 6] | 5 [2, 7] | 8 [7, 10] | 8 [7, 10] |
|  |  | %Mod | 1.00 [0.80, 1.50] | 2.20 [1.40, 4.60] | 0.60 [0.30, 1.50] | 0.50 [0.30, 0.70] | 2.00 [0.90, 3.80] | 3.10 [1.40, 5.30] | 1.60 [0.80, 4.70] | 1.80 [0.90, 5.30] |
| Shenzhen AM801 | 10 | ITA (Forehead) | 40.24 [31.95, 43.50] | 12.25 [0.72, 22.77] | -56.14 [-66.82, -43.04] | -66.82 [-71.89, -56.14] | 40.24 [31.95, 43.50] | 20.71 [3.56, 25.75] | -56.14 [-66.82, -43.04] | -65.41 [-70.62, -64.00] |
|  |  | ITA (DP) | 28.64 [15.00, 30.00] | 8.24 [-11.35, 27.24] | -66.69 [-68.79, -57.66] | -57.66 [-84.81, -54.85] | 30.00 [18.64, 33.29] | 16.77 [-1.84, 27.42] | -66.28 [-68.79, -57.66] | -61.80 [-79.06, -57.66] |
|  |  | MST (Forehead) | 2 [1, 5] | 5 [2, 8] | 8 [8, 10] | 10 [8, 10] | 2 [1, 5] | 6 [2, 7] | 8 [8, 10] | 8 [8, 10] |
|  |  | MST (DP) | 4 [1, 5] | 7 [4, 8] | 9 [8, 9] | 9 [8, 9] | 1 [1, 5] | 5 [3, 7] | 9 [8, 9] | 8 [8, 9] |
|  |  | %Mod | 1.30 [0.80, 3.10] | 3.00 [1.00, 5.40] | 5.00 [2.80, 8.00] | 3.90 [2.40, 7.40] | 1.90 [0.80, 4.50] | 4.00 [1.40, 6.10] | 4.60 [2.50, 7.60] | 3.40 [2.20, 6.60] |
| Nonin Onyx Vantage 9590 | 35 | ITA (Forehead) | 35.61 [31.54, 40.55] | 20.78 [-10.06, 22.77] | -53.32 [-60.55, -43.66] | -60.55 [-71.89, -53.32] | 35.61 [31.95, 39.72] | 12.25 [-10.06, 21.85] | -53.32 [-64.00, -48.46] | -60.55 [-66.82, -53.32] |
|  |  | ITA (DP) | 25.48 [15.00, 30.22] | 19.25 [-20.26, 27.24] | -51.76 [-68.79, -45.82] | -51.76 [-84.81, -45.82] | 23.42 [11.48, 28.92] | 4.95 [-16.34, 19.25] | -57.66 [-62.80, -45.82] | -54.85 [-61.80, -45.82] |
|  |  | MST (Forehead) | 3 [1, 5] | 5 [4, 8] | 9 [8, 10] | 9 [9, 10] | 3 [1, 5] | 5 [4, 8] | 8 [8, 10] | 8 [8, 10] |
|  |  | MST (DP) | 2 [2, 5] | 5 [4, 8] | 8 [7, 10] | 7 [7, 10] | 4 [2, 5] | 5 [4, 8] | 8 [7, 10] | 8 [7, 10] |
|  |  | %Mod | 1.30 [0.60, 4.00] | 1.40 [0.60, 3.40] | 2.00 [0.70, 3.90] | 2.20 [0.30, 4.10] | 1.70 [0.70, 4.50] | 2.50 [0.90, 4.20] | 2.60 [1.10, 5.60] | 2.60 [1.00, 5.72] |
| Choicemed MD300CN350R | 14 | ITA (Forehead) | 35.61 [31.22, 40.24] | 12.09 [-10.06, 20.71] | -53.32 [-70.27, -51.44] | -66.82 [-70.27, -53.32] | 35.61 [31.54, 42.16] | 12.25 [2.69, 23.02] | -58.96 [-66.82, -51.44] | -64.00 [-70.27, -58.94] |
|  |  | ITA (DP) | 28.64 [18.64, 31.16] | 18.51 [-20.26, 21.30] | -57.66 [-72.04, -45.82] | -57.66 [-72.04, -45.82] | 28.92 [18.64, 31.16] | 2.39 [-11.30, 18.51] | -60.10 [-66.28, -43.71] | -61.80 [-68.65, -45.82] |
|  |  | MST (Forehead) | 5 [1, 5] | 5 [2, 7] | 9 [8, 10] | 9 [8, 10] | 2 [1, 5] | 5 [2, 8] | 8 [8, 10] | 8 [8, 10] |
|  |  | MST (DP) | 4 [1, 5] | 5 [3, 7] | 9 [8, 10] | 9 [8, 10] | 4 [1, 5] | 5 [3, 7] | 8 [7, 10] | 8 [8, 10] |
|  |  | %Mod | 1.10 [0.60, 2.90] | 2.10 [0.60, 4.60] | 3.40 [0.90, 6.60] | 2.50 [0.80, 5.80] | 1.30 [0.70, 3.40] | 2.10 [0.60, 4.30] | 2.50 [1.30, 5.10] | 2.10 [1.00, 4.50] |
| Masimo (2024) Mightysat | 78 | ITA (Forehead) | 38.40 [35.35, 43.69] | 15.16 [8.70, 21.28] | -59.99 [-70.08, -43.61] | -70.08 [-70.08, -59.99] | 40.97 [38.40, 43.69] | 15.16 [4.21, 21.28] | -55.36 [-59.99, -39.60] | -59.99 [-72.28, -58.96] |
|  |  | ITA (DP) | 35.24 [27.27, 47.60] | 2.02 [-6.41, 14.96] | -60.16 [-66.83, -38.43] | -66.83 [-69.20, -60.16] | 35.24 [27.27, 37.28] | 2.02 [-6.41, 14.96] | -57.14 [-61.09, -28.63] | -61.09 [-65.81, -60.16] |
|  |  | MST (Forehead) | 2 [2, 4] | 6 [4, 8] | 8 [8, 9] | 9 [8, 9] | 2 [2, 4] | 6 [4, 7] | 8 [8, 9] | 8 [8, 9] |
|  |  | MST (DP) | 2 [2, 4] | 5 [4, 7] | 8 [8, 9] | 8 [8, 9] | 2 [1, 4] | 5 [4, 7] | 8 [7, 9] | 8 [8, 9] |
|  |  | %Mod | 2.50 [1.80, 3.80] | 3.90 [2.48, 5.30] | 1.80 [1.20, 3.50] | 1.80 [1.20, 3.80] | 2.50 [1.70, 3.80] | 3.80 [2.40, 5.20] | 1.80 [1.20, 3.30] | 1.80 [1.20, 3.60] |
| Acare (2023) Ab-F1/Ah-S1/Ah-M1 | 56 | ITA (Forehead) | 37.54 [34.88, 42.67] | 9.18 [-5.29, 16.59] | -53.32 [-60.55, -42.00] | -60.55 [-71.89, -53.32] | 35.85 [32.95, 42.67] | 9.18 [-7.51, 23.74] | -53.32 [-60.55, -42.57] | -59.99 [-71.89, -53.32] |
|  |  | ITA (DP) | 28.92 [18.29, 30.22] | 6.69 [-11.27, 9.73] | -59.38 [-68.35, -51.76] | -51.76 [-84.81, -45.82] | 26.31 [18.64, 30.22] | -4.70 [-20.26, 12.84] | -59.38 [-66.73, -45.30] | -51.76 [-66.83, -43.71] |
|  |  | MST (Forehead) | 4 [2, 5] | 5 [5, 7] | 9 [8, 10] | 9 [9, 10] | 4 [1, 5] | 6 [4, 8] | 8 [8, 10] | 9 [8, 10] |
|  |  | MST (DP) | 5 [2, 5] | 6 [2, 8] | 8 [7, 10] | 7 [7, 10] | 3 [2, 5] | 6 [2, 8] | 8 [7, 10] | 8 [7, 10] |
|  |  | %Mod | 1.20 [0.80, 3.00] | 2.80 [0.90, 4.40] | 2.70 [1.10, 5.70] | 2.10 [0.20, 5.10] | 1.70 [0.90, 3.60] | 3.20 [1.50, 5.00] | 1.40 [0.50, 4.10] | 1.70 [0.50, 3.80] |
| Philips M1020B IntelliVue with M1191BL Probe | 71 | ITA (Forehead) | 38.91 [34.33, 41.15] | 18.28 [-18.79, 22.91] | -47.91 [-60.45, -42.57] | -60.45 [-71.89, -51.44] | 41.00 [39.02, 44.48] | 21.01 [14.05, 23.62] | -51.44 [-67.50, -42.71] | -67.50 [-68.06, -60.45] |
|  |  | ITA (DP) | 25.98 [19.90, 40.20] | 25.19 [-9.08, 27.26] | -66.69 [-68.79, -43.71] | -68.65 [-84.81, -43.71] | 30.85 [21.04, 39.68] | 19.13 [2.21, 26.56] | -66.69 [-70.41, -42.89] | -70.41 [-77.28, -68.65] |
|  |  | MST (Forehead) | 4 [2, 5] | 5 [4, 8] | 8 [8, 10] | 8 [8, 10] | 2 [1, 5] | 4 [4, 8] | 8 [7, 9] | 8 [8, 9] |
|  |  | MST (DP) | 2 [2, 6] | 5 [2, 8] | 8 [8, 9] | 9 [8, 9] | 2 [1, 6] | 4 [2, 8] | 8 [7, 10] | 8 [8, 10] |
|  |  | %Mod | 2.70 [2.00, 4.08] | 2.40 [1.40, 6.50] | 1.20 [0.60, 2.32] | 0.60 [0.40, 1.50] | 3.00 [1.60, 5.70] | 5.00 [3.70, 6.90] | 0.90 [0.50, 2.50] | 0.60 [0.50, 1.50] |
| CMS Zacurate 500C | 15 | ITA (Forehead) | 38.90 [31.95, 41.13] | 16.59 [-10.06, 22.77] | -66.82 [-70.27, -43.66] | -66.82 [-71.49, -51.44] | 35.61 [31.95, 41.13] | 14.45 [3.56, 23.18] | -56.14 [-66.82, -43.66] | -64.00 [-70.27, -56.14] |
|  |  | ITA (DP) | 28.92 [15.00, 32.22] | 4.95 [-20.26, 27.24] | -57.66 [-72.04, -43.71] | -57.66 [-81.62, -43.71] | 28.92 [11.48, 32.22] | 12.50 [-1.84, 27.24] | -66.28 [-68.79, -54.85] | -61.80 [-72.04, -54.85] |
|  |  | MST (Forehead) | 3 [2, 5] | 5 [4, 8] | 8 [8, 10] | 9 [8, 10] | 5 [2, 5] | 6 [4, 8] | 8 [8, 10] | 8 [8, 10] |
|  |  | MST (DP) | 2 [2, 5] | 5 [3, 8] | 8 [8, 10] | 8 [8, 10] | 2 [2, 6] | 6 [3, 8] | 8 [8, 10] | 8 [8, 10] |
|  |  | %Mod | 0.80 [0.50, 1.40] | 2.85 [1.00, 4.60] | 1.50 [0.70, 4.50] | 2.60 [0.60, 5.70] | 1.00 [0.50, 1.90] | 4.20 [2.80, 6.50] | 2.40 [1.20, 5.60] | 2.20 [1.20, 5.30] |
| Covidien (2022)<br>Nellcor PM10N | 14 | ITA (Forehead) | 38.03 [35.32, 40.55] | 9.18 [-18.79, 24.39] | -58.69 [-58.94, -40.55] | -58.94 [-71.89, -58.69] | 40.55 [38.03, 45.92] | 9.18 [8.70, 23.88] | -51.48 [-58.69, -40.55] | -58.69 [-58.94, -51.48] |
|  |  | ITA (DP) | 25.48 [21.63, 33.04] | 6.69 [-35.93, 17.06] | -63.93 [-68.35, -59.38] | -63.93 [-84.81, -60.10] | 28.92 [21.98, 36.05] | 10.44 [-5.93, 17.13] | -59.38 [-63.93, -30.35] | -60.10 [-63.93, -44.20] |
|  |  | MST (Forehead) | 4 [1, 5] | 5 [5, 8] | 8 [8, 10] | 9 [8, 10] | 2 [1, 5] | 6 [4, 8] | 8 [8, 10] | 8 [8, 10] |
|  |  | MST (DP) | 4 [2, 5] | 8 [2, 8] | 8 [8, 10] | 9 [8, 10] | 2 [1, 5] | 6 [4, 8] | 8 [7, 10] | 8 [8, 10] |
|  |  | %Mod | 1.00 [0.70, 1.40] | 1.00 [0.70, 2.60] | 2.20 [0.70, 4.70] | 2.80 [0.30, 5.80] | 1.30 [0.70, 2.60] | 3.80 [2.50, 5.90] | 2.30 [0.70, 6.30] | 5.30 [1.00, 7.40] |
| Utech UT100 | 39 | ITA (Forehead) | 34.88 [33.85, 45.61] | 9.82 [3.37, 24.93] | -45.26 [-51.72, -39.52] | -51.72 [-60.55, -51.23] | 34.88 [32.81, 38.90] | 23.74 [5.59, 25.65] | -51.72 [-60.55, -45.26] | -60.55 [-71.89, -51.72] |
|  |  | ITA (DP) | 31.36 [16.63, 33.04] | 5.56 [-9.95, 14.79] | -51.76 [-68.35, -44.50] | -51.76 [-73.00, -44.50] | 31.36 [16.63, 33.04] | 12.84 [-4.70, 22.26] | -56.96 [-68.35, -44.50] | -51.76 [-73.00, -44.50] |
|  |  | MST (Forehead) | 4 [1, 4] | 5 [4, 8] | 9 [8, 9] | 9 [9, 9] | 2 [1, 4] | 5 [4, 8] | 9 [8, 10] | 9 [8, 10] |
|  |  | MST (DP) | 5 [2, 5] | 5 [4, 8] | 8 [8, 9] | 8 [8, 8] | 4 [2, 5] | 5 [2, 8] | 8 [8, 10] | 8 [8, 10] |
|  |  | %Mod | 0.90 [0.70, 1.10] | 2.20 [0.80, 4.20] | 0.70 [0.50, 1.60] | 0.50 [0.30, 0.70] | 1.10 [0.80, 3.10] | 2.50 [1.20, 4.20] | 1.20 [0.70, 3.30] | 1.20 [0.60, 3.70] |
| Nihon Kohden OLV-4202 with NK-SPO2-ADVT Probe | 64.03 | ITA (Forehead) | 35.61 [32.01, 41.15] | 24.34 [14.05, 26.72] | -45.26 [-67.50, -39.83] | -67.50 [-71.89, -53.32] | 39.55 [35.61, 44.83] | 22.45 [22.06, 25.88] | -42.71 [-67.97, -39.83] | -67.97 [-67.97, -53.32] |
|  |  | ITA (DP) | 28.64 [23.42, 30.22] | 19.13 [9.19, 26.58] | -45.82 [-77.28, -29.87] | -77.28 [-84.81, -45.82] | 25.98 [23.42, 31.50] | 18.38 [10.64, 27.24] | -68.77 [-69.25, -20.30] | -69.25 [-77.28, -45.82] |
|  |  | MST (Forehead) | 3 [2, 5] | 5 [5, 6] | 8 [8, 10] | 8 [8, 10] | 2 [1, 4] | 5 [4, 8] | 8 [7, 10] | 9 [8, 10] |
|  |  | MST (DP) | 4 [1, 6] | 5 [4, 6] | 8 [8, 10] | 8 [8, 10] | 4 [1, 6] | 4 [4, 8] | 9 [7, 10] | 9 [8, 10] |
|  |  | %Mod | 1.00 [0.80, 1.80] | 5.60 [4.07, 7.60] | 1.50 [0.80, 5.00] | 5.40 [4.40, 6.00] | 1.30 [0.80, 2.70] | 5.10 [3.30, 7.00] | 2.50 [1.30, 4.40] | 4.00 [1.00, 5.80] |
| EDAN (2020) H100b | 44 | ITA (Forehead) | 34.88 [32.81, 40.55] | 11.06 [-7.51, 16.79] | -51.23 [-51.72, -38.43] | -51.72 [-58.69, -51.23] | 43.48 [38.40, 45.93] | 12.82 [8.89, 21.49] | -47.91 [-58.69, -31.93] | -58.69 [-61.99, -51.72] |
|  |  | ITA (DP) | 24.45 [21.98, 33.04] | -7.35 [-31.34, 22.26] | -63.93 [-68.35, -56.96] | -63.93 [-73.00, -44.50] | 34.54 [21.98, 36.20] | 10.44 [-13.62, 18.58] | -63.93 [-75.10, -43.61] | -63.93 [-75.95, -59.07] |
|  |  | MST (Forehead) | 4 [2, 4] | 8 [4, 8] | 9 [8, 9] | 9 [9, 9] | 2 [1, 5] | 5 [4, 8] | 9 [7, 10] | 9 [9, 10] |
|  |  | MST (DP) | 4 [2, 5] | 4 [2, 8] | 8 [7, 9] | 8 [8, 9] | 2 [1, 4] | 5 [4, 8] | 9 [7, 10] | 8 [8, 10] |
|  |  | %Mod | 1.00 [0.80, 1.80] | 1.70 [0.80, 3.60] | 0.60 [0.30, 1.60] | 0.50 [0.30, 0.70] | 1.40 [0.80, 3.70] | 3.00 [1.80, 4.20] | 1.30 [0.50, 4.50] | 0.60 [0.30, 2.20] |
| Shenzhen WIT-S300C | 62 | ITA (Forehead) | 43.48 [42.16, 43.83] | 20.78 [-18.79, 25.95] | -54.63 [-58.25, -47.91] | -57.20 [-66.82, -54.63] | 43.48 [38.91, 45.29] | 17.97 [3.35, 23.02] | -54.63 [-58.25, -47.91] | -57.20 [-66.82, -54.63] |
|  |  | ITA (DP) | 30.22 [20.94, 30.29] | 11.33 [-49.67, 32.46] | -62.80 [-67.15, -45.82] | -57.66 [-67.37, -45.82] | 30.29 [26.36, 38.64] | 9.73 [-11.27, 17.06] | -62.80 [-67.15, -52.59] | -57.66 [-67.37, -45.82] |
|  |  | MST (Forehead) | 2 [1, 3] | 5 [2, 8] | 8 [8, 10] | 8 [8, 10] | 2 [1, 4] | 5 [2, 8] | 8 [8, 10] | 8 [8, 10] |
|  |  | MST (DP) | 2 [2, 4] | 5 [3, 8] | 8 [8, 10] | 8 [8, 10] | 2 [2, 4] | 5 [2, 8] | 8 [8, 10] | 8 [8, 10] |
|  |  | %Mod | 1.40 [0.50, 3.90] | 4.40 [3.20, 6.30] | 3.30 [1.30, 6.30] | 2.40 [1.10, 6.00] | 2.90 [1.20, 4.50] | 1.60 [0.80, 4.80] | 2.80 [1.20, 6.00] | 2.40 [1.10, 6.00] |
| CONTEC CMS50M | 17 | ITA (Forehead) | 35.61 [31.22, 43.48] | 10.74 [7.42, 20.78] | -51.44 [-70.27, -42.57] | -66.82 [-71.49, -51.44] | 43.48 [35.39, 45.29] | 17.78 [8.44, 20.08] | -51.44 [-70.27, -42.57] | -66.82 [-71.49, -51.44] |
|  |  | ITA (DP) | 30.00 [16.47, 30.95] | 11.33 [-4.47, 14.12] | -57.66 [-72.04, -53.68] | -57.66 [-81.62, -43.71] | 30.00 [16.47, 32.22] | 11.33 [-0.58, 14.12] | -66.28 [-72.04, -53.68] | -57.66 [-81.62, -43.71] |
|  |  | MST (Forehead) | 4 [2, 4] | 5 [4, 7] | 8 [8, 10] | 9 [8, 10] | 2 [2, 4] | 6 [4, 7] | 8 [8, 10] | 9 [8, 10] |
|  |  | MST (DP) | 4 [1, 4] | 4 [4, 7] | 8 [8, 10] | 8 [8, 10] | 2 [1, 4] | 4 [4, 7] | 8 [8, 10] | 8 [8, 10] |
|  |  | %Mod | 1.60 [0.50, 5.60] | 1.60 [0.90, 4.40] | 2.60 [1.20, 5.30] | 2.60 [0.60, 5.70] | 2.20 [0.70, 5.90] | 2.10 [0.90, 5.20] | 3.20 [1.40, 5.70] | 2.60 [0.60, 5.70] |
| Shenzhen A310L | 18 | ITA (Forehead) | 32.79 [31.95, 40.24] | 16.59 [3.56, 22.77] | -56.88 [-66.82, -43.66] | -66.82 [-66.82, -53.32] | 35.96 [32.16, 43.50] | 18.03 [3.56, 22.77] | -53.32 [-64.00, -43.66] | -64.00 [-66.82, -58.94] |
|  |  | ITA (DP) | 18.64 [11.48, 28.64] | 18.51 [-1.84, 27.42] | -63.15 [-68.79, -45.82] | -57.66 [-68.65, -45.82] | 28.64 [15.00, 37.31] | 18.51 [-1.84, 27.42] | -61.80 [-68.65, -45.82] | -61.80 [-68.65, -57.66] |
|  |  | MST (Forehead) | 3 [1, 5] | 5 [2, 7] | 8 [8, 10] | 9 [8, 10] | 2 [1, 5] | 5 [2, 8] | 8 [8, 10] | 8 [8, 10] |
|  |  | MST (DP) | 2 [2, 5] | 5 [3, 7] | 9 [8, 10] | 9 [9, 10] | 2 [2, 5] | 5 [3, 8] | 8 [8, 10] | 9 [8, 10] |
|  |  | %Mod | 2.70 [0.80, 4.50] | 2.60 [0.90, 5.40] | 3.20 [1.20, 6.00] | 4.00 [1.50, 7.40] | 3.70 [1.20, 5.80] | 2.00 [0.80, 5.20] | 2.40 [1.20, 5.80] | 3.40 [1.80, 6.10] |
| Masimo (2020)<br>Mightysat Rx | 7 | ITA (Forehead) | 41.13 [35.61, 43.50] | 10.74 [6.40, 21.96] | -53.32 [-66.82, -48.46] | -54.63 [-66.82, -53.32] | 41.96 [37.61, 43.50] | 18.20 [10.74, 22.06] | -53.32 [-66.35, -42.71] | -56.14 [-66.82, -53.32] |
|  |  | ITA (DP) | 28.92 [18.64, 30.95] | 5.06 [-4.47, 14.12] | -53.68 [-62.80, -43.71] | -45.82 [-57.66, -43.71] | 28.92 [25.06, 30.95] | 10.26 [1.33, 23.54] | -53.68 [-62.80, -43.61] | -54.85 [-74.08, -43.71] |
|  |  | MST (Forehead) | 2 [1, 5] | 6 [2, 8] | 9 [8, 10] | 9 [8, 10] | 2 [1, 5] | 5 [2, 8] | 8 [7, 10] | 8 [8, 10] |
|  |  | MST (DP) | 2 [1, 4] | 4 [3, 8] | 8 [8, 10] | 8 [8, 10] | 2 [1, 4] | 4 [3, 8] | 8 [7, 10] | 8 [8, 10] |
|  |  | %Mod | 1.20 [0.50, 2.73] | 0.90 [0.60, 3.20] | 2.00 [0.90, 6.60] | 2.10 [0.70, 6.70] | 1.80 [0.80, 4.20] | 2.00 [0.70, 6.00] | 2.30 [1.30, 5.50] | 2.40 [0.80, 6.30] |
| Masimo Rad-G with<br>Rad-G Finger Clip<br>Sensor | 55.03 | ITA (Forehead) | 41.53 [40.97, 47.58] | 15.16 [-9.75, 21.28] | -55.36 [-63.53, -43.61] | -63.53 [-73.37, -55.36] | 42.16 [39.55, 46.26] | 15.16 [-7.62, 19.62] | -55.36 [-66.35, -33.46] | -66.35 [-73.37, -63.53] |
|  |  | ITA (DP) | 35.24 [28.10, 36.70] | 2.02 [-6.41, 15.82] | -57.14 [-61.09, -24.57] | -61.09 [-70.41, -39.31] | 34.58 [24.20, 37.28] | 2.02 [-6.17, 10.26] | -43.61 [-70.41, -28.63] | -70.41 [-74.08, -61.09] |
|  |  | MST (Forehead) | 4 [2, 4] | 5 [4, 7] | 8 [8, 8] | 8 [8, 8] | 2 [1, 4] | 5 [3, 7] | 8 [8, 8] | 8 [8, 8] |
|  |  | MST (DP) | 2 [1, 5] | 6 [4, 7] | 8 [7, 9] | 8 [8, 8] | 2 [1, 5] | 7 [3, 7] | 8 [7, 9] | 8 [8, 9] |
|  |  | %Mod | 2.80 [1.70, 4.30] | 4.40 [2.60, 6.20] | 1.30 [0.50, 2.80] | 1.50 [0.60, 3.80] | 3.00 [1.70, 4.50] | 4.60 [2.30, 7.50] | 1.60 [0.70, 3.10] | 1.60 [0.70, 3.50] |
| Bistos Co., Ltd. BT-710 | 61 | ITA (Forehead) | 40.55 [38.90, 43.48] | 9.18 [-11.97, 9.22] | -45.26 [-60.55, -42.57] | -60.55 [-71.89, -53.32] | 39.30 [35.61, 42.67] | 10.74 [-5.29, 23.02] | -51.44 [-58.25, -43.66] | -57.20 [-60.55, -53.32] |
|  |  | ITA (DP) | 21.98 [16.63, 30.00] | 9.73 [-28.79, 10.44] | -51.76 [-68.79, -45.82] | -51.76 [-84.81, -45.82] | 24.39 [19.90, 30.22] | 8.17 [-11.27, 14.12] | -43.71 [-67.15, -31.01] | -51.76 [-67.15, -43.71] |
|  |  | MST (Forehead) | 2 [1, 4] | 6 [2, 8] | 8 [8, 10] | 9 [9, 10] | 2 [1, 5] | 6 [2, 8] | 8 [8, 10] | 8 [8, 10] |
|  |  | MST (DP) | 2 [2, 5] | 8 [3, 8] | 8 [7, 10] | 7 [7, 10] | 4 [2, 5] | 5 [2, 8] | 8 [7, 10] | 8 [7, 10] |
|  |  | %Mod | 1.00 [0.80, 1.70] | 5.00 [2.90, 7.80] | 1.10 [0.70, 2.30] | 2.10 [0.20, 5.10] | 1.00 [0.70, 1.90] | 2.00 [0.90, 5.20] | 1.10 [0.60, 2.10] | 1.20 [0.50, 2.40] |
| BodyMed<br>BDMOXMTRBLK | 2 | ITA (Forehead) | 35.61 [31.54, 43.48] | 17.98 [6.40, 21.96] | -53.32 [-54.63, -42.57] | -54.63 [-66.82, -53.32] | 40.24 [32.61, 42.16] | 12.31 [8.44, 20.78] | -53.32 [-58.94, -43.04] | -54.63 [-66.82, -53.32] |
|  |  | ITA (DP) | 30.22 [28.64, 31.16] | 5.06 [-4.47, 19.25] | -53.68 [-62.80, -45.82] | -45.82 [-57.66, -10.91] | 30.22 [28.64, 30.95] | 11.33 [-0.58, 13.71] | -57.66 [-66.28, -45.82] | -45.82 [-60.10, -43.71] |
|  |  | MST (Forehead) | 5 [2, 5] | 6 [2, 8] | 9 [8, 9] | 9 [8, 9] | 2 [2, 5] | 6 [2, 8] | 8 [8, 10] | 8 [8, 10] |
|  |  | MST (DP) | 4 [1, 4] | 5 [4, 8] | 9 [8, 9] | 9 [8, 9] | 2 [1, 4] | 5 [4, 8] | 9 [8, 9] | 9 [8, 9] |
|  |  | %Mod | 0.80 [0.50, 3.40] | 1.00 [0.60, 3.40] | 1.90 [0.90, 5.90] | 1.70 [0.30, 6.90] | 0.90 [0.50, 3.42] | 1.10 [0.60, 4.80] | 2.20 [1.10, 5.10] | 1.80 [0.80, 4.70] |
| Masimo Rad-97 with<br>Adult Probe, Product<br>Code 4050 | 60 | ITA (Forehead) | 40.55 [38.91, 43.48] | 8.36 [-11.97, 21.34] | -47.91 [-58.69, -43.04] | -58.69 [-71.89, -51.72] | 39.55 [32.95, 43.48] | 8.70 [-2.97, 18.20] | -51.44 [-58.69, -39.60] | -58.69 [-68.03, -51.72] |
|  |  | ITA (DP) | 21.98 [11.95, 30.22] | 1.76 [-28.79, 29.93] | -66.28 [-68.35, -63.93] | -63.93 [-84.81, -44.50] | 28.17 [19.19, 35.95] | 4.95 [-11.27, 20.73] | -60.53 [-66.28, -44.20] | -57.66 [-63.93, -43.83] |
|  |  | MST (Forehead) | 3 [1, 5] | 5 [5, 8] | 9 [8, 10] | 9 [9, 10] | 2 [1, 5] | 5 [4, 7] | 8 [7, 9] | 9 [8, 9] |
|  |  | MST (DP) | 2 [2, 5] | 5 [3, 7] | 9 [8, 10] | 9 [8, 10] | 2 [1, 5] | 5 [2, 7] | 9 [7, 10] | 8 [8, 10] |
|  |  | %Mod | 1.30 [0.70, 2.20] | 2.20 [0.70, 3.20] | 1.40 [0.40, 4.60] | 0.80 [0.40, 4.60] | 2.00 [0.80, 3.80] | 3.60 [1.80, 6.90] | 2.30 [1.00, 5.30] | 2.60 [0.60, 5.30] |
| Guangdong Biolight<br>M800 | 43 | ITA (Forehead) | 35.85 [32.81, 39.30] | 8.44 [-18.79, 21.49] | -51.72 [-58.69, -43.66] | -58.69 [-71.89, -51.72] | 38.90 [32.81, 43.50] | 15.79 [3.37, 23.74] | -51.48 [-58.25, -43.66] | -58.25 [-58.69, -51.72] |
|  |  | ITA (DP) | 25.36 [16.63, 36.13] | 5.56 [-49.67, 23.49] | -44.50 [-63.93, -37.96] | -63.93 [-84.81, -44.50] | 30.22 [18.64, 36.05] | 5.56 [-9.95, 23.49] | -44.20 [-63.94, -31.01] | -63.93 [-67.15, -44.20] |
|  |  | MST (Forehead) | 4 [2, 5] | 8 [5, 8] | 9 [8, 10] | 9 [9, 10] | 2 [1, 5] | 6 [4, 8] | 8 [8, 10] | 8 [8, 10] |
|  |  | MST (DP) | 3 [2, 5] | 5 [5, 8] | 8 [8, 10] | 9 [8, 10] | 2 [2, 5] | 6 [2, 8] | 8 [7, 10] | 8 [7, 10] |
|  |  | %Mod | 1.00 [0.70, 1.50] | 2.70 [1.40, 4.60] | 0.80 [0.60, 2.50] | 0.70 [0.30, 4.00] | 1.20 [0.80, 3.50] | 2.70 [1.30, 4.20] | 1.20 [0.60, 3.40] | 1.90 [0.80, 6.00] |
| Masimo Rad-G with Adult Probe, Product Code 4050 | 55 | ITA (Forehead) | 35.61 [34.22, 39.30] | 25.65 [9.18, 25.95] | -58.69 [-66.82, -45.26] | -66.82 [-71.89, -58.69] | 37.54 [35.32, 45.92] | 16.79 [8.89, 25.65] | -51.44 [-66.82, -40.73] | -58.96 [-71.89, -53.25] |
|  |  | ITA (DP) | 24.45 [11.95, 29.97] | 22.66 [10.44, 31.81] | -60.79 [-73.00, -56.96] | -63.93 [-84.81, -57.66] | 29.44 [17.18, 36.20] | 17.13 [-5.02, 23.49] | -57.66 [-73.00, -38.43] | -63.93 [-73.00, -43.71] |
|  |  | MST (Forehead) | 4 [1, 5] | 5 [2, 8] | 9 [8, 10] | 9 [9, 10] | 2 [1, 5] | 6 [2, 8] | 9 [7, 10] | 9 [8, 10] |
|  |  | MST (DP) | 4 [2, 5] | 5 [3, 6] | 8 [8, 10] | 8 [8, 10] | 4 [2, 5] | 5 [2, 7] | 8 [8, 10] | 8 [8, 10] |
|  |  | %Mod | 1.20 [0.90, 2.00] | 3.80 [1.90, 5.90] | 1.10 [0.50, 6.20] | 4.70 [0.40, 6.90] | 1.50 [0.90, 2.60] | 2.50 [1.30, 4.60] | 1.30 [0.60, 4.90] | 1.50 [0.50, 5.30] |
| Guangdong Biolight M70 | 37 | ITA (Forehead) | 35.61 [35.61, 43.48] | 16.59 [3.35, 20.08] | -53.32 [-60.55, -43.66] | -60.55 [-71.89, -53.32] | 39.37 [35.61, 43.50] | 12.31 [-7.51, 16.85] | -53.32 [-58.25, -48.46] | -57.20 [-60.55, -51.94] |
|  |  | ITA (DP) | 30.00 [23.42, 30.95] | 1.33 [-11.29, 11.33] | -51.76 [-62.80, -45.82] | -51.76 [-84.81, -45.82] | 30.22 [23.42, 33.29] | -0.58 [-34.75, 11.33] | -54.85 [-62.80, -44.20] | -54.85 [-67.15, -44.60] |
|  |  | MST (Forehead) | 2 [2, 5] | 5 [4, 7] | 9 [8, 10] | 9 [9, 10] | 2 [1, 5] | 6 [4, 8] | 8 [8, 10] | 8 [8, 10] |
|  |  | MST (DP) | 4 [1, 5] | 6 [4, 8] | 8 [8, 10] | 8 [8, 10] | 2 [1, 5] | 6 [4, 8] | 8 [8, 10] | 8 [8, 10] |
|  |  | %Mod | 0.80 [0.60, 1.30] | 1.50 [0.70, 3.60] | 3.20 [0.80, 5.40] | 3.20 [0.30, 4.50] | 1.60 [0.80, 2.70] | 1.80 [0.90, 4.40] | 2.40 [1.20, 5.90] | 2.60 [1.10, 6.30] |
| Acare (2023) AH-M1 | 57 | ITA (Forehead) | 35.85 [32.95, 39.30] | 9.22 [-5.29, 20.76] | -51.72 [-60.55, -42.00] | -60.55 [-71.89, -51.72] | 38.90 [35.61, 43.48] | 9.18 [-5.29, 15.79] | -51.48 [-58.25, -40.73] | -58.25 [-60.55, -51.72] |
|  |  | ITA (DP) | 25.36 [17.18, 29.97] | 1.39 [-11.27, 11.64] | -59.38 [-66.69, -51.76] | -51.76 [-84.81, -44.50] | 28.92 [17.18, 33.04] | -7.35 [-27.34, 9.73] | -63.97 [-67.37, -44.50] | -51.76 [-67.37, -44.50] |
|  |  | MST (Forehead) | 4 [2, 5] | 5 [5, 8] | 9 [8, 10] | 9 [9, 10] | 2 [1, 5] | 6 [4, 8] | 8 [7, 10] | 8 [8, 10] |
|  |  | MST (DP) | 5 [2, 5] | 5 [5, 8] | 8 [7, 10] | 8 [8, 10] | 4 [1, 5] | 6 [4, 8] | 8 [7, 10] | 8 [8, 10] |
|  |  | %Mod | 1.50 [0.80, 3.20] | 1.90 [0.50, 4.40] | 1.10 [0.30, 5.40] | 0.70 [0.30, 4.50] | 2.30 [0.80, 4.50] | 2.70 [0.70, 4.90] | 1.60 [0.70, 6.00] | 1.40 [0.80, 5.80] |
| Acare (2020) AH-MX | 42 | ITA (Forehead) | 35.61 [32.81, 38.66] | 11.06 [-2.37, 21.49] | -51.23 [-51.72, -38.43] | -51.72 [-58.69, -51.23] | 35.61 [32.81, 45.61] | 11.06 [-2.37, 21.49] | -46.24 [-57.20, -40.73] | -57.20 [-58.69, -52.64] |
|  |  | ITA (DP) | 25.48 [16.63, 36.13] | 5.56 [-31.34, 31.71] | -63.93 [-68.35, -56.96] | -63.93 [-73.00, -44.50] | 30.22 [21.72, 36.13] | 5.56 [-10.86, 22.28] | -63.93 [-68.35, -36.92] | -67.15 [-70.18, -44.50] |
|  |  | MST (Forehead) | 4 [1, 5] | 5 [4, 8] | 9 [8, 9] | 9 [9, 9] | 4 [1, 5] | 6 [4, 8] | 8 [7, 10] | 8 [8, 10] |
|  |  | MST (DP) | 4 [2, 5] | 6 [2, 8] | 8 [7, 9] | 8 [8, 9] | 4 [1, 5] | 6 [2, 8] | 8 [7, 10] | 8 [7, 10] |
|  |  | %Mod | 1.00 [0.80, 1.60] | 1.20 [0.60, 2.23] | 0.60 [0.30, 1.60] | 0.50 [0.30, 0.70] | 1.00 [0.60, 2.60] | 2.10 [1.00, 4.10] | 1.10 [0.60, 2.40] | 1.10 [0.60, 2.20] |
| American Oxygen LK-87 | 20 | ITA (Forehead) | 39.56 [32.79, 43.50] | 16.59 [8.36, 21.85] | -61.48 [-70.27, -48.46] | -66.82 [-71.89, -61.48] | 39.76 [32.79, 43.50] | 13.52 [8.75, 22.77] | -64.00 [-66.82, -42.57] | -66.82 [-70.27, -64.00] |
|  |  | ITA (DP) | 19.90 [15.00, 30.00] | 8.24 [-10.51, 21.30] | -68.79 [-72.04, -57.66] | -57.66 [-84.81, -56.25] | 19.90 [15.00, 30.00] | 1.76 [-13.62, 17.13] | -61.80 [-68.79, -57.66] | -61.80 [-72.04, -57.66] |
|  |  | MST (Forehead) | 2 [1, 5] | 5 [5, 8] | 9 [8, 10] | 9 [8, 10] | 2 [1, 5] | 5 [4, 8] | 8 [8, 10] | 8 [8, 10] |
|  |  | MST (DP) | 4 [1, 5] | 5 [5, 8] | 9 [8, 10] | 8 [8, 10] | 2 [1, 5] | 5 [4, 8] | 8 [7, 10] | 8 [8, 10] |
|  |  | %Mod | 2.50 [0.90, 3.70] | 4.20 [1.40, 5.60] | 3.30 [2.00, 6.00] | 2.80 [1.50, 5.70] | 2.70 [1.20, 4.60] | 3.70 [1.40, 5.40] | 2.80 [1.60, 5.90] | 2.70 [1.70, 6.10] |

Distribution of Individual Typology Angle (ITA) at the forehead and dorsal distal phalanx (DP), Monk Skin Tone (MST) at the forehead and DP, and percent modulation (%Mod), an indicator of participant perfusion —measured by the Nellcor PM1000N and scaled by 1/10—across four ITA-defined skin color bins: light ( $ITA > 30^\circ$ ), medium ( $-30^\circ \leq ITA \leq 30^\circ$ ), dark ( $ITA < -30^\circ$ ), and very dark ( $ITA < -50^\circ$ ); note that “very dark” is a subset of the dark bin. Data for the 24 participant cohorts are shown on the left and data for the larger cohorts meeting anticipated 2025 U.S. Food and Drug Administration (FDA) and International Organization for Standardization (ISO) enrollment criteria are reported on the right. ITA and %Mod are reported as median [Q1, Q3], and MST as mode [minimum, maximum].

## Notes

### Competing Interest Statement

The authors have declared no competing interest.

### Clinical Protocols

https://openoximetry.org/study-protocols/

### Author Declarations

IRB of the University of California, San Francisco gave ethical approval for this work

### Summary of Updates

This manuscript revision includes an updated figure (Figure 1) and minor text changes to improve clarity.

