## Supplementary material for "Pulse oximeter performance and skin pigment: comparison of 34 oximeters using current and emerging regulatory frameworks": eMethods

Supplemental Digital Content

eMethods:

**Demographics and skin pigment assessment**

Study participants’ age, sex and US National Institutes of Health (NIH) race were self-reported. Participants’ height, weight, and finger diameter were measured. Percent modulation of infrared light was recorded from the clinical monitor oximeter (PM1000N, Medtronic, USA) and divided by 10. As previously described, two clinical research coordinators assigned subjective skin pigmentation data for each participant using a professionally printed version of the Monk Skin Tone (MST) scale.^1^^,^^2^ When in disagreement, a third observer was utilized to achieve concordance, and the median value was recorded. The MST values were assessed at the forehead with participants in front of a full spectrum lamp (Aputure Amaran 60 XS with mini light dome at 6500k, Aputure Imaging Industries Co. Ltd., Shenzhen, P.R.China).

Three ITA measurements were taken at each site, and the median was reported. The KM data were collected using the CM-SA Skin Analysis Software with the illuminant set to ‘D65’ and observer angle set to 10°. The ITA was calculated using the L* (lightness) and b* (yellow/blue component) values collected by the spectrophotometer using the standard ITA equation:

$ITA = atan\left( \frac{L*-50}{b*} \right)\cdot\frac{180}{\pi}$^3^

Further details on the skin pigment collection protocol and details on skin pigment characterization are previously published (Lipnick MS, Chen D, Law T, et al. Comparison of methods for characterizing skin pigment diversity in research cohorts. Preprint. medRxiv. 2025).

**Controlled Desaturation Protocol**

As previously described, study investigators controlled partial pressures of inspired oxygen, carbon dioxide, and nitrogen to achieve six stable “plateaus” of targeted arterial functional oxygen saturation (SaO_2_).^4^ Plateaus were distributed across three saturation ranges of 70–80%, 80–90% and 90–100% SaO_2_. A computer tracked a calculated oxygen saturation (ScO_2_) using the participant’s end-tidal oxygen saturation and SpO_2_ from the clinical reference pulse oximeter. Once the computer indicated a stable plateau was reached, arterial blood samples were taken from a radial artery catheter with each plateau sample collected 20–30 seconds apart. The SpO_2_ was recorded at the exact time of arterial blood gas sampling.

Blood gas samples were immediately analyzed using Radiometer ABL90 Flex Plus (ABL) (Radiometer, Copenhagen, Denmark) blood gas analyzers at the participant’s bedside. Each blood sample was run on at least two ABLs. If there was > 0.5% difference in SaO_2_ between the ABLs, then the sample was run on a third ABL. Between the SaO_2_ values obtained, if two values were within 0.5%, then the mean was used, otherwise, the samples were excluded from analysis.

*Pulse Oximeter Probe Placement*

Most pulse oximeters were randomly assigned to all digits with the following exceptions: devices 73.09 and 78 were not used on the 1st or 5th digit in either the 24 participant or maximum participant cohort analyses. Devices 81 and 59 were not used on the 1st or 5th digit in the 24 participant cohort analysis. Device 55.03 was used exclusively on the 5th digit to test.

*Prevention of Device Interference*

Study participants wore opaque finger shields (Masimo Ambient Light Finger Shields, Masimo Corporation, Irvine, CA) to prevent signal interference between adjacent oximeters. For fingertip devices with built-in displays, finger shields were cut so that the SpO_2_ display was visible.

**Statistical and sensitivity analyses**

*SaO_2_ Thresholds*

For simplicity, we labeled our SaO_2_ thresholds to match with labels for anticipated FDA and ISO thresholds (e.g. “70–80% SaO_2_” and “70–100% SaO_2_”). However, in reality these ranges are most accurately described as SaO_2_ 67–80% SaO_2_ and 67–100% SaO_2_ respectively.

*Primary Analysis: A_RMS_ and Differential Bias*

For the primary analysis, we used cohorts of 24 participants and skin color data from the forehead and DP. We used two metrics to define a ‘passing device’ based on what had been shared publicly by FDA and ISO at the time of study design: 1. A_RMS_; and 2. differential bias. 1. A_RMS_ with 95% CI < 3% (anticipated FDA) vs A_RMS_ point estimate ≤ 3% (anticipated ISO); 2. Differential bias point estimate with 95% CI < 3.5% for the 70–85% SaO_2_ range and < 1.5% for the 85–100% SaO_2_ range (anticipated FDA), vs point estimate ≤ 4% for the 70–85% SaO_2_ range and ≤ 2% for the 85–100% range (anticipated ISO).^5,6^ We calculated the overall A_RMS_ as the square root of the average of the squared difference between SpO_2_ and SaO_2_. We estimated the differential SpO_2_ bias and 95% confidence interval (CI) using a linear mixed effects (LME) model to estimate the predicted difference in bias for each device between two theoretical participants with an ITA difference of 100° (i.e., dark ITA -50° and light ITA 50°). Of note, the FDA’s draft guidance recommends devices pass the differential bias thresholds for both pairwise comparisons of MST groups 1–4, 5–7, and 8–10 and for a 100-point change in emitter sensor site ITA, while the anticipated ISO standards only recommend differential bias thresholds for 100-point changes in ITA.

To assess the ITA-derived differential bias, we fit an LME model with SpO_2_ as the outcome, including main effects of SaO_2_ and ITA, their interaction, and random intercepts and slopes for SaO_2_ by participant ID to account for individual variation in baseline SpO_2_ and the SpO_2_ and SaO_2_ relationship. Point estimates correspond to the change in average SpO_2_ bias associated with a 100° change in ITA; standard errors were then derived via the multivariate delta method using the model’s fixed-effects covariance matrix, and two-sided 95% CI were constructed as the point estimate plus or minus 1.96 times its standard error. For MST-derived differential bias, we categorized the MST scale into three bins: 1–4 (light), 5–7 (medium), and 8–10 (dark), and fit a LME model including dummy variables for the medium and dark groups (light as reference), interactions between SaO_2_ and each MST bin, and random intercepts and slopes for SaO_2_ by participant ID to account for within-subject differences in both SpO_2_ and SaO_2_. For each medium–light and dark–light contrast, we obtained standard errors and denominator degrees of freedom via the Satterthwaite approximation (lmerTest) and built two-sided 95% CI as the point estimate plus or minus the appropriate t-quantile (for df) times the standard error.

6.	Approach for Improving the Performance Evaluation of Pulse Oximeter Devices Taking into Consideration Skin Pigmentation, Race and Ethnicity:  Discussion Paper and Request for Feedback. https://www.fda.gov/media/173905/download?attachment
